# Dissecting shared genetic susceptibility between gastroesophageal reflux disease and psychiatric disorders beyond body mass index

**DOI:** 10.64898/2026.09.26.26364056

**Authors:** Xianjin Wang, Qingyi Zhang, Qinfeng Wu, Meng Zhang, Zekun Liu, Fen Hu, Yidi Zhou, Hongwei Yu, Kalim Ullah, Nan Zhang, Yingchao Song, Hongyang Zhao, Xiao Chang

## Abstract

Gastroesophageal reflux disease (GERD) exhibits substantial comorbidity with psychiatric disorders, yet the biological basis underlying this relationship remains unclear. Here, we integrated large-scale GWAS meta-analysis, Mendelian randomization, Genomic Structural Equation Modeling, local genetic correlation analysis, and single-cell transcriptomic enrichment mapping to investigate the shared genetic architecture linking GERD, psychiatric disorders, and body mass index (BMI). GERD demonstrated strongest genetic overlap with internalizing psychiatric disorders, particularly major depressive disorder and post-traumatic stress disorder, whereas the relationship with BMI was comparatively weaker and lacked significant bidirectional causal effects. Genomic SEM further showed that GERD preferentially loaded onto an internalizing latent factor independent of obesity-related liability. At the cellular level, GERD-associated genetic signals localized predominantly to interoceptive and limbic brain systems involved in visceral sensation, stress responsivity, and autonomic regulation. Together, these findings support a distinct neuropsychiatric component underlying GERD susceptibility and implicate centrally mediated brain–gut mechanisms beyond metabolic dysfunction.

## Introduction

Despite its high prevalence and clinical burden, the biological basis of gastroesophageal reflux disease (GERD) remains incompletely understood. Traditionally conceptualized as a disorder of impaired lower esophageal sphincter function and abnormal gastric physiology, GERD is increasingly recognized as a systemic disorder shaped by coordinated interactions among epithelial integrity, immune signaling, metabolic regulation, and neural control (Guan et al. 2025; Liu and Mei 2025; Paulrasu et al. 2025; Azer and Goosenberg 2025; Caldart et al. 2025).

Emerging evidence further implicates the brain–gut axis (BGA) in GERD pathophysiology, as neural circuits governing esophageal motility, visceral sensation, stress responsivity, and autonomic regulation contribute substantially to symptom generation and disease susceptibility (Bentley et al. 2024; Holtmann et al. 2025; Zheng and Tao 2025; Savarino et al. 2025). These observations have prompted growing interest in whether GERD shares common neurobiological mechanisms with psychiatric disorders.

Epidemiological studies consistently demonstrate elevated rates of depression, anxiety, and stress-related disorders among individuals with GERD (Chen et al. 2024; Ding et al. 2025; Pavić et al. 2025). Genome-wide association studies (GWAS) similarly support widespread genetic correlations between GERD and psychiatric phenotypes (An et al. 2019; Ong et al. 2022; A et al. 2024; Wei et al. 2026). However, interpretation of these relationships remains challenging because GERD also exhibits strong genetic overlap with obesity-related traits, particularly body mass index (BMI), which itself is genetically associated with multiple psychiatric conditions. As a result, the apparent GERD _–_ psychiatric relationship may arise from several partially overlapping mechanisms: shared neurobehavioral liability within the brain–gut axis, metabolic pathways related to adiposity and systemic inflammation, or downstream consequences of chronic disease burden. Distinguishing these components is essential for understanding whether psychiatric comorbidity reflects a core dimension of GERD biology or a secondary consequence of broader metabolic dysfunction.

Recent advances in multivariate statistical genetics provide an opportunity to disentangle these overlapping sources of genetic liability (He et al. 2025; Song et al. 2025; Zhi et al. 2025; Lu et al. 2025; Frei et al. 2025; Xu et al. 2025). In particular, Genomic Structural Equation Modeling (Genomic SEM) enables genome-wide partitioning of shared and trait-specific genetic variance across correlated phenotypes, facilitating the identification of latent biological dimensions underlying complex disease relationships. Rather than treating GERD, BMI, and psychiatric disorders as isolated traits, this framework allows their shared genetic architecture to be decomposed into separable neuropsychiatric and metabolic components. Such an approach is especially relevant for disorders involving the brain– gut axis, where behavioral, autonomic, and metabolic processes are highly interconnected at both physiological and genetic levels.

At the same time, single-cell transcriptomic atlases have transformed the interpretation of polygenic disease risk by enabling genetic associations to be mapped onto specific cellular populations across human tissues (Clevenger et al. 2023; Siletti et al. 2023). Integrative analyses combining GWAS with cell-type_–_resolved expression profiles have revealed that psychiatric and neurological risk variants frequently converge on restricted neuronal subtypes and functional neural circuits (Yao et al. 2025; Duncan et al. 2025). Yet whether GERD susceptibility similarly localizes to defined cellular populations within the brain–gut axis has not been systematically investigated. Resolving the cellular context of GERD-associated genetic variation may provide critical insight into how neurobiological and metabolic liabilities converge to influence visceral function and symptom perception.

Here, we integrated large-scale genomic analyses with cell-type–resolved transcriptomic mapping to define the biological architecture of GERD susceptibility. Using multi-trait GWAS approaches and Genomic SEM, we characterized the shared and independent genetic components linking GERD, psychiatric disorders, and BMI. We further refined GERD-associated loci through gene-level prioritization and functional annotation analyses and mapped polygenic risk onto high-resolution single-cell atlases of the human brain and esophageal mucosa. Together, these analyses provide a genome-to-cell framework for understanding GERD biology and clarify the extent to which psychiatric comorbidity reflects intrinsic brain – gut neurobiology versus obesity-related metabolic liability.

## Methods

### GWAS Data

GWAS summary statistics for GERD were obtained from a meta-analysis of UK Biobank and QSKIN cohorts(An et al. 2019) as well as the Million Veteran Program (MVP)(A et al. 2024). Summary statistics for psychiatric disorders, including attention-deficit/hyperactivity disorder (ADHD), anorexia nervosa (AN), anxiety disorder (ANX), autism spectrum disorder (ASD), bipolar disorder (BIP), major depressive disorder (MDD), obsessive-compulsive disorder (OCD), post-traumatic stress disorder (PTSD), and schizophrenia (SCZ), were obtained from the Psychiatric Genomics Consortium (PGC)(Sullivan et al. 2018). To capture metabolic contributions, GWAS summary statistics for BMI(Sidorenko et al. 2024) were additionally included. Detailed sample sizes and study characteristics are provided in Table S1.

### Estimation of genetic correlation

Genetic correlations (r_g_) for GERD with a range of psychiatric disorders were computed by applying linkage disequilibrium score regression (LDSC) to the respective GWAS summary statistics. In line with established protocols, the analysis was confined to single-nucleotide polymorphisms (SNPs) present in the HapMap3 panel. Heritability estimates on the observed scale (h²) for each trait were concurrently generated using the LDSC framework. The required linkage disequilibrium (LD) reference was derived from the European ancestry subset of the 1000 Genomes Project, specifically for HapMap3 variants (Bulik-Sullivan et al. 2015).

### Mendelian Randomization

Instrumental variables (IVs) for each exposure were obtained from the corresponding GWAS summary statistics using LD clumping with an r² threshold of 0.01 and a 10-Mb window to ensure independence among selected variants. Genome-wide significant SNPs passing the predefined P-value threshold were retained as candidate instruments. For each IV set, harmonized SNP-level data were extracted from both exposure and outcome datasets, aligning effect alleles and removing ambiguous or palindromic variants to prevent strand inconsistencies. Bidirectional Mendelian randomization (MR) was performed using the Inverse Variance Weighted (IVW) approach as implemented in the TwoSampleMR R package (v0.6.4).(Hemani et al. 2017, 2018)

### Local genetic correlation analysis

Local genetic correlations between GERD and psychiatric disorders were analysed using LAVA (Werme et al. 2022). This method partitions the genome into approximately independent, 1 Mb regions based on LD structure, resulting in 2495 loci per phenotype pair. Univariate tests first estimated local SNP heritability (h^2^_SNP_) for each trait; loci with non-significant h^2^_SNP_ (p < 0.05/2495) were excluded, retaining only phenotype-relevant regions. Bivariate analyses were then performed for loci with significant h^2^SNP in both traits to estimate local genetic correlations, with multiple testing corrected via Benjamini Hochberg FDR (q < 0.05).

### Gene-based Association and Functional Enrichment Analyses

Gene-level and pathway enrichment analyses were performed to characterize the biological mechanisms underlying GERD-associated genetic variation. Gene-based association testing was carried out using MAGMA v1.0.6 on three GERD GWAS datasets: the UKB + QSKIN cohort (An et al.)(An et al. 2019), MVP(A et al. 2024), and their fixed-effect meta-analysis. In MAGMA, SNPs were mapped to genes, and gene-level test statistics were derived by aggregating SNP-wise association signals while accounting for LD structure. LD-adjusted linear regression models were used to integrate the joint effects of SNPs within each gene. Multiple testing was controlled using Bonferroni correction within the MAGMA framework (de Leeuw et al. 2015). Genes with MAGMA gene-level P-values < 0.01 were retained from each dataset for enrichment testing.

Functional characterization of significant genes was subsequently performed through Gene Ontology (GO)(Kolberg et al. 2023) enrichment analysis using g:Profiler. Enrichment was evaluated across biological processes (BP), cellular components (CC), and molecular functions (MF). Gene sets obtained from each GERD GWAS dataset (An et al.(An et al. 2019), MVP(A et al. 2024), and the meta-analysis) were analyzed separately to identify convergent and dataset-specific functional signatures. GO terms with Benjamini-Hochberg adjusted *P*-values < 0.05 were considered statistically significant and retained for interpretation.

### condFDR analysis

The conditional false discovery rate (condFDR) framework was applied to enhance the discovery of GERD-associated loci and to identify variants jointly enriched across GERD and psychiatric phenotypes. CondFDR is built upon an empirical Bayesian model that incorporates cross-trait enrichment by leveraging association strengths from two GWAS datasets (Andreassen et al. 2014). This framework re-ranks SNPs for the primary trait (GERD) by adjusting their test statistics according to the degree of association with a secondary trait (e.g., MDD or SCZ), thereby increasing power to detect true associations even when individual SNPs do not meet genome-wide significance thresholds. To ensure robust enrichment modeling, SNPs with secondary-trait P-values ≥ 0.05 were excluded from the condFDR re-ranking step. This threshold focuses the analysis on variants with at least moderate evidence of association with the secondary trait, which improves stability of the conditional FDR estimation and aligns with prior applications of the method. For the remaining SNPs, condFDR values were estimated following the standard procedure, and variants were declared conditionally significant at condFDR < 0.05, consistent with established practice. As recommended for Bayesian FDR modeling, genomic regions with extreme linkage disequilibrium were excluded before model fitting, including the extended MHC region (chr6: 25–34 Mb) and chromosome 8p23.1 (chr8: 7.2–12.5 Mb).

### Genomic Structural Equation Modeling

To investigate the shared genetic architecture among the 11 phenotypes (GERD, ADHD, AN, ANX, ASD, BIP, MDD, OCD, PTSD, SCZ, and BMI), we applied the Genomic SEM framework for multivariate genetic analysis(Grotzinger et al. 2019). We first performed exploratory factor analysis (EFA) on the LDSC-derived genetic covariance matrix to determine the number of latent factors. Parallel analysis (paLDSC) revealed that the observed eigenvalues intersected the 95th percentile of those generated from simulated data at the third component, falling below the simulated values from the fourth component onward, indicating that subsequent components no longer reflected meaningful shared genetic signal(Figure S1). Building on this, we further compared several candidate models: a 2-factor solution was evaluated as a more parsimonious alternative for sensitivity analysis, and given the residual correlations among F1, F2, and F3 in the 3-factor solution, we additionally fit a p-factor model (i.e., assuming a single general genetic factor underlying all phenotypes) to test for a higher-order common structure. Based on AIC and related fit indices, the 3-factor model outperformed both the 2-factor and the p-factor models and was therefore retained as the optimal solution(Table S2). With the factor number established, we then fit the 3-factor model using confirmatory factor analysis (CFA). In the model specification, phenotypes with EFA loadings greater than 0.2 were allowed to load onto the corresponding factor; for the final factor assignment and biological interpretation, each phenotype was assigned to the factor on which it showed the highest loading, to ensure interpretability and unambiguous factor membership. Given that GERD was the primary focus of the present study, we centered downstream analyses on F1, the factor that included GERD, and performed a multivariate GWAS of F1 within the Genomic SEM framework to identify candidate risk loci driving the shared genetic signal underlying this factor.

### Tissue and Cell-Type-Specific Enrichment Analysis

To identify disease-relevant enriched cell types associated with GERD, we performed TDEP(Yao et al. 2025) analysis at two levels. At the genome-wide level, GERD GWAS summary statistics were directly used as TDEP input to assess cell-type enrichment across genome-wide association signals. At the direction-specific level, Genomic SEM analysis revealed opposing genetic correlations between GERD and psychiatric trait factors—positive with the F1 factor (MDD, ADHD, ANX, PTSD) and negative with the F3 factor (BIP, SCZ). To investigate whether these two opposing genetic sharing mechanisms correspond to distinct cell-type biological substrates, we performed LAVA local genetic correlation analysis between GERD and each of the six psychiatric traits, obtaining the local genetic correlation coefficient (rho) for each genomic region. We then applied the following direction-specific filtering strategy to the GERD GWAS signals: regions where rho directions conflicted across traits were excluded to avoid directional confounding; the F1 input file retained only GWAS signals within conflict-free regions showing rho > 0 for F1 traits, while the F3 input file retained only signals within conflict-free regions showing rho < 0 for F3 traits; SNPs outside the retained regions had their z-values set to 0 and p-values set to 1 to mask non-target directional signals; the MHC region (chr6: 25–35 Mb) was also masked due to its high polymorphism and complex LD structure. The two resulting direction-specific summary statistics files were used as TDEP inputs for the F1 and F3 directions, respectively.

## Results

### Meta-analysis of GERD GWAS from UKB, QSKIN and MVP Consortia

We leveraged publicly available GERD GWAS summary statistics from An et al. (An et al. 2019), excluding the 23andMe cohort due to data-sharing restrictions. This dataset primarily consists of European-ancestry participants from the UK Biobank and the QSKIN study. To further increase power, we incorporated European-ancestry data from MVP (A et al. 2024) and performed a fixed-effects inverse-variance-weighted meta-analysis. In total, the combined analysis included 247,513 cases and 497,816 controls (Table S1). We identified 185 genome-wide significant loci (P < 5 × 10^-8^) that were independent at LD r^2^ < 0.05. Of these, 107 loci represent previously unreported associations with GERD (Table S3), as further illustrated in Figure 1A, which quantifies the overlap of associated loci across datasets.

**Figure 1.**
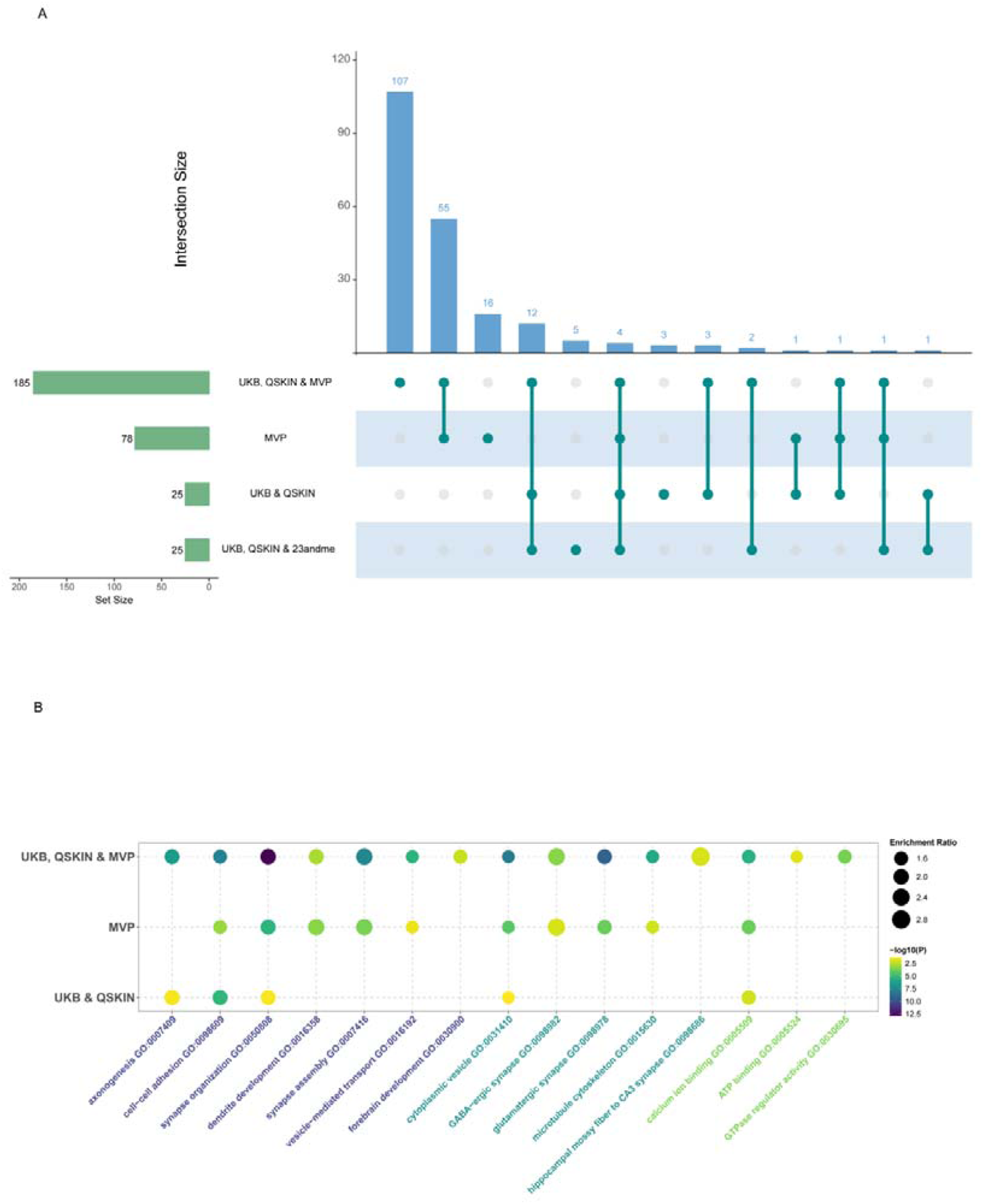
Overlap of significant loci and gene-set enrichment analysis across GERD GWAS datasets. **(A)** UpSet plot summarizing the intersections of independent genome-wide significant GERD loci identified across four GWAS datasets: UKB + QSKIN; UKB + QSKIN + 23andMe; MVP; and the UKB + QSKIN + MVP meta-analysis. Horizontal bars (left) indicate the total number of significant loci within each dataset. Vertical bars (top) represent the number of shared loci across dataset combinations, as indicated by filled dots in the matrix below. **(B)** Bubble plot showing GO biological processes and cellular components enriched among MAGMA-significant genes from three GERD GWAS datasets (UKB & QSKIN, MVP, and the combined UKB, QSKIN & MVP meta-analysis). Bubble size represents the enrichment ratio, and color denotes –log_10_(P) for each term.

To further characterize the biological architecture underlying GERD susceptibility, we performed MAGMA gene-level association and pathway enrichment analyses across the An et al. (UKB + QSKIN) (An et al. 2019), MVP (A et al. 2024), and combined meta-analysis datasets. As sample size and statistical power increased, enrichment patterns progressively converged from broad developmental processes toward increasingly specific neuronal and synaptic pathways. In the An et al. dataset (An et al. 2019), enrichment signals were relatively broad and included developmental and cellular processes such as axonogenesis, synapse organization and cell_–_ cell adhesion. In the MVP dataset(A et al. 2024), enrichment became more focused toward neuronal signaling pathways, including GABAergic and glutamatergic synapses. In the full meta-analysis, enrichment patterns converged further on discrete neurobiological programs related to synaptic organization, forebrain development and hippocampal mossy fiber_–_CA3 synaptic structure (Figure 1B), supporting a neurogenic component underlying GERD susceptibility.

### Genetic correlations and causal relationships between GERD and psychiatric disorders

Using LD score regression, we examined genome-wide genetic correlations between GERD and major psychiatric disorders(Table S4). As shown in Figure S2, GERD demonstrated strong positive genetic correlations with MDD, PTSD, and ADHD, with more moderate correlations observed for ANX and BIP. A weaker correlation was detected for autism, while SCZ showed no significant genetic overlap with GERD. To further evaluate directionality in these associations, we performed bidirectional MR analyses. As shown in Figure S3, genetically predicted GERD exerted significant causal effects on multiple psychiatric phenotypes, including MDD, PTSD, and ADHD. The causal estimates were substantially stronger for MDD and PTSD compared with ADHD, indicating a more pronounced influence of GERD on internalizing disorders. Conversely, the reverse MR analyses revealed that MDD, PTSD, ADHD, and ANX exerted significant causal effects on GERD. Interestingly, ADHD and ANX demonstrated stronger causal effects on GERD than MDD and PTSD, suggesting that neurodevelopmental and anxiety-related liability may exert a comparatively stronger influence on GERD risk than liability to MDD or PTSD.

Given the well-established epidemiological relationship between obesity and GERD, we additionally evaluated BMI as a representative trait. LDSC analysis revealed a significant positive genetic correlation between BMI and GERD, although the magnitude was weaker than those observed for MDD and PTSD (Figure S2). In contrast, bidirectional MR analyses between GERD and BMI did not identify significant causal effects in either direction (Figure S3).

### Latent neuropsychiatric architecture underlying GERD susceptibility

To further disentangle the shared genetic architecture linking GERD, psychiatric disorders, and metabolic liability, we performed Genomic SEM based on genome-wide genetic covariance estimated from LDSC (Figure 2). Correlation structure analysis identified three major latent dimensions broadly corresponding to internalizing disorders (F1), compulsive/neurodevelopmental disorders (F2), and psychotic disorders (F3) (Figure 2A-B; Table S5). GERD clustered most strongly within the internalizing dimension together with MDD, PTSD, ANX, and ADHD, whereas autism spectrum disorder (ASD) and obsessive-compulsive disorder (OCD) preferentially loaded onto the compulsive/neurodevelopmental factor, and bipolar disorder (BIP) and schizophrenia (SCZ) loaded primarily onto the psychotic factor. Notably, BMI also loaded onto the internalizing factor, consistent with its known epidemiological and genetic relationship with GERD.

**Figure 2.**
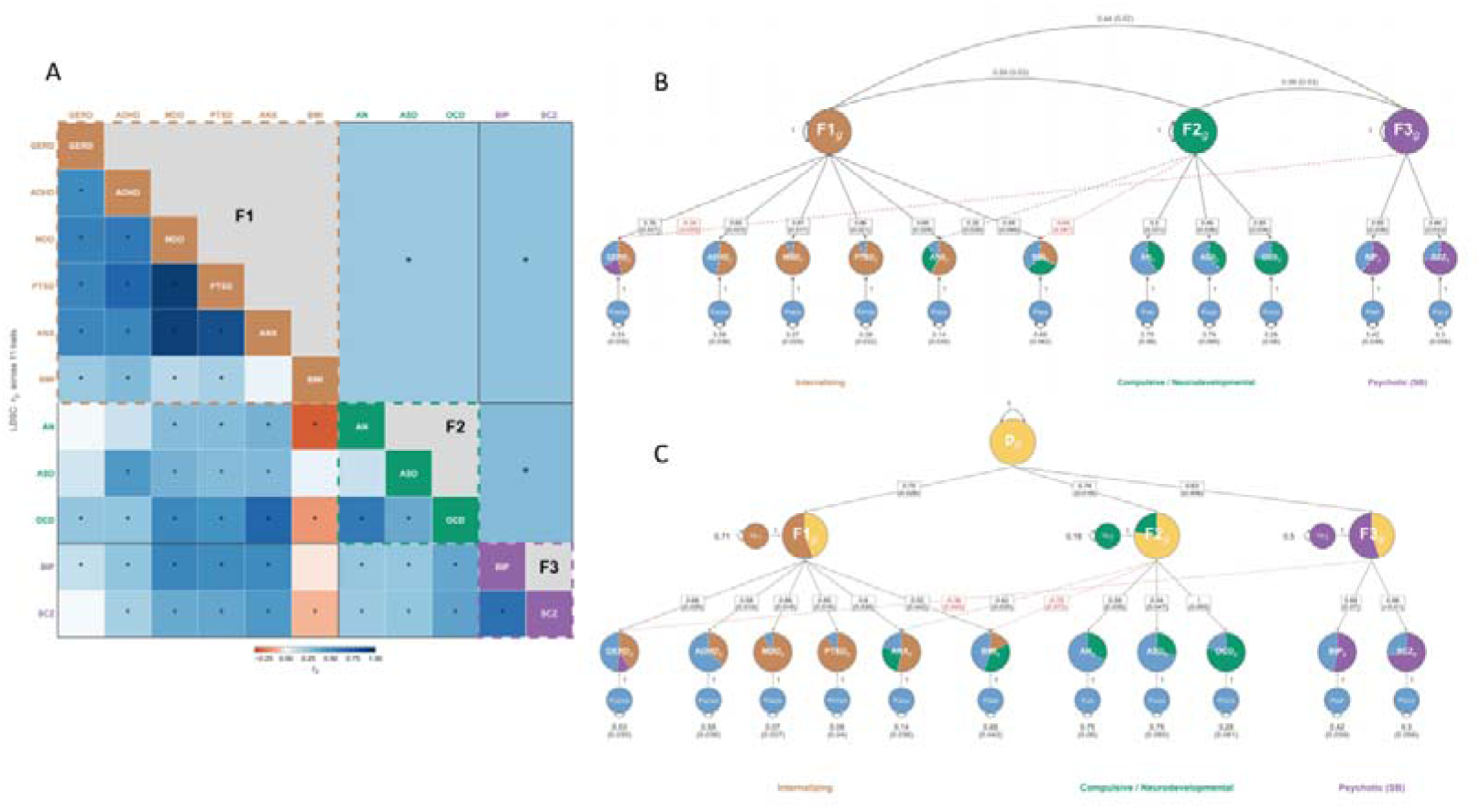
Latent neuropsychiatric architecture underlying GERD susceptibility. **(A)** Heatmap of pairwise r_g_ estimated by LDSC across 11 traits. Traits are grouped into three latent factors: F1 (Internalizing: GERD, ADHD, MDD, PTSD, ANX, BMI), F2 (Compulsive/Neurodevelopmental: AN, ASD, OCD), and F3 (Psychotic: BIP, SCZ). Dashed borders indicate factor groupings. Asterisks denote statistically significant correlations. **(B)**Correlated three-factor model from Genomic SEM. Each observed trait loads onto one of three latent factors (F1_g_, F2_g_, F3_g_), with standardized factor loadings and standard errors shown along paths. Inter-factor correlations are displayed at the top. **(C)** Higher-order factor model incorporating a general psychiatric factor (P_g_) above the three lower-order dimensions. Standardized loadings from P_g_ to each sub-factor and from sub-factors to observed traits are shown with standard errors. Residual variances for each trait are indicated at the bottom.

We next fitted a higher-order latent factor model incorporating a general psychiatric factor (Pg) above the three lower-order dimensions (Figure 2C). In this framework, GERD retained substantial loading on the internalizing factor even after accounting for shared covariance involving BMI and the broader psychiatric structure. Interestingly, GERD showed negative loading relative to the psychotic-spectrum factor, whereas BMI demonstrated a negative relationship with the compulsive/neurodevelopmental factor. These contrasting patterns suggest that GERD and BMI exhibit partially distinct latent architectures despite their positive genome-wide correlation. Overall, these findings indicate that the genetic relationship between GERD and psychiatric disorders is not solely attributable to obesity-mediated metabolic effects, but instead reflects a distinct neuropsychiatric component enriched within internalizing and stress-related liability.

### Local genetic correlation patterns between GERD and psychiatric disorders

We further performed local genetic correlation analysis using LAVA to identify specific genomic regions contributing to the shared architecture between GERD and psychiatric disorders (Table S6). Consistent with the genome-wide LDSC and Genomic SEM results, internalizing disorders showed widespread positive local genetic correlations with GERD across multiple genomic regions (Figure 3). Most significant loci demonstrated concordant positive effects, which support the presence of a latent internalizing dimension (F1) underlying the genetic relationship between GERD and psychiatric disorders.

**Figure 3.**
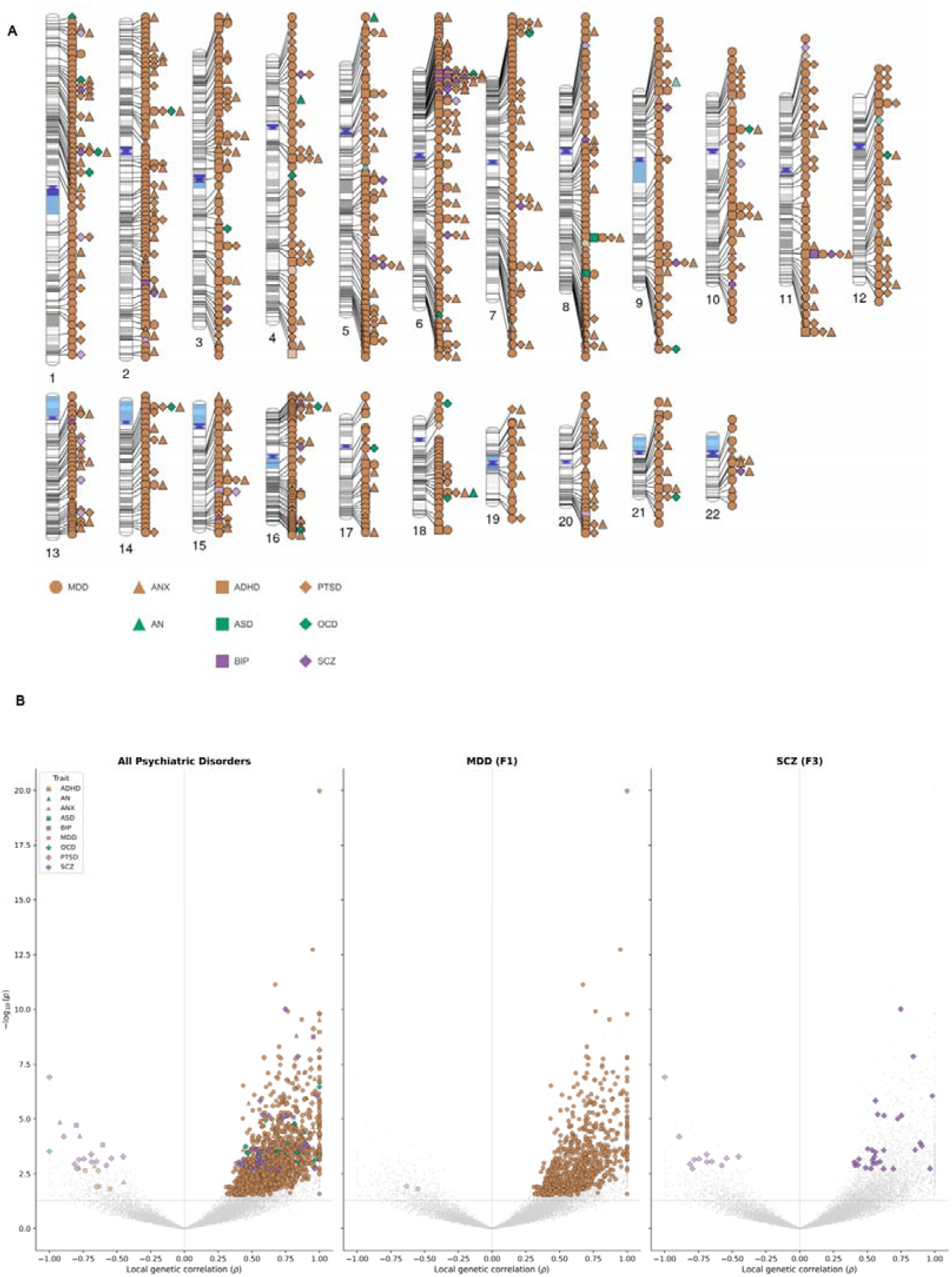
Local genetic correlation patterns between GERD and psychiatric disorders. **(A)** Genome-wide ideogram displaying significant local genetic correlation loci between GERD and nine psychiatric disorders identified by LAVA. Each chromosome (1–22) is shown with cytogenetic banding. Significant loci are marked by trait-specific symbols positioned along chromosomes, with shape and color denoting the corresponding psychiatric trait (see legend). **(B)** Volcano-style scatter plots of local genetic correlations (ρ) versus –log_10_(P) for GERD paired with psychiatric disorders. The left panel shows all psychiatric disorders combined; the middle panel highlights MDD (representative of the internalizing factor F1); the right panel highlights SCZ (representative of the psychotic factor F3). Each point represents a single genomic locus, with colored symbols indicating trait identity and gray points denoting non-significant results. The dashed horizontal line indicates the significance threshold.

In contrast, negative local genetic correlations were comparatively less frequent but were enriched within psychotic-spectrum disorders, particularly SCZ. As SCZ showed the strongest loading on F3 in the Genomic SEM framework, we observed that a substantial subset of GERD_–_SCZ local correlations exhibited opposite effect directions across the genome. This pattern is consistent with the Genomic SEM results, in which GERD showed negative loading relative to F3, suggesting that specific genomic regions may exert antagonistic effects on GERD susceptibility and psychotic liability.

### Shared pleiotropic loci between GERD and psychiatric disorders

Given the strong global and local genetic overlap between GERD and the psychiatric traits contributing to the F1 factor, we next performed cross-trait condFDR analyses to enhance GERD locus discovery. Through leveraging the polygenic enrichment between GERD and its most strongly associated psychiatric disorders, we identified a total of 352 novel GERD risk signals (Figure 4 and Table S7). Independent GWAS signals were defined using PLINK’s clumping procedure, applying an r² threshold of 0.1 to account for linkage disequilibrium. Notably, the cross-trait analysis revealed additional members of the same functional families implicated in our primary meta-analysis. For example, the meta-analysis detected immune and neurogenic loci such as *TLR4*, *GRIK3*, and *DRD3*, while the condFDR approach identified their corresponding family members *TLR9*, *GRIK2*, and *DRD2*, indicating convergent biology across related receptor systems. We also detected *GRM5*, complementing prior evidence for *GRM8* reported by An et al. (An et al. 2019) and further supporting a role for glutamatergic signaling in GERD susceptibility. The cross-trait analysis also uncovered additional GERD-associated genes involved in neuro-sensory regulation and synaptic signaling, including *PCLO*, a presynaptic scaffolding protein essential for synaptic vesicle cycling and *ROBO2*, a key axon-guidance receptor implicated in visceral sensory circuit organization; *TSNARE1*, involved in synaptic vesicle fusion and neurotransmitter release. In parallel, we performed GenomicSEM analysis integrating GERD with the psychiatric traits contributing to the F1 factor, which identified 110 additional genome-wide significant loci (Table S8). Notably, the majority of loci identified by GenomicSEM were also detected by condFDR, supporting the robustness and reproducibility of the shared genetic architecture captured by these complementary cross-trait approaches.

**Figure 4.**
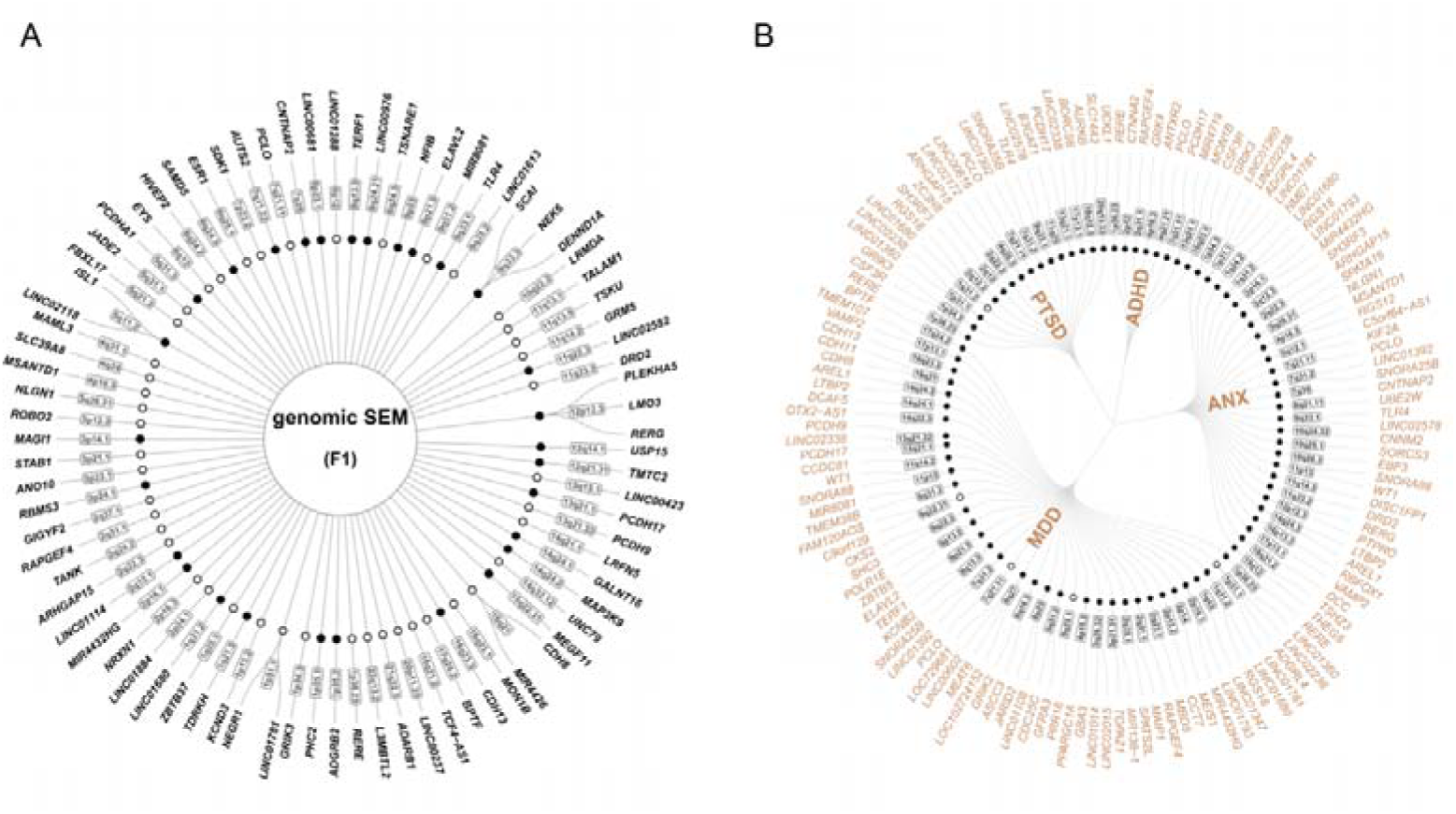
Novel pleiotropic loci shared between GERD and internalizing psychiatric disorders. **(A)** Circular dendrogram of novel loci identified through Genomic SEM common factor GWAS of the internalizing factor (F1). Loci are organized by cytogenetic band (inner ring) with mapped genes in the outer ring. At the cytoBand level, filled circles denote loci not previously reported in any GERD GWAS (new), and open circles denote loci previously reported in psychiatric disorder GWAS but newly identified as GERD-associated in the present study (new*). **(B)** Circular dendrogram of novel loci identified through conjunctional FDR analysis (conjFDR < 0.05 and condFDR < 0.05) between GERD and each of four F1-loading psychiatric traits (ADHD, ANX, MDD, PTSD). Loci already identified by the primary meta-analysis or the Genomic SEM analysis in panel A were excluded. Only loci with GERD P < 1 × 10_10_ are displayed; the complete results are provided in Table S7. Loci are organized hierarchically by conditioning trait, cytogenetic band, and mapped gene. Symbol conventions for new and new* are the same as in panel A.

### Mapping the cellular basis of GERD neuropsychiatric susceptibility in the human brain

To investigate the neurobiological basis of GERD susceptibility, we performed TDEP analysis using a large-scale human brain single-cell transcriptomic atlas (Figure S4). GERD-associated genetic signals showed predominant enrichment within neuronal populations of the cerebral cortex, particularly the primary motor cortex (M1C), middle temporal gyrus (MTG), anterior cingulate cortex (ACC), frontal insular-related regions (FI), and medial entorhinal cortex (MEC). Significant enrichment was also observed across limbic structures, including the amygdala, hippocampal CA1_–_CA4 and dentate gyrus regions, entorhinal cortex, and hypothalamic-associated neuronal populations. Many enriched cell populations consisted of both glutamatergic excitatory neurons and GABAergic interneuron subtypes, including MGE and CGE interneurons as well as LAMP5-LHX6 chandelier cells(Table S9).

To further characterize the neuropsychiatric component underlying F1, we restricted the analysis to genomic regions shared between GERD and the major F1-contributing psychiatric disorders and repeated TDEP analysis. As expected, the number of significant cell clusters was substantially reduced after restricting the analysis to shared loci, reflecting the smaller subset of genomic signals specifically contributing to the GERD_–_ internalizing overlap. Nevertheless, significant enrichment remained concentrated within cortical and limbic regions, particularly ACC, MTG, anterior olfactory nucleus (AON), and hypothalamic mammillary nucleus (MN) (Figure 5; Table S9). The cortical signals were primarily driven by deep-layer corticothalamic and near-projecting neuronal populations, whereas the hypothalamic MN-associated signal was notably enriched in astrocyte populations rather than neurons. Given the central role of the hypothalamus and mammillary region in autonomic and stress-related regulation, this finding suggests that glial-mediated neuroendocrine or autonomic mechanisms may contribute to the shared GERD_–_internalizing genetic architecture. In contrast, although Genomic SEM suggested a weak negative contribution of F3 to GERD susceptibility, TDEP analysis based on F3-associated regions did not identify significant brain-region enrichment signals.

**Figure 5.**
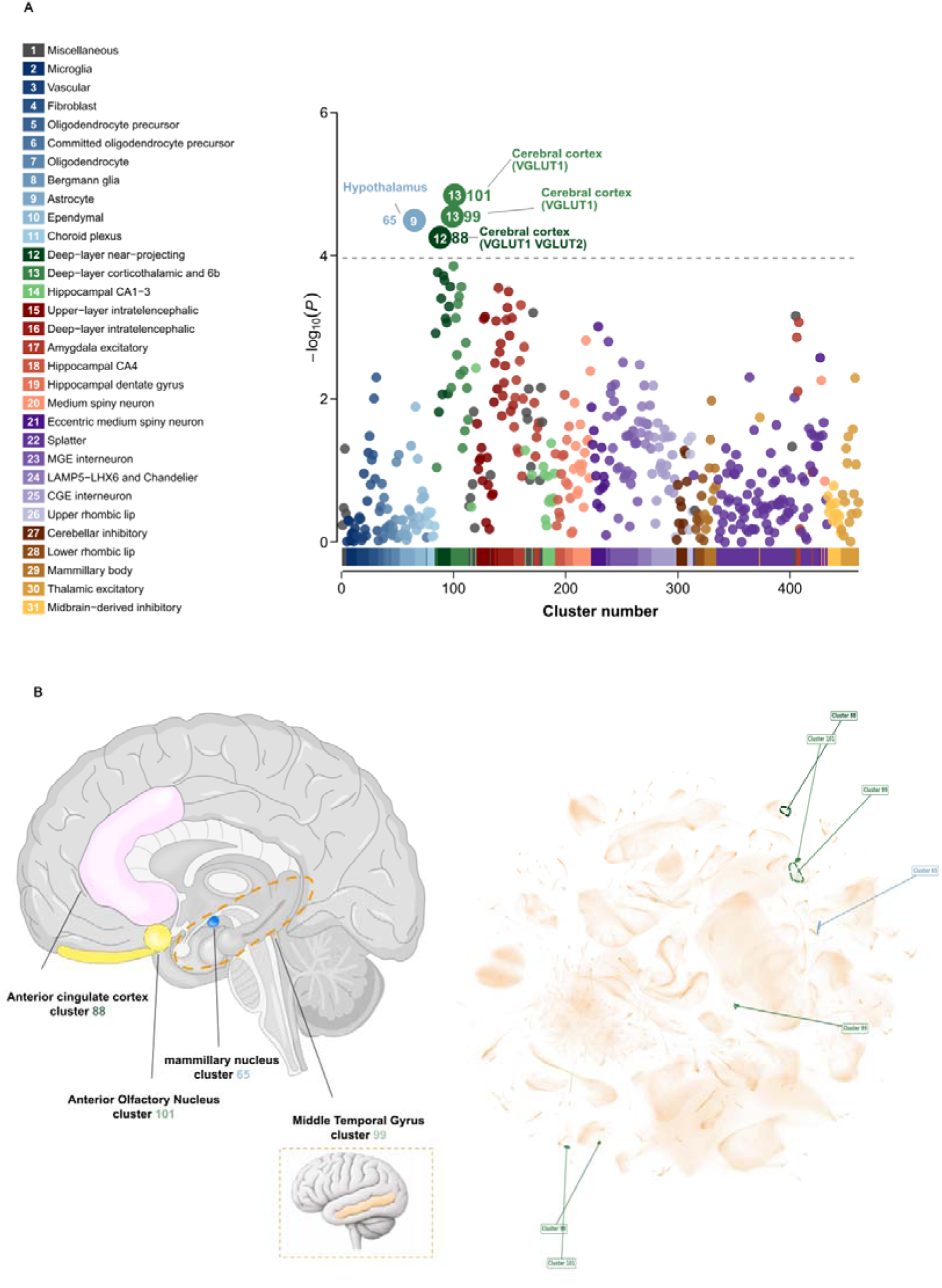
Cell-type enrichment of GERD–internalizing shared genetic signals in the human brain. **(A)** TDEP analysis of GERD-associated genetic signals restricted to genomic regions shared with F1-loading internalizing psychiatric disorders. Each point represents a single cell cluster from a large-scale human brain single-cell transcriptomic atlas, plotted by cluster number (x-axis) and –log_10_ (P) (y-axis). Points are colored according to 31 supercluster cell-type categories (left legend). The dashed horizontal line indicates the Bonferroni-corrected significance threshold. Significant clusters are labeled with their cluster number and annotated with cell-type identity and brain region of origin. **(B)** Anatomical localization of significantly enriched cell clusters. Left: sagittal brain schematic illustrating the anatomical positions of four significant clusters, including the anterior cingulate cortex (cluster 88), middle temporal gyrus (cluster 99), anterior olfactory nucleus (cluster 101), and mammillary nucleus (cluster 65). Right: UMAP embedding of the single-cell atlas with the positions of the four significant clusters highlighted. **Abbreviations:** UMAP, Uniform Manifold Approximation and Projection

## Discussion

In this study, we integrated large-scale statistical genetics, cross-trait genomic analyses, and single-cell transcriptomic mapping to investigate the biological architecture underlying GERD susceptibility. Across multiple analytical levels, we observed convergent evidence supporting the presence of an intrinsic neuropsychiatric component underlying GERD that preferentially overlaps with internalizing psychiatric liability and localizes to interoceptive, limbic, and autonomic brain systems. Importantly, although GERD also demonstrated genetic overlap with BMI, our findings indicate that the relationship between GERD and psychiatric disorders cannot be explained solely by obesity-related metabolic liability. Together, these results support a model in which GERD susceptibility emerges not only from peripheral gastrointestinal dysfunction, but also from centrally mediated neurobiological processes involved in visceral sensation, stress responsivity, and autonomic regulation.

The relationship between GERD and psychiatric disorders has long been recognized epidemiologically(Li et al. 2024), yet the underlying biological interpretation has remained unclear. Here, we found that GERD exhibited strongest genome-wide genetic correlations with internalizing psychiatric disorders, particularly MDD and PTSD, while the genetic overlap with BMI was comparatively weaker. Moreover, bidirectional MR analyses identified significant causal relationships between GERD and multiple psychiatric phenotypes, whereas no significant bidirectional causal effects were observed between GERD and BMI. These findings suggest that psychiatric liability represents a biologically meaningful component of GERD susceptibility rather than merely a secondary consequence of metabolic dysfunction.

Our Genomic SEM analyses further refined this relationship by demonstrating that GERD clustered preferentially within a latent internalizing factor composed primarily of MDD, PTSD, ANX, and ADHD. Notably, GERD retained substantial loading on this factor even after accounting for BMI and broader shared psychiatric covariance, indicating that the GERD–psychiatric relationship is partially independent of obesity-related genetic liability. In contrast, psychotic-spectrum disorders formed a distinct latent dimension that demonstrated weak or negative relationships with GERD. Consistent with this framework, local genetic correlation analyses revealed widespread positive local correlations between GERD and internalizing disorders, whereas negative local correlations were enriched primarily within schizophrenia-associated regions.

At the cellular level, TDEP analyses revealed that GERD-associated genetic signals preferentially localized to cortical and limbic systems involved in interoception, visceral sensory processing, stress responsivity, and autonomic regulation. The strongest enrichments involved neuronal populations within the anterior cingulate cortex, frontal insular-related cortex, middle temporal gyrus, entorhinal cortex, hippocampus, amygdala, and hypothalamic-associated regions, supporting convergence of GERD susceptibility on core brain–gut axis circuitry. Notably, when the analysis was restricted to loci shared specifically between GERD and F1, the number of significant cell clusters decreased substantially, yet significant enrichment persisted within the ACC, MTG, anterior olfactory nucleus, and hypothalamic mammillary nucleus. Intriguingly, the mammillary nucleus signal was driven predominantly by astrocyte populations rather than neurons, raising the possibility that glial-mediated autonomic or neuroendocrine regulation may contribute to the shared GERD– internalizing genetic architecture. Together, these findings suggest that GERD susceptibility may involve altered central processing of visceral sensory and stress-related signals rather than reflecting solely peripheral reflux physiology.

In conclusion, our study provides convergent genetic and cellular evidence that GERD susceptibility contains a distinct neuropsychiatric component preferentially linked to internalizing liability and interoceptive–limbic brain systems. These findings support a revised conceptual framework in which GERD is shaped by coordinated interactions among peripheral gastrointestinal physiology, metabolic regulation, and centrally mediated brain–gut neurobiology. More broadly, our results highlight the importance of neural and affective mechanisms in shaping susceptibility to common gastrointestinal disorders and provide a genome-to-cell framework for understanding the neurobiological basis of GERD.

## Data Availability

This study used only openly available, de-identified, summary-level human data that were publicly accessible before initiation of the study. GERD GWAS summary statistics for UK Biobank and QSKIN, excluding 23andMe data, were obtained from Figshare: https://doi.org/10.6084/m9.figshare.8986589. GERD summary statistics from the Million Veteran Program were obtained from the publicly accessible MVP dbGaP summary-results study phs002453.v1.p1: https://www.ncbi.nlm.nih.gov/projects/gap/cgi-bin/study.cgi?study_id=phs002453.v1.p1 and https://phenomics.va.ornl.gov/web/. Psychiatric-disorder GWAS summary statistics were obtained from the following publicly accessible Figshare datasets: ADHD, https://doi.org/10.6084/m9.figshare.22564390.v1; anorexia nervosa, https://doi.org/10.6084/m9.figshare.14671980.v1; anxiety disorder, https://doi.org/10.6084/m9.figshare.16602218.v1; autism spectrum disorder, https://doi.org/10.6084/m9.figshare.14671989.v1; bipolar disorder, https://doi.org/10.6084/m9.figshare.27216117.v2; major depressive disorder, https://doi.org/10.6084/m9.figshare.27061255.v4; obsessive-compulsive disorder, https://doi.org/10.6084/m9.figshare.28707155.v4; post-traumatic stress disorder, https://doi.org/10.6084/m9.figshare.26349322.v3; and schizophrenia, https://doi.org/10.6084/m9.figshare.19426775.v7. BMI GWAS summary statistics were obtained from the NHGRI-EBI GWAS Catalog, accession GCST90446645: https://www.ebi.ac.uk/gwas/studies/GCST90446645. Publicly available human brain single-cell transcriptomic data were obtained from the adult human brain atlas of Siletti et al.: https://brain-map.org/our-research/cell-type-taxonomies/whole-human-brain-taxonomy. No individual-level, identifiable, controlled-access, or application-restricted data were accessed.

## Acknowledgements

We thank the investigators and participants of the GERD, psychiatric disorder, and BMI GWAS studies, including the UK Biobank, QSKIN, Million Veteran Program, and Psychiatric Genomics Consortium, for making their summary statistics publicly available. We also thank Siletti et al. for providing the human brain single-cell transcriptomic atlas.

## Author contributions

X.W. and Q.Z. contributed equally to this work and wrote the main manuscript text. Y.S, X.C. and H.Z. conceived and supervised the study and wrote the manuscript. Q.W., M.Z. and Z.L. developed and implemented the core methodology. F.H., Y.Z., H.Y. and K.U. contributed to data preprocessing, annotation, and quality control. N.Z. assisted with interpretation of results and literature review. All authors reviewed the manuscript.

## Data availability

All GWAS summary statistics analyzed in this study are publicly available from their respective consortia and original publications. Detailed sources, accession identifiers, and download links for each dataset (including the GERD, psychiatric disorder, BMI, and single-cell transcriptomic resources) are provided in the Methods section and Supplementary Table S1. No individual-level data were used in this study.

## Code availability

The statistical analyses were performed using LDSC (https://github.com/bulik/ldsc), MAGMA v1.0.6 (https://ctg.cncr.nl/software/magma), the TwoSampleMR R package v0.6.4, LAVA (https://github.com/josefin-werme/LAVA), Genomic SEM (https://github.com/GenomicSEM/GenomicSEM), the condFDR/conjFDR framework (https://github.com/precimed/pleiofdr), TDEP (https://github.com/TissueEnrich/TDEP), PLINK v1.9 (https://www.cog-genomics.org/plink/1.9/), and g:Profiler (https://biit.cs.ut.ee/gprofiler/). All software versions and parameters are described in the Methods section.

## References

A V, Je H, A R, et al (2024) Diversity and scale: Genetic architecture of 2068 traits in the VA Million Veteran Program. PubMed

An J, Gharahkhani P, Law MH, et al (2019) Gastroesophageal reflux GWAS identifies risk loci that also associate with subsequent severe esophageal diseases. Nature Communications 10:4219. 10.1038/s41467-019-11968-2

Andreassen OA, Thompson WK, Dale AM (2014) Boosting the power of schizophrenia genetics by leveraging new statistical tools. Schizophr Bull 40:13–17. 10.1093/schbul/sbt168

Azer SA, Goosenberg E (2025) Gastroesophageal Reflux Disease (GERD). In: StatPearls. StatPearls Publishing, Treasure Island (FL)

Bentley B, Chanaa F, Cecil A, Clayton S (2024) The impact of gastroesophageal reflux disease on upper esophageal sphincter function: Insights from PH impedance and high-resolution manometry. Physiol Rep 12:e70011. 10.14814/phy2.70011

Bulik-Sullivan BK, Loh P-R, Finucane HK, et al (2015) LD Score regression distinguishes confounding from polygenicity in genome-wide association studies. Nat Genet 47:291–295. 10.1038/ng.3211

Caldart F, Gabriel C, Vauquelin B, et al (2025) Overlap of Esophageal Disorders of Gut-Brain Interactions and Gastroesophageal Reflux Disease Is Highly Prevalent in Patients With Refractory Reflux Symptoms. Am J Gastroenterol 120:1770–1778. 10.14309/ajg.0000000000003542

Chen S, Chen Z, Jiang X, et al (2024) Modifiable risk factors mediate the effect of gastroesophageal reflux disease on stroke and subtypes: A Mendelian randomization study. J Stroke Cerebrovasc Dis 33:107612. 10.1016/j.jstrokecerebrovasdis.2024.107612

Clevenger MH, Karami AL, Carlson DA, et al (2023) Suprabasal cells retain progenitor cell identity programs in eosinophilic esophagitis-driven basal cell hyperplasia. JCI Insight 8:. 10.1172/jci.insight.171765

de Leeuw CA, Mooij JM, Heskes T, Posthuma D (2015) MAGMA: generalized gene-set analysis of GWAS data. PLoS Comput Biol 11:e1004219. 10.1371/journal.pcbi.1004219

Ding H, Jiang Y, Sun Q, et al (2025) Integrating genetics and transcriptomics to characterize shared mechanisms in digestive diseases and psychiatric disorders. Commun Biol 8:47. 10.1038/s42003-025-07481-6

Duncan LE, Li T, Salem M, et al (2025) Mapping the cellular etiology of schizophrenia and complex brain phenotypes. Nat Neurosci 28:248–258. 10.1038/s41593-024-01834-w

Frei E, Filiz TT, Frei O, et al (2025) Genome-wide analysis of screen behaviors among adolescents identifies novel loci and overlap with educational attainment and mental disorders. Sci Rep 15:34420. 10.1038/s41598-025-17450-y

Grotzinger AD, Rhemtulla M, de Vlaming R, et al (2019) Genomic structural equation modelling provides insights into the multivariate genetic architecture of complex traits. Nature Human Behaviour 3:513–525. 10.1038/s41562-019-0566-x

Guan Y, Cheng H, Zhang N, et al (2025) The role of the esophageal and intestinal microbiome in gastroesophageal reflux disease: past, present, and future. Front Immunol 16:1558414. 10.3389/fimmu.2025.1558414

He X, Ma Q, Liu J, et al (2025) Investigating the shared genetic architecture between schizophrenia and sex hormone traits. Transl Psychiatry 15:83. 10.1038/s41398-025-03305-7

Hemani G, Tilling K, Davey Smith G (2017) Orienting the causal relationship between imprecisely measured traits using GWAS summary data. PLoS Genet 13:e1007081. 10.1371/journal.pgen.1007081

Hemani G, Zheng J, Elsworth B, et al (2018) The MR-Base platform supports systematic causal inference across the human phenome. Elife 7:. 10.7554/eLife.34408

Holtmann G, Moniruzzaman M, Shah A (2025) Decoding the Gut-Brain Axis: A Journey toward Targeted Interventions for Disorders-of-Gut-Brain Interaction. Dig Dis 43:257–265. 10.1159/000543845

Kolberg L, Raudvere U, Kuzmin I, et al (2023) g:Profiler-interoperable web service for functional enrichment analysis and gene identifier mapping (2023 update). Nucleic Acids Res 51:W207–W212. 10.1093/nar/gkad347

Li Q, Duan H, Wang Q, et al (2024) Analyzing the correlation between gastroesophageal reflux disease and anxiety and depression based on ordered logistic regression. Sci Rep 14:6594. 10.1038/s41598-024-57101-2

Liu T, Mei Z (2025) Anti-reflux mucosectomy superior to argon plasma coagulation in managing Barrett’s esophagus by improving lower esophageal sphincter pressure and cardiac sphincter valve function: a single-center, retrospective study. Therap Adv Gastroenterol 18:17562848251331787. 10.1177/17562848251331787

Lu W, He X, Peng H, et al (2025) Exploring the shared genetic architecture between testosterone traits and major depressive disorder. BMC Psychiatry 25:651. 10.1186/s12888-025-07096-5

Ong J-S, An J, Han X, et al (2022) Multitrait genetic association analysis identifies 50 new risk loci for gastro-oesophageal reflux, seven new loci for Barrett’s oesophagus and provides insights into clinical heterogeneity in reflux diagnosis. Gut 71:1053–1061. 10.1136/gutjnl-2020-323906

Paulrasu K, Caspa Gokulan R, El-Rifai W, et al (2025) Chronic Gastroesophageal Reflux Dysregulates Proteostasis in Esophageal Epithelial Cells. Cell Mol Gastroenterol Hepatol 19:101434. 10.1016/j.jcmgh.2024.101434

Pavić I, Topalušić I, Močić Pavić A, et al (2025) Linking Gastroesophageal Reflux Characteristics to Airway Inflammation: Insights from Bronchoalveolar Lavage Cytology in Severe Preschool Wheeze. Life (Basel) 15:. 10.3390/life15101561

Savarino V, Visaggi P, Marabotto E, et al (2025) Topical Protection of Esophageal Mucosa as a New Treatment of GERD. J Clin Gastroenterol 59:197–205. 10.1097/MCG.0000000000002128

Sidorenko J, Couvy-Duchesne B, Kemper KE, et al (2024) Genetic architecture reconciles linkage and association studies of complex traits. Nat Genet 56:2352–2360. 10.1038/s41588-024-01940-2

Siletti K, Hodge R, Mossi Albiach A, et al (2023) Transcriptomic diversity of cell types across the adult human brain. Science 382:eadd7046. 10.1126/science.add7046

Song Y, Li L, Jiang Y, et al (2025) Multitrait Genetic Analysis Identifies Novel Pleiotropic Loci for Depression and Schizophrenia in East Asians. Schizophr Bull 51:684–695. 10.1093/schbul/sbae145

Sullivan PF, Agrawal A, Bulik CM, et al (2018) Psychiatric Genomics: An Update and an Agenda. Am J Psychiatry 175:15–27. 10.1176/appi.ajp.2017.17030283

Wei Y, Meng Z, Wang X, et al (2026) Transformer-based InsightGWAS improves GERD genetic discovery via pretraining on GWAS for major depressive disorder. Commun Biol 9:2. 10.1038/s42003-025-09177-3

Werme J, van der Sluis S, Posthuma D, de Leeuw CA (2022) An integrated framework for local genetic correlation analysis. Nat Genet 54:274–282. 10.1038/s41588-022-01017-y

Xu J, Du X, Zhai Y, et al (2025) Shared neuroimaging and molecular profiles in type 2 diabetes mellitus and major depressive disorder: an integrative analysis of genetic, transcriptomic, and neuroimaging data. Transl Psychiatry 15:352. 10.1038/s41398-025-03585-z

Yao S, Harder A, Darki F, et al (2025) Connecting genomic results for psychiatric disorders to human brain cell types and regions reveals convergence with functional connectivity. Nat Commun 16:395. 10.1038/s41467-024-55611-1

Zheng J, Tao L (2025) Multidimensional mechanisms and therapies underlying gastroesophageal reflux disease: focus on immunity, signaling pathways, and the microbiota-gut-brain axis. Front Immunol 16:1629944. 10.3389/fimmu.2025.1629944

Zhi L, Zheng Q, Jiang Y, et al (2025) Multi-trait genetic analysis of asthma and eosinophils uncovers pleiotropic loci in East Asians. Nature Communications 16:5081. 10.1038/s41467-025-60405-0

