## Supplementary Figures for "Dissecting shared genetic susceptibility between gastroesophageal reflux disease and psychiatric disorders beyond body mass index"

**Figure S1. Parallel analysis scree plot for determining the number of latent factors in Genomic SEM.** Observed eigenvalues (open triangles) and simulated 95th percentile eigenvalues (filled triangles) from parallel analysis of the LDSC-derived genetic covariance matrix across 11 traits. The observed eigenvalues exceeded the simulated threshold for the first three components, supporting a three-factor solution.

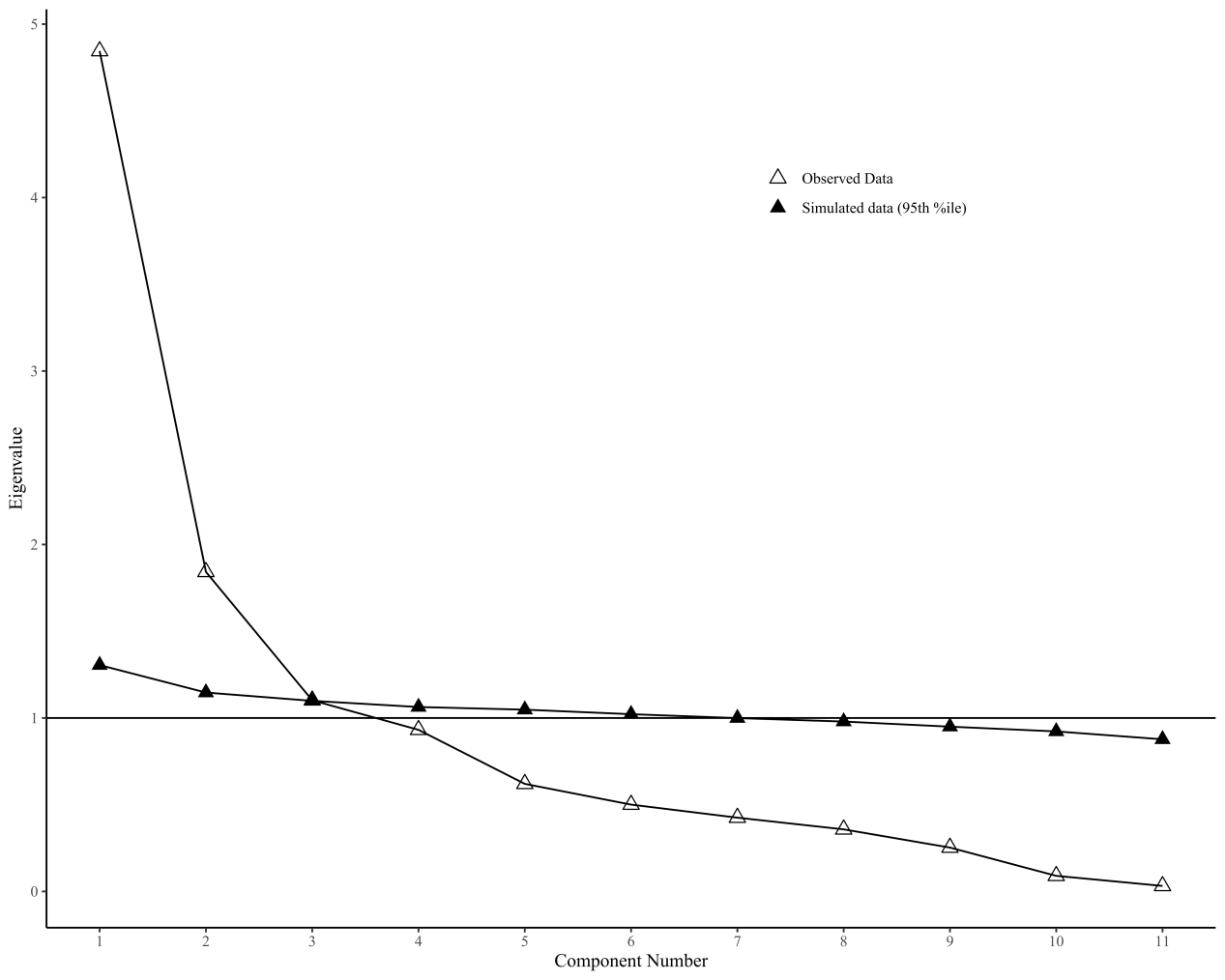

**Figure S2. Genome-wide genetic correlations between GERD and psychiatric disorders and BMI.** Forest plot of r_g_ estimated by LDSC between GERD and ten traits. Points represent r_g_ estimates; horizontal bars indicate 95% confidence intervals. The red dashed line indicates r_g_ = 0.

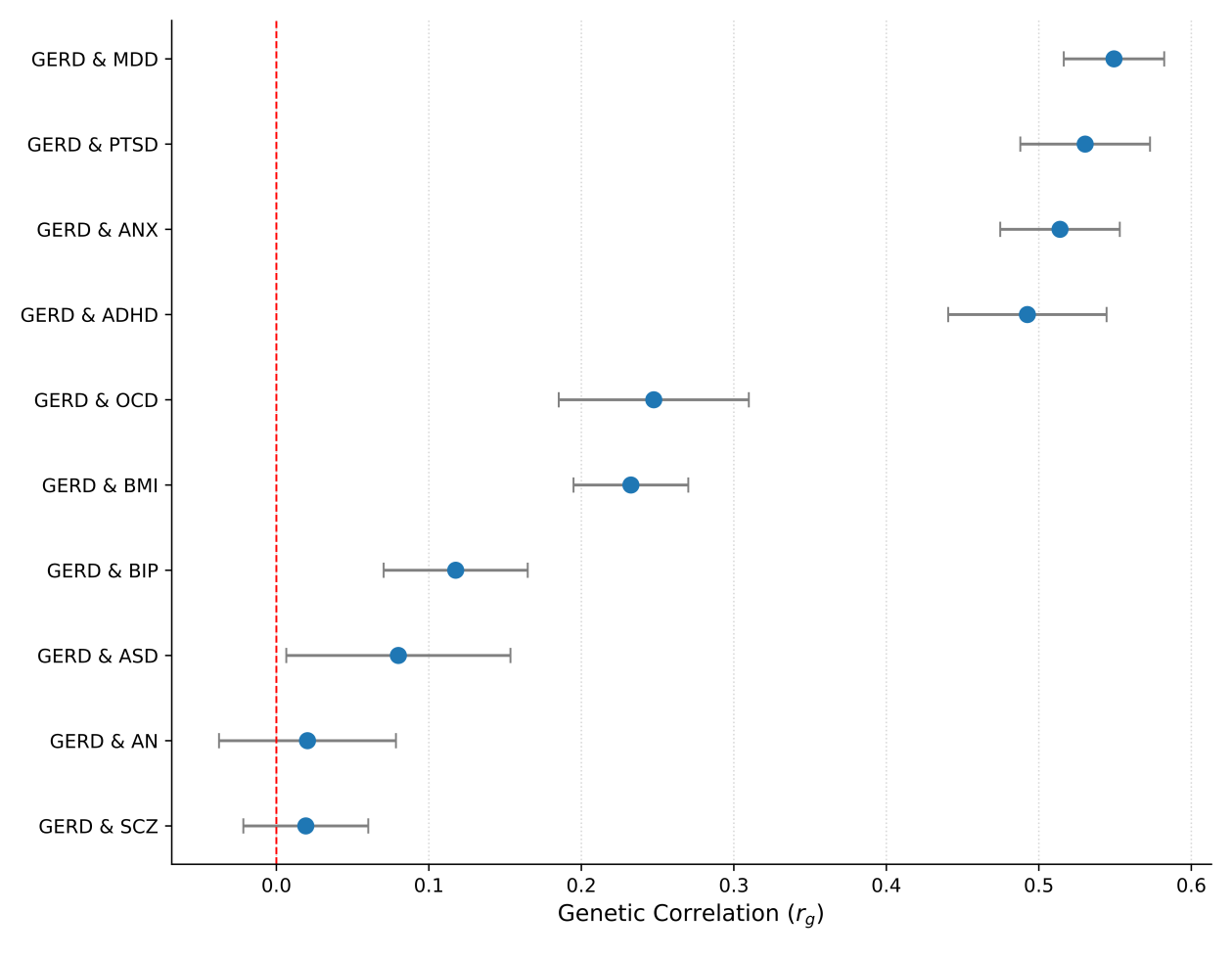

**Figure S3. Bidirectional Mendelian randomization between GERD and psychiatric disorders and BMI.** (A) MR estimates for the causal effect of GERD (exposure) on each psychiatric disorder and BMI (outcome). (B) MR estimates for the causal effect of each psychiatric disorder and BMI (exposure) on GERD (outcome). Odds ratios (OR) with 95% confidence intervals were estimated using the inverse variance weighted method. P-values are shown to the right of each estimate.

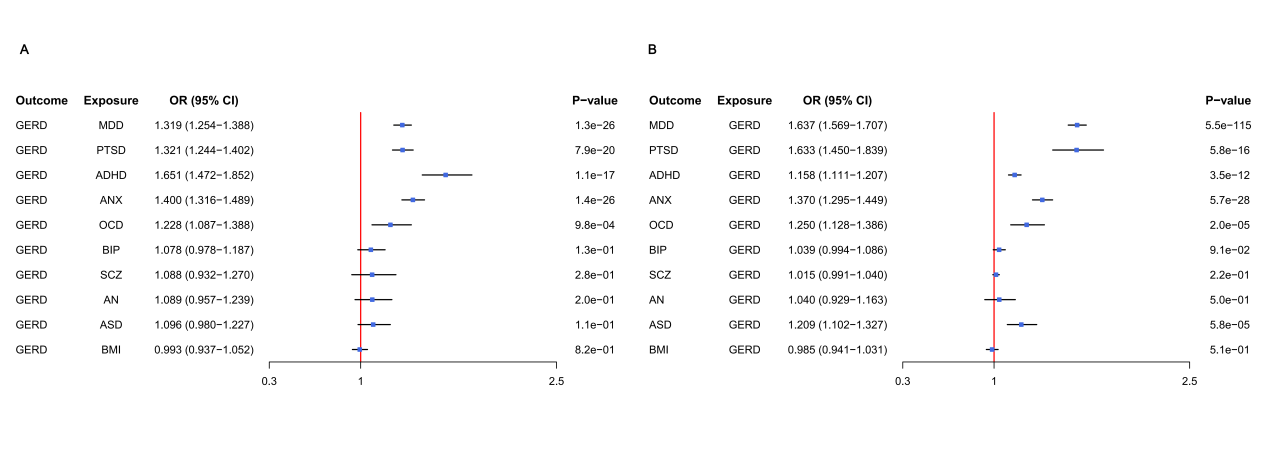

**Figure S4. Genome-wide TDEP cell-type enrichment analysis of GERD-associated genetic signals.** (A) TDEP analysis using GERD GWAS summary statistics and a large-scale human brain single-cell transcriptomic atlas. Each point represents a single cell cluster, plotted by cluster number (x-axis) and –log₁₀(P) (y-axis). Points are colored according to 31 supercluster cell-type categories (left legend). The dashed horizontal line indicates the Bonferroni-corrected significance threshold. (B) Anatomical localization of significantly enriched cell clusters, highlighting enrichment in the cerebral cortex, hippocampus, amygdala, basal forebrain, and cerebellum.

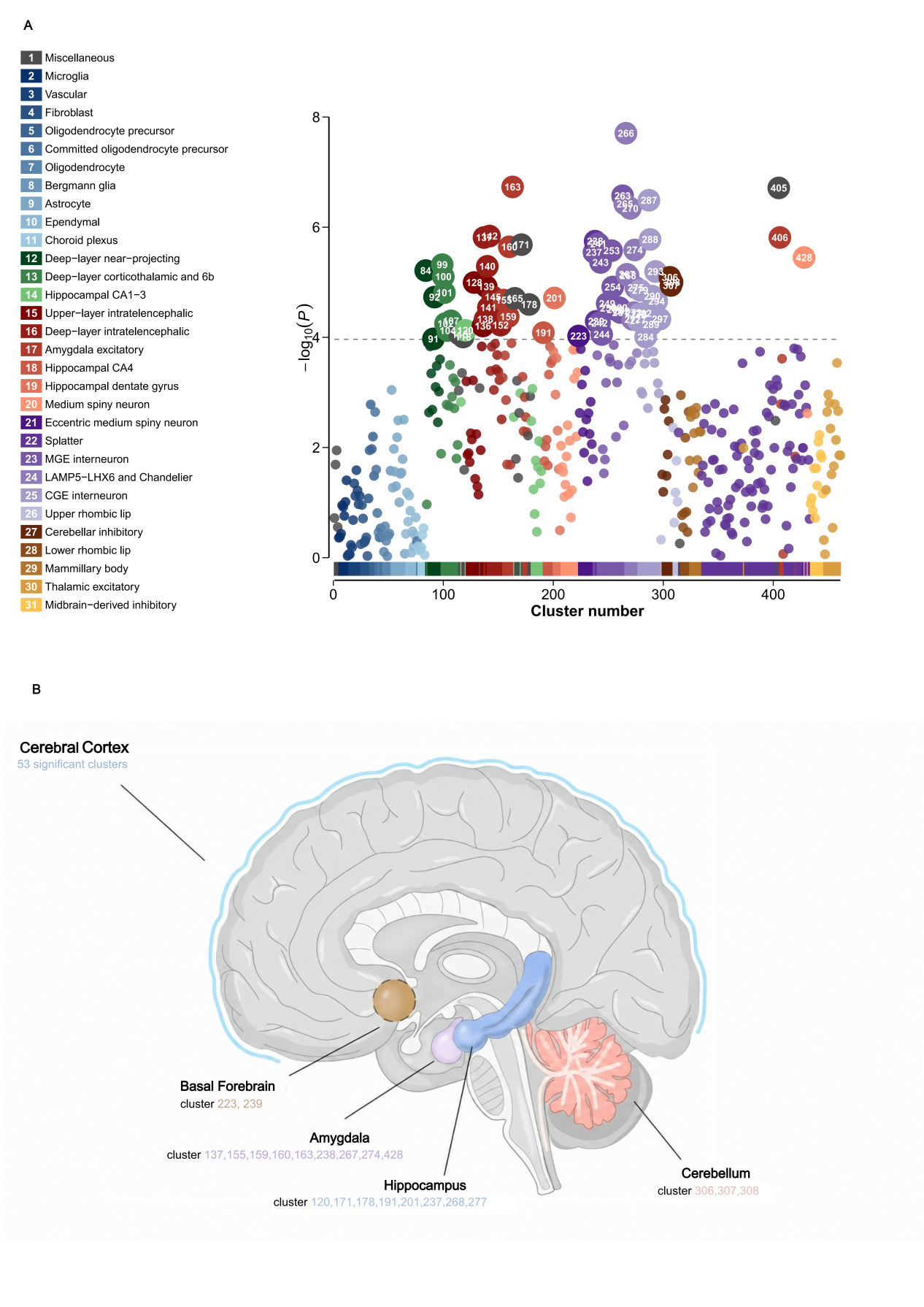
