## Supplementary Tables for "Dissecting shared genetic susceptibility between gastroesophageal reflux disease and psychiatric disorders beyond body mass index"

**Table S1.** Summary statistics for traits used in this study.

| Phenotype | Data source | Total sample size | Ncase | Ncontrol |
| --- | --- | --- | --- | --- |
| Gastroesophageal reflux disease | An J, Gharahkhani P, Law MH, et al. Gastroesophageal reflux GWAS identifies risk loci that also associate with subsequent severe esophageal diseases. Nat Commun. 2019;10(1):4219. | 332,601 | 71,522 | 261,079 |
| Gastroesophageal reflux disease | Verma A, Huffman JE, Rodriguez A, et al. Diversity and scale: Genetic architecture of 2068 traits in the VA Million Veteran Program. Science. 2024;385(6706):eadj1182. | 412,728 | 175,991 | 236,737 |
| Attention Deficit Hyperactivity Disorder | Sullivan, Patrick (2023). adhd2022. figshare. Dataset. https://doi.org/10.6084/m9.figshare.22564390.v1 | 225,534 | 38,691 | 186,843 |
| Anorexia Nervosa | Sullivan, Patrick (2021). an2019. figshare. Dataset. https://doi.org/10.6084/m9.figshare.14671980.v1 | 72,517 | 16,992 | 55,525 |
| Anxiety Disorder | Sullivan, Patrick (2021). panic2019. figshare. Dataset. https://doi.org/10.6084/m9.figshare.16602218.v1 | 9,907 | 2,147 | 7,760 |
| Autism Spectrum Disorder | Sullivan, Patrick (2021). asd2019. figshare. Dataset. https://doi.org/10.6084/m9.figshare.14671989.v1 | 46,350 | 18,381 | 27,969 |
| Bipolar Disorder | Sullivan, Patrick (2024). bip2024. figshare. Dataset. https://doi.org/10.6084/m9.figshare.27216117.v2 | 840,309 | 59,287 | 781,022 |
| Body mass index | Sidorenko J, et al. (2024). GWAS summary statistics for body mass index [Dataset]. GWAS Catalog, Study Accession GCST90446645. https://www.ebi.ac.uk/gwas/studies/GCST90446645 | 650,000 | / | / |
| Major Depressive Disorder | Adams, Mark James; Lewis, Cathryn; Mcintosh, Andrew (2025). GWAS summary statistics for major depression (PGC MDD2025). figshare. Dataset. https://doi.org/10.6084/m9.figshare.27061255.v4 | 2,000,702 | 412,305 | 1,588,397 |
| Obsessive Compulsive Disorder | Sullivan, Patrick (2025). ocd2025. figshare. Dataset. https://doi.org/10.6084/m9.figshare.28707155.v4 | 1,138,106 | 23,493 | 1,114,613 |
| Post Traumatic Stress Disorder | Sullivan, Patrick (2024). ptsd2024. figshare. Dataset. https://doi.org/10.6084/m9.figshare.26349322.v3 | 1,222,882 | 137,136 | 1,085,746 |
| Schizophrenia | Sullivan, Patrick (2022). scz2022. figshare. Dataset. https://doi.org/10.6084/m9.figshare.19426775.v7 | 127,906 | 52,017 | 75,889 |

**Table S2.** Model fit indices for candidate Genomic SEM factor models.

|  | **chisq** | **df** | **p_chisq** | **AIC** | **CFI** | **SRMR** |
| --- | --- | --- | --- | --- | --- | --- |
| Two factor | 1461.685 | 40 | 8.67E-281 | 1513.685 | 0.905604 | 0.096298 |
| Three factor | 557.8551 | 37 | 3.25E-94 | **615.8551** | 0.965417 | 0.064427 |
| P factor | 557.8214 | 33 | 1.22E-96 | 623.8214 | 0.965153 | 0.064428 |

Note:

chisq: chi-square test statistic;

df: degrees of freedom;

p_chisq: p-value for chi-square test;

AIC: Akaike Information Criterion;

CFI: Comparative Fit Index;

SRMR: Standardized Root Mean Square Residual.

**Table S3. Genome-wide significant loci from meta-analysis of UK Biobank/QSKIN and MVP datasets.**

| **SNP** | **CHR** | **BP** | **A1** | **A2** | ***P_meta*** | **BETA_meta** | ***P1*** | **BETA1** | ***P2*** | **BETA2** | **Nearest protein-coding genes**  **(20kb)** | **Nearest protein-coding genes**  **(250kb)** | **Nearest non-protein-coding genes (20kb)** | **Nearest non-protein-coding genes**  **(250kb)** | **cytoBand** | **annotation** |
| --- | --- | --- | --- | --- | --- | --- | --- | --- | --- | --- | --- | --- | --- | --- | --- | --- |
| rs2097532 | 1 | 20225252 | T | C | 8.19E-09 | -0.0206 | 8.73E-05 | -0.024 | 1.96E-05 | -0.018821754 |  | UBXN10 | LINC01757 | LINC01757 | 1p36.12 | new |
| rs6677366 | 1 | 36721903 | A | G | 4.28E-08 | 0.021 | 0.005727 | 0.0178 | 1.93E-06 | -0.022739487 |  | CSF3R;GRIK3 |  |  | 1p34.3 | new |
| rs4546893 | 1 | 66017839 | A | T | 3.96E-10 | 0.0229 | 2.97E-06 | 0.0291 | 1.58E-05 | 0.01959065 | PDE4B | PDE4B |  |  | 1p31.3 | known |
| rs12031924 | 1 | 96411849 | T | C | 3.96E-09 | 0.0238 | 0.0003688 | 0.0246 | 2.80E-06 | 0.023370986 |  |  |  | LINC01787 | 1p21.3 | new |
| rs75641275 | 1 | 97861577 | A | C | 6.44E-09 | -0.0267 | 1.49E-07 | -0.044 | 0.0004472 | 0.019284761 | DPYD | DPYD |  |  | 1p21.3 | known |
| rs111760035 | 1 | 111800478 | A | C | 1.50E-08 | 0.0245 | 0.00109 | 0.0249 | 3.87E-06 | 0.024292693 | KCND3 | KCND3 |  |  | 1p13.2 | new |
| rs143847321 | 1 | 150294896 | CAAAAAA | C | 1.89E-08 | 0.021 | 0.0001254 | 0.0236 | 3.90E-05 | 0.019488677 | MRPS21 | MRPS21 |  |  | 1q21.2 | new |
| rs577033996 | 1 | 174259097 | G | GT | 5.25E-13 | 0.0299 | 5.88E-06 | 0.0304 | 1.96E-08 | -0.029558802 | RABGAP1L | RABGAP1L |  |  | 1q25.1 | known |
| rs2296296 | 1 | 183136264 | C | G | 2.15E-11 | 0.0227 | 0.000134 | 0.0226 | 3.99E-08 | -0.022739487 | LAMC1 | LAMC1 |  |  | 1q25.3 | known |
| rs1912942 | 1 | 189138502 | A | G | 4.10E-10 | 0.0229 | 2.39E-06 | 0.0278 | 2.22E-05 | -0.019802627 |  |  |  |  | 1q31.1 | new |
| rs12091558 | 1 | 243290608 | T | C | 2.01E-10 | 0.0238 | 0.01443 | 0.0154 | 1.05E-09 | 0.028399475 | SDCCAG8 | SDCCAG8 |  |  | 1q43 | known |
| rs2289081 | 2 | 20682080 | C | G | 8.73E-10 | 0.0202 | 2.69E-08 | 0.0342 | 0.0001602 | 0.014707629 | GDF7;LDAH | GDF7;LDAH |  |  | 2p24.1 | known |
| rs693 | 2 | 21009323 | A | G | 4.73E-10 | -0.022 | 4.28E-05 | -0.0241 | 2.33E-06 | -0.020782539 | APOB | APOB |  |  | 2p24.1 | known |
| rs6759679 | 2 | 24758829 | A | C | 9.89E-09 | 0.0371 | 3.25E-05 | 0.0426 | 6.64E-05 | 0.03334998 | NCOA1 | NCOA1 |  |  | 2p23.3 | new |
| rs940052 | 2 | 45754003 | A | G | 4.67E-09 | -0.0208 | 4.86E-05 | -0.0241 | 2.05E-05 | 0.018877056 | PRKCE | PRKCE |  |  | 2p21 | new |
| rs2061028 | 2 | 52737131 | A | G | 9.41E-12 | 0.0236 | 0.05275 | 0.0117 | 3.22E-12 | -0.029558802 |  |  |  | MIR4431 | 2p16.2 | known |
| rs75439645 | 2 | 55821809 | A | G | 1.30E-08 | 0.0298 | 6.92E-07 | 0.0432 | 0.0007316 | 0.022143365 |  | PNPT1;EFEMP1 |  |  | 2p16.1 | new |
| rs1430785 | 2 | 67642160 | A | G | 2.05E-19 | 0.0392 | 2.30E-08 | 0.041 | 1.76E-12 | -0.038258712 |  | ETAA1 |  | LINC01812 | 2p14 | known |
| rs12477289 | 2 | 76083391 | T | C | 4.59E-09 | -0.0272 | 2.40E-05 | -0.034 | 3.04E-05 | -0.023716527 |  |  |  |  | 2p12 | new |
| rs61668542 | 2 | 83019019 | T | G | 4.52E-08 | -0.0272 | 0.0005727 | -0.0303 | 1.95E-05 | 0.025728149 |  |  |  | DHFRP3 | 2p12 | new |
| rs7607172 | 2 | 99875673 | T | C | 2.30E-12 | -0.025 | 2.77E-06 | -0.028 | 1.33E-07 | 0.023370986 | AFF3 | AFF3 |  |  | 2q11.2 | known |
| rs36025546 | 2 | 109221979 | A | AT | 2.29E-08 | 0.0201 | 6.78E-05 | 0.0244 | 6.39E-05 | -0.017839918 | SH3RF3 | SH3RF3 |  |  | 2q12.3 | new |
| rs7581524 | 2 | 112476739 | T | C | 1.52E-11 | 0.0257 | 1.14E-05 | 0.029 | 2.55E-07 | 0.024087796 | TTL | RGPD8;TTL |  |  | 2q13 | new |
| rs3754855 | 2 | 118851006 | A | G | 6.18E-10 | -0.0221 | 0.0002125 | -0.0224 | 6.41E-07 | 0.021938908 | EN1 | EN1;MARCO |  |  | 2q14.2 | new |
| rs5834968 | 2 | 143738258 | A | AT | 1.80E-10 | 0.0224 | 0.0001083 | 0.0238 | 3.39E-07 | -0.021761492 | ARHGAP15 | ARHGAP15 |  |  | 2q22.3 | new |
| rs9287989 | 2 | 175853013 | T | C | 2.84E-10 | 0.0216 | 8.33E-05 | 0.0232 | 7.70E-07 | -0.020782539 |  | LNPK |  |  | 2q31.1 | new |
| rs4305233 | 2 | 181177718 | T | C | 1.16E-09 | -0.0224 | 5.31E-05 | -0.025 | 4.49E-06 | 0.021019367 |  |  | LINC01934 | LINC01934 | 2q31.3 | new |
| rs10208097 | 2 | 193142779 | T | C | 1.84E-14 | -0.0275 | 9.06E-06 | -0.0271 | 4.32E-10 | 0.027679569 |  |  |  |  | 2q32.3 | known |
| rs10931833 | 2 | 198819343 | T | G | 1.82E-13 | 0.0296 | 0.0002728 | 0.0248 | 1.10E-10 | 0.032110054 |  |  |  |  | 2q33.1 | known |
| rs6435049 | 2 | 200336095 | A | G | 5.17E-10 | -0.0257 | 0.0002762 | -0.0251 | 4.68E-07 | 0.026036015 | SPATS2L | SPATS2L |  |  | 2q33.1 | new |
| rs2160328 | 2 | 205777613 | A | G | 4.12E-12 | 0.0244 | 0.0001779 | 0.0227 | 5.87E-09 | 0.025317808 | NRP2 | NRP2 |  |  | 2q33.3 | known |
| rs10191395 | 2 | 210499273 | A | G | 2.87E-08 | -0.0203 | 0.08387 | -0.0106 | 1.81E-08 | -0.025667747 | CPS1 | CPS1 |  |  | 2q34 | known |
| rs12995875 | 2 | 221494322 | T | C | 1.23E-09 | 0.0232 | 6.54E-06 | 0.03 | 2.39E-05 | -0.019802627 | EPHA4 | EPHA4 |  |  | 2q36.1 | new |
| rs6704768 | 2 | 232727791 | A | G | 2.79E-08 | 0.0194 | 0.111 | 0.0095 | 1.15E-08 | 0.024702613 | GIGYF2 | GIGYF2 |  |  | 2q37.1 | known |
| rs71312384 | 3 | 8429450 | T | C | 1.38E-11 | 0.0383 | 0.0007104 | 0.0327 | 3.60E-09 | -0.041141943 |  |  | LMCD1-AS1 | LMCD1-AS1 | 3p26.1 | known |
| rs1404481 | 3 | 43965174 | T | C | 1.89E-10 | -0.0226 | 1.18E-05 | -0.0259 | 3.04E-06 | -0.020782539 |  | ABHD5 |  | MIR138-1 | 3p21.33 | new |
| rs1858828 | 3 | 50204954 | T | G | 3.49E-10 | 0.0214 | 3.23E-08 | 0.0327 | 0.0001664 | 0.01572296 | SLC38A3 | SLC38A3 |  |  | 3p21.31 | known |
| rs34908451 | 3 | 52351099 | T | C | 4.55E-11 | -0.0295 | 0.0007461 | -0.0257 | 1.32E-08 | -0.031498667 | DNAH1 | DNAH1 |  |  | 3p21.1 | known |
| rs11130474 | 3 | 55455807 | C | G | 1.13E-09 | -0.0216 | 0.0003419 | -0.0212 | 8.79E-07 | -0.021761492 | WNT5A | WNT5A |  |  | 3p14.3 | new |
| rs35975336 | 3 | 58465907 | A | G | 4.05E-08 | 0.0271 | 2.83E-07 | 0.0415 | 0.00283 | 0.018571386 |  | PDHB;KCTD6 |  |  | 3p14.3 | new |
| rs6549363 | 3 | 70903502 | C | G | 1.22E-11 | 0.0247 | 1.81E-08 | 0.0358 | 1.25E-05 | 0.019386714 |  | FOXP1 |  |  | 3p13 | known |
| rs4856535 | 3 | 84624395 | A | C | 2.39E-10 | 0.0238 | 0.0003238 | 0.0229 | 1.70E-07 | 0.024292693 |  |  | NONE;LINC00971 | NONE;LINC00971 | 3p12.1 | new |
| rs7633827 | 3 | 87722474 | T | C | 2.32E-08 | -0.0203 | 0.01299 | -0.0156 | 3.57E-07 | 0.022654691 |  | HTR1F |  |  | 3p11.2 | new |
| rs6802736 | 3 | 105402968 | T | C | 3.20E-08 | -0.0201 | 0.01547 | -0.0155 | 4.49E-07 | 0.022245609 | ALCAM | ALCAM |  |  | 3q13.11 | known |
| rs1800828 | 3 | 114172702 | C | G | 2.04E-08 | 0.0223 | 0.003995 | 0.0195 | 1.26E-06 | -0.023716527 | DRD3 | DRD3 |  |  | 3q13.31 | new |
| rs9883955 | 3 | 134653644 | T | G | 9.22E-10 | -0.0211 | 2.15E-05 | -0.0259 | 6.42E-06 | 0.018877056 | KY | KY;EPHB1 |  |  | 3q22.2 | known |
| rs10804686 | 3 | 143148948 | T | C | 5.95E-09 | 0.0209 | 0.000548 | 0.021 | 2.73E-06 | 0.02081514 |  | CHST2;SLC9A9 |  |  | 3q24 | new |
| rs223103 | 3 | 169382655 | A | G | 4.39E-10 | -0.0203 | 0.01251 | -0.0147 | 5.47E-09 | -0.022739487 | MECOM | MECOM |  |  | 3q26.2 | known |
| rs556464 | 3 | 173395890 | C | G | 1.13E-08 | -0.0196 | 3.96E-05 | -0.0243 | 4.58E-05 | 0.017146159 | NLGN1 | NLGN1 |  |  | 3q26.31 | new |
| rs1362828 | 3 | 177920817 | C | G | 3.26E-08 | -0.0195 | 1.33E-05 | -0.0268 | 0.0002551 | -0.015873349 |  |  |  | LINC02015 | 3q26.32 | new |
| rs4855125 | 3 | 179950091 | T | C | 1.33E-09 | 0.0328 | 0.02507 | 0.0207 | 4.55E-09 | 0.039260714 | PEX5L | PEX5L |  |  | 3q26.33 | known |
| rs35862164 | 3 | 183649736 | T | TA | 1.10E-12 | 0.028 | 0.06824 | 0.0125 | 1.33E-13 | -0.035367144 | KLHL24 | KLHL24 |  |  | 3q27.1 | known |
| rs2051559 | 4 | 3297073 | T | C | 5.11E-09 | -0.0304 | 0.008284 | -0.023 | 1.04E-07 | 0.034591445 | RGS12 | MSANTD1;RGS12 |  |  | 4p16.3 | known |
| rs13130879 | 4 | 23484360 | A | G | 1.60E-09 | -0.0221 | 2.65E-05 | -0.0267 | 8.66E-06 | -0.019802627 |  |  |  |  | 4p15.2 | new |
| rs1031035 | 4 | 101301927 | T | C | 2.30E-10 | -0.0242 | 2.35E-05 | -0.0274 | 1.79E-06 | 0.022450129 | PPP3CA | PPP3CA |  |  | 4q24 | new |
| rs11933762 | 4 | 118385971 | A | G | 7.54E-09 | -0.0276 | 0.365 | -0.0075 | 1.12E-10 | 0.037805715 |  | PRSS12 |  | CEP170P1 | 4q26 | known |
| rs1445842 | 4 | 137680624 | T | C | 4.29E-08 | -0.0253 | 0.0002642 | -0.0283 | 3.60E-05 | -0.023716527 |  |  |  | LINC02172 | 4q28.3 | new |
| rs1510719 | 4 | 140016962 | T | C | 5.02E-12 | 0.0244 | 5.21E-09 | 0.0354 | 1.37E-05 | -0.018821754 | MAML3 | MAML3 |  |  | 4q31.1 | known |
| rs10019224 | 4 | 173534758 | T | C | 7.10E-12 | -0.0231 | 7.21E-05 | -0.024 | 2.54E-08 | -0.022739487 |  |  | HAND2-AS1 | HAND2-AS1 | 4q34.1 | known |
| rs558995127 | 5 | 675470 | T | C | 7.59E-13 | -0.0363 | 4.67E-07 | -0.0435 | 2.16E-07 | 0.032419891 | TPPP | TPPP |  |  | 5p15.33 | known |
| rs13160920 | 5 | 29021441 | T | C | 8.94E-13 | 0.0258 | 1.96E-07 | 0.0312 | 4.93E-07 | -0.022739487 |  |  |  | LSP1P3;LOC101929645 | 5p13.3 | known |
| rs1809045 | 5 | 44256418 | T | C | 1.83E-10 | 0.0246 | 0.00956 | 0.017 | 1.94E-09 | -0.028587457 |  | FGF10 |  |  | 5p12 | known |
| rs10941692 | 5 | 45099676 | T | C | 1.88E-10 | 0.0283 | 0.0008923 | 0.0245 | 4.21E-08 | -0.030529205 |  | HCN1 |  |  | 5p12 | known |
| rs7732492 | 5 | 51490578 | A | G | 7.67E-11 | -0.0229 | 0.0005462 | -0.0205 | 3.28E-08 | 0.024292693 |  | ISL1 |  |  | 5q11.2 | known |
| rs10054434 | 5 | 62238708 | C | G | 1.63E-08 | 0.0277 | 0.003335 | 0.0241 | 1.21E-06 | 0.029737818 |  | KIF2A |  |  | 5q12.1 | new |
| rs12514758 | 5 | 77168785 | A | G | 4.20E-09 | 0.0214 | 0.1228 | 0.0095 | 7.92E-10 | -0.027615167 | PDE8B | PDE8B |  | ZBED3-AS1 | 5q13.3 | known |
| rs3105118 | 5 | 83568893 | C | G | 5.98E-09 | 0.0251 | 3.85E-08 | 0.0407 | 0.001298 | 0.017044435 | VCAN | VCAN |  |  | 5q14.3 | known |
| rs335210 | 5 | 123166170 | A | G | 3.65E-11 | -0.0241 | 2.47E-06 | -0.0287 | 2.05E-06 | 0.021530119 | PRDM6 | PRDM6 |  |  | 5q23.2 | new |
| rs736801 | 5 | 132497907 | T | C | 2.58E-09 | -0.022 | 0.0002257 | -0.0224 | 2.73E-06 | -0.021761492 | IRF1 | IRF1;IL5 |  |  | 5q31.1 | new |
| rs329117 | 5 | 134524410 | T | C | 1.06E-09 | 0.0224 | 2.14E-06 | 0.0283 | 4.77E-05 | -0.018821754 | JADE2 | JADE2 |  |  | 5q31.1 | new |
| rs703249 | 5 | 135637589 | T | C | 2.21E-10 | -0.0253 | 7.29E-05 | -0.0263 | 7.71E-07 | -0.024692613 | SLC25A48 | SLC25A48 |  |  | 5q31.1 | new |
| rs6894833 | 5 | 138357132 | A | G | 4.06E-12 | 0.0244 | 0.0003464 | 0.0219 | 2.61E-09 | -0.025667747 | KDM3B | KDM3B |  |  | 5q31.2 | known |
| rs7356639 | 5 | 144670490 | T | C | 1.28E-09 | 0.0216 | 2.26E-06 | 0.0297 | 3.67E-05 | -0.017839918 |  | KCTD16 |  |  | 5q31.3 | new |
| rs4867926 | 5 | 170148172 | T | G | 7.36E-12 | 0.0252 | 5.83E-06 | 0.0278 | 2.71E-07 | -0.023716527 |  | FOXI1 |  | LINC01187 | 5q35.1 | new |
| rs4959678 | 6 | 2502939 | A | C | 7.77E-12 | -0.0233 | 0.000126 | -0.0226 | 1.41E-08 | -0.023716527 |  |  |  | GMDS-DT;LINC01600 | 6p25.2 | known |
| rs59136961 | 6 | 34690788 | A | AGTTTT | 2.21E-18 | 0.0435 | 7.26E-06 | 0.0387 | 5.05E-14 | -0.045928932 |  |  |  |  |  | known |
| rs9472010 | 6 | 43117009 | A | G | 6.41E-09 | 0.02 | 3.88E-07 | 0.03 | 0.0004711 | 0.014809116 | PTK7 | PTK7 |  |  | 6p21.1 | new |
| rs11756556 | 6 | 64102820 | T | C | 1.41E-10 | 0.0253 | 0.03834 | 0.0138 | 1.21E-10 | -0.031498667 | EYS | EYS |  |  | 6q12 | known |
| rs7742854 | 6 | 98080267 | T | C | 7.76E-10 | -0.0222 | 3.31E-06 | -0.0281 | 3.01E-05 | -0.018821754 |  |  |  | MIR2113 | 6q16.1 | new |
| rs62429515 | 6 | 139968252 | T | C | 6.79E-10 | -0.0263 | 0.02616 | -0.0163 | 2.02E-09 | -0.031498667 |  |  | LOC100507477 | FILNC1;LOC100507477 | 6q24.1 | known |
| rs198683 | 6 | 142879410 | T | C | 7.13E-12 | -0.0232 | 5.03E-05 | -0.0241 | 3.62E-08 | -0.022739487 | HIVEP2 | HIVEP2 |  |  | 6q24.2 | known |
| rs949075 | 6 | 147652062 | A | G | 5.26E-10 | -0.0237 | 4.61E-06 | -0.0298 | 1.38E-05 | 0.020508877 |  | SAMD5 |  |  | 6q24.3 | new |
| rs3757323 | 6 | 151880872 | T | C | 9.58E-10 | 0.0225 | 0.0003229 | 0.022 | 7.78E-07 | -0.022739487 | ESR1 | ESR1 |  |  | 6q25.1 | new |
| rs12195182 | 6 | 161108526 | C | G | 4.13E-08 | -0.0204 | 0.02514 | -0.0135 | 1.87E-07 | -0.024692613 | MAP3K4 | MAP3K4 |  |  | 6q26 | new |
| rs1845532 | 6 | 165883713 | C | G | 1.67E-08 | -0.0187 | 0.0003576 | -0.0211 | 1.12E-05 | 0.017654935 |  | PDE10A |  | LINC00473 | 6q27 | new |
| rs11766944 | 7 | 1848415 | A | G | 1.92E-12 | -0.0301 | 4.79E-08 | -0.0417 | 1.72E-06 | -0.024692613 | MAD1L1 | MAD1L1 |  |  | 7p22.3 | known |
| rs73035954 | 7 | 3524172 | T | C | 1.01E-08 | -0.0233 | 4.56E-05 | -0.0279 | 4.39E-05 | -0.020782539 | SDK1 | SDK1 |  |  | 7p22.2 | known |
| rs7802254 | 7 | 8657542 | C | G | 4.66E-12 | 0.0262 | 0.003407 | 0.0193 | 1.61E-10 | -0.029558802 | NXPH1 | NXPH1 |  |  | 7p21.3 | known |
| rs10264177 | 7 | 32331250 | A | G | 9.42E-10 | -0.0212 | 8.56E-06 | -0.0277 | 1.26E-05 | 0.018265809 | PDE1C | PDE1C |  |  | 7p14.3 | new |
| rs1533305 | 7 | 109065335 | A | G | 3.36E-08 | 0.0217 | 0.01777 | 0.016 | 3.61E-07 | -0.024692613 |  |  |  | C7orf66 | 7q31.1 | new |
| rs8180817 | 7 | 114407487 | C | G | 5.66E-09 | -0.0202 | 1.39E-06 | -0.0289 | 0.0001946 | -0.015873349 | FOXP2 | FOXP2 | NONE | NONE | 7q31.1 | known |
| rs2237717 | 7 | 116765333 | T | C | 3.47E-09 | -0.02 | 0.0006348 | -0.0202 | 1.47E-06 | 0.019896632 | MET | MET |  |  | 7q31.2 | new |
| rs7786867 | 7 | 121067688 | T | C | 3.12E-11 | -0.0236 | 0.0132 | -0.0153 | 1.81E-10 | -0.027615167 | CPED1 | CPED1 |  |  | 7q31.31 | known |
| rs12706746 | 7 | 126802805 | A | G | 1.54E-10 | 0.0242 | 4.24E-09 | 0.0369 | 0.0003059 | 0.017044435 | GRM8 | GRM8 |  |  | 7q31.33 | known |
| rs10232060 | 7 | 146995375 | A | G | 5.60E-09 | -0.0222 | 0.001377 | -0.0211 | 1.07E-06 | -0.022739487 | CNTNAP2 | CNTNAP2 |  |  | 7q35 | new |
| rs11774014 | 8 | 12840665 | A | C | 6.48E-09 | -0.0349 | 0.002638 | -0.0278 | 4.25E-07 | 0.040093094 |  | TRMT9B |  | LINC00681 | 8p23.1 | new |
| rs2410129 | 8 | 13938532 | C | G | 9.70E-09 | 0.0205 | 0.1069 | 0.0099 | 3.54E-09 | 0.026036015 |  | SGCZ |  |  | 8p22 | known |
| rs2929052 | 8 | 55450765 | A | T | 1.51E-09 | -0.0228 | 0.0001776 | -0.0249 | 2.17E-06 | -0.021761492 |  |  | SBF1P1 | SBF1P1 | 8q12.1 | new |
| rs11779044 | 8 | 67169660 | A | G | 1.04E-09 | -0.0225 | 8.47E-05 | -0.024 | 2.93E-06 | 0.0216323 | CSPP1 | CSPP1 |  |  | 8q13.2 | new |
| rs17291214 | 8 | 73456854 | A | G | 1.56E-08 | 0.0195 | 0.009083 | 0.0156 | 3.70E-07 | 0.021427947 | STAU2 | STAU2 |  |  | 8q21.11 | new |
| rs35687905 | 8 | 73914330 | T | C | 4.77E-09 | -0.0316 | 9.21E-05 | -0.0364 | 1.08E-05 | 0.029119899 |  | ELOC |  | LINC01617 | 8q21.11 | new |
| rs527272483 | 8 | 108206595 | T | C | 1.90E-08 | 0.0183 | 0.005815 | 0.0167 | 8.67E-07 | 0.018978967 | EIF3E | EIF3E |  |  | 8q23.1 | new |
| rs13274202 | 8 | 142678748 | T | C | 1.72E-10 | 0.0224 | 0.0001628 | 0.024 | 2.27E-07 | -0.021761492 | JRK;PSCA | JRK;PSCA |  |  | 8q24.3 | known |
| rs61090372 | 9 | 31201623 | A | T | 1.21E-08 | 0.023 | 7.36E-05 | 0.0273 | 3.03E-05 | -0.020782539 |  |  |  | LINC01243 | 9p21.1 | new |
| rs10761282 | 9 | 93823893 | A | G | 4.46E-18 | -0.0299 | 0.005034 | -0.0168 | 6.49E-18 | -0.036331929 |  | BARX1 | MIR4291 | MIR4291 | 9q22.31 | known |
| rs10993394 | 9 | 94916629 | A | G | 1.63E-08 | -0.0203 | 0.001256 | -0.0193 | 3.39E-06 | -0.020782539 | AOPEP | AOPEP |  |  | 9q22.32 | new |
| rs78073895 | 9 | 117745032 | CA | C | 2.37E-08 | -0.0195 | 0.001724 | -0.0189 | 4.01E-06 | -0.019802627 |  | TLR4 |  |  | 9q33.1 | new |
| rs10118767 | 9 | 123761017 | T | C | 5.29E-09 | 0.0248 | 0.01095 | 0.0189 | 9.26E-08 | 0.027782381 | DENND1A | DENND1A |  |  | 9q33.3 | new |
| rs10760348 | 9 | 124301272 | A | G | 3.45E-08 | 0.0191 | 0.003011 | 0.0176 | 3.29E-06 | 0.019896632 | NEK6 | NEK6 |  |  | 9q33.3 | new |
| rs7024396 | 9 | 125892044 | T | C | 3.52E-22 | 0.0351 | 6.29E-06 | 0.0283 | 3.90E-18 | 0.03853295 | PBX3 | PBX3 |  |  | 9q33.3 | known |
| rs4072289 | 9 | 126652045 | T | C | 1.73E-10 | 0.0218 | 8.19E-05 | 0.0244 | 4.41E-07 | 0.020713042 | LMX1B | LMX1B |  |  | 9q33.3 | new |
| rs1802669 | 10 | 21538867 | A | G | 5.19E-10 | 0.0225 | 4.29E-05 | 0.0255 | 2.66E-06 | 0.020917248 | MLLT10 | MLLT10 |  |  | 10p12.31 | new |
| rs4144586 | 10 | 22498773 | T | G | 1.88E-08 | 0.0241 | 0.005326 | 0.0197 | 7.35E-07 | -0.026641931 |  | PIP4K2A |  | LOC100499489 | 10p12.2 | known |
| rs4562750 | 10 | 76600464 | T | C | 5.31E-10 | -0.021 | 0.0009723 | -0.0195 | 1.39E-07 | -0.021761492 |  | LRMDA |  |  | 10q22.3 | new |
| rs10786736 | 10 | 103089359 | C | G | 3.21E-09 | -0.0344 | 0.009217 | -0.0255 | 5.44E-08 | -0.039220713 | CNNM2;NT5C2 | CNNM2;NT5C2 |  |  | 10q24.32 | new |
| rs11596214 | 10 | 104694074 | A | G | 9.63E-09 | -0.0208 | 0.0006233 | -0.0207 | 3.73E-06 | -0.020782539 | SORCS3 | SORCS3 |  |  | 10q25.1 | new |
| rs2806446 | 10 | 133134745 | A | G | 6.73E-10 | 0.0247 | 2.65E-05 | 0.0287 | 5.05E-06 | 0.022552404 | ADGRA1 | ADGRA1;KNDC1 |  |  | 10q26.3 | new |
| rs12792379 | 11 | 6247316 | A | G | 3.61E-14 | 0.0313 | 1.31E-07 | 0.0375 | 3.11E-08 | 0.02809088 | CNGA4;CCKBR | CNGA4;CCKBR |  |  | 11p15.4 | known |
| rs11042765 | 11 | 10382719 | A | G | 3.36E-08 | -0.0231 | 0.004052 | -0.0201 | 2.08E-06 | -0.024692613 |  |  | CAND1.11 | CAND1.11 | 11p15.4 | new |
| rs1481890 | 11 | 13212845 | T | C | 1.48E-09 | 0.0205 | 0.03909 | 0.0122 | 2.97E-09 | 0.024497632 |  | RASSF10 |  | ARNTL | 11p15.2 | known |
| rs4440995 | 11 | 31816580 | A | G | 1.87E-10 | 0.029 | 3.13E-06 | 0.0353 | 7.32E-06 | 0.025420377 |  |  | PAX6-AS1 | PAX6-AS1 | 11p13 | new |
| rs10160794 | 11 | 32485294 | T | C | 9.21E-16 | -0.0275 | 2.48E-05 | -0.0251 | 6.03E-12 | -0.028587457 |  | EIF3M |  | WT1-AS | 11p13 | known |
| rs35893832 | 11 | 35410159 | A | AT | 1.32E-08 | -0.0228 | 3.84E-05 | -0.0285 | 5.51E-05 | 0.019896632 | SLC1A2 | SLC1A2 |  |  | 11p13 | new |
| rs566204 | 11 | 95847762 | A | G | 2.97E-08 | -0.0197 | 0.00127 | -0.0196 | 6.26E-06 | -0.019802627 | MTMR2 | MTMR2 |  |  | 11q21 | new |
| rs4533068 | 11 | 112962896 | A | G | 6.08E-09 | 0.0309 | 1.44E-06 | 0.0445 | 0.0002154 | 0.024087796 |  |  | LOC101928847 | LOC101928847 | 11q23.2 | new |
| rs12226068 | 11 | 118717175 | A | G | 3.10E-08 | -0.0272 | 7.59E-06 | -0.0369 | 0.0003903 | -0.021761492 |  | TREH;DDX6 |  |  | 11q23.3 | new |
| rs1500105 | 12 | 114027 | T | C | 6.34E-09 | 0.0225 | 0.03314 | 0.0131 | 9.09E-09 | 0.02850236 | IQSEC3 | IQSEC3 |  |  | 12p13.33 | known |
| rs73057214 | 12 | 15320836 | T | C | 4.87E-09 | -0.0235 | 2.16E-07 | -0.0382 | 0.0002802 | 0.017349638 | PTPRO | RERG;PTPRO |  |  | 12p12.3 | new |
| rs2279537 | 12 | 19275075 | A | T | 3.21E-08 | 0.0253 | 0.0007828 | 0.0259 | 1.13E-05 | 0.025010163 | PLEKHA5 | PLEKHA5 |  |  | 12p12.3 | new |
| rs773110 | 12 | 55981353 | C | G | 1.10E-09 | 0.0221 | 4.99E-08 | 0.0341 | 0.0003903 | -0.015873349 | RAB5B | RAB5B |  |  | 12q13.2 | known |
| rs12308242 | 12 | 77857660 | T | C | 8.83E-16 | -0.0282 | 3.31E-06 | -0.0274 | 5.11E-11 | -0.028587457 | NAV3 | NAV3 |  |  | 12q21.2 | known |
| rs35350651 | 12 | 111469627 | A | AC | 2.45E-21 | 0.0319 | 4.35E-08 | 0.0329 | 9.91E-15 | -0.031498667 | ATXN2 | ATXN2 |  |  | 12q24.12 | known |
| rs7303456 | 12 | 122909471 | A | T | 4.86E-09 | -0.0218 | 0.03523 | -0.0131 | 1.00E-08 | -0.026641931 | VPS37B;ABCB9 | VPS37B;ABCB9 |  |  | 12q24.31 | known |
| rs4943172 | 13 | 33999808 | T | C | 2.20E-12 | 0.0262 | 0.0003214 | 0.0232 | 1.26E-09 | -0.027615167 |  | RFC3 |  |  | 13q13.2 | known |
| rs9544418 | 13 | 35477382 | A | T | 3.87E-15 | -0.0287 | 2.63E-06 | -0.0289 | 3.60E-10 | -0.028587457 | NBEA | NBEA |  |  | 13q13.3 | known |
| rs9538483 | 13 | 59645292 | A | T | 4.62E-09 | 0.0337 | 0.001025 | 0.0323 | 1.24E-06 | -0.034401427 |  | DIAPH3 |  |  | 13q21.2 | new |
| rs1822212 | 13 | 66754426 | A | T | 1.04E-10 | 0.0222 | 0.000728 | 0.021 | 3.63E-08 | -0.022739487 | PCDH9 | PCDH9 |  |  | 13q21.32 | known |
| rs9574701 | 13 | 80617715 | T | C | 1.24E-09 | 0.0247 | 0.02611 | 0.0156 | 4.23E-09 | 0.029428811 |  |  |  |  | 13q31.1 | known |
| rs9549625 | 13 | 111963965 | A | G | 2.94E-10 | 0.0228 | 8.91E-06 | 0.0273 | 5.93E-06 | 0.02040681 |  |  | SOX1-OT | LINC00354;SOX1-OT | 13q34 | new |
| rs10132715 | 14 | 41712900 | A | G | 1.59E-08 | -0.0185 | 0.0004722 | -0.0207 | 8.49E-06 | 0.017553159 | LRFN5 | LRFN5 |  |  | 14q21.1 | new |
| rs7151970 | 14 | 101128268 | A | G | 1.25E-10 | -0.0258 | 5.30E-05 | -0.028 | 5.54E-07 | -0.024692613 |  |  | LINC02285 | LINC02285 | 14q32.31 | new |
| rs4906269 | 14 | 102895680 | A | C | 3.99E-09 | -0.0276 | 3.40E-05 | -0.0327 | 2.05E-05 | 0.024805119 | TRAF3 | TRAF3 |  |  | 14q32.32 | new |
| rs58264392 | 15 | 41058574 | T | C | 2.01E-09 | -0.0232 | 9.16E-05 | -0.0255 | 5.19E-06 | 0.021938908 | INO80 | INO80 |  |  | 15q15.1 | new |
| rs2733330 | 15 | 57259591 | A | G | 1.54E-11 | 0.0276 | 4.45E-05 | 0.0275 | 8.72E-08 | -0.027615167 | TCF12 | TCF12 |  |  | 15q21.3 | known |
| rs150302 | 15 | 89403558 | C | G | 4.20E-12 | 0.0236 | 0.0002829 | 0.0215 | 3.39E-09 | 0.024702613 |  |  | LOC105371031 | LOC105371031 | 15q26.1 | known |
| rs34652584 | 15 | 96413575 | T | TA | 4.95E-09 | -0.0315 | 0.009799 | -0.0244 | 1.03E-07 | 0.035005609 |  | NR2F2 |  |  | 15q26.2 | new |
| rs75023450 | 16 | 4231665 | T | C | 4.74E-08 | -0.0228 | 0.0014 | -0.0229 | 9.03E-06 | -0.022739487 | SRL | SRL |  |  | 16p13.3 | new |
| rs2289314 | 16 | 4673450 | A | G | 2.95E-08 | -0.0197 | 0.0003391 | -0.0212 | 2.29E-05 | 0.018877056 | MGRN1 | MGRN1 |  |  | 16p13.3 | new |
| rs11075253 | 16 | 15054789 | A | C | 1.03E-08 | -0.0211 | 5.49E-05 | -0.026 | 2.70E-05 | -0.018821754 | NTAN1;PDXDC1 | NTAN1;PDXDC1 |  |  | 16p13.11 | new |
| rs9673356 | 16 | 53530276 | A | G | 2.35E-23 | -0.0347 | 1.71E-05 | -0.0268 | 8.82E-20 | 0.038429027 |  | AKTIP;RPGRIP1L |  |  | 16q12.2 | known |
| rs5816929 | 16 | 54162319 | A | AT | 2.79E-08 | -0.0215 | 0.004544 | -0.018 | 1.31E-06 | 0.023473356 |  | FTO |  | LINC02169 | 16q12.2 | new |
| rs62046253 | 16 | 60610717 | T | C | 1.99E-09 | -0.022 | 1.30E-08 | -0.0352 | 0.001053 | -0.014888612 |  |  |  |  | 16q21 | known |
| rs4281739 | 16 | 64266921 | T | C | 8.32E-10 | 0.0219 | 0.003187 | 0.0182 | 5.32E-08 | -0.023716527 |  |  | NONE | NONE | 16q21 | known |
| rs6416749 | 16 | 73066409 | T | C | 4.81E-14 | -0.0288 | 0.0005275 | -0.0221 | 8.82E-12 | -0.03246719 | ZFHX3 | ZFHX3 |  | HCCAT5 | 16q22.3 | known |
| rs35449523 | 16 | 77088398 | C | G | 1.40E-09 | -0.0216 | 2.64E-06 | -0.0286 | 4.40E-05 | 0.017960325 |  | MON1B |  | MIR4719 | 16q23.1 | new |
| rs4888635 | 16 | 77471140 | A | G | 2.92E-11 | 0.0244 | 5.13E-05 | 0.026 | 1.33E-07 | 0.023678127 |  | ADAMTS18 |  |  | 16q23.1 | new |
| rs62051937 | 16 | 86462921 | C | G | 2.31E-09 | 0.0251 | 5.21E-05 | 0.0294 | 7.76E-06 | 0.022961612 |  |  | FENDRR | LINC00917;FENDRR | 16q24.1 | new |
| rs13339485 | 16 | 89506697 | A | C | 3.16E-10 | -0.0225 | 5.42E-06 | -0.0274 | 8.81E-06 | 0.019794627 | ANKRD11;SPG7 | ANKRD11;SPG7 |  |  | 16q24.3 | new |
| rs36040607 | 17 | 1419944 | A | G | 3.49E-11 | -0.0321 | 4.38E-06 | -0.0396 | 1.07E-06 | -0.028587457 | CRK | CRK |  |  | 17p13.3 | new |
| rs35299172 | 17 | 3931304 | T | C | 1.16E-10 | -0.0313 | 0.002193 | -0.0257 | 9.97E-09 | 0.034177452 | ATP2A3 | ATP2A3 |  |  | 17p13.2 | known |
| rs55915523 | 17 | 8195447 | A | G | 2.92E-08 | 0.0207 | 0.01126 | 0.0164 | 5.63E-07 | -0.022739487 | BORCS6;AURKB | BORCS6;AURKB |  |  | 17p13.1 | new |
| rs1053739 | 17 | 45105740 | A | G | 1.02E-08 | 0.0211 | 7.65E-07 | 0.0306 | 0.0005139 | 0.015926151 | NMT1 | NMT1 |  |  | 17q21.31 | new |
| rs2158079 | 17 | 47741184 | T | C | 4.36E-08 | 0.0304 | 7.19E-05 | 0.0375 | 0.0001114 | -0.026641931 | TBX21 | TBX21 |  |  | 17q21.32 | known |
| rs35895680 | 17 | 48982960 | A | C | 3.27E-10 | -0.0246 | 1.43E-05 | -0.0275 | 5.08E-06 | -0.022739487 | GIP;IGF2BP1 | GIP;IGF2BP1 |  |  | 17q21.32 | new |
| rs12602632 | 17 | 52230528 | A | G | 6.57E-17 | 0.0297 | 4.51E-11 | 0.0398 | 3.90E-08 | 0.024190239 |  | CA10 |  | LINC01982 | 17q22 | known |
| rs60218670 | 17 | 52924072 | A | T | 5.80E-10 | -0.0238 | 0.07833 | -0.0118 | 2.48E-10 | 0.029634805 |  |  |  | LINC02089;LINC02876 | 17q22 | known |
| rs1567962 | 17 | 80945758 | T | C | 4.19E-11 | -0.0239 | 2.28E-05 | -0.0261 | 3.42E-07 | -0.022739487 | RPTOR | RPTOR |  |  | 17q25.3 | new |
| rs12963022 | 18 | 22558027 | A | G | 6.36E-09 | -0.0206 | 0.03038 | -0.0135 | 2.71E-08 | 0.023985363 |  | CTAGE1 |  | LOC101927571 | 18q11.2 | known |
| rs11665070 | 18 | 37572600 | A | G | 2.02E-10 | -0.0239 | 3.14E-05 | -0.0261 | 1.27E-06 | -0.022739487 | CELF4 | CELF4 |  | MIR4318 | 18q12.2 | new |
| rs4617956 | 18 | 44756457 | C | G | 7.26E-09 | -0.0201 | 0.001514 | -0.0189 | 1.18E-06 | -0.020782539 | SETBP1 | SETBP1 |  |  | 18q12.3 | new |
| rs299725 | 18 | 48642874 | A | G | 3.38E-09 | 0.025 | 2.16E-05 | 0.0311 | 2.40E-05 | 0.021938908 | CTIF | CTIF |  |  | 18q21.1 | new |
| rs2337106 | 18 | 48934533 | C | G | 4.22E-09 | -0.0212 | 0.001031 | -0.0194 | 1.04E-06 | 0.022245609 | SMAD7 | SMAD7 |  |  | 18q21.1 | new |
| rs8096647 | 18 | 52429667 | A | T | 4.17E-08 | -0.0199 | 0.003448 | -0.0175 | 3.00E-06 | 0.021325787 | DCC | DCC |  |  | 18q21.2 | new |
| rs35717258 | 18 | 53273732 | CT | C | 7.99E-09 | 0.0218 | 0.0002616 | 0.0227 | 8.13E-06 | 0.021223636 | DCC | DCC |  |  | 18q21.2 | new |
| rs1011392 | 18 | 55374672 | A | G | 3.64E-13 | -0.0261 | 1.17E-06 | -0.0307 | 4.61E-08 | 0.02388294 | TCF4 | TCF4 |  |  | 18q21.2 | known |
| rs11152213 | 18 | 60185715 | A | C | 6.29E-09 | 0.0236 | 0.021 | 0.016 | 3.86E-08 | -0.027615167 |  | MC4R |  |  | 18q21.32 | known |
| rs12462498 | 19 | 18722140 | T | C | 8.19E-22 | -0.0402 | 4.97E-10 | -0.0466 | 1.50E-13 | -0.037295785 | CRTC1 | CRTC1 |  |  | 19p13.11 | known |
| rs8105984 | 19 | 19263259 | T | C | 1.17E-10 | -0.0307 | 7.85E-05 | -0.0312 | 3.56E-07 | 0.030459207 | HAPLN4 | HAPLN4 |  |  | 19p13.11 | new |
| rs8106322 | 19 | 31533324 | A | G | 2.27E-08 | -0.0203 | 5.61E-05 | -0.0251 | 7.11E-05 | 0.017756722 |  | TSHZ3 |  | THEG5 | 19q12 | new |
| rs429358 | 19 | 44908684 | T | C | 4.67E-11 | 0.0316 | 0.01207 | 0.0205 | 2.78E-10 | -0.037295785 | APOE | APOE |  |  | 19q13.32 | known |
| rs7256524 | 19 | 45721396 | T | C | 3.56E-09 | 0.022 | 7.07E-05 | 0.0244 | 9.55E-06 | 0.020610954 | FBXO46 | FBXO46 |  |  | 19q13.32 | new |
| rs13043980 | 20 | 9065125 | T | C | 2.13E-09 | -0.0246 | 0.0009524 | -0.0228 | 5.62E-07 | -0.025667747 | PLCB4 | PLCB1;PLCB4 |  |  | 20p12.3 | new |
| rs6011155 | 20 | 63819311 | T | C | 4.16E-09 | 0.0208 | 1.34E-05 | 0.0266 | 3.94E-05 | -0.017839918 |  |  | ZBTB46-AS1 | ZBTB46-AS1 | 20q13.33 | new |
| rs4470425 | 22 | 40320484 | A | C | 8.26E-10 | 0.0235 | 0.001438 | 0.0203 | 1.19E-07 | 0.025317808 | TNRC6B | TNRC6B |  |  | 22q13.1 | new |
| rs11090045 | 22 | 41357599 | A | G | 2.96E-08 | 0.0212 | 0.007802 | 0.0172 | 8.15E-07 | 0.023370986 | ZC3H7B | ZC3H7B |  |  | 22q13.2 | new |

Note:

SNP: lead single-nucleotide polymorphism representing each genome-wide significant locus;

CHR: chromosome;

BP: base-pair position;

A1/A2: effect allele and non-effect allele;

P_meta: meta-analysis P-value from the combined UKB+QSKIN and MVP datasets;

BETA_meta: inverse-variance-weighted meta-analysis effect size;

P1/BETA1: association P-value and effect size for the UKB+QSKIN dataset;

P2/BETA2: association P-value and effect size for the MVP dataset;

Nearest protein-coding genes (20 kb/250 kb): protein-coding genes located within ±20 kb or ±250 kb of the lead SNP;

Nearest non-protein-coding genes (20 kb/250 kb): non-coding genes located within ±20 kb or ±250 kb of the lead SNP;

cytoBand: cytogenetic band location;

annotation: locus novelty status (new, not previously reported in any GERD GWAS; known, previously reported).

**Table S4.** Genetic correlation between GERD and psychiatric traits.

| **Trait1** | **Trait2** | **r_g_** | **SE** | ***P*** |
| --- | --- | --- | --- | --- |
| GERD | MDD | 0.5493 | 0.0168 | 6.2826E-234 |
|  | ANX | 0.5139 | 0.02 | 3.5752E-146 |
|  | PTSD | 0.5304 | 0.0217 | 2.6446E-132 |
|  | ADHD | 0.4925 | 0.0265 | 4.7901E-77 |
|  | BMI | 0.2325 | 0.0192 | 1.0775E-33 |
|  | OCD | 0.2475 | 0.0318 | 6.7316E-15 |
|  | BIP | 0.1176 | 0.0241 | 0.000001012 |
|  | ASD | 0.08 | 0.0375 | 0.0331 |
|  | SCZ | 0.0193 | 0.0209 | 0.3553 |
|  | AN | 0.0204 | 0.0296 | 0.4905 |

Note:

Trait1/Trait2: trait pair analyzed;

r_g_: genetic correlation estimate;

SE: standard error of rg;

*P*: P-value for genetic correlation.

**Table S5.** Confirmatory factor analysis (CFA) parameter estimates for the three-factor Genomic SEM model.

| **lhs** | **op** | **rhs** | **Unstand Est** | **Unstand SE** | **STD Genotype** | **STD Genotype SE** | **STD All** | ***p*** |
| --- | --- | --- | --- | --- | --- | --- | --- | --- |
| F1 | =~ | GERD | 0.238574 | 0.008558 | 0.761997 | 0.027334 | 0.761997 | 5.03E-171 |
| F1 | =~ | ADHD | 0.244179 | 0.008666 | 0.648772 | 0.023026 | 0.648772 | 1.15E-174 |
| F1 | =~ | ANX | 0.135255 | 0.005522 | 0.675615 | 0.027585 | 0.675615 | 1.80E-132 |
| F1 | =~ | BIP | 0.059255 | 0.008918 | 0.203165 | 0.030579 | 0.203165 | 3.05E-11 |
| F1 | =~ | MDD | 0.224531 | 0.003983 | 0.966 | 0.017136 | 0.966 | < 5e-300 |
| F1 | =~ | PTSD | 0.202682 | 0.004436 | 0.959497 | 0.021 | 0.959496 | < 5e-300 |
| F1 | =~ | BMI | 0.263569 | 0.020485 | 0.590493 | 0.045895 | 0.590493 | 6.98E-38 |
| F2 | =~ | ANX | 0.070355 | 0.005629 | 0.351431 | 0.02812 | 0.351431 | 7.70E-36 |
| F2 | =~ | AN | 0.182645 | 0.011174 | 0.499275 | 0.030545 | 0.499275 | 4.70E-60 |
| F2 | =~ | ASD | 0.153991 | 0.012208 | 0.459834 | 0.036453 | 0.459835 | 1.76E-36 |
| F2 | =~ | OCD | 0.242765 | 0.009787 | 0.84969 | 0.034256 | 0.84969 | 8.08E-136 |
| F2 | =~ | BMI | -0.28595 | 0.021181 | -0.64063 | 0.047452 | -0.64063 | 1.55E-41 |
| F3 | =~ | BIP | 0.189713 | 0.011144 | 0.650468 | 0.03821 | 0.650468 | 5.48E-65 |
| F3 | =~ | SCZ | 0.374375 | 0.014616 | 0.838987 | 0.032754 | 0.838987 | 1.05E-144 |
| F3 | =~ | GERD | -0.10633 | 0.008943 | -0.33961 | 0.028565 | -0.33961 | 1.35E-32 |
| F1 | ~~ | F2 | 0.585909 | 0.029106 | 0.585909 | 0.029106 | 0.585909 | 4.02E-90 |
| F1 | ~~ | F3 | 0.443718 | 0.024292 | 0.44372 | 0.024292 | 0.44372 | 1.55E-74 |
| F2 | ~~ | F3 | 0.588788 | 0.026966 | 0.588789 | 0.026966 | 0.588789 | 1.10E-105 |
| MDD | ~~ | MDD | 0.003611 | 0.001359 | 0.066845 | 0.025151 | 0.066845 | 0.007866962 |
| OCD | ~~ | OCD | 0.022695 | 0.004936 | 0.278028 | 0.060468 | 0.278027 | 4.27E-06 |
| SCZ | ~~ | SCZ | 0.058956 | 0.011141 | 0.2961 | 0.055952 | 0.2961 | 1.21E-07 |
| PTSD | ~~ | PTSD | 0.003541 | 0.001442 | 0.079367 | 0.032307 | 0.079367 | 0.014026059 |
| GERD | ~~ | GERD | 0.052314 | 0.00344 | 0.533679 | 0.035091 | 0.533679 | 3.10E-52 |
| ADHD | ~~ | ADHD | 0.082032 | 0.005403 | 0.579094 | 0.038145 | 0.579095 | 4.70E-52 |
| AN | ~~ | AN | 0.100466 | 0.00801 | 0.750724 | 0.059856 | 0.750725 | 4.39E-36 |
| ANX | ~~ | ANX | 0.005684 | 0.001405 | 0.141815 | 0.035057 | 0.141815 | 5.23E-05 |
| ASD | ~~ | ASD | 0.088435 | 0.009495 | 0.788549 | 0.084667 | 0.788552 | 1.24E-20 |
| BIP | ~~ | BIP | 0.035586 | 0.003323 | 0.418338 | 0.039068 | 0.418338 | 9.32E-27 |
| BMI | ~~ | BMI | 0.136312 | 0.008375 | 0.684194 | 0.042037 | 0.684193 | 1.47E-59 |
| F1 | ~~ | F1 | 1 |  | 1 |  | 1 | NA |
| F2 | ~~ | F2 | 1 |  | 1 |  | 1 | NA |
| F3 | ~~ | F3 | 1 |  | 1 |  | 1 | NA |

Note:

lhs: left-hand side variable (latent factor or observed trait);

op: operator (=~, factor loading; ~~, variance or covariance);

rhs: right-hand side variable;

Unstand Est: unstandardized parameter estimate;

Unstand SE: standard error of unstandardized estimate;

STD Genotype: standardized estimate on the genetic variance scale;

STD Genotype SE: standard error of standardized estimate;

STD All: fully standardized estimate;

*p*: *P*-value for parameter estimate.

**Table S6.** Local genetic correlation between GERD and related traits.

| **chr** | **start** | **stop** | **phen1** | **phen2** | **rho** | **p** | **direction** | **genes** |
| --- | --- | --- | --- | --- | --- | --- | --- | --- |
| 1 | 14720132 | 15755230 | GERD | ADHD | 0.63698 | 0.00316877 | positive | KAZN-AS1,FHAD1,TMEM51,TMEM51-AS1,KAZN,FHAD1-AS1,EFHD2,C1orf195 |
| 1 | 19238925 | 20556202 | GERD | ADHD | 0.637809 | 0.000467674 | positive | LOC100506730,HTR6,PLA2G5,LOC117779438,AKR7L,TMCO4,PLA2G2A,UBXN10,AKR7A2,MICOS10,PLA2G2E,CAPZB,MICOS10-NBL1,LOC105378614,IFFO2,UBR4,SLC66A1,EMC1,NBL1,MRTO4,RPS14P3,PLA2G2D,OTUD3,EMC1-AS1,PLA2G2F,RNF186,AKR7A3,PLA2G2C |
| 2 | 59251997 | 60775066 | GERD | ADHD | 0.512917 | 0.000100165 | positive | MIR4432HG,BCL11A,LINC01122,LINC01793,MIR4432 |
| 2 | 100202437 | 101556799 | GERD | ADHD | 0.652968 | 0.000301428 | positive | AFF3,LINC01104,LINC01849,LONRF2,NMS,CHST10,PDCL3,NPAS2 |
| 2 | 144503689 | 145977920 | GERD | ADHD | 0.67197 | 3.98628E-05 | positive | LINC01412,GTDC1,TEX41,ZEB2-AS1,ZEB2,LOC101928386,LOC105373656,ARHGAP15 |
| 2 | 149568876 | 151428988 | GERD | ADHD | 0.627996 | 0.00399016 | positive | LYPD6B,MIR9899,LINC01931,LINC01817,MMADHC-DT,KIF5C,RND3,LYPD6,LINC01920,MMADHC |
| 2 | 215899571 | 217566011 | GERD | ADHD | 0.428718 | 0.0034264 | positive | XRCC5,LINC00607,SMARCAL1,IGFBP5,RPL37A,LINC01963,TMEM169,LOC102724849,FN1,MREG,LINC01280,ABCA12,LINC01614,ATIC,MARCHF4,IGFBP2,PECR |
| 3 | 3128153 | 4109393 | GERD | ADHD | 0.603027 | 0.000259105 | positive | LOC100130207,IL5RA,TRNT1,LRRN1,CRBN |
| 3 | 42301658 | 44141693 | GERD | ADHD | 0.443327 | 0.0033612 | positive | HHATL-AS1,HIGD1A,VIPR1-AS1,CYP8B1,ANO10,NKTR,CCK,LYZL4,GASK1A,CCDC13,ACKR2,SNRK-AS1,KLHL40,KRBOX1,ABHD5,POMGNT2,SEC22C,LINC02158,HHATL,ZBTB47,LOC101928323,KRBOX1-AS1,CCDC13-AS1,SS18L2,ZNF662,SNRK,VIPR1 |
| 3 | 65326752 | 66702084 | GERD | ADHD | 1 | 1.78517E-05 | positive | MAGI1-AS1,MAGI1,MAGI1-IT1,SLC25A26,LRIG1 |
| 3 | 72334705 | 73606612 | GERD | ADHD | 0.553232 | 0.00406983 | positive | GXYLT2,LOC101927296,PDZRN3,EBLN2,PPP4R2,SHQ1,LOC105377162,RYBP |
| 3 | 83529395 | 84698480 | GERD | ADHD | 0.74745 | 0.00085769 | positive | LINC00971 |
| 3 | 127495174 | 129091975 | GERD | ADHD | 0.699877 | 0.00114592 | positive | H1-10-AS1,MIR12124,RPN1,LOC653712,MIR6826,CFAP92,GATA2,SEC61A1,COPG1,RUVBL1-AS1,LOC90246,EFCC1,EEFSEC,RAB7A,MGLL,CNBP,ISY1-RAB43,ACAD9,LINC01565,H1-10,KBTBD12,DNAJB8-AS1,GATA2-AS1,ISY1,RUVBL1,GP9,DNAJB8,RAB43,HMCES |
| 3 | 135288395 | 137372141 | GERD | ADHD | 0.76221 | 0.000329714 | positive | NCK1-DT,STAG1,PCCB,NCK1,SLC35G2,IL20RB,MSL2,PPP2R3A |
| 3 | 149998412 | 151131307 | GERD | ADHD | 0.653733 | 0.00419803 | positive | P2RY12,TSC22D2,EIF2A,MINDY4B,CLRN1-AS1,MED12L,LINC01214,ERICH6-AS1,GPR171,ERICH6,LOC101928105,P2RY13,SERP1,P2RY14,GPR87,CLRN1,SIAH2,SELENOT |
| 4 | 61971162 | 63337954 | GERD | ADHD | 0.917958 | 0.000434046 | positive | ADGRL3,LINC02271,ADGRL3-AS1 |
| 4 | 111733580 | 112830229 | GERD | ADHD | 0.821493 | 0.00320519 | positive | MIR297 |
| 4 | 139553761 | 141087047 | GERD | ADHD | 0.74058 | 1.03645E-06 | positive | NDUFC1,LOC105377448,NOCT,RAB33B-AS1,MGARP,SETD7,NAA15,MGST2,MAML3,RAB33B,ELF2,LOC101927516 |
| 4 | 186881569 | 187985393 | GERD | ADHD | -0.736904 | 0.00220039 | negative | F11-AS1,CYP4V2,KLKB1,MTNR1A,F11,TLR3,FAM149A,FAT1,FLJ38576 |
| 5 | 44049879 | 45305566 | GERD | ADHD | 0.864935 | 5.28184E-05 | positive | LINC02224,FGF10,FGF10-AS1,HCN1,MRPS30,MRPS30-DT |
| 5 | 91956906 | 93814604 | GERD | ADHD | 0.814407 | 3.74522E-05 | positive | NR2F1-AS1,POU5F2,KIAA0825,FAM172A,MIR2277,NR2F1 |
| 5 | 103788461 | 104850490 | GERD | ADHD | 0.664246 | 0.00365294 | positive | RAB9BP1 |
| 5 | 107265912 | 108641784 | GERD | ADHD | 0.826717 | 0.000347831 | positive | FBXL17,LOC285638,LINC01023,FER |
| 5 | 132554691 | 134556570 | GERD | ADHD | 0.502647 | 7.71972E-05 | positive | MIR3661,UBE2B,LINC01843,MIR1289-2,PITX1,SEC24A,WSPAR,DDX46,CAMLG,SKP1,JADE2,LOC102546229,C5orf66-AS1,LOC105379183,CDKL3,CATSPER3,TCF7,VDAC1,CDKN2AIPNL,C5orf24,C5orf66,SAR1B,FSTL4,PPP2CA,C5orf15,PCBD2,TXNDC15 |
| 5 | 162386028 | 163781268 | GERD | ADHD | 0.775391 | 0.00371943 | positive | MAT2B,HMMR-AS1,CCNG1,NUDCD2,HMMR |
| 5 | 170898341 | 172285682 | GERD | ADHD | 0.861031 | 0.000647046 | positive | FBXW11,ERGIC1,LOC100288254,NEURL1B,SMIM23,UBTD2,EFCAB9,STK10,LOC101928093,MIR5003,DUSP1,SH3PXD2B,LINC01944 |
| 6 | 19449153 | 20418020 | GERD | ADHD | 0.928879 | 8.68388E-06 | positive | MBOAT1,LOC105374960,E2F3,LNC-LBCS,ID4 |
| 6 | 33864263 | 34979270 | GERD | ADHD | 0.783153 | 0.000191461 | positive | TAF11,ANKS1A,SPDEF,PACSIN1,MIR7159,HMGA1,MIR6835,RPS10-NUDT3,UHRF1BP1,MIR1275,LOC101929243,ILRUN,GRM4,SMIM29,RPS10,NUDT3,LINC01016,SNRPC |
| 6 | 39785908 | 40778401 | GERD | ADHD | 0.82148 | 0.000135575 | positive | LRFN2,DAAM2-AS1,DAAM2,TDRG1,LINC00951,MOCS1 |
| 6 | 70363349 | 71610787 | GERD | ADHD | 0.791099 | 0.000149237 | positive | COL19A1,SDHAF4,LMBRD1,EVADR,FAM135A-AS1,B3GAT2,COL9A1,SMAP1,FAM135A |
| 6 | 93696826 | 95176337 | GERD | ADHD | 0.993817 | 0.000709077 | positive | TSG1,EPHA7 |
| 6 | 101793543 | 102637706 | GERD | ADHD | 1 | 1.94206E-05 | positive | GRIK2 |
| 6 | 163336362 | 164687462 | GERD | ADHD | 0.555158 | 0.0039528 | positive | CAHM,PACRG-AS3,DKFZp451B082,QKI,PACRG-AS1,PACRG-AS2,LOC102724152,PACRG |
| 7 | 1366973 | 2473749 | GERD | ADHD | 0.658508 | 0.000116729 | positive | NUDT1,LOC100128653,ELFN1,MIR4655,MRM2,CHST12,TFAMP1,INTS1,PSMG3,TMEM184A,SNX8,MAFK,ELFN1-AS1,SNORA114,EIF3B,MAD1L1,MIR6836,LOC105375303,MICALL2,PSMG3-AS1 |
| 7 | 113339387 | 115321301 | GERD | ADHD | 0.568175 | 0.00109173 | positive | MIR3666,LINC01392,LINC01393,PPP1R3A,FOXP2,SNORA25B,MDFIC |
| 8 | 36641175 | 38803980 | GERD | ADHD | 0.648243 | 0.000701016 | positive | GOT1L1,C8orf86,ADRB3,KCNU1,LOC101929622,FGFR1,RAB11FIP1,LETM2,LSM1,PLEKHA2,EIF4EBP1,LOC728024,RNF5P1,PLPP5,MIR1268A,BRF2,LINC01605,ZNF703,ASH2L,TACC1,ERLIN2,BAG4,LOC102723701,DDHD2,ADGRA2,PLPBP,NSD3,STAR |
| 8 | 92876621 | 94999066 | GERD | ADHD | 0.728666 | 1.29422E-06 | positive | C8orf87,LOC102724710,MIR378D2,RUNX1T1,LINC00535,FLJ46284,TMEM67,RBM12B,RBM12B-AS1,PDP1,CIBAR1,MIR8084,TRIQK |
| 9 | 15839404 | 16658377 | GERD | ADHD | 0.535955 | 0.00131527 | positive | CCDC171,BNC2,C9orf92 |
| 9 | 119001228 | 119890464 | GERD | ADHD | 0.80378 | 0.00143729 | positive | PAPPA-AS1,PAPPA,ASTN2-AS1,TRIM32,ASTN2 |
| 9 | 121008530 | 122677719 | GERD | ADHD | 1 | 1.07846E-09 | positive | BRINP1,LINC02578 |
| 9 | 127184583 | 128785782 | GERD | ADHD | 0.73983 | 0.00280881 | positive | MIR181B2,NR5A1,SCAI,MIR181A2HG,ARPC5L,LOC105376271,PPP6C,NR6A1,MIR181A2,RPL35,WDR38,GOLGA1,MAPKAP1,PBX3,HSPA5,LOC51145,GAPVD1,OLFML2A,RABEPK |
| 10 | 12581572 | 13208061 | GERD | ADHD | 0.91356 | 0.00100608 | positive | OPTN,CCDC3,MCM10,CAMK1D,MIR548Q,MIR4481,MIR4480 |
| 10 | 33655459 | 35178224 | GERD | ADHD | 0.551087 | 0.00222562 | positive | PARD3-AS1,PARD3,LINC00838 |
| 10 | 104206838 | 106142283 | GERD | ADHD | 0.684608 | 0.000282208 | positive | MIR609,WBP1L,ITPRIP,CFAP58,CFAP43,LOC107984265,GSTO2,ACTR1A,PCGF6,CYP17A1,NEURL1-AS1,INA,TAF5,COL17A1,RPARP-AS1,CNNM2,C10orf95,NEURL1,GSTO1,ARL3,CFAP58-DT,TRIM8,MFSD13A,SUFU,CALHM1,CALHM2,BORCS7,SFR1,SLK,CALHM3,ATP5MD,NT5C2,SFXN2,PDCD11,AS3MT,SH3PXD2A-AS1,MIR4482,SH3PXD2A,MIR1307,RPEL1,STN1,BORCS7-ASMT,ITPRIP-AS1,MIR936 |
| 10 | 106142284 | 107877787 | GERD | ADHD | 0.591044 | 0.000263655 | positive | LINC02627,CFAP58,SORCS3-AS1,SORCS3,LINC02620 |
| 11 | 112755447 | 113889019 | GERD | ADHD | 0.82841 | 3.23207E-06 | positive | HTR3B,TTC12,NCAM1,NCAM1-AS1,TMPRSS5,HTR3A,ZW10,USP28,DRD2,LOC101928847,MIR4301,CLDN25,ANKK1 |
| 11 | 133521643 | 134351064 | GERD | ADHD | 0.689779 | 5.09725E-05 | positive | SNORD153,LINC02743,IGSF9B,THYN1,GLB1L3,VPS26B,B3GAT1-DT,MIR4697,JAM3,ACAD8,GLB1L2,LINC02731,SPATA19,NCAPD3,B3GAT1 |
| 12 | 23923799 | 25058714 | GERD | ADHD | 0.535609 | 0.00392621 | positive | SOX5,BCAT1,MIR920,LINC00477,SOX5-AS1 |
| 13 | 107037865 | 108521978 | GERD | ADHD | 0.518106 | 0.000953476 | positive | EFNB2,SNORD31B,LINC00551,ARGLU1,FAM155A,MIR1267,LINC00443,FAM155A-IT1 |
| 14 | 29029225 | 30831154 | GERD | ADHD | 0.65586 | 2.79148E-05 | positive | LINC01551,LINC02282,PRKD1,FOXG1-AS1,FOXG1 |
| 14 | 74245429 | 75699501 | GERD | ADHD | 0.777821 | 0.00359393 | positive | ENTPD5,YLPM1,SYNDIG1L,FCF1,COQ6,ISCA2,LIN52,LTBP2,MLH3,ZC2HC1C,BBOF1,PTGR2,LINC02274,EIF2B2,NEK9,LOC100506476,ZNF410,DLST,PROX2,ALDH6A1,ACYP1,ABCD4,AREL1,PGF,NPC2,FAM161B,TMED10,MIR4709,VSX2,RPS6KL1,MIDEAS,VRTN |
| 15 | 69089816 | 70767983 | GERD | ADHD | 0.82786 | 4.01007E-07 | positive | EWSAT1,TLE3,PAQR5,PCAT29,GLCE,RPLP1,LOC145694,NOX5,KIF23,ANP32A-IT1,MIR4312,LINC00593,ANP32A,DRAIC,MIR629,SPESP1 |
| 15 | 98025685 | 98828261 | GERD | ADHD | 0.589345 | 0.0006331 | positive | LOC101927310,LINC02254,LINC00923,LINC02251,LINC01582,ARRDC4 |
| 16 | 61092845 | 62607090 | GERD | ADHD | 0.923478 | 7.2562E-06 | positive | CDH8 |
| 17 | 49909299 | 51110385 | GERD | ADHD | 0.822622 | 0.000853346 | positive | LINC02089,LINC01982,CA10,LINC02876 |
| 17 | 65582328 | 66757556 | GERD | ADHD | 0.714995 | 0.000912859 | positive | C17orf58,PRKAR1A,NOL11,BPTF,PITPNC1,KPNA2,MIR635,LINC00674,WIPI1,LINC01482,SLC16A6,FAM20A,AMZ2,SNORA38B,ARHGAP27P2,ARSG |
| 18 | 45718493 | 46558306 | GERD | ADHD | 0.681612 | 0.000309891 | positive | CTIF,C18orf12,LOC100129878,SMAD7,MIR4743,ZBTB7C |
| 18 | 52512524 | 53762996 | GERD | ADHD | 0.780301 | 0.000062898 | positive | LINC01539,TCF4,CCDC68,LINC01905,TCF4-AS1,RAB27B,LINC01415,MIR4529,LINC01929,LOC642484,LINC01416 |
| 18 | 77148138 | 78017072 | GERD | ADHD | 0.936829 | 0.000995181 | positive | TXNL4A,PARD6G,HSBP1L1,KCNG2,PARD6G-AS1,LOC284241,ADNP2,CTDP1,LOC284240,NFATC1,RBFADN,RBFA,SLC66A2 |
| 20 | 19957929 | 21682377 | GERD | ADHD | 0.797637 | 0.000113725 | positive | NAA20,KIZ-AS1,CRNKL1,LINC01727,NKX2-2,LINC01726,CFAP61,KIZ,XRN2,LINC00237,INSM1,RALGAPA2,RIN2,NKX2-4 |
| 21 | 18222038 | 19479024 | GERD | ADHD | 0.583398 | 0.001898 | positive | BTG3-AS1,LINC01549,BTG3,C21orf91,CHODL-AS1,CHODL,CXADR,C21orf91-OT1 |
| 21 | 39252060 | 40480831 | GERD | ADHD | 0.847138 | 0.0030591 | positive | KCNJ6,LINC01700,ERG,LINC01423,LOC101928398,KCNJ15,ETS2,DSCR10,DSCR4,LOC400867,DSCR8,LINC00114 |
| 8 | 92876621 | 94999066 | GERD | ASD | 1 | 2.03102E-07 | positive | C8orf87,LOC102724710,MIR378D2,RUNX1T1,LINC00535,FLJ46284,TMEM67,RBM12B,RBM12B-AS1,PDP1,CIBAR1,MIR8084,TRIQK |
| 8 | 107787164 | 108647463 | GERD | ASD | 0.90547 | 0.000182421 | positive | ANGPT1 |
| 2 | 181309940 | 182307877 | GERD | BIP | 0.876428 | 0.000170839 | positive | MIR4437,SCHLAP1,UBE2E3,LINC01934 |
| 11 | 112755447 | 113889019 | GERD | BIP | 0.714127 | 7.24179E-06 | positive | HTR3B,TTC12,NCAM1,NCAM1-AS1,TMPRSS5,HTR3A,ZW10,USP28,DRD2,LOC101928847,MIR4301,CLDN25,ANKK1 |
| 15 | 82191078 | 84209760 | GERD | BIP | -0.800435 | 1.89806E-05 | negative | LINC01583,RPS17,ACTG1P17,FSD2,C15orf40,SCARNA15,WHAMM,GOLGA6L10,HDGFL3,SNHG21,TM6SF1,HOMER2,BTBD1,EFL1,MEX3B,UBE2Q2P2,CPEB1,GOLGA6L9,AP3B2,ADAMTS7P1,LOC727751,LOC338963,SH3GL3,SAXO2,BNC1,MIR4515,RAMAC,GOLGA2P10,CPEB1-AS1,LOC102724034,GOLGA6L17P |
| 20 | 56951774 | 57924037 | GERD | BIP | -0.604452 | 0.000155667 | negative | SLMO2-ATP5E,APCDD1L,CTSZ,MIR298,VAPB,LOC101927932,STX16-NPEPL1,LINC01711,TUBB1,ATP5F1E,STX16,LOC105372695,APCDD1L-DT,ZNF831,GNAS-AS1,EDN3,NELFCD,NPEPL1,MIR296,PRELID3B,GNAS |
| 1 | 4281477 | 4773403 | GERD | MDD | 0.493049 | 0.0216431 | positive | LINC01646,AJAP1,LINC01777 |
| 1 | 6136815 | 7711794 | GERD | MDD | 0.573976 | 0.00692749 | positive | KCNAB2,KLHL21,ZBTB48,PHF13,HES2,ACOT7,MIR4252,RNF207,PLEKHG5,TNFRSF25,NOL9,THAP3,CAMTA1-DT,CAMTA1-AS2,CAMTA1,HES3,LINC00337,CHD5,ESPN,DNAJC11,LINC01672,RPL22,ICMT,RNF207-AS1,TAS1R1,GPR153 |
| 1 | 7711795 | 8580987 | GERD | MDD | 0.677061 | 0.000158709 | positive | RERE-AS1,ERRFI1,SLC45A1,VAMP3,SNORD128,RERE,LINC01714,UTS2,PER3,TNFRSF9,CAMTA1,PARK7 |
| 1 | 8580988 | 9475966 | GERD | MDD | 0.840987 | 7.77677E-05 | positive | MIR34A,SLC2A5,LNCTAM34A,MIR34AHG,SPSB1,H6PD,CA6,GPR157,RERE,ENO1-AS1,MIR6728,SLC2A7,ENO1 |
| 1 | 17557747 | 18427820 | GERD | MDD | 0.698039 | 0.000157735 | positive | ARHGEF10L,PADI4,LINC01654,PADI6,PADI1,MIR3972,ACTL8,RCC2,PADI3 |
| 1 | 19238925 | 20556202 | GERD | MDD | 0.482169 | 0.00518318 | positive | LOC100506730,HTR6,PLA2G5,LOC117779438,AKR7L,TMCO4,PLA2G2A,UBXN10,AKR7A2,MICOS10,PLA2G2E,CAPZB,MICOS10-NBL1,LOC105378614,IFFO2,UBR4,SLC66A1,EMC1,NBL1,MRTO4,RPS14P3,PLA2G2D,OTUD3,EMC1-AS1,PLA2G2F,RNF186,AKR7A3,PLA2G2C |
| 1 | 20556203 | 21416121 | GERD | MDD | 0.688043 | 0.00124867 | positive | PINK1-AS,FAM43B,LINC01141,DDOST,MIR1256,MUL1,EIF4G3,LINC01757,SH2D5,CDA,VWA5B1,PINK1,CAMK2N1,MIR6084,HP1BP3,KIF17 |
| 1 | 29188427 | 30405182 | GERD | MDD | 0.633666 | 0.0108655 | positive | OPRD1,SRSF4,MECR,TMEM200B,EPB41,PTPRU,LINC01756 |
| 1 | 34371023 | 35201626 | GERD | MDD | 0.702334 | 0.000429046 | positive | MIR552,CSMD2,C1orf94 |
| 1 | 35201627 | 36855139 | GERD | MDD | 0.557792 | 0.00247865 | positive | GJB5,GJA4,GJB4,CLSPN,MAP7D1,TFAP2E,PSMB2,ZMYM4,TRAPPC3,AGO3,GJB3,LOC653160,SH3D21,SFPQ,KIAA0319L,ZMYM1,NCDN,THRAP3,C1orf216,AGO1,TEKT2,ADPRS,ZMYM4-AS1,ZMYM6,DLGAP3,AGO4,SMIM12,STK40,EVA1B,COL8A2,TMEM35B |
| 1 | 36855140 | 38474036 | GERD | MDD | 0.70979 | 3.3552E-08 | positive | MIR6732,SNIP1,MRPS15,ZC3H12A,MANEAL,FHL3,YRDC,CSF3R,INPP5B,C1orf122,CDCA8,MIR5581,MEAF6,OSCP1,C1orf109,SF3A3,DNALI1,MTF1,GRIK3,LINC01137,EPHA10,GNL2,LSM10,MIR4255,SNORA63C,RSPO1 |
| 1 | 38474037 | 40200950 | GERD | MDD | 0.714965 | 1.70077E-08 | positive | HPCAL4,MACF1,HEYL,PABPC4,AKIRIN1,SNORA55,RHBDL2,UTP11,NT5C1A,PPIEL,GJA9,PABPC4-AS1,MYCBP,RRAGC,NDUFS5,POU3F1,OXCT2P1,GJA9-MYCBP,LINC01343,LOC105378663,BMP8A,MIR3659,MIR3659HG,LINC01685,KIAA0754 |
| 1 | 41093876 | 41977966 | GERD | MDD | 0.66225 | 0.0119418 | positive | NFYC,SLFNL1,RIMS3,CTPS1,FOXO6,KCNQ4,MIR30C1,MIR30E,NFYC-AS1,EDN2,HIVEP3,SLFNL1-AS1,FOXO6-AS1,CITED4,SCMH1 |
| 1 | 41977967 | 43512669 | GERD | MDD | 0.816048 | 1.71281E-05 | positive | C1orf50,ERMAP,SVBP,GUCA2B,GUCA2A,ZNF691,P3H1,YBX1,TMEM269-DT,LOC339539,SLC2A1,PPIH,FOXJ3,CCDC30,CLDN19,PPCS,TMEM269,SLC2A1-AS1,HIVEP3,ZMYND12,RIMKLA |
| 1 | 47257262 | 48105233 | GERD | MDD | 0.644807 | 0.0171821 | positive | STIL,CYP4X1,CYP4Z2P,CYP4B1,FOXE3,CYP4A11,CYP4Z1,CYP4A22,LINC01389,FOXD2,PDZK1IP1,FOXD2-AS1,CMPK1,LINC00853,TAL1 |
| 1 | 49185415 | 51777577 | GERD | MDD | 0.633756 | 0.00989424 | positive | MIR4421,AGBL4-AS1,C1orf185,TTC39A,BEND5,AGBL4-IT1,RNF11,FAF1,ELAVL4,MIR6500,LINC01562,AGBL4,CDKN2C,DMRTA2 |
| 1 | 51777578 | 53114186 | GERD | MDD | 0.8553 | 0.00258072 | positive | TUT4,TXNDC12-AS1,RAB3B,TTC39A,OSBPL9,NRDC,ZFYVE9,TTC39A-AS1,EPS15,TXNDC12,ORC1,MIR761,KTI12,CALR4P,CC2D1B,PRPF38A,SHISAL2A,GPX7,BTF3L4 |
| 1 | 53114187 | 54771293 | GERD | MDD | 0.384421 | 0.0181365 | positive | TMEM59,CPT2,NDC1,LRRC42,PODN,SLC1A7,CYB5RL,ECHDC2,CZIB,LDLRAD1,DMRTB1,TCEANC2,LINC01771,CDCP2,MIR4781,ZYG11A,SSBP3,MAGOH,SLC25A3P1,SSBP3-AS1,YIPF1,LRP8,LOC100507564,COA7,SHISAL2A,MRPL37,SCP2,DIO1,HSPB11,GLIS1,ZYG11B |
| 1 | 65894185 | 66778015 | GERD | MDD | 0.546635 | 0.0263901 | positive | LEPR,LEPROT,PDE4B,PDE4B-AS1 |
| 1 | 68633861 | 69438909 | GERD | MDD | 0.529409 | 0.0248938 | positive | MIR1262,DEPDC1,WLS,DEPDC1-AS1,GNG12-AS1,RPE65 |
| 1 | 70992906 | 72513119 | GERD | MDD | 0.614721 | 0.00209997 | positive | ZRANB2-AS2,MIR186,LINC01788,ZRANB2-AS1,NEGR1,NEGR1-IT1,PTGER3,ZRANB2 |
| 1 | 72513120 | 73992170 | GERD | MDD | 0.846359 | 7.86309E-08 | positive | NEGR1,LINC01360 |
| 1 | 73992171 | 75132092 | GERD | MDD | 0.696054 | 1.08626E-05 | positive | FPGT,ERICH3-AS1,FPGT-TNNI3K,LINC02238,ERICH3,LRRC53,TNNI3K,LRRIQ3 |
| 1 | 79065287 | 80099353 | GERD | MDD | 0.717243 | 0.0138885 | positive | ADGRL4,IFI44,IFI44L |
| 1 | 82123369 | 83461215 | GERD | MDD | 0.559469 | 0.00453525 | positive | LINC01361,ADGRL2,LINC01362 |
| 1 | 84728348 | 86017267 | GERD | MDD | 1 | 2.12426E-05 | positive | LINC01461,MCOLN3,LPAR3,UOX,LINC01555,BCL10,SAMD13,GNG5,LOC646626,DNAI3,SYDE2,MCOLN2,MIR4423,DDAH1,RPF1,CTBS,SSX2IP,SPATA1,C1orf52,DNASE2B |
| 1 | 87667573 | 88656689 | GERD | MDD | 0.439017 | 0.00970837 | positive | LMO4,LINC02801,LINC01364 |
| 1 | 88656690 | 89753964 | GERD | MDD | 0.673184 | 0.000333806 | positive | PKN2,GBP5,RBMXL1,PKN2-AS1,GBP3,GBP1,KYAT3,GBP7,GBP2,GBP4,GTF2B |
| 1 | 89753965 | 90941549 | GERD | MDD | 0.640543 | 0.00249615 | positive | LOC729930,GBP6,LRRC8B,LRRC8C,LRRC8D,LRRC8C-DT,ZNF326,GBP1P1,GEMIN8P4 |
| 1 | 96147577 | 97721186 | GERD | MDD | 0.744865 | 5.22371E-06 | positive | LINC02790,DPYD-AS1,DPYD,PTBP2,LINC01787 |
| 1 | 97721187 | 99425000 | GERD | MDD | 0.690505 | 0.00267576 | positive | MIR137,DPYD-AS1,DPYD-AS2,LINC01930,MIR137HG,SNX7,DPYD,MIR2682,LINC01776,PLPPR5 |
| 1 | 111134063 | 112489248 | GERD | MDD | 0.579901 | 0.000285346 | positive | INKA2-AS1,WDR77,LOC101928718,LRIF1,KCND3,CHIAP2,KCND3-AS1,C1orf162,PIFO,DRAM2,DENND2D,DDX20,KCNA2,TMIGD3,KCND3-IT1,RAP1A,CD53,OVGP1,ADORA3,LINC01160,PGBP,ATP5PB,CHI3L2,CEPT1,CHIA,KCNA3,LOC107985184,INKA2 |
| 1 | 114664388 | 116207297 | GERD | MDD | 0.410914 | 0.010065 | positive | TSHB,VANGL1,DENND2C,TRIM33,NGF,AMPD1,SIKE1,SYT6,CSDE1,SYCP1,NGF-AS1,NRAS,TSPAN2,BCAS2 |
| 1 | 149716038 | 151146407 | GERD | MDD | 0.876904 | 0.00196203 | positive | SEMA6C,PLEKHO1,ARNT,H4C15,APH1A,H3C13,GABPB2,SCNM1,ADAMTSL4-AS1,BOLA1,FCGR1A,TARS2,SNORD13C,ANXA9,MTMR11,SF3B4,GOLPH3L,H3P4,H2AC21,PRPF3,H4C14,H2BC20P,FALEC,H2BC21,ADAMTSL4,MIR6878,MIR4257,TNFAIP8L2,TMOD4,TNFAIP8L2-SCNM1,MCL1,H2AC18,H3C14,PRUNE1,SV2A,C1orf54,CERS2,LOC105371433,LYSMD1,C1orf56,H2AC19,PDE4DIPP4,CA14,CTSK,OTUD7B,HORMAD1,H2BC18,ANP32E,CIART,VPS45,RPRD2,CTSS,SETDB1,H3C15,ENSA,MLLT11,ECM1,MRPS21,CDC42SE1,MINDY1,CTXND2,BNIPL,H2AC20 |
| 1 | 154685546 | 156813845 | GERD | MDD | 0.507382 | 5.57789E-05 | positive | CKS1B,HDGF,MEF2D,CLK2,ISG20L2,DPM3,RIT1,RXFP4,HAPLN2,MIR4258,KHDC4,SYT11,GBA,MIR555,LMNA,SHC1,NAXE,BCAN,MTX1,YY1AP1,CCT3,SSR2,EFNA4,FDPS,RRNAD1,CRABP2,RUSC1,DCST1,BGLAP,MRPL24,GBAP1,EFNA3,RHBG,RAB25,GPATCH4,SLC50A1,DAP3,LOC101928120,TMEM79,LENEP,PAQR6,TSACC,PMVK,PRCC,MSTO1,MEX3A,THBS3,POU5F1P4,SMG5,KRTCAP2,ZBTB7B,LOC101928177,SCARNA26B,ASH1L-AS1,SLC25A44,GON4L,KCNN3,MIR9-1,MIR9-1HG,PYGO2,TRIM46,SNORA80E,UBQLN4,SEMA4A,TTC24,MSTO2P,VHLL,FAM189B,LAMTOR2,HCN3,MUC1,PKLR,SCARNA4,NES,RUSC1-AS1,DCST1-AS1,GLMP,PMF1,INSRR,SH2D2A,SCAMP3,EFNA1,SCARNA26A,ADAM15,PMF1-BGLAP,MIR92B,DCST2,FLAD1,MIR6738,PBXIP1,ARHGEF2,IQGAP3,NTRK1,ASH1L |
| 1 | 156813846 | 158081627 | GERD | MDD | 0.801522 | 0.000133909 | positive | FCRL2,KIRREL1-IT1,LRRC71,FCRL1,MIR765,ETV3,FCRL5,CYCSP52,INSRR,KIRREL1,FCRL3,LOC105371458,NTRK1,CD5L,FCRL4,ARHGEF11,ETV3L,PEAR1 |
| 1 | 168179087 | 169521551 | GERD | MDD | 0.516907 | 0.00185351 | positive | SFT2D2,LINC00970,CCDC181,TBX19,LINC00626,DPT,XCL1,LOC100505918,MIR557,NME7,F5,BLZF1,ANKRD36BP1,LOC101928565,XCL2,SLC19A2,LOC101928596,ATP1B1 |
| 1 | 173582512 | 175385220 | GERD | MDD | 0.82156 | 2.90799E-05 | positive | RABGAP1L,SNORD44,SNORD47,CACYBP,TEX50,ZBTB37,SERPINC1,KLHL20,ANKRD45,SNORD81,DARS2,SNORD80,RABGAP1L-DT,SNORD74,GAS5-AS1,KIAA0040,SNORA103,SNORD78,MRPS14,SNORD75,CENPL,SNORD76,SNORD77,SNORD79,TNN,GPR52,RABGAP1L-AS1,RC3H1,GAS5,TNR |
| 1 | 176449097 | 177746472 | GERD | MDD | 0.731666 | 0.0019089 | positive | MIR488,BRINP2,LINC01645,LINC01741,ASTN1,PAPPA2 |
| 1 | 182420541 | 183920616 | GERD | MDD | 1 | 0.000374935 | positive | RNASEL,ARPC5,LOC647070,SHCBP1L,COLGALT2,NPL,LAMC1-AS1,LAMC2,LAMC1,NCF2,SMG7,LINC01688,RGSL1,LINC01686,APOBEC4,RGS16,RGS8,SMG7-AS1,DHX9,RGL1,NMNAT2 |
| 1 | 189904638 | 191419275 | GERD | MDD | 0.689814 | 0.00765997 | positive | LINC01720,LINC01680,LINC01351,BRINP3 |
| 1 | 196179111 | 197935821 | GERD | MDD | 0.844352 | 4.05503E-05 | positive | F13B,C1orf53,CFHR5,ZBTB41,ASPM,CFHR1,CFH,MIR4735,KCNT2,CRB1,LHX9,CFHR3,CFHR2,DENND1B,CFHR4 |
| 1 | 197935822 | 199263389 | GERD | MDD | 0.488503 | 0.00848912 | positive | LINC02789,LINC01222,LINC01221,ATP6V1G3,MIR181B1,MIR181A1HG,PTPRC,MIR181A1,NEK7 |
| 1 | 204092538 | 205009623 | GERD | MDD | 0.498088 | 0.00150467 | positive | PIK3C2B,KISS1,MDM4,LINC00628,NFASC,REN,ETNK2,PLEKHA6,PPP1R15B,SOX13,LRRN2,ERLNC1,GOLT1A |
| 1 | 205009624 | 205917548 | GERD | MDD | 0.419095 | 0.0245927 | positive | CNTN2,LEMD1,TMCC2,CDK18,PM20D1,SLC26A9,ELK4,RAB29,LEMD1-AS1,NUCKS1,LEMD1-DT,SLC45A3,LOC284581,SLC41A1,KLHDC8A,MIR135B,DSTYK,BLACAT1,MFSD4A-AS1,RBBP5,MFSD4A,NUAK2,TMEM81 |
| 1 | 208162952 | 209711169 | GERD | MDD | 0.470187 | 0.000599578 | positive | LINC01774,MIR205,LINC01696,PLXNA2,MIR205HG,LINC01698,LINC01717 |
| 1 | 209711170 | 211082892 | GERD | MDD | 0.371239 | 0.028229 | positive | LAMB3,HHAT,SERTAD4,UTP25,HSD11B1-AS1,IRF6,CAMK1G,KCNH1,SYT14,TRAF3IP3,G0S2,C1orf74,MIR4260,HSD11B1,SERTAD4-AS1 |
| 1 | 212347583 | 213958292 | GERD | MDD | 0.48749 | 0.0128769 | positive | ANGEL2,SPATA45,FAM71A,ATF3,NENF,FLVCR1-DT,PPP2R5A,LINC01740,BATF3,LINC02608,FLVCR1,TATDN3,NSL1,VASH2,PACC1,RPS6KC1,SNORA16B |
| 1 | 213958293 | 214783872 | GERD | MDD | 0.573793 | 1.08269E-06 | positive | PTPN14,PROX1-AS1,PROX1,LINC00538,CENPF,SMYD2 |
| 1 | 229075157 | 230227247 | GERD | MDD | 0.63119 | 0.0229234 | positive | URB2,LOC105373159,LINC01682,ACTA1,LINC01736,MIR4454,ABCB10,CCSAP,RAB4A,GALNT2,TAF5L,LOC101927478,NUP133 |
| 1 | 241033585 | 241813534 | GERD | MDD | 0.726713 | 0.000113024 | positive | MIR3123,OPN3,FH,KMO,CHML,RGS7 |
| 1 | 242417924 | 243048003 | GERD | MDD | 0.619492 | 0.00760783 | positive | PLD5 |
| 1 | 243048004 | 244723196 | GERD | MDD | 0.308787 | 0.0127306 | positive | SDCCAG8,LINC01347,CATSPERE,C1orf100,LINC02774,CEP170,ADSS2,ZBTB18,AKT3,MIR4677 |
| 2 | 1875061 | 2680063 | GERD | MDD | 0.685608 | 0.00300009 | positive | MYT1L-AS1,MYT1L |
| 2 | 5568654 | 6306606 | GERD | MDD | 0.693861 | 8.84622E-05 | positive | MIR7158,LINC01810,SOX11,SILC1,LOC400940,LINC01248 |
| 2 | 6306607 | 7108208 | GERD | MDD | 0.839711 | 0.00389855 | positive | LINC01246,LINC00487,NRIR,MIR7515,RSAD2,CMPK2,LINC01247,RNF144A,GRASLND,MIR7515HG |
| 2 | 7966710 | 8978798 | GERD | MDD | 0.63329 | 0.000357617 | positive | LINC00299,LOC101929551,ID2-AS1,LINC01814,ID2,LINC00298,KIDINS220 |
| 2 | 9960671 | 10611654 | GERD | MDD | 0.611328 | 0.0120591 | positive | CYS1,ODC1-DT,MIR4261,TAF1B,LOC101929882,SNORA80B,GRHL1,HPCAL1,ODC1,RRM2,KLF11 |
| 2 | 10611655 | 11230349 | GERD | MDD | 0.847818 | 0.00052311 | positive | ATP6V1C2,KCNF1,PDIA6,NOL10,RN7SL832P,LINC01954 |
| 2 | 12732507 | 13625450 | GERD | MDD | 0.521113 | 0.0177924 | positive | MIR3125,TRIB2,LOC100506474 |
| 2 | 21084899 | 22429640 | GERD | MDD | 0.525345 | 0.00833233 | positive | APOB,TDRD15,LINC01822 |
| 2 | 22429641 | 23538527 | GERD | MDD | 0.839367 | 4.63292E-06 | positive | LINC01884 |
| 2 | 23538528 | 24669224 | GERD | MDD | 0.771151 | 0.000245388 | positive | MFSD2B,ATAD2B,KLHL29,FAM228A,UBXN2A,SF3B6,FAM228B,PFN4,TP53I3,FKBP1B,ITSN2,WDCP |
| 2 | 26894103 | 28819510 | GERD | MDD | 0.365558 | 0.0140964 | positive | PLB1,KRTCAP3,LOC100505716,MAPRE3-AS1,PREB,FOSL2,KCNK3,LINC01460,CGREF1,ZNF512,ATRAID,GTF3C2,SLC30A3,DNAJC5G,FNDC4,IFT172,UCN,ZNF513,ABHD1,MRPL33,CCDC121,CENPA,BABAM2-AS1,FLJ31356,TCF23,TMEM214,OST4,PRR30,MIR4263,DPYSL5,AGBL5-AS1,SUPT7L,LOC105374378,TRIM54,CAD,BABAM2,GTF3C2-AS1,KHK,EIF2B4,SLC35F6,PPM1G,C2orf16,RBKS,SNX17,NRBP1,AGBL5,GPN1,FTH1P3,GCKR,MAPRE3,SLC4A1AP,EMILIN1,SLC5A6,LOC100505736,MPV17 |
| 2 | 36120744 | 36867389 | GERD | MDD | 0.75695 | 0.000509523 | positive | CRIM1-DT,FEZ2,CRIM1 |
| 2 | 36867390 | 37603360 | GERD | MDD | 0.794063 | 0.00647528 | positive | CEBPZOS,STRN,NDUFAF7,VIT,HEATR5B,PRKD3,CEBPZ,SULT6B1,EIF2AK2,QPCT,GPATCH11 |
| 2 | 49114133 | 49849185 | GERD | MDD | 0.741002 | 0.00903456 | positive | FSHR,MIR548BA |
| 2 | 50756241 | 51669493 | GERD | MDD | 0.753418 | 2.09044E-05 | positive | NRXN1,LOC730100,MIR8485 |
| 2 | 52494112 | 53032976 | GERD | MDD | 0.74734 | 0.00316487 | positive | MIR4431,LINC01867,LOC730100 |
| 2 | 57952946 | 59251996 | GERD | MDD | 0.395872 | 0.00805454 | positive | LINC01795,FANCL,VRK2,LINC01122 |
| 2 | 59251997 | 60775066 | GERD | MDD | 0.672554 | 7.48413E-12 | positive | MIR4432HG,BCL11A,LINC01122,LINC01793,MIR4432 |
| 2 | 63672119 | 64696089 | GERD | MDD | 1 | 0.0017312 | positive | WDPCP,MIR4433A,UGP2,PELI1,LGALSL,LOC100507006,MIR4433B,VPS54,LINC00309,MDH1 |
| 2 | 64696090 | 65938002 | GERD | MDD | 0.629964 | 0.0027878 | positive | LOC101927402,LINC02245,SLC1A4,CEP68,LINC02579,LINC01800,SERTAD2,SPRED2,RAB1A,AFTPH,LINC01805,ACTR2,MIR4434 |
| 2 | 65938003 | 67224027 | GERD | MDD | 0.63074 | 9.54431E-05 | positive | LINC01798,MIR4778,LINC01797,MEIS1,LINC01799,MEIS1-AS2,LINC01628,MEIS1-AS3,LINC01873 |
| 2 | 67224028 | 67979828 | GERD | MDD | 0.427093 | 0.0263919 | positive | LINC01829,LINC01828,ETAA1,LOC101927661 |
| 2 | 79900056 | 80737811 | GERD | MDD | 0.400241 | 0.0180327 | positive | CTNNA2,LRRTM1,MIR8080 |
| 2 | 97339302 | 98989360 | GERD | MDD | 0.96668 | 0.000413193 | positive | CNNM3,FAHD2B,ANKRD39,TMEM131,ZAP70,SEMA4C,FAM178B,ANKRD36B,LOC100506076,CNNM4,ANKRD36,C2orf92,CNGA3,CNNM3-DT,LOC100506123,ANKRD23,COX5B,MIR3127,VWA3B,ACTR1B,LMAN2L,FER1L5,LOC101927053 |
| 2 | 98989361 | 100202436 | GERD | MDD | 0.791868 | 0.00118176 | positive | MRPL30,TSGA10,TXNDC9,LYG2,AFF3,INPP4A,LIPT1,MITD1,REV1,EIF5B,LINC02611,MGAT4A,CRACDL,LYG1,CNGA3,UNC50,COA5,C2orf15 |
| 2 | 100202437 | 101556799 | GERD | MDD | 0.573013 | 0.000023139 | positive | AFF3,LINC01104,LINC01849,LONRF2,NMS,CHST10,PDCL3,NPAS2 |
| 2 | 103305258 | 105120578 | GERD | MDD | 0.555591 | 0.00031172 | positive | LINC01831,LOC100287010,SLC9A2,LINC01102,LINC01796,LINC01935,TMEM182,LINC01103,MFSD9 |
| 2 | 112644829 | 113930668 | GERD | MDD | 0.78572 | 0.00218099 | positive | POLR1B,CKAP2L,IL1A,CHCHD5,SLC20A1,RGPD5,FLJ42351,LOC105373562,NT5DC4,IL36B,IL36RN,RGPD8,IL1F10,TMEM87B,ZC3H8,FBLN7,IL36G,MERTK,IL1RN,TTL,IL37,ZC3H6,IL36A,IL1B |
| 2 | 115203836 | 116415816 | GERD | MDD | 0.630046 | 0.00110721 | positive | DPP10-AS3,DPP10-AS1,DPP10 |
| 2 | 124730992 | 125741528 | GERD | MDD | 0.742395 | 0.000039982 | positive | CNTNAP5,LOC107985820 |
| 2 | 125741529 | 126754027 | GERD | MDD | 1 | 1.63679E-05 | positive | NA |
| 2 | 127895645 | 129486897 | GERD | MDD | 0.612435 | 0.00484672 | positive | SAP130,PROC,AMMECR1L,MIR4783,LIMS2,MAP3K2,LOC105373609,GPR17,CYP27C1,IWS1,POLR2D,ERCC3,HS6ST1,UGGT1,MYO7B,WDR33,SFT2D3 |
| 2 | 133352951 | 134359059 | GERD | MDD | 0.673697 | 0.0111287 | positive | NCKAP5-AS1,GPR39,NCKAP5,LYPD1,NCKAP5-AS2 |
| 2 | 138073792 | 139505819 | GERD | MDD | 0.673961 | 0.000733539 | positive | LOC101928273,LINC01832,NXPH2,HNMT,SPOPL,THSD7B |
| 2 | 141802665 | 142494980 | GERD | MDD | 0.707409 | 0.00977703 | positive | LRP1B |
| 2 | 142494981 | 143259780 | GERD | MDD | 0.862067 | 6.30219E-05 | positive | LRP1B |
| 2 | 143259781 | 144503688 | GERD | MDD | 0.814476 | 4.23325E-07 | positive | KYNU,ARHGAP15 |
| 2 | 144503689 | 145977920 | GERD | MDD | 0.608039 | 8.33032E-05 | positive | LINC01412,GTDC1,TEX41,ZEB2-AS1,ZEB2,LOC101928386,LOC105373656,ARHGAP15 |
| 2 | 147252197 | 149568875 | GERD | MDD | 0.491306 | 0.00170317 | positive | MBD5,EPC2,PABPC1P2,ORC4,ACVR2A |
| 2 | 149568876 | 151428988 | GERD | MDD | 0.473506 | 0.0240529 | positive | LYPD6B,MIR9899,LINC01931,LINC01817,MMADHC-DT,KIF5C,RND3,LYPD6,LINC01920,MMADHC |
| 2 | 153999310 | 155207969 | GERD | MDD | 0.87334 | 1.24069E-05 | positive | RPRM,GALNT13 |
| 2 | 155207970 | 156130947 | GERD | MDD | 0.61288 | 0.00188524 | positive | KCNJ3,LOC100144595,GALNT13 |
| 2 | 157556097 | 159040979 | GERD | MDD | 0.694618 | 0.00286341 | positive | ACVR1C,CCDC148-AS1,FAM133DP,CCDC148,UPP2,ACVR1,GALNT5,CYTIP,ERMN |
| 2 | 159040980 | 160200379 | GERD | MDD | 0.676258 | 0.0031993 | positive | WDSUB1,DAPL1,CCDC148-AS1,MIR6888,CCDC148,TANC1,PKP4,PKP4-AS1,BAZ2B |
| 2 | 160200380 | 161432089 | GERD | MDD | 0.456208 | 0.00712603 | positive | LINC02478,LY75-CD302,CD302,RBMS1,LY75,MARCHF7,MIR4785,BAZ2B,ITGB6,PLA2R1,LOC643072 |
| 2 | 161432090 | 163607635 | GERD | MDD | 0.53621 | 0.000753526 | positive | LOC101929512,LOC101929532,KCNH7,TANK,FAP,DPP4-DT,IFIH1,GCG,LINC01806,AHCTF1P1,PSMD14,SLC4A10,TBR1,DPP4,GCA |
| 2 | 172501316 | 173287857 | GERD | MDD | 0.569033 | 0.0138654 | positive | SLC25A12,DYNC1I2,DLX2,DLX2-DT,DLX1,HAT1,METAP1D |
| 2 | 173287858 | 174118389 | GERD | MDD | 0.895053 | 7.16301E-08 | positive | ITGA6,RAPGEF4,RAPGEF4-AS1,MAP3K20-AS1,ITGA6-AS1,PDK1,MAP3K20 |
| 2 | 174118390 | 174927563 | GERD | MDD | 0.672866 | 2.48597E-05 | positive | CDCA7,SP3,MAP3K20-AS1,LINC01960,MAP3K20 |
| 2 | 174927564 | 176734750 | GERD | MDD | 0.417766 | 0.00214389 | positive | GPR155,LINC01305,RNU6-1,H3P6,CIR1,OLA1,RNU6-7,RNU6-2,CHN1,RNU6-9,ATF2,SCRN3,RNU6-8,SP9,ATP5MC3,WIPF1,MIR933,CHRNA1 |
| 2 | 193033983 | 195300250 | GERD | MDD | 0.720089 | 0.000332296 | positive | PCGEM1,LINC01821,TMEFF2 |
| 2 | 199654970 | 201324132 | GERD | MDD | 0.778786 | 0.00148699 | positive | C2orf69,SPATS2L,LINC01877,SATB2-AS1,MAIP1,FTCDNL1,SATB2,TYW5 |
| 2 | 205038120 | 206821199 | GERD | MDD | 0.463752 | 0.000327985 | positive | PARD3B,NRP2 |
| 2 | 207726595 | 208674588 | GERD | MDD | 0.576023 | 0.00272298 | positive | CREB1,LINC01857,CCNYL1,MYOSLID-AS1,MYOSLID,MIR4775,KLF7,LINC01802,MIR1302-4,METTL21A,MIR2355,CPO,FZD5,MIR7845 |
| 2 | 208674589 | 209940230 | GERD | MDD | 0.717326 | 0.000196632 | positive | CRYGD,PLEKHM3,PTH2R,IDH1,C2orf80,IDH1-AS1,CRYGA,PIKFYVE,LOC100507443,CRYGB,LOC101927960,CRYGC |
| 2 | 210842458 | 212520453 | GERD | MDD | 0.458869 | 0.00980678 | positive | UNC80,KANSL1L,KANSL1L-AS1,CPS1-IT1,RPE,ERBB4,LANCL1-AS1,LANCL1,MYL1,CPS1,ACADL |
| 2 | 212520454 | 213194362 | GERD | MDD | 0.692902 | 0.000373599 | positive | ERBB4 |
| 2 | 215899571 | 217566011 | GERD | MDD | 0.404773 | 0.00827961 | positive | XRCC5,LINC00607,SMARCAL1,IGFBP5,RPL37A,LINC01963,TMEM169,LOC102724849,FN1,MREG,LINC01280,ABCA12,LINC01614,ATIC,MARCHF4,IGFBP2,PECR |
| 2 | 227557842 | 228812485 | GERD | MDD | 0.880137 | 3.17507E-05 | positive | MIR5703,SLC19A3,C2orf83,MFF,AGFG1,RHBDD1,CCL20,LOC729968,IRS1,DAW1,TM4SF20,COL4A4,COL4A3,MFF-DT |
| 2 | 234945578 | 235625789 | GERD | MDD | 0.567727 | 0.00729274 | positive | LINC01891,ARL4C,SPP2,LINC01173 |
| 2 | 239634351 | 240325600 | GERD | MDD | 0.468662 | 0.00749808 | positive | LOC100287387,MIR4440,LINC01940,TWIST2,HDAC4,MIR4269,MIR4441,MIR2467,MGC16025,HDAC4-AS1 |
| 2 | 240325601 | 241113279 | GERD | MDD | 0.491337 | 0.0122462 | positive | OR6B2,NDUFA10,OTOS,LOC150935,OR6B3,MIR4786,COPS9 |
| 3 | 2140012 | 3128152 | GERD | MDD | 0.868634 | 2.85591E-10 | positive | IL5RA,CNTN4-AS1,CNTN4,CNTN4-AS2 |
| 3 | 3128153 | 4109393 | GERD | MDD | 0.777189 | 1.03863E-06 | positive | LOC100130207,IL5RA,TRNT1,LRRN1,CRBN |
| 3 | 5496183 | 6421145 | GERD | MDD | 0.558954 | 0.00956505 | positive | NA |
| 3 | 16084006 | 17182494 | GERD | MDD | 0.562786 | 0.000360964 | positive | MIR3714,LINC00690,OXNAD1,PLCL2,RFTN1,GALNT15,DAZL,DPH3 |
| 3 | 17182495 | 19030988 | GERD | MDD | 0.395125 | 0.027685 | positive | SATB1-AS1,TBC1D5,LOC339862,SATB1,LOC105376975 |
| 3 | 20754843 | 21790867 | GERD | MDD | 0.701332 | 0.000395853 | positive | ZNF385D,VENTXP7,ZNF385D-AS1 |
| 3 | 21790868 | 22648001 | GERD | MDD | 0.858145 | 4.42882E-05 | positive | ZNF385D,ZNF385D-AS2 |
| 3 | 23426045 | 24270189 | GERD | MDD | 0.491659 | 0.00311618 | positive | RPL15,UBE2E1-AS1,THRB-AS2,UBE2E2,LINC00691,THRB,NKIRAS1,UBE2E1,NR1D2 |
| 3 | 28649811 | 29877022 | GERD | MDD | 0.586016 | 0.000234059 | positive | RBMS3,LINC00693,RBMS3-AS3 |
| 3 | 35009560 | 35913033 | GERD | MDD | 0.705595 | 0.0026266 | positive | LOC101928135,ARPP21,MIR128-2 |
| 3 | 35913034 | 36840136 | GERD | MDD | 0.675294 | 0.000444836 | positive | STAC,DCLK3 |
| 3 | 36840137 | 38729767 | GERD | MDD | 0.54177 | 0.000456705 | positive | ITGA9,MIR26A1,ACVR2B,SLC22A14,C3orf35,MYD88,LINC02033,VILL,EXOG,PLCD1,SLC22A13,SCN5A,TRANK1,XYLB,LOC152048,MLH1,UBE2FP1,ACAA1,OXSR1,GOLGA4,ITGA9-AS1,LRRFIP2,DLEC1,CTDSPL,ACVR2B-AS1,EPM2AIP1 |
| 3 | 42301658 | 44141693 | GERD | MDD | 0.498615 | 0.00140569 | positive | HHATL-AS1,HIGD1A,VIPR1-AS1,CYP8B1,ANO10,NKTR,CCK,LYZL4,GASK1A,CCDC13,ACKR2,SNRK-AS1,KLHL40,KRBOX1,ABHD5,POMGNT2,SEC22C,LINC02158,HHATL,ZBTB47,LOC101928323,KRBOX1-AS1,CCDC13-AS1,SS18L2,ZNF662,SNRK,VIPR1 |
| 3 | 44141694 | 45844191 | GERD | MDD | 0.543838 | 0.000470813 | positive | ZNF502,KIF15,KIAA1143,EXOSC7,ZNF445,TMEM42,LIMD1-AS1,C3orf86,LARS2-AS1,TMEM158,TGM4,ZNF501,ZNF197,TCAIM,LINC01988,LIMD1,ZNF852,CDCP1,ZNF197-AS1,ZNF35,MIR564,SLC6A20,TOPAZ1,SACM1L,ZKSCAN7,LARS2,ZDHHC3,ZNF660-ZNF197,MIR138-1,ZKSCAN7-AS1,ZNF660,CLEC3B |
| 3 | 47588462 | 50387742 | GERD | MDD | 0.815898 | 3.4732E-08 | positive | APEH,GNAI2,MAP4,SLC26A6,SLC25A20,MIR4443,LSMEM2,C3orf84,NPRL2,NCKIPSD,SEMA3B-AS1,SNORA94,MIR191,ZNF589,MIR5787,MIR6824,GPX1,TREX1,DALRD3,PLXNB1,CCDC51,NDUFAF3,LINC02585,RBM6,C3orf62,TMEM89,MIR5193,HYAL1,FCF1P2,CDC25A,RHOA,UCN2,ARIH2OS,RBM5,MIR4271,HYAL3,CAMP,SEMA3B,QARS1,CELSR3,QRICH1,RASSF1-AS1,NICN1,IFRD2,IMPDH2,CDHR4,NAA80,USP4,ATRIP,SHISA5,NME6,ARIH2,DHX30,SEMA3F,GNAT1,TMA7,KLHDC8B,MIR2115,MIR6890,SPINK8,IP6K2,COL7A1,RNF123,RBM5-AS1,SMARCC1,MIR4793,CCDC71,GMPPB,IP6K1,WDR6,RASSF1,UQCRC1,IHO1,UBA7,BSN-DT,TUSC2,INKA1,LAMB2,ATRIP-TREX1,PRKAR2A,MIR711,FBXW12,MON1A,TCTA,CSPG5,MIR1226,MIR425,MIR6872,MIR6823,MST1,P4HTM,BSN,SLC38A3,HYAL2,DAG1,CAMKV,AMT,ZMYND10,PRKAR2A-AS1,TRAIP,USP19,SNORD146,LAMB2P1,MST1R,PFKFB4,SEMA3F-AS1,AMIGO3 |
| 3 | 50387743 | 51953968 | GERD | MDD | 0.713651 | 0.00368596 | positive | CYB561D2,IQCF3,MIR4787,HEMK1,IQCF6,GRM2,TEX264,IQCF5,RAD54L2,NPRL2,IQCF4P,DOCK3,RBM15B,DCAF1,CISH,CACNA2D2,IQCF1,IQCF2,MAPKAPK3,C3orf18,MANF,TMEM115,IQCF5-AS1 |
| 3 | 51953969 | 54074844 | GERD | MDD | 0.586017 | 0.000109139 | positive | STAB1,PHF7,NISCH,MIRLET7G,MIR8064,TNNC1,DNAH1,DUSP7,PARP3,LOC101929054,SPCS1,SEMA3G,POC1A,MUSTN1,SNORD136,NT5DC2,BAP1,PBRM1,CACNA1D,GLT8D1,RPL29,ABHD14B,SNORD19C,GLYCTK,CHDH,PCBP4,ABHD14A-ACY1,ACTR8,ITIH4,ABHD14A,RRP9,STIMATE,WDR82,ALAS1,SNORD38C,PRKCD,ITIH1,RFT1,NEK4,ITIH3,GLYCTK-AS1,DCP1A,SFMBT1,TKT,IL17RB,ACY1,SNORD19,GNL3,SNORD69,ITIH4-AS1,TLR9,STIMATE-MUSTN1,SELENOK,SMIM4,MIR135A1,GPR62,TWF2,LINC00696,PPM1M,SNORD19B |
| 3 | 55616626 | 56954878 | GERD | MDD | 0.501289 | 0.0280507 | positive | CCDC66,ARHGEF3,TASOR,ERC2,MIR3938,ERC2-IT1 |
| 3 | 60571140 | 61277587 | GERD | MDD | 0.949499 | 0.000221728 | positive | FHIT,MIR548BB |
| 3 | 61277588 | 62609818 | GERD | MDD | 0.377355 | 0.00167838 | positive | CADPS,C3orf14,FEZF2,PTPRG-AS1,PTPRG |
| 3 | 63670804 | 64662373 | GERD | MDD | 0.510497 | 0.0112602 | positive | PRICKLE2,ADAMTS9-AS1,C3orf49,THOC7-AS1,THOC7,ATXN7,PSMD6,SCAANT1,PSMD6-AS2,PRICKLE2-AS1,ADAMTS9,PRICKLE2-AS2,PRICKLE2-AS3,LINC00994 |
| 3 | 64662374 | 65326751 | GERD | MDD | 0.805275 | 0.000177787 | positive | ADAMTS9,ADAMTS9-AS2 |
| 3 | 65326752 | 66702084 | GERD | MDD | 0.550825 | 9.48857E-05 | positive | MAGI1-AS1,MAGI1,MAGI1-IT1,SLC25A26,LRIG1 |
| 3 | 68319771 | 69784969 | GERD | MDD | 0.387364 | 0.0122108 | positive | EOGT,UBA3,TAFA4,MIR3136,LMOD3,ARL6IP5,TMF1,FRMD4B,LOC105377146,TAFA1 |
| 3 | 71223282 | 72334704 | GERD | MDD | 0.567188 | 1.23625E-05 | positive | LINC00877,FOXP1,MIR1284,LINC00870,EIF4E3,GPR27,PROK2,FOXP1-AS1 |
| 3 | 76157471 | 77507832 | GERD | MDD | 0.489646 | 0.00189636 | positive | ROBO2 |
| 3 | 83529395 | 84698480 | GERD | MDD | 0.782919 | 0.0232321 | positive | LINC00971 |
| 3 | 89612940 | 94250144 | GERD | MDD | 0.726188 | 0.00230655 | positive | PROS1,STX19,NSUN3,DHFR2,ARL13B |
| 3 | 106174686 | 107736519 | GERD | MDD | 0.499577 | 0.00741113 | positive | LOC101929579,LINC00636,CCDC54,LINC01990,DUBR,LINC00635,LINC00882,BBX |
| 3 | 112408514 | 113657665 | GERD | MDD | 0.664441 | 0.00773055 | positive | SPICE1,NAA50,USF3,CD200R1,LINC02044,MIR9900,NEPRO,GTPBP8,CFAP44-AS1,CFAP44,ATP6V1A,BOC,CD200R1L-AS1,LINC02042,MIR8076,GRAMD1C,MIR4446,SIDT1,CD200R1L |
| 3 | 115649910 | 116498570 | GERD | MDD | 0.483969 | 0.0211236 | positive | LSAMP-AS1,LSAMP,LINC00903,TUSC7 |
| 3 | 116498571 | 117241644 | GERD | MDD | 0.468375 | 0.0216187 | positive | MIR4447,LINC00901 |
| 3 | 117241645 | 118086929 | GERD | MDD | 0.739303 | 0.000591263 | positive | LINC02024 |
| 3 | 125992651 | 127495173 | GERD | MDD | 0.450789 | 0.0159098 | positive | ABTB1,CFAP100,LINC01471,CHST13,TPRA1,LINC02016,KLF15,MCM2,NUP210P1,MGLL,MIR7976,MIR6825,UROC1,PLXNA1,PODXL2,C3orf56,ZXDC,CCDC37-DT,CHCHD6,C3orf22,TXNRD3,PRR20G |
| 3 | 135288395 | 137372141 | GERD | MDD | 0.78937 | 0.0106805 | positive | NCK1-DT,STAG1,PCCB,NCK1,SLC35G2,IL20RB,MSL2,PPP2R3A |
| 3 | 141898979 | 142976707 | GERD | MDD | 0.499519 | 0.0148629 | positive | U2SURP,PAQR9,PAQR9-AS1,TRPC1,ATR,PCOLCE2,LOC100289361,LOC100507389,XRN1,CHST2,PLS1,GK5 |
| 3 | 142976708 | 144408821 | GERD | MDD | 0.596346 | 0.00228785 | positive | SLC9A9-AS1,DIPK2A,SLC9A9 |
| 3 | 145903839 | 147358243 | GERD | MDD | 0.625013 | 0.00576847 | positive | ZIC4,PLSCR4,PLSCR1,PLSCR2,LOC440982,PLSCR5,ZIC1,LINC02010 |
| 3 | 147358244 | 148638308 | GERD | MDD | 0.50977 | 0.00130694 | positive | AGTR1,CPA3,CPB1,LINC02046,LINC02032 |
| 3 | 149998412 | 151131307 | GERD | MDD | 1 | 1.69332E-05 | positive | P2RY12,TSC22D2,EIF2A,MINDY4B,CLRN1-AS1,MED12L,LINC01214,ERICH6-AS1,GPR171,ERICH6,LOC101928105,P2RY13,SERP1,P2RY14,GPR87,CLRN1,SIAH2,SELENOT |
| 3 | 167118894 | 168972662 | GERD | MDD | 0.69242 | 1.19996E-05 | positive | GOLIM4,EGFEM1P,LRRC77P,LINC02082,ZBBX,WDR49,PDCD10,SERPINI2,SERPINI1,MECOM,LINC01327,MIR551B |
| 3 | 172004761 | 173415591 | GERD | MDD | 0.637462 | 0.000155106 | positive | FNDC3B,LINC02068,ECT2,SPATA16,TNFSF10,GHSR,NLGN1,NCEH1 |
| 3 | 173415592 | 174684707 | GERD | MDD | 0.479033 | 0.0160292 | positive | NAALADL2,NLGN1,NLGN1-AS1 |
| 3 | 174684708 | 175671424 | GERD | MDD | 0.667744 | 0.00242541 | positive | NAALADL2-AS1,NAALADL2-AS2,MIR4789,NAALADL2,NAALADL2-AS3 |
| 3 | 176931164 | 178110322 | GERD | MDD | 1 | 8.56172E-06 | positive | LINC02015,LINC00578,LINC00501 |
| 3 | 179372166 | 181424323 | GERD | MDD | 0.716597 | 4.9802E-06 | positive | FXR1,LINC02053,PEX5L-AS2,LOC101928882,RNU6-1,DNAJC19,LOC102724604,RNU6-7,TTC14,RNU6-9,RNU6-2,USP13,CCDC39,RNU6-8,PEX5L,SOX2-OT |
| 3 | 191229358 | 192049688 | GERD | MDD | 0.522066 | 0.0202905 | positive | FGF12,LINCR-0002,FGF12-AS1 |
| 3 | 193122091 | 194217140 | GERD | MDD | 0.511827 | 0.00988223 | positive | CPN2,LINC02036,OPA1,ATP13A4-AS1,ATP13A4,HES1,GP5,LINC00884,ATP13A3,LINC02037,LINC02038,LRRC15,LINC00887,OPA1-AS1,LINC02026,LINC02028,DPPA2P3 |
| 4 | 2468936 | 3549229 | GERD | MDD | 0.623078 | 0.0171311 | positive | NOP14-AS1,RNF4,ADD1,HGFAC,TNIP2,HTT,LRPAP1,HTT-AS,DOK7,SH3BP2,MFSD10,NOP14,RGS12,FAM193A,MSANTD1,GRK4 |
| 4 | 5051835 | 5972709 | GERD | MDD | 0.599602 | 0.00351101 | positive | EVC,MIR378D1,C4orf50,LINC01587,STK32B,EVC2,CRMP1 |
| 4 | 11050120 | 12209028 | GERD | MDD | 0.949153 | 0.000834646 | positive | LINC02360,MIR12113,HS3ST1,MIR572 |
| 4 | 13127577 | 14395886 | GERD | MDD | 0.604683 | 0.0101735 | positive | LINC01096,LINC01085,LINC01097,RAB28,BOD1L1,LINC01182,NKX3-2,MIR5091 |
| 4 | 15910802 | 17290236 | GERD | MDD | 0.439143 | 0.00727653 | positive | TAPT1,FGFBP1,FGFBP2,LDB2,PROM1,TAPT1-AS1,LINC02493 |
| 4 | 17290237 | 18753263 | GERD | MDD | 0.74793 | 0.000333026 | positive | FAM184B,CLRN2,QDPR,DCAF16,SNORA75B,NCAPG,LAP3,MED28,LCORL |
| 4 | 18753264 | 20086838 | GERD | MDD | 0.629531 | 0.0110628 | positive | NA |
| 4 | 21416381 | 22835185 | GERD | MDD | 0.794619 | 0.0131685 | positive | LOC105374516,GBA3,KCNIP4,LOC100505912,MIR12115,KCNIP4-IT1,ADGRA3,MIR7978 |
| 4 | 22835186 | 23858350 | GERD | MDD | 0.632018 | 0.00118682 | positive | PPARGC1A |
| 4 | 28177793 | 28927438 | GERD | MDD | 0.802051 | 0.0114521 | positive | MIR4275 |
| 4 | 34721846 | 35411236 | GERD | MDD | 0.784618 | 0.000866408 | positive | NA |
| 4 | 37880861 | 38984838 | GERD | MDD | 0.600404 | 5.29104E-05 | positive | TBC1D1,KLF3,PTTG2,TLR10,TLR6,MIR574,LINC01259,FAM114A1,TLR1,KLF3-AS1,LINC02278,LINC01258,TMEM156,LINC02513 |
| 4 | 45186768 | 46358360 | GERD | MDD | 0.915167 | 0.00549085 | positive | GABRG1,GABRA2 |
| 4 | 46358361 | 47408265 | GERD | MDD | 0.713648 | 0.00468355 | positive | COX7B2,GABRB1,GABRA2,GABRA4 |
| 4 | 60746710 | 61971161 | GERD | MDD | 0.969566 | 5.14919E-07 | positive | MIR548AG1 |
| 4 | 61971162 | 63337954 | GERD | MDD | 0.826576 | 1.52902E-06 | positive | ADGRL3,LINC02271,ADGRL3-AS1 |
| 4 | 66597447 | 67616362 | GERD | MDD | 0.566227 | 0.00761218 | positive | MIR1269A |
| 4 | 101132996 | 102544803 | GERD | MDD | 0.349644 | 0.0167934 | positive | PPP3CA,LOC101929353,FLJ20021,SNORA101A,MIR1255A,MIR8066,LINC01216,EMCN |
| 4 | 102544804 | 104384534 | GERD | MDD | 0.514636 | 0.0130523 | positive | LOC102723704,LINC02428,SLC9B1,NFKB1,BDH2,CISD2,UBE2D3-AS1,SLC9B2,MANBA,BANK1,UBE2D3,CENPE,LOC105377621,SLC39A8 |
| 4 | 113916003 | 115306149 | GERD | MDD | 0.611142 | 0.0190439 | positive | MIR1243,MIR8082,CAMK2D,ANK2,ARSJ |
| 4 | 118395263 | 119138097 | GERD | MDD | 0.788338 | 0.00104145 | positive | NDST3,LINC01378,LINC02264 |
| 4 | 122655435 | 123768693 | GERD | MDD | 0.765831 | 0.00337749 | positive | IL2,ADAD1,EXOSC9,PP12613,IL21,FGF2,BBS12,SMIM43,TRPC3,BBS7,CCNA2,CETN4P,KIAA1109,IL21-AS1 |
| 4 | 135809543 | 137400031 | GERD | MDD | 0.495228 | 0.0219547 | positive | LINC02485,LINC00613 |
| 4 | 137400032 | 138537344 | GERD | MDD | 0.57271 | 0.000265449 | positive | LINC02172,LINC02511,LINC02510,PCDH18 |
| 4 | 139553761 | 141087047 | GERD | MDD | 0.596143 | 5.13658E-06 | positive | NDUFC1,LOC105377448,NOCT,RAB33B-AS1,MGARP,SETD7,NAA15,MGST2,MAML3,RAB33B,ELF2,LOC101927516 |
| 4 | 153520884 | 155044409 | GERD | MDD | -0.632456 | 0.0115209 | negative | TMEM154,TLR2,TIGD4,FHDC1,TRIM2,ARFIP1,RNF175,SFRP2,ANXA2P1,TMEM131L,MND1,LOC100419170,LOC729870 |
| 4 | 155044410 | 156520288 | GERD | MDD | 0.504193 | 0.00733066 | positive | FGG,PLRG1,FGA,LRAT,MAP9,FGB,NPY2R,LOC102724776,DCHS2,RBM46 |
| 4 | 157597310 | 159176073 | GERD | MDD | 0.624706 | 0.00555224 | positive | LINC02433,PDGFC,GRIA2,TMEM144,FAM198B-AS1,GASK1B,GLRB |
| 4 | 159176074 | 160828480 | GERD | MDD | 0.763839 | 0.00283824 | positive | MIR3688-2,C4orf45,FNIP2,RXFP1,PPID,RAPGEF2,LINC02233,TMEM144,C4orf46,ETFDH,MIR3688-1 |
| 4 | 163937104 | 164781847 | GERD | MDD | 0.76999 | 0.000529948 | positive | TMA16,NAF1,TKTL2,MARCHF1,MIR4454,NPY5R,NPY1R |
| 4 | 175959698 | 177129678 | GERD | MDD | 0.458417 | 0.0138474 | positive | SPATA4,GPM6A,LOC101928590,WDR17 |
| 4 | 177129679 | 178314527 | GERD | MDD | 0.813411 | 0.00182298 | positive | VEGFC,LINC02509,ASB5,NEIL3,SPCS3 |
| 4 | 181122434 | 182065646 | GERD | MDD | 0.715194 | 0.000864145 | positive | LINC00290 |
| 4 | 184338585 | 185483262 | GERD | MDD | 0.689215 | 0.00377795 | positive | CDKN2AIP,RWDD4,ING2,LINC02363,LINC02362,TRAPPC11,ENPP6,LOC389247,IRF2,STOX2,LINC02427 |
| 5 | 1206610 | 2106885 | GERD | MDD | 0.56448 | 0.000949115 | positive | NDUFS6,IRX4,LINC01511,MIR4457,TERT,LPCAT1,MIR6075,MIR4277,SLC6A18,CLPTM1L,SDHAP3,LINC02116,LOC728613,CTD-2194D22.4,MRPL36,SLC6A19,SLC6A3 |
| 5 | 6936734 | 8079415 | GERD | MDD | 0.581028 | 0.000138192 | positive | ADCY2,C5orf49,MTRR,FASTKD3,MIR4454,LINC02123,LOC442132 |
| 5 | 18791411 | 20397013 | GERD | MDD | 0.634325 | 0.00538914 | positive | CDH18-AS1,CDH18 |
| 5 | 24362662 | 25722097 | GERD | MDD | 0.691929 | 6.44992E-05 | positive | LINC02239,LINC02211,LINC02228,CDH10 |
| 5 | 26938179 | 28315653 | GERD | MDD | 0.723424 | 1.64136E-05 | positive | CDH9,LINC02103,PURPL |
| 5 | 36431968 | 37925487 | GERD | MDD | 0.662785 | 0.00548385 | positive | CPLANE1,SLC1A3,LOC105374727,WDR70,GDNF,NUP155,NIPBL,GDNF-AS1,NIPBL-DT |
| 5 | 44049879 | 45305566 | GERD | MDD | 0.610648 | 0.000713917 | positive | LINC02224,FGF10,FGF10-AS1,HCN1,MRPS30,MRPS30-DT |
| 5 | 50164836 | 51203265 | GERD | MDD | 0.89031 | 9.07713E-05 | positive | ISL1,LOC642366,LINC02106 |
| 5 | 51203266 | 52139773 | GERD | MDD | 0.69342 | 8.55296E-05 | positive | ITGA1,LINC02118,PELO |
| 5 | 52139774 | 52837225 | GERD | MDD | 0.844902 | 0.00088347 | positive | ITGA2,LOC257396,ITGA1,MOCS2,FST |
| 5 | 60930754 | 62180368 | GERD | MDD | 0.930579 | 1.24687E-07 | positive | LRRC70,C5orf64,IPO11,LOC101928651,DIMT1,IPO11-LRRC70,C5orf64-AS1,KIF2A |
| 5 | 63214090 | 64438518 | GERD | MDD | 0.814965 | 0.000299657 | positive | CWC27,SHISAL2B,SREK1IP1,RGS7BP,HTR1A,RNF180 |
| 5 | 71468652 | 73314061 | GERD | MDD | 0.405458 | 0.0102915 | positive | LOC105379030,MAP1B,LOC102503427,FCHO2,LOC340090,TMEM174,ARHGEF28,BTF3,TNPO1,LINC01386,MRPS27,ZNF366,FOXD1,MIR4804,LINC02056,PTCD2,UTP15,ANKRA2,LINC02230,TMEM171 |
| 5 | 73314062 | 74245354 | GERD | MDD | 0.680318 | 0.0137825 | positive | LINC01335,LINC01333,ENC1,HEXB,GFM2,NSA2,LOC441086,FAM169A,LINC01331 |
| 5 | 82720249 | 84094797 | GERD | MDD | 0.678127 | 2.90565E-05 | positive | EDIL3,HAPLN1,VCAN-AS1,VCAN |
| 5 | 91956906 | 93814604 | GERD | MDD | 0.59214 | 0.000335833 | positive | NR2F1-AS1,POU5F2,KIAA0825,FAM172A,MIR2277,NR2F1 |
| 5 | 97316602 | 98567769 | GERD | MDD | 0.53722 | 0.0130464 | positive | LOC100289230,RGMB,RGMB-AS1,CHD1,LINC02062,LINC01846 |
| 5 | 102903986 | 103788460 | GERD | MDD | 0.757543 | 1.77433E-06 | positive | NA |
| 5 | 103788461 | 104850490 | GERD | MDD | 0.750995 | 0.000320689 | positive | RAB9BP1 |
| 5 | 104850491 | 106389388 | GERD | MDD | 0.456503 | 0.00683838 | positive | LINC01950 |
| 5 | 106389389 | 107265911 | GERD | MDD | 0.333257 | 0.0173434 | positive | FBXL17,EFNA5 |
| 5 | 116208001 | 117348497 | GERD | MDD | 0.929444 | 4.29819E-05 | positive | LINC00992,LINC02147 |
| 5 | 119664174 | 120371857 | GERD | MDD | 1 | 1.48339E-06 | positive | PRR16 |
| 5 | 122005292 | 123191941 | GERD | MDD | 0.694757 | 0.000680213 | positive | PPIC,CSNK1G3,SNX24,PRDM6,SNX2,LINC02201,CEP120,LOC105379152 |
| 5 | 123191942 | 124109485 | GERD | MDD | 0.645348 | 0.00219846 | positive | ZNF608,LINC01170 |
| 5 | 124109486 | 125131720 | GERD | MDD | 0.619776 | 8.0571E-08 | positive | LOC101927421,LINC02240 |
| 5 | 132554691 | 134556570 | GERD | MDD | 0.517707 | 7.99843E-07 | positive | MIR3661,UBE2B,LINC01843,MIR1289-2,PITX1,SEC24A,WSPAR,DDX46,CAMLG,SKP1,JADE2,LOC102546229,C5orf66-AS1,LOC105379183,CDKL3,CATSPER3,TCF7,VDAC1,CDKN2AIPNL,C5orf24,C5orf66,SAR1B,FSTL4,PPP2CA,C5orf15,PCBD2,TXNDC15 |
| 5 | 135344944 | 136949853 | GERD | MDD | 0.472677 | 0.00400088 | positive | TGFBI,SMAD5-AS1,LOC105379192,SMAD5,VTRNA2-1,SMIM32,MIR4454,SPOCK1,TRPC7-AS2,TRPC7 |
| 5 | 139408117 | 141253830 | GERD | MDD | 0.482359 | 0.00188433 | positive | PCDHGB8P,PCDHA1,PCDH1,PCDHGB2,HBEGF,PCDHGC3,PCDHGA10,PCDHGA12,IK,DIAPH1-AS1,RELL2,PFDN1,MIR6831,HDAC3,PCDHA4,LOC112267934,PCDHB13,CD14,VTRNA1-2,HARS2,PCDHA11,IGIP,PCDHA7,PCDHB7,PURA,PCDHGA11,SLC25A2,PCDHB16,PCDHA10,PCDHA6,PCDHGC5,PCDHB19P,PCDHGC4,PCDHB17P,FCHSD1,PCDHA2,ZMAT2,ANKHD1,PCDHA12,HARS1,TMCO6,ARAP3,NRG2,VTRNA1-1,PCDHA8,EIF4EBP3,PCDHAC2,PCDHB15,PCDHGB7,PCDHB11,PCDHGA5,PCDHB18P,SLC4A9,LOC101926905,PCDHGA3,WDR55,LOC101929719,VTRNA1-3,NDUFA2,CYSTM1,PCDHB12,PCDHA9,PCDHGA9,PCDHB9,PCDHAC1,PCDHB3,SRA1,DIAPH1,ANKHD1-EIF4EBP3,PCDHB1,PCDHB8,PCDHGB5,DND1,PCDHGA8,PCDHGA7,PCDHB6,PCDHB5,TAF7,PCDHA5,PCDHA13,PCDHGA6,MIR3655,PCDHB14,PCDHGB6,PCDHGA2,PCDHGA1,PCDHGB4,PCDHGA4,APBB3,PCDHB10,PCDHGB1,PCDHA3,SLC35A4,PCDHGB3,MALINC1,PCDHB2,PCDHB4 |
| 5 | 141253831 | 142532956 | GERD | MDD | 0.397142 | 0.0270712 | positive | SPRY4,ARHGAP26-AS1,ARHGAP26,PCDH1,PCDH12,RNF14,NDFIP1,LOC729080,SPRY4-IT1,SPRY4-AS1,DELE1,FGF1,LINC01844,GNPDA1 |
| 5 | 145319097 | 147180171 | GERD | MDD | 0.682766 | 0.0004783 | positive | LARS1,PPP2R2B,PLAC8L1,TCERG1,STK32A,RBM27,JAKMIP2-AS1,JAKMIP2,GPR151,PPP2R2B-IT1,SH3RF2,DPYSL3,POU4F3,STK32A-AS1 |
| 5 | 152878628 | 153734494 | GERD | MDD | 0.553265 | 0.00657181 | positive | GRIA1,MFAP3,LINC01861,FAM114A2,MIR1294,GALNT10 |
| 5 | 156345452 | 157191081 | GERD | MDD | 0.919447 | 0.000794795 | positive | LOC102724404,NIPAL4,CYFIP2,FNDC9,MED7,SOX30,THG1L,HAVCR2,C5orf52,HAVCR1,LSM11,ADAM19,TIMD4,ITK,FAM71B |
| 5 | 162386028 | 163781268 | GERD | MDD | 0.409688 | 0.00246349 | positive | MAT2B,HMMR-AS1,CCNG1,NUDCD2,HMMR |
| 5 | 163781269 | 165289211 | GERD | MDD | 0.316487 | 0.0259993 | positive | LINC02143,LOC102546299 |
| 5 | 165289212 | 166863413 | GERD | MDD | 0.503493 | 0.000109565 | positive | LOC101927908,LINC01947,TENM2 |
| 5 | 166863414 | 168342743 | GERD | MDD | 0.46666 | 0.000733908 | positive | MIR218-2,MIR103B1,PANK3,RARS1,SLIT3,FBLL1,TENM2,WWC1,CTB-178M22.2,MIR12125,MIR103A1,SLIT3-AS2 |
| 5 | 168342744 | 169506966 | GERD | MDD | 0.424599 | 0.0175202 | positive | SPDL1,DOCK2,MIR585,SLIT3,SLIT3-AS1,INSYN2B,MIR378E |
| 5 | 169506967 | 170898340 | GERD | MDD | 0.438246 | 0.00161231 | positive | MIR3912,DOCK2,RANBP17,LINC01366,KCNIP1-AS1,KCNIP1-OT1,KCNIP1,LOC100128059,FOXI1,KCNMB1,LINC01187,SNORA70J,GABRP,TLX3,NPM1,LCP2,C5orf58,FGF18 |
| 5 | 170898341 | 172285682 | GERD | MDD | 0.659039 | 0.00408555 | positive | FBXW11,ERGIC1,LOC100288254,NEURL1B,SMIM23,UBTD2,EFCAB9,STK10,LOC101928093,MIR5003,DUSP1,SH3PXD2B,LINC01944 |
| 5 | 173606996 | 174662885 | GERD | MDD | 0.335852 | 0.0273346 | positive | MSX2,LINC01411,LINC01951,MIR4634 |
| 5 | 174662886 | 176180301 | GERD | MDD | 0.380212 | 0.0214579 | positive | LOC100507387,LINC01574,MIR4281,LOC100996385,CLTB,ARL10,MIR1271,HRH2,HIGD2A,LOC643201,TSPAN17,DRD1,KIAA1191,CDHR2,THOC3,SNCB,RNF44,GPRIN1,EIF4E1B,SFXN1,CPLX2,FAM153B,SIMC1,FAF2,NOP16 |
| 5 | 178595253 | 179794710 | GERD | MDD | 0.45866 | 0.00107483 | positive | RUFY1,RASGEF1C,MIR1229,LOC100996419,LTC4S,HNRNPH1,LOC105377763,MAPK9,CANX,MIR340,C5orf60,MGAT4B,TBC1D9B,ADAMTS2,MRNIP,MAML1,RUFY1-AS1,RNF130,GFPT2,SQSTM1,CBY3 |
| 6 | 63979 | 1380679 | GERD | MDD | 0.454327 | 0.0046076 | positive | LINC01622,LINC00266-3,IRF4,HUS1B,FOXQ1,LOC285766,EXOC2,DUSP22,LOC101927691 |
| 6 | 1380680 | 2746531 | GERD | MDD | 0.425612 | 0.00874444 | positive | FOXCUT,GMDS,FOXC1,GMDS-DT,FOXF2,MYLK4,MIR6720,LINC01600,LINC02521 |
| 6 | 9020711 | 10416550 | GERD | MDD | 0.492999 | 0.00940183 | positive | TFAP2A-AS2,TFAP2A,TFAP2A-AS1 |
| 6 | 11790672 | 13352326 | GERD | MDD | 0.881012 | 0.000293424 | positive | TBC1D7-LOC100130357,EDN1,LOC101928253,LOC100130357,PHACTR1,LINC02530,HIVEP1,TBC1D7 |
| 6 | 13352327 | 15069421 | GERD | MDD | 0.532287 | 0.000256845 | positive | CD83,LINC01108,GFOD1,SIRT5,NOL7,RANBP9,MCUR1,RNF182 |
| 6 | 16566883 | 17391994 | GERD | MDD | 0.998749 | 2.68162E-06 | positive | ATXN1-AS1,RBM24,STMND1,ATXN1 |
| 6 | 17391995 | 18208676 | GERD | MDD | 0.801555 | 0.000409434 | positive | LOC105374952,NHLRC1,KDM1B,CAP2,LOC101928491,TPMT,KIF13A,NUP153,FAM8A1 |
| 6 | 19449153 | 20418020 | GERD | MDD | 0.65549 | 0.00643572 | positive | MBOAT1,LOC105374960,E2F3,LNC-LBCS,ID4 |
| 6 | 22199714 | 23192337 | GERD | MDD | 0.701676 | 0.000782438 | positive | HDGFL1,LOC105374972,PRL |
| 6 | 23192338 | 23939306 | GERD | MDD | 0.550795 | 0.00184708 | positive | NA |
| 6 | 23939307 | 24950379 | GERD | MDD | 0.74379 | 0.00473572 | positive | RIPOR2,MRS2,KAAG1,GMNN,KIAA0319,ACOT13,NRSN1,TDP2,GPLD1,ARMH2,ALDH5A1,DCDC2,C6orf62 |
| 6 | 39785908 | 40778401 | GERD | MDD | 0.692198 | 2.47964E-06 | positive | LRFN2,DAAM2-AS1,DAAM2,TDRG1,LINC00951,MOCS1 |
| 6 | 40778402 | 42103738 | GERD | MDD | 0.534741 | 0.00154332 | positive | LINC01276,USP49,MED20,APOBEC2,TREML3P,FOXP4-AS1,TREML5P,MIR4641,PRICKLE4,TFEB,FRS3,BYSL,TREML4,TSPO2,NFYA,C6orf132,TREML1,NCR2,LOC101929555,ADCY10P1,OARD1,TOMM6,PGC,TREM2,TREML2,TAF8,FOXP4,MIR10398,CCND3,UNC5CL,TREM1,MDFI |
| 6 | 42103739 | 43770626 | GERD | MDD | 0.68485 | 0.00177819 | positive | CUL7,TJAP1,C6orf226,MRPL2,DNPH1,MEA1,GUCA1ANB,CUL9,CNPY3,RSPH9,DLK2,XPO5,KLC4,BICRAL,VEGFA,MRPS10,PPP2R5D,MIR6780B,GNMT,LOC401261,ABCC10,PRPH2,SRF,MRPS18A,POLR1C,KLHDC3,TTBK1,CNPY3-GNMT,TBCC,SLC22A7,C6orf132,RRP36,RPL7L1,MAD2L1BP,ATP6V0CP3,GTPBP2,PTK7,LRRC73,UBR2,ZNF318,GUCA1A,CRIP3,PTCRA,POLH,PEX6,TRERF1,YIPF3,GUCA1B |
| 6 | 49528165 | 50940767 | GERD | MDD | 1 | 2.27395E-05 | positive | PGK2,DEFB110,DEFB113,CRISP1,DEFB133,LOC101927020,CRISP3,CRISP2,RHAG,LOC101927048,TFAP2D,TFAP2B,C6orf141,DEFB114,DEFB112 |
| 6 | 64010287 | 65759861 | GERD | MDD | 1 | 1.60988E-08 | positive | PTP4A1,LGSN,EYS,SCAT8,PHF3 |
| 6 | 68547495 | 69294227 | GERD | MDD | 0.667773 | 0.000793649 | positive | LOC102723883,LINC02549 |
| 6 | 70363349 | 71610787 | GERD | MDD | 0.763272 | 0.00210802 | positive | COL19A1,SDHAF4,LMBRD1,EVADR,FAM135A-AS1,B3GAT2,COL9A1,SMAP1,FAM135A |
| 6 | 83571954 | 85038134 | GERD | MDD | 0.913636 | 0.000331938 | positive | RWDD2A,CEP162,DOP1A,ME1,SNAP91,RIPPLY2,PRSS35,MRAP2,LOC105377879,CYB5R4,UBE3D,PGM3 |
| 6 | 91097025 | 92015279 | GERD | MDD | 1 | 0.000748242 | positive | MAP3K7 |
| 6 | 93696826 | 95176337 | GERD | MDD | 0.64384 | 0.000217645 | positive | TSG1,EPHA7 |
| 6 | 96480154 | 98173003 | GERD | MDD | 0.47675 | 0.00745348 | positive | GPR63,MMS22L,NDUFAF4,LOC101927314,KLHL32,FHL5,UFL1,FUT9 |
| 6 | 98173004 | 99678876 | GERD | MDD | 0.709668 | 1.98383E-06 | positive | FBXL4,PNKY,POU3F2,MIR548AI,MIR2113 |
| 6 | 100878646 | 101793542 | GERD | MDD | 0.620986 | 0.00147792 | positive | ASCC3,SIM1 |
| 6 | 101793543 | 102637706 | GERD | MDD | 0.57443 | 0.0176555 | positive | GRIK2 |
| 6 | 106053916 | 107309327 | GERD | MDD | 0.970154 | 0.000368831 | positive | MIR587,RTN4IP1,CRYBG1,LOC105377924,QRSL1,PRDM1,LINC02532,LINC02526,ATG5 |
| 6 | 107309328 | 109043244 | GERD | MDD | 0.75401 | 6.55037E-05 | positive | SCML4,SNX3,FOXO3,SEC63,OSTM1,NR2E1,SOBP,OSTM1-AS1,BEND3,MTRES1,AFG1L,PDSS2 |
| 6 | 122929291 | 123854857 | GERD | MDD | 0.645695 | 0.0107077 | positive | SMPDL3A,TRDN,TRDN-AS1,CLVS2,PKIB,FABP7 |
| 6 | 123854858 | 125365054 | GERD | MDD | 0.448874 | 0.00351526 | positive | TRDN,NKAIN2,RNF217-AS1,RNF217 |
| 6 | 125365055 | 127545459 | GERD | MDD | 0.774968 | 0.00998147 | positive | LINC02523,RNF217,NCOA7,TRMT11,CENPW,HDDC2,NCOA7-AS1,MIR588,RSPO3,HINT3,TPD52L1,HEY2,LOC100126584 |
| 6 | 129850179 | 130550137 | GERD | MDD | 0.545917 | 0.00109928 | positive | SAMD3,TMEM244,ARHGAP18,L3MBTL3 |
| 6 | 131457165 | 132890694 | GERD | MDD | 0.556049 | 0.00428302 | positive | ENPP1,TAAR8,STX7,OR2A4,CTAGE9,MIR548AJ1,MOXD1,AKAP7,CCN2,LINC01013,TAAR9,MED23,ENPP3,ARG1,MIR548H5 |
| 6 | 139716714 | 141449453 | GERD | MDD | 0.572736 | 0.000212209 | positive | FILNC1,MIR4465,LINC01625,MIR3668,LOC100507477,LOC103352541 |
| 6 | 141449454 | 143083347 | GERD | MDD | 0.582641 | 0.0106679 | positive | LOC153910,VTA1,NMBR,GJE1,HIVEP2,ADGRG6 |
| 6 | 143083348 | 144791455 | GERD | MDD | 0.599806 | 7.71736E-06 | positive | LTV1,HIVEP2,SF3B5,PHACTR2,SNORA98,STX11,PLAGL1,UTRN,ADAT2,LINC01277,PEX3,ZC2HC1B,HYMAI,PHACTR2-AS1,FUCA2,AIG1 |
| 6 | 147015883 | 148184844 | GERD | MDD | 0.539795 | 0.0201077 | positive | STXBP5-AS1,ADGB,SAMD5,STXBP5,LUADT1,KATNBL1P6 |
| 6 | 148184845 | 149278798 | GERD | MDD | 0.825539 | 0.000191557 | positive | UST,UST-AS1,SASH1 |
| 6 | 151629955 | 152376499 | GERD | MDD | 0.494489 | 0.000640045 | positive | ARMT1,AKAP12,CCDC170,ESR1,RMND1,ZBTB2 |
| 6 | 155523958 | 156762740 | GERD | MDD | 0.865512 | 0.000431426 | positive | CLDN20,NOX3,MIR1202,LOC105378068,SNORD28B,TFB1M,TIAM2 |
| 6 | 161371015 | 162254229 | GERD | MDD | 0.462503 | 0.00149153 | positive | LOC105378098,AGPAT4,AGPAT4-IT1,MAP3K4,PRKN |
| 6 | 162254230 | 163336361 | GERD | MDD | 0.424771 | 0.00959116 | positive | PACRG,PRKN |
| 6 | 163336362 | 164687462 | GERD | MDD | 0.604819 | 3.4738E-07 | positive | CAHM,PACRG-AS3,DKFZp451B082,QKI,PACRG-AS1,PACRG-AS2,LOC102724152,PACRG |
| 6 | 164687463 | 165835953 | GERD | MDD | 0.631065 | 0.000210638 | positive | C6orf118,MEAT6,PDE10A |
| 6 | 165835954 | 166853325 | GERD | MDD | 1 | 1.18978E-05 | positive | LOC101929297,LNCDAT,LINC00602,PDE10A,MPC1,SFT2D1,LOC729681,LINC00473,RPS6KA2,LOC100289495,TBXT,PRR18 |
| 7 | 617320 | 1366972 | GERD | MDD | 1 | 0.000295497 | positive | ZFAND2A-DT,UNCX,ADAP1,PRKAR1B-AS1,PRKAR1B,GPR146,MIR339,DNAAF5,SUN1,GPER1,CYP2W1,GET4,ZFAND2A,C7orf50,COX19 |
| 7 | 1366973 | 2473749 | GERD | MDD | 0.814686 | 4.27866E-08 | positive | NUDT1,LOC100128653,ELFN1,MIR4655,MRM2,CHST12,TFAMP1,INTS1,PSMG3,TMEM184A,SNX8,MAFK,ELFN1-AS1,SNORA114,EIF3B,MAD1L1,MIR6836,LOC105375303,MICALL2,PSMG3-AS1 |
| 7 | 2473750 | 3100211 | GERD | MDD | 0.608137 | 0.00129137 | positive | BRAT1,TTYH3,CHST12,IQCE,GNA12,MIR4648,AMZ1,CARD11,LFNG,GRIFIN |
| 7 | 3100212 | 4139431 | GERD | MDD | 0.830801 | 1.57863E-07 | positive | SDK1,LOC100129603 |
| 7 | 4139432 | 4938442 | GERD | MDD | 0.746442 | 0.000290557 | positive | RADIL,FOXK1,MMD2,LOC105375131,PAPOLB,AP5Z1,MIR4656,SDK1,SNORD165 |
| 7 | 4938443 | 6020653 | GERD | MDD | 0.574444 | 0.0163839 | positive | WIPI2,MIR589,RSPH10B2,LOC100129484,SLC29A4,CCZ1,ACTB,RNF216-IT1,OCM,PMS2,RBAKDN,TNRC18,RNF216P1,RBAK-RBAKDN,ZNF815P,FBXL18,RSPH10B,RNF216,FSCN1,ZNF890P,LOC221946,MMD2,MIR6874,RBAK |
| 7 | 6020654 | 6716905 | GERD | MDD | 0.868091 | 0.000875387 | positive | ZNF853,SNORA80D,DAGLB,ZDHHC4,ZNF316,CYTH3,USP42,ANKRD61,PMS2,AIMP2,C7orf26,EIF2AK1,RAC1,GRID2IP,FAM220A,KDELR2 |
| 7 | 7167611 | 7926500 | GERD | MDD | 0.488146 | 0.0178974 | positive | UMAD1,MIOS,LOC101927391,C1GALT1,RPA3,COL28A1,LOC101927354 |
| 7 | 7926501 | 8718351 | GERD | MDD | 0.753758 | 0.000023192 | positive | NXPH1,ICA1,GLCCI1-DT,LOC100505938,GLCCI1 |
| 7 | 11232448 | 11855413 | GERD | MDD | 1 | 0.00223896 | positive | THSD7A |
| 7 | 11855414 | 12694915 | GERD | MDD | 1 | 1.24073E-07 | positive | SCIN,VWDE,TMEM106B,THSD7A,LOC102725191 |
| 7 | 13676748 | 15013694 | GERD | MDD | 0.528395 | 0.000231166 | positive | DGKB,ETV1 |
| 7 | 15877565 | 16739013 | GERD | MDD | 0.559157 | 0.0197628 | positive | SOSTDC1,CRPPA-AS1,LRRC72,CRPPA,BZW2,ANKMY2 |
| 7 | 16739014 | 17596578 | GERD | MDD | 0.746538 | 0.00186355 | positive | TSPAN13,AGR2,LINC02888,AHR,BZW2,LINC02889,AGR3 |
| 7 | 20122180 | 21390644 | GERD | MDD | 0.782672 | 0.00250887 | positive | SP8,ITGB8,MACC1-AS1,ABCB5,LOC101927811,LOC101927769,MACC1,LOC100506098,RPL23P8,LOC101927668,LINC01162 |
| 7 | 21390645 | 22165060 | GERD | MDD | 0.765335 | 2.71318E-05 | positive | SP4,MIR1183,RAPGEF5,DNAH11,CDCA7L |
| 7 | 28890887 | 30175113 | GERD | MDD | 1 | 1.54101E-08 | positive | PRR15,DPY19L2P3,FKBP14,LOC101928168,WIPF3,CHN2,MTURN,TRIL,CPVL,LOC100506497,LOC646762,ZNRF2P2,SCRN1,MIR550A3,PLEKHA8,CHN2-AS1 |
| 7 | 30175114 | 31730554 | GERD | MDD | 0.813806 | 0.000237016 | positive | NOD1,LOC105375218,ZNRF2,DKFZP586I1420,GARS1,PPP1R17,MIR550B1,LINC01176,INMT,NEUROD6,GHRHR,MTURN,INMT-MINDY4,GARS1-DT,MIR550A1,ITPRID1,CRHR2,AQP1,MINDY4,GGCT,ADCYAP1R1 |
| 7 | 31730555 | 32460228 | GERD | MDD | 0.689747 | 0.000469095 | positive | PDE1C,PPP1R17 |
| 7 | 38966484 | 40261240 | GERD | MDD | 0.813051 | 0.00675896 | positive | POU6F2,YAE1,POU6F2-AS2,RALA,LINC00265,CDK13,POU6F2-AS1,SNORA20B,SUGCT,LOC112267983,MPLKIP |
| 7 | 40261241 | 41415925 | GERD | MDD | 0.625617 | 0.0131724 | positive | LINC01450,SUGCT,LINC01449 |
| 7 | 43448816 | 44989571 | GERD | MDD | 0.422286 | 0.0291046 | positive | UBE2D4,LINC00957,COA1,OGDH,H2AZ2,PURB,STK17A,CAMK2B,URGCP-MRPS24,NPC1L1,HECW1,LUARIS,MRPS24,ZMIZ2,MIR6837,BLVRA,PGAM2,URGCP,DBNL,YKT6,MIR4657,POLM,MIR6838,PPIA,AEBP1,NUDCD3,MYL7,MIR4649,POLD2,SPDYE1,RASA4CP,GCK,POLR2J4,DDX56,TMED4 |
| 7 | 44989572 | 46247335 | GERD | MDD | 0.638261 | 0.00147102 | positive | SNORA9,NACAD,SNHG15,TBRG4,SNORA5A,IGFBP1,SNORA5C,SNORA5B,MYO1G,ADCY1,CCM2,SEPTIN7P2,LOC730234,IGFBP3,RAMP3 |
| 7 | 49632427 | 50894508 | GERD | MDD | 0.851831 | 0.00131693 | positive | GRB10,ZPBP,DDC-AS1,FIGNL1,IKZF1,SPATA48,VWC2,DDC |
| 7 | 50894509 | 51951647 | GERD | MDD | 0.629281 | 0.00568305 | positive | COBL,LOC107986794 |
| 7 | 70144615 | 71215741 | GERD | MDD | 0.669408 | 0.00093603 | positive | MIR3914-2,AUTS2,GALNT17,MIR3914-1 |
| 7 | 71215742 | 73128830 | GERD | MDD | 0.829511 | 0.000214265 | positive | NSUN5,MLXIPL,BUD23,BCL7B,STAG3L1,PMS2P7,SPDYE13,FZD9,SPDYE11,TBL2,DNAJC30,VPS37D,STX1A,GTF2IP1,SPDYE7P,LOC100101148,MIR4650-1,SPDYE14,SPDYE15,SPDYE10P,PMS2P5,SPDYE9,SBDSP1,TYW1B,PMS2P2,STAG3L3,MIR4284,GTF2IP4,FKBP6,NSUN5P2,LOC541473,NCF1B,TRIM73,SPDYE8,TRIM50,BAZ1B,MIR4650-2,TRIM74,CALN1,POM121 |
| 7 | 77663965 | 78685009 | GERD | MDD | 0.640167 | 8.00401E-07 | positive | MAGI2,MAGI2-AS2,RPL13AP17 |
| 7 | 81533567 | 82759283 | GERD | MDD | 0.760931 | 1.67364E-07 | positive | CACNA2D1-AS1,CACNA2D1,PCLO |
| 7 | 95174848 | 96564981 | GERD | MDD | 0.851324 | 0.000640609 | positive | PDK4,DYNC1I1,MIR591,SLC25A13,SEM1 |
| 7 | 96564982 | 97420207 | GERD | MDD | 0.744908 | 0.00193272 | positive | SDHAF3,DLX5,DLX6,DLX6-AS1,TAC1 |
| 7 | 100849308 | 101950748 | GERD | MDD | 0.445496 | 0.0271266 | positive | COL26A1,MYL10,PLOD3,MIR4285,IFT22,EMSLR,FIS1,ZNHIT1,LNCPRESS1,LINC01007,CLDN15,SH2B2,CUX1 |
| 7 | 107658604 | 109095558 | GERD | MDD | 0.679375 | 0.000155561 | positive | DNAJB9,NRCAM,LAMB4,C7orf66,PNPLA8,THAP5 |
| 7 | 109095559 | 110479479 | GERD | MDD | 0.46288 | 0.00316758 | positive | EIF3IP1,IMMP2L |
| 7 | 110479480 | 111795299 | GERD | MDD | 0.634787 | 0.027496 | positive | DOCK4-AS1,IMMP2L,DOCK4,LRRN3 |
| 7 | 113339387 | 115321301 | GERD | MDD | 0.659536 | 5.8543E-07 | positive | MIR3666,LINC01392,LINC01393,PPP1R3A,FOXP2,SNORA25B,MDFIC |
| 7 | 116835337 | 118342090 | GERD | MDD | 0.646407 | 0.0104095 | positive | ST7,CFTR,CFTR-AS1,ASZ1,ANKRD7,WNT2,LSM8,ST7-OT3,CTTNBP2 |
| 7 | 125389587 | 126640872 | GERD | MDD | 0.665527 | 0.00308129 | positive | GRM8 |
| 7 | 126640873 | 127903738 | GERD | MDD | 0.434857 | 0.0165212 | positive | LRRC4,LEP,MIR592,GCC1,ZNF800,MIR593,SND1-IT1,MIR129-1,SND1,LOC100506682,PAX4,FSCN3,ARF5,GRM8-AS1,GRM8 |
| 7 | 128779174 | 130418704 | GERD | MDD | 0.629358 | 0.000246952 | positive | CEP41,UBE2H,MEST,SMO,TSPAN33,NRF1,MIR182,MIR335,COPG2,KLHDC10,ZC3HC1,LOC105375504,MESTIT1,CPA1,CPA4,AHCYL2,LOC100128325,STRIP2,TMEM209,SSMEM1,SMKR1,KLF14,CPA5,TSGA13,MIR183,CPA2,MIR96 |
| 7 | 130418705 | 131856481 | GERD | MDD | 0.430716 | 0.00613261 | positive | PODXL,LINC00513,MKLN1-AS,MIR29B1,PLXNA4,LINC-PINT,MKLN1,MIR29A,KLF14,LOC101928782 |
| 7 | 131856482 | 133807216 | GERD | MDD | 0.418624 | 0.00783398 | positive | FLJ40288,LOC101928861,PLXNA4,LOC100506937,LOC105375512,CHCHD3,EXOC4,MIR3654,MIR6133,LOC101928807 |
| 7 | 134943222 | 136113467 | GERD | MDD | 1 | 0.000685072 | positive | LUZP6,STMP1,FAM180A,STRA8,SLC13A4,MTPN,NUP205,CNOT4 |
| 7 | 136113468 | 137657459 | GERD | MDD | 0.404413 | 0.0287907 | positive | LOC349160,CHRM2,CREB3L2,MIR490,CREB3L2-AS1,DGKI,PTN |
| 7 | 154407488 | 155280610 | GERD | MDD | 0.529246 | 0.00564721 | positive | EN2,BLACE,HTR5A,HTR5A-AS1,PAXIP1-AS2,PAXIP1,INSIG1,LOC100286906,PAXIP1-DT,DPP6 |
| 8 | 3386668 | 3787248 | GERD | MDD | 0.647795 | 2.91035E-05 | positive | CSMD1 |
| 8 | 3787249 | 4273083 | GERD | MDD | 0.696876 | 1.66586E-05 | positive | CSMD1 |
| 8 | 4893121 | 5566269 | GERD | MDD | 0.534642 | 0.0172558 | positive | NA |
| 8 | 8064601 | 8589770 | GERD | MDD | 0.368407 | 0.0247413 | positive | CLDN23,FAM85B,PRAG1,FAM86B3P |
| 8 | 12296850 | 12947935 | GERD | MDD | 0.644541 | 0.00740235 | positive | LONRF1,LINC00681,MIR5692A2,MIR3926-2,DLC1,LOC340357,MIR5692A1,LOC729732,TRMT9B,MIR3926-1,LOC100506990 |
| 8 | 13395052 | 13948095 | GERD | MDD | 0.603169 | 0.0152692 | positive | SGCZ,DLC1,LOC102725080,C8orf48 |
| 8 | 13948096 | 14389305 | GERD | MDD | 0.557594 | 0.00871659 | positive | SGCZ |
| 8 | 31134787 | 32454962 | GERD | MDD | 0.45329 | 0.0186062 | positive | NRG1-IT1,NRG1,NRG1-IT3 |
| 8 | 32454963 | 33982537 | GERD | MDD | 0.829285 | 0.00448249 | positive | DUSP26,MAK16,NRG1,TTI2,SNORD13,FUT10,RNF122 |
| 8 | 33982538 | 34792220 | GERD | MDD | 0.97988 | 0.000291377 | positive | LINC01288 |
| 8 | 36641175 | 38803980 | GERD | MDD | 0.483344 | 0.000029526 | positive | GOT1L1,C8orf86,ADRB3,KCNU1,LOC101929622,FGFR1,RAB11FIP1,LETM2,LSM1,PLEKHA2,EIF4EBP1,LOC728024,RNF5P1,PLPP5,MIR1268A,BRF2,LINC01605,ZNF703,ASH2L,TACC1,ERLIN2,BAG4,LOC102723701,DDHD2,ADGRA2,PLPBP,NSD3,STAR |
| 8 | 41725003 | 43825633 | GERD | MDD | 0.550222 | 0.0267513 | positive | DKK4,RNF170,CHRNB3,ANK1,PLAT,SMIM19,POMK,HGSNAT,LOC105379393,THAP1,IKBKB,CHRNA6,MIR4469,VDAC3,KAT6A,AP3M2,POLB,FNTA,POTEA,LOC101929897,SLC20A2,HOOK3 |
| 8 | 55275355 | 56346878 | GERD | MDD | 0.547753 | 0.00105579 | positive | XKR4,RP1,SOX17 |
| 8 | 59555978 | 60476248 | GERD | MDD | 0.705701 | 0.00102379 | positive | TOX,NSMAF,LOC100505501 |
| 8 | 60476249 | 61324889 | GERD | MDD | 0.634193 | 0.0242623 | positive | CA8,LINC01301 |
| 8 | 64215359 | 66018204 | GERD | MDD | 0.547255 | 0.0155698 | positive | LOC401463,MIR124-2HG,LINC01289,LOC102724612,CYP7B1,BHLHE22,MIR124-2,LINC01414 |
| 8 | 66018205 | 66960438 | GERD | MDD | 0.613269 | 0.00265657 | positive | PDE7A,MTFR1,LINC00251,LINC01299,DNAJC5B,ARMC1 |
| 8 | 66960439 | 68683823 | GERD | MDD | 0.628104 | 0.00569954 | positive | LINC00967,MYBL1,ARFGEF1,RRS1-AS1,DNAJC5B,CPA6,TCF24,CSPP1,C8orf44-SGK3,PPP1R42,SGK3,MCMDC2,C8orf44,VCPIP1,LOC102724708,TRIM55,CRH,SNORD87,SNHG6,COPS5,RRS1,PTTG3P,ADHFE1,VXN |
| 8 | 70004666 | 70872088 | GERD | MDD | 0.717708 | 0.0100711 | positive | LINC01592,SULF1,LINC01603,SLCO5A1 |
| 8 | 72013185 | 72917489 | GERD | MDD | 0.983091 | 0.000473191 | positive | MSC,MSC-AS1,EYA1 |
| 8 | 73838028 | 75011699 | GERD | MDD | 0.747328 | 2.09076E-06 | positive | RDH10,KCNB2,SBSPON,TMEM70,RPL7,C8orf89,TERF1,LY96,STAU2,RDH10-AS1,STAU2-AS1,ELOC,UBE2W,LINC01617 |
| 8 | 76295484 | 77377989 | GERD | MDD | 0.494883 | 0.0245784 | positive | HNF4G,LINC01111 |
| 8 | 79756170 | 81340231 | GERD | MDD | 0.610406 | 0.00930055 | positive | MIR12123,HEY1,LINC01607,STMN2,MIR5708,MRPS28,LOC101927040,TPD52 |
| 8 | 81340232 | 82878769 | GERD | MDD | 0.620498 | 0.00647542 | positive | ZNF704,PAG1,FABP9,CHMP4C,IMPA1P1,SLC10A5,FABP5,IMPA1,ZBTB10,ZFAND1,SNX16,FABP12,PMP2,FABP4 |
| 8 | 87563146 | 89081177 | GERD | MDD | 0.532191 | 0.00793398 | positive | CNBD1,CNGB3,MMP16,CPNE3,DCAF4L2 |
| 8 | 89081178 | 90364149 | GERD | MDD | 0.750359 | 0.00453834 | positive | MMP16 |
| 8 | 90364150 | 91457962 | GERD | MDD | 0.861484 | 0.000302274 | positive | LOC101929709,LINC00534,NBN,RIPK2,OSGIN2,DECR1,CALB1 |
| 8 | 91457963 | 92876620 | GERD | MDD | 1 | 7.43356E-06 | positive | MIR4661,SLC26A7,TMEM64,OTUD6B,LINC01030,C8orf88,NECAB1,LRRC69,PIP4P2,OTUD6B-AS1 |
| 8 | 92876621 | 94999066 | GERD | MDD | 0.733641 | 7.44659E-07 | positive | C8orf87,LOC102724710,MIR378D2,RUNX1T1,LINC00535,FLJ46284,TMEM67,RBM12B,RBM12B-AS1,PDP1,CIBAR1,MIR8084,TRIQK |
| 8 | 98132520 | 99388944 | GERD | MDD | -0.550613 | 0.0150748 | negative | RPL30,LOC101927066,MTDH,TSPYL5,RIDA,ERICH5,CPQ,POP1,SNORA72,MATN2,NIPAL2,LAPTM4B,SNORD3H |
| 8 | 102548681 | 103716053 | GERD | MDD | 0.566326 | 0.008784 | positive | NCALD,LOC105375683,LOC101927245,RRM2B,MIR5680,GRHL2,UBR5,UBR5-AS1,LOC104054148,KLF10,ODF1 |
| 8 | 104467846 | 105370130 | GERD | MDD | 0.587663 | 0.0279009 | positive | DCSTAMP,RIMS2,LOC105375690 |
| 8 | 105370131 | 106500950 | GERD | MDD | 0.586537 | 0.00078398 | positive | MIR548A3,LRP12,DPYS,ZFPM2 |
| 8 | 107787164 | 108647463 | GERD | MDD | 0.906254 | 0.000257325 | positive | ANGPT1 |
| 8 | 111207350 | 112601513 | GERD | MDD | 0.821349 | 0.00737585 | positive | LINC01609,LINC01608,LINC02237 |
| 8 | 124001020 | 125453322 | GERD | MDD | 0.444539 | 0.00693851 | positive | ATAD2,FAM91A1,DERL1,FER1L6-AS2,FAM83A,FER1L6,LOC101927588,ZHX1,FER1L6-AS1,C8orf76,KLHL38,MIR4663,ZHX1-C8orf76,ANXA13,FAM83A-AS1,MIR548D1,TMEM65,NTAQ1,TBC1D31,MIR548AA1,FBXO32 |
| 8 | 129291119 | 130383563 | GERD | MDD | 0.694385 | 0.00120967 | positive | LINC00824,LINC00976,LINC00977,CCDC26 |
| 8 | 132534637 | 133461551 | GERD | MDD | 0.593202 | 0.0186946 | positive | KCNQ3,HHLA1,OC90,EFR3A |
| 8 | 133461552 | 134509448 | GERD | MDD | 0.695146 | 3.14166E-05 | positive | TMEM71,MIR7848,PTCSC1,HPYR1,ST3GAL1,LRRC6,SLA,PHF20L1,KCNQ3,CCN4,TG,NDRG1 |
| 8 | 135313674 | 136220342 | GERD | MDD | 0.677886 | 0.000564684 | positive | MIR30B,NCRNA00250,LOC101927845,ZFAT-AS1,MIR30D,ZFAT |
| 8 | 136220343 | 137527617 | GERD | MDD | 0.620432 | 0.00260487 | positive | KHDRBS3,LINC01591 |
| 8 | 138564844 | 139404631 | GERD | MDD | 0.65595 | 0.00184863 | positive | LOC401478,FAM135B |
| 8 | 140355989 | 141527881 | GERD | MDD | 0.506011 | 0.00427061 | positive | TRAPPC9,CHRAC1,KCNK9,PEG13 |
| 8 | 142611890 | 143611461 | GERD | MDD | 0.391744 | 0.00771295 | positive | MIR1302-7,LINC00051,ADGRB1,MIR4472-1,TSNARE1,MIR4539 |
| 8 | 144934289 | 146303787 | GERD | MDD | 0.761933 | 0.00083179 | positive | ZNF16,TONSL-AS1,MIR6850,ZNF252P,SLC52A2,MIR6893,ARHGAP39,SMPD5,DGAT1,TMEM249,MIR661,PARP10,SCRT1,FOXH1,ZNF34,LRRC14,MIR6846,COMMD5,ZNF250,HSF1,LRRC24,LOC101928902,CYC1,PPP1R16A,MIR6847,ZNF517,FBXL6,RPL8,C8orf82,LOC100130027,BOP1,KIFC2,MIR7112,EXOSC4,CPSF1,MIR6848,OPLAH,MIR6849,PLEC,SPATC1,SHARPIN,WDR97,MROH1,RECQL4,GRINA,GPT,CYHR1,MIR10400,SCX,ZNF7,MFSD3,GPAA1,ZNF251,ADCK5,EPPK1,VPS28,ZNF252P-AS1,TMED10P1,MIR939,C8orf33,HGH1,MAF1,TONSL,SLC39A4 |
| 9 | 1874962 | 2441120 | GERD | MDD | 0.834358 | 0.000458383 | positive | SMARCA2 |
| 9 | 2441121 | 3079045 | GERD | MDD | 0.669658 | 0.0155462 | positive | VLDLR,PUM3,VLDLR-AS1,KCNV2 |
| 9 | 3079046 | 4025355 | GERD | MDD | 0.925704 | 4.68887E-05 | positive | GLIS3-AS1,GLIS3,RFX3-AS1,LINC01231,RFX3 |
| 9 | 4025356 | 4596614 | GERD | MDD | 0.464597 | 0.00486016 | positive | SLC1A1,SPATA6L,GLIS3 |
| 9 | 7388203 | 8262303 | GERD | MDD | 0.500863 | 0.0135656 | positive | DMAC1 |
| 9 | 8262304 | 9170120 | GERD | MDD | 0.362359 | 0.0025579 | positive | PTPRD-AS1,PTPRD |
| 9 | 9920259 | 10675474 | GERD | MDD | 0.54986 | 0.00835228 | positive | PTPRD,PTPRD-AS2 |
| 9 | 14244748 | 14902924 | GERD | MDD | 0.387934 | 0.00907492 | positive | ZDHHC21,FREM1,NFIB,CER1 |
| 9 | 15839404 | 16658377 | GERD | MDD | 0.406645 | 0.0224033 | positive | CCDC171,BNC2,C9orf92 |
| 9 | 20483881 | 21674032 | GERD | MDD | 0.736764 | 0.000347148 | positive | IFNA22P,IFNA8,MIR31,IFNA17,IFNA1,IFNA16,IFNA10,IFNA5,MIR491,IFNA14,IFNA6,SNORA30B,MIR4474,IFNW1,IFNE,MLLT3,FOCAD,MIR31HG,IFNA4,IFNA7,HACD4,IFNB1,IFNA13,IFNA21,IFNA2,KLHL9,FOCAD-AS1 |
| 9 | 23091194 | 23859374 | GERD | MDD | 0.429404 | 0.0120981 | positive | ELAVL2,LOC101929563 |
| 9 | 28067455 | 28766060 | GERD | MDD | 0.741281 | 0.000922325 | positive | LINGO2 |
| 9 | 30410079 | 31115153 | GERD | MDD | 0.703563 | 0.0103215 | positive | NA |
| 9 | 32253284 | 33194892 | GERD | MDD | 0.574396 | 0.0293063 | positive | B4GALT1,MIR12117,DNAJA1,TOPORS,NDUFB6,DDX58,SMU1,APTX,B4GALT1-AS1,TAF1L,ACO1,SMIM27,TMEM215 |
| 9 | 36749274 | 37687112 | GERD | MDD | 0.377539 | 0.0173027 | positive | TOMM5,GRHPR,MIR4476,FBXO10,EBLN3P,PAX5,MIR4540,MIR4475,POLR1E,FRMPD1,ZCCHC7,ZBTB5 |
| 9 | 72288879 | 73915494 | GERD | MDD | 0.569288 | 0.0125526 | positive | PTAR1,MAMDC2-AS1,KLF9,C9orf135-DT,SMC5,SMC5-AS1,C9orf135,MAMDC2,TRPM3,MIR204 |
| 9 | 76197745 | 77862308 | GERD | MDD | 0.725542 | 0.000152368 | positive | LOC101927358,RORB-AS1,CARNMT1-AS1,OSTF1,NMRK1,C9orf40,RORB,TRPM6,CARNMT1 |
| 9 | 78630916 | 79451035 | GERD | MDD | 0.68969 | 0.014829 | positive | GCNT1,RFK,PRUNE2,PCA3,PCSK5,RPSAP9 |
| 9 | 81161598 | 82601465 | GERD | MDD | 0.587344 | 2.06701E-05 | positive | TLE4,LOC101927450,LINC01507 |
| 9 | 91643256 | 92500277 | GERD | MDD | 0.713835 | 0.00470854 | positive | MIR3153,SECISBP2,SEMA4D,CKS2,UNQ6494,SHC3,GADD45G |
| 9 | 95854926 | 96670171 | GERD | MDD | 0.394779 | 0.00504638 | positive | FAM120AOS,MIR4291,C9orf129,FAM120A,NINJ1,WNK2,CARD19,PHF2,MIR548AU |
| 9 | 101596751 | 103165725 | GERD | MDD | 0.910745 | 0.000509781 | positive | TEX10,ERP44,ALG2,TGFBR1,GALNT12,NAMA,INVS,NR4A3,COL15A1,SEC61B,STX17,STX17-AS1,LOC101928438 |
| 9 | 108203096 | 109150717 | GERD | MDD | 0.819243 | 0.000219548 | positive | RALGAPA1P1,FSD1L,TMEM38B,FKTN,TAL2 |
| 9 | 112865521 | 113646149 | GERD | MDD | 0.720027 | 0.00894303 | positive | TXNDC8,LPAR1,C9orf152,SVEP1,TXN,PALM2AKAP2,MUSK |
| 9 | 119001228 | 119890464 | GERD | MDD | 0.644631 | 1.00658E-05 | positive | PAPPA-AS1,PAPPA,ASTN2-AS1,TRIM32,ASTN2 |
| 9 | 119890465 | 121008529 | GERD | MDD | 0.475886 | 0.00328199 | positive | SNORA70C,TLR4,ASTN2,LOC101928797 |
| 9 | 121008530 | 122677719 | GERD | MDD | 0.465188 | 4.21629E-05 | positive | BRINP1,LINC02578 |
| 9 | 126092932 | 127184582 | GERD | MDD | 0.484789 | 0.0010559 | positive | PSMB7,LOC613206,MIR7150,LHX2,LOC100505588,NEK6,MIR601,DENND1A,CRB2,LOC100129034 |
| 9 | 128785783 | 129617771 | GERD | MDD | 0.331747 | 0.0118242 | positive | ZBTB43,LOC101929116,LMX1B,NRON,MVB12B |
| 9 | 132193799 | 132999452 | GERD | MDD | 0.803655 | 2.40754E-07 | positive | TOR1A,TOR1B,GPRACR,MIR6855,NCS1,PTGES,ASB6,C9orf78,FNBP1,NTMT1,C9orf50,LINC00963,USP20,GPR107,PRRX2,PRRX2-AS1 |
| 9 | 132999453 | 134141936 | GERD | MDD | 0.72101 | 7.25423E-05 | positive | ABL1,EXOSC2,QRFP,PRDM12,NCS1,MIR12126,LOC100272217,FUBP3,NUP214,FAM78A,ASS1,HMCN2,MIR6856,AIF1L,LAMC3,FIBCD1 |
| 9 | 134141937 | 135135887 | GERD | MDD | 0.667103 | 0.00422576 | positive | MED27,UCK1,SNORD62A,FAM78A,PLPP7,POMT1,NTNG2,RAPGEF1,PRRC2B,PRRT1B,SNORD62B |
| 9 | 135135888 | 136042490 | GERD | MDD | 0.700669 | 0.000307134 | positive | DDX31,GTF3C5,RALGDS,TSC1,CELP,BARHL1,SETX,CFAP77,SNORD141B,SNORD141A,MIR548AW,GFI1B,CEL,LOC105376306,AK8,GBGT1,MIR6877,SPACA9,TTF1,GTF3C4 |
| 9 | 136042491 | 136770926 | GERD | MDD | 1 | 0.000216398 | positive | DBH,ADAMTS13,SNORD24,ABO,DBH-AS1,STKLD1,SURF1,SARDH,SURF4,ADAMTSL2,SLC2A6,MED22,SNORD36B,MYMK,RPL7A,SURF2,FAM163B,SNORD36C,OBP2B,SURF6,CACFD1,SNORD36A,REXO4,VAV2 |
| 9 | 137593528 | 138202112 | GERD | MDD | 0.600289 | 0.000380483 | positive | MIR3689B,LOC401557,MIR3689A,MIR3689C,MIR3689E,FCN2,MIR3689D1,LOC101448202,MIR3689F,COL5A1,FCN1,OLFM1,MIR3689D2 |
| 10 | 3880323 | 4813182 | GERD | MDD | 0.505981 | 0.0123845 | positive | LOC101927964,MANCR,LINC00705,LINC00702,LINC00703,MIR6078 |
| 10 | 5396243 | 6234718 | GERD | MDD | 0.424367 | 0.00441389 | positive | IL15RA,FBH1,CALML5,CALML3,GDI2,IL2RA,MIR3155B,TUBAL3,NET1,TASOR2,UCN3,CALML3-AS1,PFKFB3,LASTR,ANKRD16,RBM17,MIR3155A,ASB13 |
| 10 | 7659873 | 8771920 | GERD | MDD | 0.723533 | 1.70769E-06 | positive | ATP5F1C,ITIH5,GATA3,TAF3,KIN,GATA3-AS1,LINC00708,ITIH2,LOC105376398 |
| 10 | 8771921 | 10081279 | GERD | MDD | 0.594593 | 0.00282461 | positive | LOC101928272,LINC02676,LINC00709 |
| 10 | 11856925 | 12581571 | GERD | MDD | 0.858435 | 0.000163454 | positive | CDC123,MIR548AK,UPF2,CAMK1D,SEC61A2,PROSER2,NUDT5,PROSER2-AS1,DHTKD1 |
| 10 | 12581572 | 13208061 | GERD | MDD | 0.589087 | 0.000120158 | positive | OPTN,CCDC3,MCM10,CAMK1D,MIR548Q,MIR4481,MIR4480 |
| 10 | 19965625 | 20616432 | GERD | MDD | 0.764722 | 0.00271042 | positive | LOC101928834,PLXDC2,MALRD1 |
| 10 | 21477693 | 23365685 | GERD | MDD | 0.52891 | 0.00306976 | positive | MIR1915,COMMD3,SKIDA1,MIR1915HG,LINC02643,COMMD3-BMI1,LOC100499489,MLLT10,EBLN1,SPAG6,NEBL,BMI1,LOC100130992,ARMC3,DNAJC1,PIP4K2A |
| 10 | 33655459 | 35178224 | GERD | MDD | 0.487705 | 0.000969568 | positive | PARD3-AS1,PARD3,LINC00838 |
| 10 | 35178225 | 36475838 | GERD | MDD | 0.712926 | 0.000681366 | positive | MIR4683,CREM,CCNY,PCAT5,GJD4,FZD8,CUL2,MIR3611 |
| 10 | 62626466 | 64069687 | GERD | MDD | 0.447823 | 0.0011588 | positive | LINC00845,ARID5B,TMEM26-AS1,RHOBTB1,CABCOCO1,LOC107984236,TMEM26,RTKN2 |
| 10 | 67769823 | 68623274 | GERD | MDD | 0.737768 | 0.0060618 | positive | CTNNA3 |
| 10 | 72659224 | 73514214 | GERD | MDD | 0.595244 | 4.87761E-05 | positive | SLC29A3,UNC5B-AS1,LINC02622,C10orf105,CDH23-AS1,VSIR,UNC5B,CDH23 |
| 10 | 73514215 | 75694023 | GERD | MDD | 0.82695 | 2.77513E-05 | positive | CHCHD1,PLAU,MYOZ1,VSIR,MSS51,BMS1P4,C10orf55,CHST3,MIR7152,ANAPC16,FAM149B1,DNAJB12,ANXA7,MRPS16,DNAJC9,MICU1,USP54,PPP3CB-AS1,SYNPO2L,GLUD1P3,SPOCK2,PLA2G12B,PSAP,SEC24C,MIR4676,MCU,DNAJC9-AS1,CDH23,ZSWIM8,NDST2,SNORA11F,BMS1P4-AGAP5,PPP3CB,P4HA1,OIT3,ECD,ASCC1,ZSWIM8-AS1,AGAP5,FUT11,DDIT4,NUDT13,CAMK2G,CFAP70 |
| 10 | 77144962 | 78665481 | GERD | MDD | 0.590658 | 0.000209893 | positive | MIR606,LRMDA,KCNMA1,LOC101929234,ZNF503-AS2,LOC105378367,KCNMA1-AS1,ZNF503 |
| 10 | 83596351 | 84679429 | GERD | MDD | 0.55041 | 0.0139382 | positive | NRG3-AS1,NRG3 |
| 10 | 102509299 | 104206837 | GERD | MDD | 0.483758 | 0.016414 | positive | MIR3158-2,FBXL15,LBX1,PITX3,BTRC,FGF8,DPCD,PPRC1,LINC01514,KCNIP2-AS1,MRPL43,FBXW4,ELOVL3,CUEDC2,NPM3,OGA,PDZD7,ARMH3,GBF1,MIR146B,NFKB2,PSD,NOLC1,PAX2,KAZALD1,LDB1,TLX1NB,LBX1-AS1,KCNIP2,TWNK,POLL,MIR608,SEMA4G,HPS6,LZTS2,LINC02681,SLF2,SFXN3,TLX1,MIR3158-1 |
| 10 | 104206838 | 106142283 | GERD | MDD | 0.593873 | 0.000128983 | positive | MIR609,WBP1L,ITPRIP,CFAP58,CFAP43,LOC107984265,GSTO2,ACTR1A,PCGF6,CYP17A1,NEURL1-AS1,INA,TAF5,COL17A1,RPARP-AS1,CNNM2,C10orf95,NEURL1,GSTO1,ARL3,CFAP58-DT,TRIM8,MFSD13A,SUFU,CALHM1,CALHM2,BORCS7,SFR1,SLK,CALHM3,ATP5MD,NT5C2,SFXN2,PDCD11,AS3MT,SH3PXD2A-AS1,MIR4482,SH3PXD2A,MIR1307,RPEL1,STN1,BORCS7-ASMT,ITPRIP-AS1,MIR936 |
| 10 | 106142284 | 107877787 | GERD | MDD | 0.757478 | 1.67347E-07 | positive | LINC02627,CFAP58,SORCS3-AS1,SORCS3,LINC02620 |
| 10 | 107877788 | 108725997 | GERD | MDD | 0.905011 | 0.00111502 | positive | SORCS1,LINC02624 |
| 10 | 114255955 | 115588903 | GERD | MDD | 0.680483 | 1.51109E-06 | positive | MIR4295,SNORA87,MIR4483,HABP2,PLEKHS1,TCF7L2,CASP7,NRAP,LOC103344931,VTI1A |
| 10 | 119528731 | 120592839 | GERD | MDD | 0.525645 | 0.00811248 | positive | RAB11FIP2,CASC2,PRLHR,LINC00867,FAM204A,CACUL1 |
| 10 | 125798320 | 126875816 | GERD | MDD | 0.443606 | 0.00456098 | positive | ABRAXAS2,OAT,ZRANB1,CTBP2,MIR4296,LHPP,EEF1AKMT2,NKX1-2,FAM53B,CHST15,FAM53B-AS1 |
| 10 | 127997709 | 129134738 | GERD | MDD | 0.79572 | 0.000987691 | positive | LINC00601,C10orf90,ADAM12,DOCK1,INSYN2A,LOC728158 |
| 10 | 130694009 | 131694738 | GERD | MDD | 0.406955 | 0.00510273 | positive | LINC02667,MIR4297,EBF3,MGMT |
| 10 | 131694739 | 132576793 | GERD | MDD | 0.776391 | 0.00271245 | positive | C10orf143,EBF3,CTAGE7P,GLRX3 |
| 10 | 133818037 | 134856054 | GERD | MDD | 0.590131 | 0.000305183 | positive | DPYSL4,STK32C,PWWP2B,LINC02870,LINC01167,JAKMIP3,INPP5A,LRRC27,LOC107984282,CFAP46,LINC01168,LINC01166,NKX6-2 |
| 10 | 134856055 | 135524372 | GERD | MDD | 0.960053 | 6.83308E-07 | positive | PAOX,CALY,CYP2E1,MTG1,MIR202,ADAM8,FUOM,SYCE1,ECHS1,KNDC1,ADGRA1,TUBGCP2,SPRN,MIR202HG,SCART1,UTF1,MIR3944,SPRNP1,PRAP1,VENTX,ADGRA1-AS1,FRG2B,ZNF511 |
| 11 | 87150 | 1061399 | GERD | MDD | 0.644397 | 0.000755744 | positive | BET1L,MIR210HG,LOC143666,SIRT3,IFITM5,HRAS,PNPLA2,RIC8A,TMEM80,SCGB1C2,SLC25A22,PGGHG,GATD1,RASSF7,PHRF1,PIDD1,LMNTD2-AS1,CHID1,SIGIRR,CDHR5,EPS8L2,LOC171391,MIR210,DEAF1,ANO9,B4GALNT4,IFITM1,SCT,MUC6,PTDSS2,NLRP6,SCGB1C1,CEND1,LMNTD2,PKP3,DRD4,IRF7,SNORA52,TALDO1,MIR6743,IFITM3,IFITM2,POLR2L,PSMD13,CD151,ODF3,LINC02688,CRACR2B,AP2A2,TSPAN4,RPLP2,LINC01001,LRRC56,RNH1 |
| 11 | 11632651 | 12470449 | GERD | MDD | 0.745201 | 0.00126302 | positive | MIR6124,DKK3,PARVA,MICALCL,GALNT18,MIR4299,MICAL2,LINC02547,MIR8070,USP47 |
| 11 | 12470450 | 13364727 | GERD | MDD | 0.444782 | 0.0214194 | positive | PARVA,TEAD1,ARNTL,RASSF10,LINC00958 |
| 11 | 13364728 | 15126767 | GERD | MDD | 0.424703 | 0.0276866 | positive | LINC02548,SPON1,CALCA,LINC02545,LINC02683,BTBD10,CYP2R1,PSMA1,PDE3B,CALCB,FAR1,COPB1,ARNTL,RRAS2,PTH |
| 11 | 18740268 | 19780784 | GERD | MDD | 0.717182 | 9.41563E-05 | positive | E2F8,CSRP3,LOC100126784,NAV2,PTPN5,ZDHHC13,MRGPRX1,MIR4486,NAV2-AS5,IGSF22,MRGPRX2,NAV2-AS4 |
| 11 | 19780785 | 20686336 | GERD | MDD | 0.476368 | 0.0191861 | positive | SLC6A5,HTATIP2,NAV2-AS2,NAV2,MIR4694,DBX1,PRMT3 |
| 11 | 28577719 | 29530460 | GERD | MDD | 0.752806 | 0.00123867 | positive | NA |
| 11 | 29530461 | 30570129 | GERD | MDD | 0.662201 | 0.000424385 | positive | LINC01616,MPPED2,FSHB,LINC02546,ARL14EP,KCNA4 |
| 11 | 30570130 | 32065771 | GERD | MDD | 0.595476 | 6.93857E-05 | positive | MPPED2,ELP4,PAX6,PAX6-AS1,IMMP1L,LOC100506675,DNAJC24,DCDC1,PAUPAR |
| 11 | 32065772 | 32830422 | GERD | MDD | 0.374255 | 0.0291453 | positive | WT1,RCN1,SNORA88,CCDC73,EIF3M,WT1-AS |
| 11 | 39329141 | 40362940 | GERD | MDD | 0.495457 | 0.018526 | positive | LRRC4C |
| 11 | 43949378 | 45019559 | GERD | MDD | 0.517456 | 0.0172775 | positive | ALX4,TP53I11,LINC02685,CD82,C11orf96,ACCSL,EXT2,TSPAN18,ACCS |
| 11 | 56623495 | 57675986 | GERD | MDD | 1 | 0.000394971 | positive | PRG3,SELENOH,YPEL4,P2RX3,SLC43A1,CTNND1,TIMM10,BTBD18,OR5AK4P,PRG2,CLP1,UBE2L6,RTN4RL2,OR5AK2,APLNR,LINC02735,MED19,SMTNL1,SSRP1,SLC43A3,TMX2-CTNND1,MIR130A,LRRC55,SERPING1,TNKS1BP1,ZDHHC5,TMX2 |
| 11 | 57675987 | 58452708 | GERD | MDD | 0.850246 | 0.00128866 | positive | ZFP91,CNTF,OR5B21,OR1S1,ZFP91-CNTF,OR9I1,OR5B2,OR9Q1,OR5B3,OR6Q1,OR5B12,OR10W1,OR9Q2,LPXN,OR10Q1,OR1S2,OR5B17 |
| 11 | 62800369 | 64594822 | GERD | MDD | 0.637717 | 0.000619825 | positive | PLAAT3,MAP4K2,RCOR2,MIR1237,LOC114841035,STIP1,TRPT1,MARK2,CDC42BPG,TRMT112,LGALS12,KCNK4,SLC22A10,PYGM,MEN1,PPP1R14B,NUDT22,GPR137,PLAAT2,DNAJC4,RASGRP2,SLC22A24,SLC22A25,SLC22A11,RPS6KA4,PPP1R14B-AS1,SLC22A12,PLCB3,FERMT3,ATL3,BAD,SF1,PLAAT4,KCNK4-TEX40,PLAAT5,CCDC88B,OTUB1,LINC02724,NRXN2,CATSPERZ,SLC22A9,NAA40,MIR7155,COX8A,FLRT1,VEGFB,FKBP2,RTN3,C11orf95,ESRRA,PRDX5,SPINDOC,MACROD1 |
| 11 | 64594823 | 66782661 | GERD | MDD | 0.707192 | 9.88541E-07 | positive | PC,SLC22A20P,MIR194-2,SNX15,MIR612,C11orf68,YIF1A,RELA,EIF1AD,LOC101927789,TMEM262,FOSL1,KCNK7,CCDC85B,ZNRD2,RBM14-RBM4,BRMS1,MAJIN,PACS1,ATG2A,RBM4B,POLA2,KAT5,LOC728975,AP5B1,C11orf80,FRMD8,MRPL11,FIBP,EHBP1L1,MUS81,CCS,CTSW,SCYL1,CATSPER1,BATF2,MIR10392,TALAM1,CST6,CD248,TMEM151A,MIR6749,MIR6879,C11orf86,CDCA5,B4GAT1,NAALADL1,LOC101928069,SYVN1,MALAT1,SIPA1,ZFPL1,FAU,NPAS4,BANF1,RBM4,ACTN3,MIR194-2HG,SNORD13F,MIR6751,ZDHHC24,DPF2,SLC25A45,MIR1234,ZNHIT2,ARL2,MIR6750,CDC42BPG,SNX32,TM7SF2,GPHA2,LOC102724064,LRFN4,ZNRD2-AS1,CTSF,SPTBN2,CFL1,RCE1,ARL2-SNX15,RAB1B,EHD1,KLC2,CNIH2,RIN1,CAPN1,PCNX3,MIR4489,PPP2R5B,CCDC87,RNASEH2C,CDC42EP2,VPS51,MAP3K11,TIGD3,RBM14,FAM89B,MIR192,NEAT1,MASCRNA,SAC3D1,BBS1,MIR4690,OVOL1,TSGA10IP,EFEMP2,SF3B2,OVOL1-AS1,PELI3,SART1,SLC29A2,LTBP3,SPDYC,MRPL49,MIR3163,GAL3ST3,DPP3,DRAP1 |
| 11 | 75445254 | 76518906 | GERD | MDD | 0.536987 | 0.0188333 | positive | LOC101928837,DGAT2,LRRC32,UVRAG,EMSY,GUCY2EP,UVRAG-DT,WNT11,TSKU,THAP12,GVQW3,LOC283214 |
| 11 | 79802287 | 80726012 | GERD | MDD | 0.617296 | 0.0153182 | positive | LINC02720 |
| 11 | 84613230 | 85869059 | GERD | MDD | 0.758201 | 0.00920521 | positive | CREBZF,TMEM126A,CCDC89,CCDC83,DLG2,PICALM,TMEM126B,SYTL2 |
| 11 | 88359288 | 89209738 | GERD | MDD | 0.917542 | 8.15728E-05 | positive | GRM5,TYR,NOX4 |
| 11 | 95327211 | 96150134 | GERD | MDD | 0.570899 | 0.0143736 | positive | FAM76B,MTMR2,MIR1260B,JRKL,CEP57,CCDC82,MAML2 |
| 11 | 111117781 | 112755446 | GERD | MDD | 0.716132 | 0.0011596 | positive | HSPB2,COLCA2,HOATZ,BTG4,MIR34C,C11orf52,MIR34B,ALG9,BCO2,IL18,NKAPD1,CRYAB,PLET1,COLCA1,DIXDC1,LINC02762,C11orf53,POU2AF1,SDHD,SIK2,HSPB2-C11orf52,PTS,LOC100132078,FDXACB1,DLAT,LINC02763,LAYN,LOC100132686,TIMM8B,MIR4491,PPP2R1B,TEX12,C11orf1,LOC728196,PIH1D2,LINC02764 |
| 11 | 112755447 | 113889019 | GERD | MDD | 1 | 1.07163E-20 | positive | HTR3B,TTC12,NCAM1,NCAM1-AS1,TMPRSS5,HTR3A,ZW10,USP28,DRD2,LOC101928847,MIR4301,CLDN25,ANKK1 |
| 11 | 116247378 | 117295641 | GERD | MDD | 1 | 5.93031E-05 | positive | SIK3,BACE1,LOC100652768,SIDT2,APOC3,TAGLN,ZPR1,CEP164,BUD13,PCSK7,APOA4,BACE1-AS,APOA5,APOA1-AS,APOA1,LINC02702,RNF214,PAFAH1B2 |
| 11 | 117295642 | 118901213 | GERD | MDD | 0.463535 | 0.00198435 | positive | SMIM35,TREH,BCL9L,IL10RA,KMT2A,FXYD6,TTC36,RPL23AP64,UBE4A,PHLDB1,CXCR5,SCN4B,MIR6716,ATP5MG,MPZL2,RPS25,TMEM25,JAML,CD3G,DDX6,DSCAML1,FXYD6-FXYD2,MPZL3,ARCN1,FXYD2,IFT46,CENATAC,TMPRSS13,FOXR1,TRAPPC4,LOC100131626,SCN2B,CD3D,LOC101929089,UPK2,MIR4492,TMPRSS4,CD3E,SLC37A4 |
| 11 | 118901214 | 120064528 | GERD | MDD | 0.696551 | 0.00524193 | positive | NLRX1,HINFP,USP2,C1QTNF5,LOC102724301,H2AX,TRIM29,HYOU1,CCDC153,C2CD2L,MFRP,USP2-AS1,MIR6756,CBL,THY1,MCAM,ABCG4,LOC107984399,DPAGT1,VPS11,PDZD3,SNORD150,HMBS,SLC37A4,NECTIN1,RNF26 |
| 11 | 121208187 | 122589224 | GERD | MDD | 0.448698 | 0.00489266 | positive | MIRLET7A2,LOC105369535,BLID,MIR125B1,UBASH3B,MIR100,MIR10526,SORL1,MIR100HG |
| 11 | 124403478 | 125266684 | GERD | MDD | 0.949671 | 7.15506E-07 | positive | TBRG1,ESAM,MSANTD2,PANX3,SPA17,SIAE,PKNOX2-AS1,CCDC15,TMEM218,LOC101929340,VSIG2,ROBO4,OR8A1,SLC37A2,PKNOX2,OR8B12,HEPN1,ROBO3,NRGN,LOC100507283,HEPACAM |
| 11 | 126304241 | 127306871 | GERD | MDD | 0.397269 | 0.011627 | positive | LINC02712,LOC101929473,KIRREL3-AS3,KIRREL3,MIR3167,KIRREL3-AS2,LOC101929427 |
| 11 | 128262551 | 129451784 | GERD | MDD | 0.739164 | 0.00323168 | positive | ETS1-AS1,FLI1,BARX2,ARHGAP32,ETS1,SENCR,LOC101929538,MIR6090,LOC399975,KCNJ5,TP53AIP1,KCNJ1,C11orf45 |
| 11 | 131424362 | 132584401 | GERD | MDD | 0.441862 | 0.00201054 | positive | NTM-AS1,NTM,NTM-IT,OPCML |
| 11 | 133521643 | 134351064 | GERD | MDD | 0.619165 | 6.35626E-05 | positive | SNORD153,LINC02743,IGSF9B,THYN1,GLB1L3,VPS26B,B3GAT1-DT,MIR4697,JAM3,ACAD8,GLB1L2,LINC02731,SPATA19,NCAPD3,B3GAT1 |
| 12 | 60317 | 1078397 | GERD | MDD | 0.749422 | 0.00202576 | positive | SLC6A13,KDM5A,IQSEC3,WNK1,LOC574538,CCDC77,RAD52,SLC6A12-AS1,LOC102723544,LOC105369595,NINJ2,B4GALNT3,FAM138D,NINJ2-AS1,SLC6A12,WASH8P |
| 12 | 16136743 | 17374895 | GERD | MDD | 0.636161 | 0.00144205 | positive | SLC15A5,LMO3,SKP1P2,DERA,MGST1 |
| 12 | 17374896 | 18470023 | GERD | MDD | 0.505426 | 0.0105127 | positive | LINC02378,PIK3C2G,MIR3974,RERGL |
| 12 | 22928056 | 23923798 | GERD | MDD | 1 | 7.90025E-05 | positive | SOX5,LOC101928441 |
| 12 | 23923799 | 25058714 | GERD | MDD | 0.383835 | 0.00103508 | positive | SOX5,BCAT1,MIR920,LINC00477,SOX5-AS1 |
| 12 | 32368906 | 33076722 | GERD | MDD | 0.692406 | 0.00573425 | positive | BICD1,DNM1L,YARS2,PKP2,FGD4 |
| 12 | 48960266 | 50344346 | GERD | MDD | 0.606898 | 0.0168468 | positive | SNORA2A,ARF3,SPATS2,BCDIN3D-AS1,SNORA2B,KANSL2,ADCY6-DT,FKBP11,TROAP,CCDC65,MIR1291,KCNH3,TUBA1A,DNAJC22,DDN,MCRS1,RND1,TMBIM6,DDX23,LMBR1L,TEX49,FAIM2,BCDIN3D,PRKAG1,FMNL3,RHEBL1,WNT10B,NCKAP5L,WNT1,C1QL4,LOC100335030,KMT2D,PRPF40B,FAM186B,CCNT1,LINC02395,DHH,SNORA2C,DDN-AS1,MIR4701,TUBA1C,ADCY6,LALBA,LOC101927267,TUBA1B,LINC02396,PRPH,CACNB3 |
| 12 | 51769420 | 53039987 | GERD | MDD | 0.41735 | 0.0217776 | positive | GALNT6,KRT74,KRT85,OR7E47P,KRT84,KRT2,KRT72,ANKRD33,KRT73-AS1,KRT81,KRT71,KRT6A,KRT5,KRT87P,ATG101,KRT82,ACVRL1,KRT7,TAMALIN-AS1,LINC02874,SMIM41,KRT73,FIGNL2-DT,LINC00592,KRT86,SCN8A,KRT6C,KRT7-AS,NR4A1,KRT6B,TAMALIN,SLC4A8,KRT83,KRT75,KRT80,FIGNL2,ACVR1B |
| 12 | 60000364 | 61737953 | GERD | MDD | 0.567254 | 0.0176438 | positive | SLC16A7 |
| 12 | 61737954 | 63055936 | GERD | MDD | 0.723614 | 3.80424E-05 | positive | PPM1H,MIRLET7I,TAFA2,LINC01465,USP15,MIR6125,MON2 |
| 12 | 64403858 | 66114643 | GERD | MDD | 0.955982 | 0.00148105 | positive | TBK1,LINC02389,RASSF3,LINC02231,WIF1,C12orf56,TBC1D30,C12orf66,MSRB3,MIR548C,SRGAP1,GNS,LOC100507065,XPOT,LINC02454,LEMD3,MIR548Z,LOC105369187 |
| 12 | 67183621 | 68006682 | GERD | MDD | 0.811297 | 0.00798228 | positive | LINC02420,LINC02408,LOC102724421,GRIP1,CAND1 |
| 12 | 77800464 | 79315178 | GERD | MDD | 0.411704 | 0.00596637 | positive | SYT1,NAV3,LINC02424 |
| 12 | 96017783 | 96828314 | GERD | MDD | 0.723812 | 0.00832391 | positive | LOC105369921,LINC02410,HAL,CCDC38,PGAM1P5,SNRPF,AMDHD1,LTA4H,NTN4,ELK3,CDK17 |
| 12 | 102966634 | 104140831 | GERD | MDD | 0.631345 | 0.000121859 | positive | STAB2,LOC101929058,LOC105369945,PAH,ASCL1,LINC00485,C12orf42,LINC02401 |
| 12 | 106964978 | 108183188 | GERD | MDD | 0.611102 | 0.0117709 | positive | PWP1,CRY1,PRDM4,LOC100505978,BTBD11,TMEM263,RFX4,MTERF2,ASCL4,LOC101929162,RIC8B,LOC100287944 |
| 12 | 108183189 | 109031819 | GERD | MDD | 0.656543 | 0.00119872 | positive | MIR4496,SELPLG,TMEM119,SART3,ISCU,LOC728739,LINC01498,CMKLR1,WSCD2,FICD |
| 12 | 109031820 | 110111514 | GERD | MDD | 0.517413 | 0.0266388 | positive | USP30,SVOP,ALKBH2,MVK,LINC01486,UBE3B,FOXN4,SSH1,MIR619,MMAB,CORO1C,MYO1H,UNG,ACACB,KCTD10,DAO,USP30-AS1 |
| 12 | 111592382 | 113947983 | GERD | MDD | 0.430895 | 0.0194898 | positive | MIR6762,OAS1,DDX54,LHX5-AS1,CFAP73,ATXN2,TRAFD1,SH2B3,PTPN11,RPL6,IQCD,MIR6761,MIR3657,HECTD4,ATXN2-AS,RPH3A,MIR7106,MIR1302-1,ACAD10,MAPKAPK5,DTX1,PHETA1,TMEM116,PLBD2,SDS,OAS2,RITA1,OAS3,ALDH2,MAPKAPK5-AS1,CUX2,BRAP,TPCN1,ERP29,MIR6861,RASAL1,NAA25,MIR6760,SLC8B1,SDSL,LHX5 |
| 12 | 113947984 | 114685670 | GERD | MDD | 0.831032 | 0.00366321 | positive | RBM19,LINC02459,LINC01234 |
| 12 | 119301062 | 120567740 | GERD | MDD | 0.896847 | 3.26219E-05 | positive | RAB35,GCN1,PRKAB1,HSPB8,SRRM4,LINC00934,TMEM233,CCDC60,CIT,MIR1178,LOC105370024,BICDL1 |
| 12 | 120567741 | 121817509 | GERD | MDD | 0.686877 | 0.000401276 | positive | NRAV,PXN-AS1,RNF10,MIR4498,ACADS,RNU4-1,HNF1A,P2RX4,SPPL3,ANAPC5,UNC119B,RNU4-2,CABP1,HNF1A-AS1,P2RX7,SIRT4,POP5,MLEC,TRIAP1,PLA2G1B,COX6A1,RPLP0,MIR4700,CAMKK2,XLOC_009911,GCN1,OASL,GATC,C12orf43,COQ5,SRSF9,MSI1,DYNLL1,PXN |
| 12 | 121817510 | 123396634 | GERD | MDD | 0.806282 | 2.70172E-06 | positive | MIR9902-2,RSRC2,CLIP1,ZCCHC8,DIABLO,LRRC43,TMEM120B,CLIP1-AS1,CCDC62,HCAR3,VPS37B,KNTC1,KDM2B,LOC100506691,MIR9902-1,B3GNT4,IL31,RNF34,VPS33A,MORN3,ORAI1,DENR,HCAR2,PSMD9,MIR7107,LINC01089,RHOF,MIR548AQ,BCL7A,HIP1R,HCAR1,HPD,CFAP251,LOC101593348,MLXIP,SETD1B |
| 12 | 123396635 | 124843768 | GERD | MDD | 0.689634 | 2.17573E-05 | positive | MIR4304,C12orf65,ZNF664-RFLNA,SNORA9B,CDK2AP1,DNAH10,ZNF664,ABCB9,RFLNA,TMED2-DT,PITPNM2-AS1,NCOR2,ARL6IP4,SBNO1-AS1,EIF2B1,TMED2,RILPL1,MIR6880,OGFOD2,RILPL2,MPHOSPH9,PITPNM2,TCTN2,CCDC92,MIR3908,GTF2H3,ATP6V0A2,SBNO1,KMT5A,MIR8072,SNRNP35,DDX55 |
| 12 | 129170297 | 129887418 | GERD | MDD | 0.595196 | 0.00442383 | positive | TMEM132D-AS2,SLC15A4,TMEM132C,TMEM132D-AS1,GLT1D1,TMEM132D,LOC100128276 |
| 12 | 129887419 | 130629840 | GERD | MDD | 0.503538 | 0.000589104 | positive | LINC02418,TMEM132D,LINC02419 |
| 13 | 21540734 | 22795188 | GERD | MDD | 0.539908 | 0.000219649 | positive | LOC101928764,ZDHHC20,GRK6P1,LINC00539,SKA3,LATS2,MICU2,SAP18,LINC00424,FGF9,LINC01046,MRPL57,LINC00540,MIPEPP3 |
| 13 | 30068526 | 31433218 | GERD | MDD | 0.501206 | 0.00186936 | positive | LINC00384,KATNAL1,SLC7A1,LINC00365,ALOX5AP,UBE2L5,LINC00426,LINC00544,UBL3,LINC00398,LINC00297,USPL1,LINC01058,LINC00572,HMGB1,MTUS2,LOC102723345 |
| 13 | 31433219 | 32986686 | GERD | MDD | 0.645893 | 0.000110406 | positive | RXFP2,BRCA2,ZAR1L,TEX26-AS1,MEDAG,TEX26,N4BP2L1,LINC00545,FRY-AS1,B3GLCT,HSPH1,FRY,EEF1DP3 |
| 13 | 32986687 | 34722391 | GERD | MDD | 0.601076 | 0.000271879 | positive | N4BP2L2,MICOS10P1,N4BP2L2-IT2,LINC00423,PDS5B,N4BP2L1,RFC3,KL,STARD13,STARD13-AS,LINC02344 |
| 13 | 40091624 | 41003943 | GERD | MDD | 0.401375 | 0.0127325 | positive | LINC00598,LINC00548,MIR4305,LINC00332,LHFPL6,COG6 |
| 13 | 49381728 | 50419896 | GERD | MDD | 0.447568 | 0.0280275 | positive | RCBTB1,SETDB2,ARL11,PHF11,SETDB2-PHF11,CAB39L,FNDC3A,LOC105370203,MLNR,KPNA3,EBPL,CDADC1 |
| 13 | 53336572 | 54684856 | GERD | MDD | 0.893086 | 1.17694E-06 | positive | LINC01065,MIR759,OLFM4,PCDH8,LINC00558 |
| 13 | 54684857 | 55576912 | GERD | MDD | 1 | 5.35671E-05 | positive | LINC00458,MIR1297 |
| 13 | 58245844 | 59751795 | GERD | MDD | 0.903816 | 5.73106E-06 | positive | LINC00374,PCDH17,LINC02338 |
| 13 | 59751796 | 61268155 | GERD | MDD | 0.555869 | 6.65459E-05 | positive | DIAPH3-AS2,LINC00434,TDRD3,LINC00378,DIAPH3,DIAPH3-AS1 |
| 13 | 61268156 | 62135827 | GERD | MDD | 0.630752 | 0.0117406 | positive | LINC02339,LINC00378,MIR3169,PCDH20 |
| 13 | 66382287 | 67718879 | GERD | MDD | 0.603636 | 2.6491E-06 | positive | PCDH9,PCDH9-AS2,LINC01052,PCDH9-AS4,MIR548X2,MIR4704,PCDH9-AS3 |
| 13 | 67718880 | 69004702 | GERD | MDD | 0.590117 | 0.0133685 | positive | PCDH9,LINC00364 |
| 13 | 69004703 | 69751491 | GERD | MDD | 0.770393 | 0.000650872 | positive | LINC00550 |
| 13 | 75882385 | 76671493 | GERD | MDD | 0.674703 | 0.0138123 | positive | LMO7DN-IT1,UCHL3,LMO7-AS1,LMO7,LINC00561,LMO7DN,TBC1D4,COMMD6 |
| 13 | 79489046 | 80778339 | GERD | MDD | 0.718657 | 0.00260298 | positive | LINC01068,NDFIP2,LINC01080,NDFIP2-AS1,RBM26,LINC00382,LINC01038,RBM26-AS1 |
| 13 | 80778340 | 81724089 | GERD | MDD | 0.636343 | 0.000179806 | positive | LINC00377,SPRY2 |
| 13 | 83829836 | 85035400 | GERD | MDD | 0.734424 | 0.00185636 | positive | SLITRK1,LINC00333 |
| 13 | 85035401 | 85650680 | GERD | MDD | 1 | 0.00000444 | positive | LINC00375,SNORA107,LINC00333 |
| 13 | 96132648 | 97516609 | GERD | MDD | 0.832728 | 0.00364386 | positive | UGGT2,MIR4501,DNAJC3,DNAJC3-DT,DZIP1,CLDN10-AS1,CLDN10,HS6ST3 |
| 13 | 98527336 | 99402294 | GERD | MDD | 0.894596 | 0.00390911 | positive | STK24,FARP1,STK24-AS1,RNF113B,SLC15A1,MIR3170,IPO5 |
| 13 | 100470869 | 101573736 | GERD | MDD | 0.393621 | 0.0269117 | positive | TMTC4,ZIC2,PCCA-DT,CLYBL,GGACT,NALCN-AS1,PCCA-AS1,LOC101927437,LINC00554,PCCA,ZIC5 |
| 13 | 101573737 | 103050417 | GERD | MDD | 0.606085 | 0.00478958 | positive | FGF14-IT1,MIR4705,FGF14-AS1,FGF14,NALCN-AS1,LINC00411,NALCN,MIR2681,ITGBL1,FGF14-AS2 |
| 13 | 103050418 | 104378435 | GERD | MDD | 0.429116 | 0.00522098 | positive | METTL21C,CCDC168,POGLUT2,METTL21EP,FGF14,ERCC5,TPP2,LINC01309,SLC10A2,BIVM,BIVM-ERCC5,TEX30 |
| 13 | 104378436 | 105672500 | GERD | MDD | 0.670758 | 0.000230239 | positive | NA |
| 13 | 107037865 | 108521978 | GERD | MDD | 0.696571 | 7.30816E-06 | positive | EFNB2,SNORD31B,LINC00551,ARGLU1,FAM155A,MIR1267,LINC00443,FAM155A-IT1 |
| 13 | 109813577 | 110995432 | GERD | MDD | 0.436083 | 0.0238474 | positive | COL4A1,LINC00676,MYO16,COL4A2,LINC00396,MIR8073,IRS2,LINC00399,MYO16-AS1,LOC105370361 |
| 13 | 110995433 | 111621244 | GERD | MDD | 0.54092 | 0.00290492 | positive | RAB20,COL4A2-AS1,COL4A2,LINC00567,ING1,ANKRD10,PRECSIT,COL4A2-AS2,CARS2,NAXD,LOC105370362,LINC00431 |
| 13 | 111621245 | 112319064 | GERD | MDD | 0.929715 | 0.000274971 | positive | LINC02337,LOC101060553,ARHGEF7-AS1,LINC00368,ARHGEF7,ARHGEF7-AS2,TEX29,LINC00431 |
| 13 | 112319065 | 113573118 | GERD | MDD | 0.428714 | 0.00249595 | positive | LINC00354,ATP11A,LOC101928730,SOX1-OT,LOC100506016,TUBGCP3,ATP11A-AS1,SPACA7,LINC01070,LINC01044,ATP11AUN,SOX1,LINC01043 |
| 14 | 22760701 | 23985936 | GERD | MDD | 0.55276 | 0.0013531 | positive | PSMB5,MYH7,MHRT,LMLN2,LOC101926933,CDH24,OXA1L,LOC105370705,PSMB11,ABHD4,LINC02332,MIR4707,MIR208B,ACIN1,MMP14,MIR208A,LRP10,MYH6,HOMEZ,CMTM5,PRMT5,RBM23,RNF212B,SLC7A8,NGDN,HAUS4,PRMT5-AS1,THTPA,EFS,BCL2L2,CEBPE,PABPN1,SLC7A7,AJUBA,PPP1R3E,IL25,C14orf93,REM2,SLC22A17,DAD1,C14orf119,BCL2L2-PABPN1,LOC105370401,OR6J1,MRPL52 |
| 14 | 26395691 | 27720240 | GERD | MDD | 0.457404 | 0.0143408 | positive | MIR4307,LINC02294,LINC02588,NOVA1,MIR4307HG,LINC02293 |
| 14 | 29029225 | 30831154 | GERD | MDD | 0.822085 | 1.80945E-08 | positive | LINC01551,LINC02282,PRKD1,FOXG1-AS1,FOXG1 |
| 14 | 33591114 | 34695195 | GERD | MDD | 0.69774 | 1.40583E-08 | positive | EGLN3,NPAS3,SNORA89 |
| 14 | 36422078 | 37282012 | GERD | MDD | 0.679602 | 0.00189386 | positive | MBIP,SFTA3,LINC00609,PTCSC3,PAX9,NKX2-1-AS1,SLC25A21,NKX2-1,NKX2-8 |
| 14 | 38722115 | 40343754 | GERD | MDD | 0.436281 | 0.0275776 | positive | GEMIN2,SEC23A,TRAPPC6B,CLEC14A,LOC112268136,LOC100288846,FBXO33,LINC00639,MIA2,PNN,SEC23A-AS1,LOC105370457 |
| 14 | 41614834 | 42562550 | GERD | MDD | 0.787411 | 5.9932E-06 | positive | LRFN5 |
| 14 | 42562551 | 43321168 | GERD | MDD | 0.58358 | 0.0173529 | positive | NA |
| 14 | 46748239 | 48177483 | GERD | MDD | 0.641024 | 0.00164276 | positive | MDGA2,LINC00871,RPL10L |
| 14 | 49002066 | 50500564 | GERD | MDD | 0.77985 | 0.000350601 | positive | RN7SL2,MIR6076,POLE2,RPL36AL,RN7SL3,KLHDC1,MGAT2,ARF6,DNAAF2,LINC01599,NEMF,RN7SL1,KLHDC2,RPS29,LRR1,LINC01588 |
| 14 | 56206431 | 57460781 | GERD | MDD | 0.436022 | 0.00340563 | positive | OTX2-AS1,TMEM260,OTX2,RPL13AP3,LOC101927690,PELI2,LINC00520 |
| 14 | 58447799 | 59447537 | GERD | MDD | 0.571786 | 0.00693378 | positive | TOMM20L,KIAA0586,LINC00216,PSMA3,ACTR10,ARMH4,TIMM9,LINC01500,ARID4A,PSMA3-AS1,DACT1 |
| 14 | 64033238 | 65229391 | GERD | MDD | 0.874789 | 0.000880951 | positive | ZBTB1,PLEKHG3,LOC102723809,ESR2,WDR89,HSPA2,MTHFD1,SPTB,SGPP1,PPP1R36,TEX21P,ZBTB25,SYNE2,MIR548H1,AKAP5 |
| 14 | 68976913 | 69794542 | GERD | MDD | 0.575932 | 0.00111672 | positive | ACTN1,ACTN1-AS1,EXD2,ZFP36L1,LOC100996664,DCAF5,RAD51B,GALNT16 |
| 14 | 74245429 | 75699501 | GERD | MDD | 0.777442 | 6.36144E-06 | positive | ENTPD5,YLPM1,SYNDIG1L,FCF1,COQ6,ISCA2,LIN52,LTBP2,MLH3,ZC2HC1C,BBOF1,PTGR2,LINC02274,EIF2B2,NEK9,LOC100506476,ZNF410,DLST,PROX2,ALDH6A1,ACYP1,ABCD4,AREL1,PGF,NPC2,FAM161B,TMED10,MIR4709,VSX2,RPS6KL1,MIDEAS,VRTN |
| 14 | 78374054 | 79199451 | GERD | MDD | 0.639144 | 0.00347764 | positive | ADCK1,NRXN3 |
| 14 | 88208596 | 89482883 | GERD | MDD | 0.636917 | 0.0117559 | positive | EML5,ZC3H14,LINC01146,GALC,KCNK10,SPATA7,LINC01147,TTC8,PTPN21,GPR65 |
| 14 | 89482884 | 90662836 | GERD | MDD | 0.829479 | 0.00159064 | positive | KCNK13,FOXN3-AS1,TDP1,EFCAB11,FOXN3,FOXN3-AS2 |
| 14 | 90662837 | 92101228 | GERD | MDD | 0.468305 | 0.0164776 | positive | TTC7B,RPS6KA5,SNORA11B,CATSPERB,CCDC88C,LOC101928909,LINC02317,CALM1,DGLUCY,PPP4R3A,GPR68,LINC00642,PSMC1,NRDE2,LOC105370619,LOC105370622 |
| 14 | 93386329 | 94892240 | GERD | MDD | 0.356202 | 0.0134151 | positive | BTBD7,CCDC197,SERPINA10,ASB2,IFI27L1,MOAP1,IFI27L2,SERPINA6,PPP4R4,MIR4506,IFI27,ITPK1-AS1,SERPINA2,UNC79,OTUB2,CHGA,COX8C,GON7,PRIMA1,ITPK1,FAM181A-AS1,FAM181A,TMEM251,SERPINA1,UBR7,DDX24 |
| 14 | 98268392 | 98885322 | GERD | MDD | 0.458263 | 0.00635328 | positive | LINC01550 |
| 14 | 99474534 | 100786189 | GERD | MDD | 0.450275 | 0.00347887 | positive | CCDC85C,SLC25A29,CCNK,MIR6764,MIR342,SETD3,EVL,EML1,MIR345,BCL11B,DEGS2,YY1,CYP46A1,HHIPL1,MIR151B |
| 14 | 100786190 | 102004377 | GERD | MDD | 0.491187 | 0.000238334 | positive | MIR544A,MIR329-1,MEG8,MEG9,SNORD114-29,WARS1,MIR369,MIR382,MIR409,MIR136,LINC02285,MIR370,SNORD114-10,SNORD113-7,SNORD114-19,MIR1193,MIR381HG,MIR134,MIR656,MIR431,SNORD113-2,MIR487A,LINC02314,MEG3,MIR379,SNORD114-11,RTL1,MIR770,LINC00524,SNORD114-18,SNORD114-26,MIR376A1,SNORD114-2,SNORD114-12,SNORD114-16,MIR496,BEGAIN,SNORD114-3,MIR432,MIR376B,MIR758,SNORD114-14,WDR25,SNORD114-4,MIR433,MIR337,MIR376C,MIR410,SNORD113-9,SNORD114-25,MIR889,SNORD114-31,SNORD113-6,SNORD114-20,SNORD114-24,SNORD114-23,SNORD113-8,MIR1185-2,SNORD114-9,SNORD114-5,LINC00523,SNORD114-7,MIR127,SNORD114-6,MIR323B,MIR411,SNORD114-15,SLC25A47,MIR655,SNORD114-27,SNORD114-28,MIR1185-1,SNORD114-1,SNORD113-4,SNORD113-5,SNORD114-22,SNORD113-3,SNORD114-13,MIR377,MIR376A2,MIR665,MIR380,MIR412,MIR487B,MIR539,MIR300,DLK1,MIR543,SNORD112,SNORD114-21,SNORD113-1,MIR329-2,MIR493,MIR323A,MIR668,MIR2392,SNORD114-30,MIR494,MIR485,MIR1197,MIR495,MIR299,SNORD114-17,MIR541,MIR654,SNORD114-8,MIR381,MIR154 |
| 14 | 102004378 | 103617472 | GERD | MDD | 0.43154 | 0.0143233 | positive | CINP,TNFAIP2,MIR1247,DIO3,ANKRD9,LINC00677,RCOR1,PPP2R5C,LINC00239,EXOC3L4,TECPR2,WDR20,MIR4309,CDC42BPB,MOK,LINC02323,TRAF3,DYNC1H1,LBHD2,ZNF839,HSP90AA1,DIO3OS,AMN |
| 15 | 36583328 | 37962915 | GERD | MDD | 0.434956 | 0.0019284 | positive | CDIN1,MEIS2,LOC145845,CSNK1A1P1,MIR8063 |
| 15 | 37962916 | 39238840 | GERD | MDD | 0.701926 | 8.09433E-08 | positive | LINC02345,LINC01852,FAM98B,TMCO5A,RASGRP1,LINC02694,SPRED1 |
| 15 | 39238841 | 40604780 | GERD | MDD | 0.384304 | 0.0273084 | positive | C15orf56,EIF2AK4,BUB1B-PAK6,BUB1B,PLCB2,FSIP1,SRP14,BMF,SRP14-AS1,C15orf54,LOC105370941,GPR176,ANKRD63,THBS1,PAK6 |
| 15 | 40604781 | 42332685 | GERD | MDD | 0.807379 | 1.93597E-05 | positive | VPS18,GCHFR,PPP1R14D,TYRO3,RPAP1,CHST14,PHGR1,NUSAP1,INO80,DNAJC17,JMJD7,RHOV,MRPL42P5,DISP2,EXD1,RMDN3,CHP1,C15orf62,OIP5,RPUSD2,CCDC32,DLL4,EHD4,IVD,KNL1,BAHD1,LOC105370943,OIP5-AS1,ITPKA,RAD51,CCDC9B,EHD4-AS1,MIR4310,NDUFAF1,SPINT1-AS1,INAFM2,RTF1,RAD51-AS1,MGA,MAPKBP1,PLA2G4B,MIR626,SPTBN5,PLA2G4E,KNSTRN,JMJD7-PLA2G4B,ZFYVE19,SPINT1,LOC105370792,CHAC1,LTK,PLA2G4E-AS1 |
| 15 | 47693317 | 49260006 | GERD | MDD | 0.579314 | 0.00398996 | positive | MYEF2,LINC01491,FBN1,SLC12A1,SHC4,EID1,SEMA6D,CTXN2,SLC24A5,CEP152,DUT |
| 15 | 53263127 | 54332799 | GERD | MDD | 0.87461 | 0.00340941 | positive | WDR72,LINC02490,UNC13C |
| 15 | 56322589 | 57641164 | GERD | MDD | 0.926366 | 4.76296E-05 | positive | TCF12,SNORD13D,ZNF280D,LINC01413,TEX9,MNS1,LINC00926,LOC145783,RFX7 |
| 15 | 63218979 | 64339945 | GERD | MDD | 0.657631 | 0.0112961 | positive | RAB8B,RPS27L,FBXL22,TPM1-AS,TPM1,HERC1,LOC101928988,DAPK2,USP3-AS1,APH1B,MIR422A,LACTB,CA12,LINC02568,USP3 |
| 15 | 64339946 | 66355634 | GERD | MDD | 0.834945 | 0.000157083 | positive | PLEKHO2,PDCD7,IGDCC4,ZNF609,RNU5B-1,CIAO2A,PIF1,SLC24A1,IGDCC3,DPP8,SLC51B,KBTBD13,PPIB,RNU5A-1,RBPMS2,MIR4511,CSNK1G1,PARP16,MIR4311,DENND4A,SNX1,UBAP1L,PCLAF,INTS14,ANKDD1A,OAZ2,TRIP4,SNX22,MEGF11,MIR1272,RAB11A,CILP,SNORA24B,SNORD13E,SPG21,CLPX,HACD3,MTFMT,RASL12 |
| 15 | 66355635 | 67396520 | GERD | MDD | 0.577078 | 0.000566344 | positive | LINC01169,SNORD18A,SMAD6,LOC102723493,DIS3L,TIPIN,MAP2K1,SNAPC5,SNORD18C,LINC02206,SCARNA14,MIR4512,SMAD3,SNORD18B,LCTL,RPL4,MEGF11,ZWILCH,SNORD16 |
| 15 | 67396521 | 69089815 | GERD | MDD | 0.707199 | 0.000648329 | positive | RNU6-9,MAP2K5,FEM1B,CORO2B,AAGAB,ANP32A,CLN6,C15orf61,CALML4,RNU6-1,SKOR1,SMAD3,IQCH-AS1,IQCH,LOC101929076,RNU6-7,ITGA11,RNU6-2,RNU6-8,PIAS1 |
| 15 | 69089816 | 70767983 | GERD | MDD | 0.70166 | 5.05994E-09 | positive | EWSAT1,TLE3,PAQR5,PCAT29,GLCE,RPLP1,LOC145694,NOX5,KIF23,ANP32A-IT1,MIR4312,LINC00593,ANP32A,DRAIC,MIR629,SPESP1 |
| 15 | 73375719 | 74458113 | GERD | MDD | 0.691615 | 0.00058818 | positive | INSYN1,STOML1,NPTN-IT1,LOXL1,REC114,GOLGA6A,ISLR2,INSYN1-AS1,COMMD4P2,LOXL1-AS1,MIR12135,NPTN,HCN4,CD276,LOC283731,PML,NEO1,TBC1D21 |
| 15 | 82191078 | 84209760 | GERD | MDD | 0.55814 | 0.0208563 | positive | LINC01583,RPS17,ACTG1P17,FSD2,C15orf40,SCARNA15,WHAMM,GOLGA6L10,HDGFL3,SNHG21,TM6SF1,HOMER2,BTBD1,EFL1,MEX3B,UBE2Q2P2,CPEB1,GOLGA6L9,AP3B2,ADAMTS7P1,LOC727751,LOC338963,SH3GL3,SAXO2,BNC1,MIR4515,RAMAC,GOLGA2P10,CPEB1-AS1,LOC102724034,GOLGA6L17P |
| 15 | 88035758 | 89385187 | GERD | MDD | 0.932356 | 4.57791E-06 | positive | NTRK3,MRPL46,NTRK3-AS1,MIR7-2,ACAN,ISG20,MRPS11,DET1,LINC01586,MIR1179,MIR3529,MIR11181,LINC00052,AEN |
| 15 | 89385188 | 90632718 | GERD | MDD | 0.728802 | 1.50275E-06 | positive | MFGE8,MIR6766,ZNF710-AS1,ACAN,ANPEP,LINC00928,IDH2,FANCI,ARPIN,ARPIN-AP3S2,RLBP1,MIR9-3,MIR9-3HG,PLIN1,PEX11A,ZNF710,MIR3174,HAPLN3,WDR93,ABHD2,MESP2,AP3S2,POLG,TICRR,MIR5094,MIR5009,RHCG,MESP1,KIF7,LOC105371031 |
| 15 | 90632719 | 91593623 | GERD | MDD | 0.421319 | 0.0100823 | positive | IQGAP1,GDPGP1,FURIN,CRTC3,SEMA4B,IDH2-DT,PRC1-AS1,BLM,IDH2,LINC01585,VPS33B-DT,ZNF774,FES,RCCD1,MAN2A2,TTLL13P,GABARAPL3,VPS33B,UNC45A,CRTC3-AS1,PRC1,HDDC3,CIB1,NGRN |
| 15 | 91593624 | 92675405 | GERD | MDD | 0.710582 | 9.44602E-06 | positive | SV2B,SLCO3A1,CRAT37 |
| 15 | 96864279 | 98025684 | GERD | MDD | 0.632449 | 2.57818E-07 | positive | NR2F2-AS1,NR2F2,LINC02254,SPATA8-AS1,MIR1469,SPATA8 |
| 15 | 100742118 | 101550750 | GERD | MDD | 0.698167 | 0.00352926 | positive | SPATA41,ASB7,LINS1,CERS3-AS1,LOC105369201,LOC102723335,ADAMTS17,ALDH1A3,PRKXP1,CERS3,LOC101927751,LRRK1 |
| 16 | 60169 | 1049079 | GERD | MDD | 0.539151 | 0.000116803 | positive | CEROX1,WFIKKN1,RPUSD1,LOC100287175,LUC7L,PGAP6,DDX11L10,WDR24,HAGHL,JMJD8,ARHGDIG,POLR3K,MIR662,LINC02867,NME4,SNRNP25,NHLRC4,CHTF18,MIR3176,MIR6859-4,METTL26,ANTKMT,RHOT2,HBM,HBA2,HBA1,STUB1,CIAO3,HBZ,HBQ1,PRR35,RHBDF1,FBXL16,MPG,METRN,PIGQ,CAPN15,LOC105371038,DECR2,RAB11FIP3,RGS11,LINC00235,NPRL3,GNG13,CCDC78,PRR25,MSLN,MIR5587,PDIA2,RAB40C,MIR6859-2,FAM234A,AXIN1,SOX8,MRPL28,LMF1,LMF1-AS1,MCRIP2,WASIR2,RHBDL1,MIR6859-1,LOC100134368,MIR6859-3,WDR90 |
| 16 | 1049080 | 1996492 | GERD | MDD | 0.656338 | 0.00270431 | positive | MAPK8IP3,MIR3177,CLCN7,LOC105371046,TMEM204,TPSAB1,FAHD1,C16orf91,TPSG1,CACNA1H,JPT2,IFT140,MRPS34,RPL3L,HS3ST6,TELO2,GNPTG,UNKL,PERCC1,UBE2I,CCDC154,PTX4,NME3,IGFALS,LINC02124,SSTR5-AS1,NUBP2,C1QTNF8,BAIAP3,EME2,LINC00254,MEIOB,SPSB3,TPSB2,SSTR5,CRAMP1,TPSD1,MSRB1,TSR3,HAGH |
| 16 | 3379997 | 4816145 | GERD | MDD | 0.682683 | 1.26035E-07 | positive | LINC01569,ZNF597,ZNF174,ADCY9,CORO7-PAM16,GLIS2-AS1,ANKS3,LINC02861,MGRN1,DNAJA3,VASN,MTRNR2L4,CLUAP1,NLRC3,ZSCAN32,PAM16,SLX4,UBALD1,NAA60,DNASE1,CDIP1,GLIS2,TFAP4,NUDT16L1,C16orf90,CORO7,MIR6126,NMRAL1,SRL,C16orf96,C16orf71,CREBBP,HMOX2,ZNF500,OR2C1,TRAP1,MIR6769A |
| 16 | 5782969 | 6446081 | GERD | MDD | 0.506215 | 4.07615E-06 | positive | RBFOX1 |
| 16 | 6446082 | 7051429 | GERD | MDD | 0.380074 | 0.0183466 | positive | RBFOX1 |
| 16 | 7051430 | 7704354 | GERD | MDD | 0.512042 | 0.00017782 | positive | RBFOX1 |
| 16 | 8341013 | 9104469 | GERD | MDD | 0.624601 | 0.00118156 | positive | ABAT,PMM2,TMEM186,USP7,CARHSP1,METTL22,TMEM114,LOC100130283 |
| 16 | 9104470 | 9754796 | GERD | MDD | 0.451256 | 0.0207761 | positive | LINC01195,LINC02177,C16orf72,LINC01177 |
| 16 | 12793150 | 13893407 | GERD | MDD | 0.410404 | 0.00354973 | positive | MIR4718,CPPED1,SHISA9 |
| 16 | 17343278 | 19104257 | GERD | MDD | 0.558309 | 0.0083579 | positive | COQ7,MIR6511A1,MIR3180-2,PKD1P5-LOC105376752,MIR3180-1,LOC100288162,MIR6511A4,MIR6511A2,MIR3670-2,XYLT1,MIR6511A3,MIR6770-2,LOC102723385,NPIPA7,MIR3180-3,MIR3179-2,RPS15A,NOMO2,TMC7,PKD1P4-NPIPA8,MIR3179-4,ARL6IP1,MIR3670-3,MIR6770-3,MIR6770-1,MIR3179-1,MIR3670-1,NPIPA8,MIR3670-4,SMG1,ABCC6P1,MIR3179-3 |
| 16 | 22667049 | 24014705 | GERD | MDD | 0.804409 | 0.00223567 | positive | DCTN5,SCNN1G,HS3ST2,SCNN1B,PRKCB,USP31,EARS2,PLK1,UBFD1,PALB2,NDUFAB1,CHP2,ERN2,GGA2,COG7 |
| 16 | 24014706 | 25235251 | GERD | MDD | 0.452861 | 0.000834199 | positive | MIR1273H,TNRC6A,ARHGAP17,LINC01567,LCMT1-AS2,LOC554206,LCMT1,LCMT1-AS1,AQP8,PRKCB,RBBP6,LINC02175,SLC5A11,LINC02194,CACNG3 |
| 16 | 25235252 | 26154122 | GERD | MDD | 0.69311 | 1.3867E-07 | positive | HS3ST4,AQP8,MIR548W,ZKSCAN2 |
| 16 | 26154123 | 27443061 | GERD | MDD | 0.546172 | 8.41067E-05 | positive | NSMCE1,KDM8,IL4R,C16orf82,NSMCE1-DT,IL21R |
| 16 | 35259454 | 49010761 | GERD | MDD | 0.822576 | 0.000454866 | positive | ITFG1,ORC6,ANKRD26P1,LINC02133,SHCBP1,GPT2,LINC02192,ITFG1-AS1,C16orf87,SIAH1,LINC02134,SNORD148,VPS35,PHKB,LONP2,NETO2,ABCC12,MYLK3,N4BP1,DNAJA2,ABCC11 |
| 16 | 49010762 | 50467033 | GERD | MDD | 0.766205 | 1.26878E-10 | positive | TENT4B,LINC02178,CBLN1,ADAM3B,ADCY7,ZNF423,CNEP1R1,HEATR3,C16orf78,BRD7,MIR6771 |
| 16 | 52041337 | 53393882 | GERD | MDD | 0.400185 | 0.00870525 | positive | CHD9,CASC16,LINC02180,LINC00919,TOX3,CASC22,LOC105371267 |
| 16 | 53393883 | 54866095 | GERD | MDD | 0.435374 | 3.08803E-07 | positive | FTO-IT1,RPGRIP1L,IRX3,LINC02140,FTO,LOC102723373,LOC643802,LINC02169,AKTIP,RBL2 |
| 16 | 54866096 | 55907389 | GERD | MDD | 0.78294 | 0.000148025 | positive | IRX5,CES1P2,SLC6A2,CES5A,CES1,CES1P1,LPCAT2,LOC101927480,CAPNS2,MMP2-AS1,IRX6,MMP2,CRNDE |
| 16 | 55907390 | 57119889 | GERD | MDD | 0.472434 | 0.0215276 | positive | NLRC5,MT1IP,MT2A,MT1JP,MIR6863,MT1A,MT1G,AMFR,MIR3935,GNAO1,NUP93,MT1DP,MT1X,NUDT21,SLC12A3,MT1F,MT1E,MT1M,CETP,OGFOD1,MT1B,MT1H,MT4,CES5A,LOC283856,HERPUD1,MIR138-2,MT1L,BBS2,MT3,DKFZP434H168 |
| 16 | 57119890 | 58508978 | GERD | MDD | 0.60791 | 0.000655616 | positive | CCDC113,ADGRG3,ADGRG5,CNGB1,MIR6772,ARL2BP,POLR2C,CSNK2A2,PRSS54,CCL22,PLLP,GINS3,KATNB1,LOC388282,CFAP20,CPNE2,ADGRG1,NDRG4,DOK4,COQ9,DRC7,CX3CL1,ZNF319,USB1,MMP15,TEPP,KIFC3,CIAPIN1,PSME3IP1,CCDC102A,CCL17,RSPRY1 |
| 16 | 59885500 | 61092844 | GERD | MDD | 0.762238 | 1.99267E-06 | positive | LINC02141,MIR4426 |
| 16 | 61092845 | 62607090 | GERD | MDD | 0.706602 | 0.00215091 | positive | CDH8 |
| 16 | 62607091 | 64207176 | GERD | MDD | 0.710551 | 0.000108073 | positive | NA |
| 16 | 65072404 | 65882820 | GERD | MDD | 0.525474 | 0.0137029 | positive | CDH11,LINC02126,LINC00922 |
| 16 | 68435729 | 69549149 | GERD | MDD | 0.673763 | 0.0141156 | positive | NIP7,VPS4A,TANGO6,CDH1,SNTB2,DERPC,CHTF8,TERF2,SMPD3,CDH3,TMED6,CYB5B,UTP4,HAS3,ZFP90,PDF,COG8 |
| 16 | 69549150 | 71062712 | GERD | MDD | 0.734166 | 0.00650885 | positive | DDX19A,CLEC18C,LOC105371328,VAC14-AS1,DDX19B,IL34,WWP2,EXOSC6,SF3B3,MIR1972-2,SMG1P7,HYDIN,VAC14,NOB1,CLEC18A,LOC100506083,PDPR,ST3GAL2,MIR1538,MIR140,AARS1,SNORD111B,LOC400541,SNORD111,SNORD13H,FCSK,NFAT5,NQO1,MIR1972-1,COG4,PDXDC2P-NPIPB14P,MTSS2 |
| 16 | 73140782 | 74135275 | GERD | MDD | 0.461081 | 0.00570597 | positive | ZFHX3,LINC01568 |
| 16 | 77270666 | 77739132 | GERD | MDD | 1 | 2.59686E-05 | positive | ADAMTS18 |
| 16 | 78973581 | 79662634 | GERD | MDD | 0.483806 | 0.00133442 | positive | MAF,WWOX |
| 16 | 81437862 | 82310204 | GERD | MDD | 0.888161 | 0.00201578 | positive | HSD17B2,MIR7854,CMIP,MPHOSPH6,MIR6504,SDR42E1,LOC100129617,PLCG2 |
| 16 | 82310205 | 83077869 | GERD | MDD | 0.555782 | 3.72977E-06 | positive | LOC101928417,MIR8058,CDH13,LOC101928446 |
| 16 | 83077870 | 83778371 | GERD | MDD | 0.498648 | 0.00442189 | positive | CDH13,MIR3182 |
| 16 | 83778372 | 84440154 | GERD | MDD | 0.710679 | 3.56989E-05 | positive | HSBP1,ADAD2,OSGIN1,LOC102724163,SLC38A8,ATP2C2,TAF1C,CDH13,WFDC1,KCNG4,LOC654780,DNAAF1,NECAB2,HSDL1,MBTPS1,MLYCD |
| 16 | 85146806 | 86058597 | GERD | MDD | 0.482901 | 0.0173127 | positive | MIR5093,LINC00311,EMC8,GSE1,MIR1910,LINC02139,GINS2,C16orf74,COX4I1,IRF8,MIR6774,LOC101928557,MIR12128 |
| 16 | 86058598 | 86748867 | GERD | MDD | 0.449059 | 0.000999333 | positive | LINC01081,FOXC2,FOXL1,LINC02135,LINC00917,LINC01082,FOXC2-AS1,FOXF1,FLJ30679,FENDRR,MTHFSD |
| 16 | 88387229 | 89106304 | GERD | MDD | 0.489604 | 0.0119626 | positive | MIR4722,ZC3H18,CBFA2T3,IL17C,SNAI3-AS1,CYBA,LOC339059,LOC100129697,PIEZO1,RNF166,ZC3H18-AS1,LOC100289580,APRT,ZNF469,CDT1,PABPN1L,MIR5189,LOC101927793,MVD,TRAPPC2L,CTU2,SNAI3,ZFPM1,GALNS,LOC100128882 |
| 16 | 89106305 | 90292787 | GERD | MDD | 0.646008 | 0.00205036 | positive | ACSF3,SPATA33,LINC00304,LOC100287036,PRDM7,GAS8,SNORA119,VPS9D1,SPATA2L,VPS9D1-AS1,URAHP,CPNE7,ZNF778,LOC101927817,ANKRD11,LINC02138,MC1R,CDK10,DEF8,CENPBD1,FAM157B,CDH15,GAS8-AS1,CHMP1A,AFG3L1P,TUBB3,SLC22A31,FANCA,SPIRE2,LOC105371414,SNORD68,TCF25,DPEP1,SPG7,ZNF276,DBNDD1,FAM157C,RPL13 |
| 17 | 1480057 | 2699135 | GERD | MDD | 0.801993 | 2.95244E-05 | positive | SERPINF2,MIR22HG,RILP,PAFAH1B1,MIR6776,METTL16,MIR22,SMG6,SRR,MNT,RPA1,RTN4RL1,RAP1GAP2,SCARF1,LOC105371592,DPH1,WDR81,SMYD4,LOC101927839,MIR1253,CCDC92B,TLCD2,HIC1,SLC43A2,SGSM2,SERPINF1,TSR1,LOC105371485,MIR212,OVCA2,CLUH,LOC284009,SNORD91B,PRPF8,MIR132,SNORD91A |
| 17 | 7264459 | 8554763 | GERD | MDD | 0.338791 | 0.0125131 | positive | TNFSF13,SLC35G6,PLSCR3,TNFSF12-TNFSF13,SNORA67,SPEM3,FXR2,LOC100996842,SLC25A35,LOC100128288,NDEL1,NAA38,PER1,TMEM102,ALOX12B,SNORA48,EIF4A1,TP53,TNFSF12,ALOXE3,MIR4521,RPL26,MYH10,ATP1B2,PFAS,MIR4314,CYB5D1,KRBA2,LINC00324,BORCS6,DNAH2,KDM6B,RANGRF,MPDU1,FGF11,TRAPPC1,TMEM256-PLSCR3,POLR2A,CHRNB1,TMEM256,SNORD118,SHBG,HES7,CHD3,NLGN2,VAMP2,TNK1,SOX15,TMEM107,RNF227,ODF4,RPL29P2,CNTROB,MIR6883,EFNB3,TMEM88,CD68,SPEM2,SENP3-EIF4A1,CTC1,SNORD10,ZBTB4,ALOX15B,RNF222,SCARNA21,KCNAB3,AURKB,SPEM1,ARHGEF15,GUCY2D,SAT2,SENP3,WRAP53 |
| 17 | 25872111 | 27344401 | GERD | MDD | 0.8878 | 1.31356E-06 | positive | PHF12,KSR1,TMEM199,FAM222B,SLC13A2,VTN,RSKR,ERAL1,RAB34,MIR144,FOXN1,NEK8,TMEM97,SDF2,LOC101927018,SUPT6H,ALDOC,RPL23A,SPAG5-AS1,MIR451B,PYY2,KRT18P55,POLDIP2,KIAA0100,TRAF4,PIGS,NLK,FLOT2,MIR451A,SNORD4A,SPAG5,NOS2,LYRM9,LINC01992,MIR4723,SLC46A1,SNORD42B,SEZ6,PROCA1,SARM1,UNC119,DHRS13,PPY2P,SNORD4B,MIR4732,LGALS9,SNORD42A,TNFAIP1,IFT20,TLCD1,SEBOX |
| 17 | 33614190 | 34474611 | GERD | MDD | 1 | 0.00044054 | positive | SLFN14,CCL3,CCL14,AP2B1,TAF15,LYZL6,HEATR9,SLFN12L,CCL16,SNORD7,GAS2L2,CCL18,LRRC37A8P,MMP28,C17orf50,CCL15-CCL14,CCL5,RDM1,LOC107985033,SLFN12,SLFN13,RASL10B,SLFN11,CCL15,CCL23,SNHG30,PEX12,CCL4 |
| 17 | 47516225 | 48722590 | GERD | MDD | 0.457317 | 0.00117272 | positive | LOC100288866,DLX3,MIR6165,PICART1,CHAD,MRPL27,EPN3,LINC02075,NGFR,SGCA,PDK2,MYCBPAP,RSAD1,DLX4,SPOP,PPP1R9B,FLJ45513,SAMD14,SLC35B1,NXPH3,EME1,ITGA3,TAC4,LINC01969,CACNA1G-AS1,CACNA1G,FAM117A,TMEM92-AS1,KAT7,ABCC3,LRRC59,COL1A1,H1-9P,TMEM92,SPATA20,ACSF2,XYLT2,LOC105371824 |
| 17 | 49909299 | 51110385 | GERD | MDD | 0.999726 | 1.22562E-06 | positive | LINC02089,LINC01982,CA10,LINC02876 |
| 17 | 60453121 | 62112373 | GERD | MDD | 0.667876 | 0.0139254 | positive | DCAF7,PRR29,ICAM2,PSMC5,ACE,SMARCD2,DDX42,TANC2,CCDC47,SCN4A,LIMD2,KCNH6,CSHL1,TCAM1P,CYB561,METTL2A,MAP3K3,STRADA,LOC105371855,MIR633,MARCHF10,LOC729683,TACO1,FTSJ3,EFCAB3,MRC2,CSH2,CD79B,GH2,MARCHF10-DT,GH1,CSH1,TLK2,PRR29-AS1 |
| 17 | 62112374 | 63548724 | GERD | MDD | 1 | 3.96026E-05 | positive | ARHGAP27P1-BPTFP1-KPNA2P3,AMZ2P1,MIR6080,POLG2,MILR1,PECAM1,LRRC37A3,ARHGAP27P1,SNORA50C,MIR3064,LOC100507002,DDX5,MIR5047,SNHG25,SMURF2,MIR4315-1,AXIN2,GNA13,PLEKHM1P1,ERN1,LINC02563,CEP95,SNORD104,TEX2,MIR4315-2,RGS9 |
| 17 | 64438388 | 65582327 | GERD | MDD | 0.457795 | 0.00259198 | positive | PRKCA,PSMD12,CACNG5,PITPNC1,CACNG4,HELZ,LOC101928021,MIR548AA2,CACNG1,MIR548D2,MIR634 |
| 17 | 72383473 | 73741321 | GERD | MDD | 0.504173 | 0.00484795 | positive | RECQL5,SMIM6,TMEM94,SUMO2,HID1,LLGL2,SAP30BP,CD300LD,SLC9A3R1,CD300A,MIR6785,JPT1,ITGB4,CASKIN2,CDR2L,CD300E,GPRC5C,CD300C,ARMC7,LOC100287042,NT5C,SLC16A5,GGA3,GRB2,NAT9,MIR3615,LOC101928343,TMEM104,SMIM5,LOC100130520,CD300H,MIR3678,MRPL58,CD300LB,MYO15B,ATP5PD,USH1G,OTOP3,TSEN54,CD300LF,FADS6,OTOP2,MIF4GD,RAB37,MRPS7,HID1-AS1,KCTD2,C17orf77,NUP85,GRIN2C,SLC25A19,FDXR |
| 17 | 73741322 | 74908266 | GERD | MDD | 0.693237 | 0.000623091 | positive | QRICH2,MIR6868,H3-3B,MIR4538,ITGB4,MXRA7,EVPL,JMJD6,UNK,RHBDF2,GALK1,CDK3,UBALD2,SRSF2,SRP68,RNF157-AS1,LINC00868,LINC02080,ST6GALNAC1,MIR636,EXOC7,SNORD1C,CYGB,UBE2O,SPHK1,TEN1-CDK3,SNHG16,TRIM65,TEN1,PRPSAP1,METTL23,GALR2,MRPL38,SNORD1A,ZACN,TRIM47,AANAT,FBF1,ST6GALNAC2,WBP2,SNORD1B,UNC13D,RNF157,ACOX1,FOXJ1,PRCD,LOC105274304,MGAT5B,MIR4738,MFSD11 |
| 17 | 74908267 | 75619256 | GERD | MDD | 0.769203 | 2.25509E-05 | positive | SEPTIN9,MIR6516,SCARNA16,LOC105371899,MIR4316,LOC400622,SNHG20,LOC100507351,SEC14L1,MGAT5B,SEPTIN9-DT |
| 17 | 75619257 | 76596287 | GERD | MDD | 0.995864 | 6.91241E-07 | positive | C17orf99,TMC8,AFMID,LINC01987,DNAH17,LOC101928674,TNRC6C,LINC01973,BIRC5,SOCS3,PGS1,LINC01993,TK1,SYNGR2,TMC6,DNAH17-AS1,LOC105371910,TMEM235 |
| 17 | 77412787 | 78438332 | GERD | MDD | 0.674184 | 0.00164401 | positive | TBC1D16,MIR1268B,RNF213,RNF213-AS1,MIR4739,CARD14,LINC01979,CCDC40,CBX4,ENDOV,SLC26A11,LINC02078,RBFOX3,GAA,CBX8,CBX2,LINC01978,MIR4730,EIF4A3,ENPP7,LINC01977,SGSH |
| 17 | 78438333 | 79333774 | GERD | MDD | 0.607153 | 0.00591202 | positive | LOC101928855,NDUFAF8,NPTX1,AATK,BAIAP2-DT,LINC00482,MIR3065,TMEM105,CEP131,PVALEF,RPTOR,BAIAP2,SLC38A10,LOC400627,MIR1250,LOC105371925,MIR338,CHMP6,TEPSIN,MIR657 |
| 17 | 79333775 | 80459149 | GERD | MDD | 0.824683 | 2.76415E-06 | positive | RAC3,MRPL12,FAAP100,BAHCC1,LRRC45,RFNG,HGS,LOC100130370,CCDC57,MCRIP1,TEX19,MIR3186,DUS1L,MIR6786,NOTUM,MAFG,ANAPC11,ARHGDIA,CENPX,SNORD134,PPP1R27,P4HB,MAFG-DT,ACTG1,SIRT7,SECTM1,CD7,HEXD,DCXR,UTS2R,MIR4740,NARF,MYADML2,LINC01970,PCYT2,TSPAN10,OGFOD3,ARL16,NPLOC4,CSNK1D,GCGR,PDE6G,OXLD1,NPB,PYCR1,FSCN2,FASN,DCXR-DT,CCDC137,SLC16A3,ASPSCR1,ALYREF,MIR6787,SLC25A10,CYBC1,GPS1 |
| 18 | 4603962 | 5575812 | GERD | MDD | 0.73256 | 3.59247E-05 | positive | ZBTB14,LINC00667,LINC01892,AKAIN1,EPB41L3,LINC00526 |
| 18 | 6756497 | 7862479 | GERD | MDD | 0.612634 | 0.0012877 | positive | LRRC30,LOC112577592,LOC101927188,PTPRM,LAMA1,ARHGAP28,LINC00668 |
| 18 | 7862480 | 8788524 | GERD | MDD | 0.631159 | 0.0141085 | positive | LOC100192426,GACAT2,PTPRM,RAB12,MTCL1 |
| 18 | 28792735 | 30401128 | GERD | MDD | 0.436387 | 0.00601105 | positive | TTR,B4GALT6,DSG2,KLHL14,DSG3,RNF125,MEP1B,WBP11P1,DSG4,SLC25A52,RNF138,DSG1,DSG2-AS1,TRAPPC8,DSG1-AS1,GAREM1 |
| 18 | 30401129 | 32458907 | GERD | MDD | 0.592271 | 0.0110837 | positive | CCDC178,ASXL3,NOL4,DTNA |
| 18 | 32458908 | 34153297 | GERD | MDD | 0.870917 | 0.000871996 | positive | MIR3975,GALNT1,ZNF397,INO80C,DTNA,ZSCAN30,ZNF271P,MOCOS,ZNF396,MIR187,COSMOC,SLC39A6,C18orf21,FHOD3,MAPRE2,ELP2,RPRD1A,MIR3929,ZNF24 |
| 18 | 34153298 | 36056932 | GERD | MDD | 0.764842 | 1.98594E-05 | positive | SNORA111,LOC105372069,TPGS2,MIR4318,KIAA1328,LOC105372071,FHOD3,LOC105372068,CELF4 |
| 18 | 38498382 | 39410165 | GERD | MDD | 0.738223 | 0.000196592 | positive | KC6 |
| 18 | 39410166 | 40471407 | GERD | MDD | 0.633312 | 0.00297501 | positive | LINC00907,RIT2,PIK3C3 |
| 18 | 40471408 | 41425454 | GERD | MDD | 0.647244 | 0.0150365 | positive | RIT2,SYT4 |
| 18 | 41425455 | 42974165 | GERD | MDD | 0.485862 | 0.00028813 | positive | SLC14A2,MIR4319,LINC01478,SETBP1,LINC01601 |
| 18 | 42974166 | 44300145 | GERD | MDD | 0.714972 | 0.0102636 | positive | ATP5F1A,SLC14A2,EPG5,ST8SIA5,HAUS1,LOXHD1,SLC14A1,PSTPIP2,SIGLEC15,C18orf25,SLC14A2-AS1,RNF165 |
| 18 | 45718493 | 46558306 | GERD | MDD | 0.587708 | 0.00622751 | positive | CTIF,C18orf12,LOC100129878,SMAD7,MIR4743,ZBTB7C |
| 18 | 49790997 | 50372823 | GERD | MDD | 0.77046 | 0.000016262 | positive | DCC |
| 18 | 50372824 | 51060986 | GERD | MDD | 1 | 5.17628E-08 | positive | DCC,MIR4528 |
| 18 | 51060987 | 52512523 | GERD | MDD | 0.846806 | 0.000285314 | positive | LINC01919,MBD2,STARD6,DYNAP,DCC,RAB27B,SNORA37,POLI,LINC01917,C18orf54 |
| 18 | 52512524 | 53762996 | GERD | MDD | 0.432334 | 9.4745E-06 | positive | LINC01539,TCF4,CCDC68,LINC01905,TCF4-AS1,RAB27B,LINC01415,MIR4529,LINC01929,LOC642484,LINC01416 |
| 18 | 55809316 | 56832312 | GERD | MDD | 0.805573 | 0.00109701 | positive | ALPK2,LINC01926,SEC11C,MALT1,OACYLP,ZNF532,LOC101927322,NEDD4L,SNORA108,MIR122,MIR3591 |
| 18 | 57635822 | 58527236 | GERD | MDD | 1 | 0.000182036 | positive | MC4R |
| 18 | 65294196 | 66407960 | GERD | MDD | 0.535544 | 0.00804804 | positive | TMX3,CCDC102B,LOC643542,LINC01903 |
| 18 | 74471746 | 75240760 | GERD | MDD | 0.453855 | 0.0264234 | positive | MBP,ZNF236,ZNF236-DT,GALR1 |
| 18 | 75240761 | 76231685 | GERD | MDD | 0.511025 | 0.0117262 | positive | LINC01029 |
| 18 | 77148138 | 78017072 | GERD | MDD | 0.549817 | 0.00233969 | positive | TXNL4A,PARD6G,HSBP1L1,KCNG2,PARD6G-AS1,LOC284241,ADNP2,CTDP1,LOC284240,NFATC1,RBFADN,RBFA,SLC66A2 |
| 19 | 10028841 | 11681978 | GERD | MDD | 0.521541 | 0.00995938 | positive | TSPAN16,ZGLP1,SMARCA4,MIR199A1,MIR638,LOC105372273,TIMM29,CCDC159,PPAN-P2RY11,LDLR,MIR4748,OLFM2,EIF3G,RDH8,KEAP1,FDX2,ANGPTL8,RAVER1,RAB3D,DOCK6,ICAM1,CDKN2D,KANK2,MIR1181,ICAM5,SNORD105,ZNF653,SPC24,MIR6886,ILF3,ELAVL3,CNN1,ECSIT,MIR6793,KRI1,EPOR,S1PR5,CDC37,DNM2,MIR5589,C3P1,ICAM3,QTRT1,ANGPTL6,C19orf38,YIPF2,TMED1,PLPPR2,P2RY11,PPAN,CARM1,COL5A3,LDLR-AS1,SNORD105B,ICAM4,SWSAP1,DNMT1,MIR1238,PDE4A,MRPL4,AP1M2,S1PR2,TMEM205,ELOF1,MIR4322,ILF3-DT,ATG4D,RGL3,TYK2,MIR7974,CCDC151,SLC44A2,PRKCSH,SHFL |
| 19 | 11681979 | 13213186 | GERD | MDD | 0.552467 | 0.00464953 | positive | PRDX2,MIR6794,DAND5,FARSA,MAN2B1,KLF1,RNASEH2A,ZNF491,FBXW9,SYCE2,NFIX,HOOK2,LYL1,MAST1,CALR,MIR5695,ZNF136,MIR6515,JUNB,ACP5,ZNF563,BEST2,ZNF627,SNORD135,WDR83OS,ZNF564,ZNF439,ZNF442,FARSA-AS1,ZNF799,ZNF44,ZNF709,ZNF791,RTBDN,ZNF490,ZNF823,ZNF433-AS1,ZNF844,ZNF20,DHPS,ZNF763,ZNF443,RAD23A,WDR83,GCDH,ZNF625-ZNF20,MIR5684,LOC100289333,ZNF878,HNRNPA1P10,ZNF440,ZNF625,TRIR,ZNF441,ZNF788P,GET3,SNORD41,DNASE2,GADD45GIP1,ZNF833P,ZNF700,ZNF69,TNPO2,ZNF433,GNG14 |
| 19 | 16079007 | 17045963 | GERD | MDD | 1 | 0.000631264 | positive | EPS15L1,LINC01855,AP1M1,SMIM7,C19orf44,LINC00661,SLC35E1,TPM4,LINC00905,TMEM38A,NWD1,FAM32A,CALR3,SIN3B,CPAMD8,CIB3,F2RL3,KLF2,CHERP,MED26,HSH2D,RAB8A |
| 19 | 17750519 | 18504867 | GERD | MDD | 0.592702 | 4.32378E-05 | positive | RAB3A,IQCN,ARRDC2,CCDC124,LSM4,FCHO1,IL12RB1,JAK3,MAP1S,RPL18A,PIK3R2,IFI30,LOC102725254,UNC13A,LOC729966,MPV17L2,JUND,MIR3188,SLC5A5,SNORA68,PDE4C,GDF15,INSL3,B3GNT3,MAST3,MIR3189,LRRC25,PGPEP1,KCNN1 |
| 19 | 18504868 | 19749166 | GERD | MDD | 0.574334 | 1.97499E-05 | positive | MIR640,CRTC1,TMEM59L,KLHL26,UPF1,ARMC6,HOMER3,KXD1,CERS1,TMEM161A,SUGP1,BORCS8-MEF2B,ELL,DDX49,NR2C2AP,BORCS8,HOMER3-AS1,YJEFN3,SSBP4,CILP2,REX1BD,COPE,COMP,CRLF1,HAPLN4,GATAD2A,PBX4,LPAR2,SLC25A42,ISYNA1,FKBP8,TM6SF2,LRRC25,NDUFA13,RFXANK,MEF2B,GDF1,MAU2,NCAN,GMIP,SUGP2,TSSK6,UBA52 |
| 19 | 19749167 | 20485499 | GERD | MDD | 0.952191 | 1.74193E-05 | positive | ZNF56,ZNF253,ZNF682,ZNF101,ZNF90,ZNF93,ZNF506,LINC00663,ZNF486,ATP13A1,GMIP,ZNF14 |
| 19 | 29389091 | 30375792 | GERD | MDD | 0.806695 | 0.000749455 | positive | VSTM2B,LOC102724958,LINC00906,PLEKHF1,LINC01532,C19orf12,LOC284395,UQCRFS1,CCNE1,POP4 |
| 19 | 31763009 | 33028099 | GERD | MDD | 0.563737 | 0.00134688 | positive | ZNF507,DPY19L3,LINC01782,LOC400684,TSHZ3,LINC01533,THEG5 |
| 19 | 33785836 | 34633274 | GERD | MDD | 0.583207 | 0.00404682 | positive | PEPD,CEBPA-DT,KCTD15,CEBPA,CHST8,CEBPG |
| 19 | 51259179 | 51903804 | GERD | MDD | 0.410741 | 0.0236776 | positive | LIM2,MIR8074,CLDND2,KLK3,SIGLECL1,SNORD88A,KLK8,VSIG10L,KLK10,CD33,SNORD88B,KLK14,LOC105372440,KLK9,KLK12,SNORD88C,SIGLEC9,LOC105372441,IGLON5,KLK7,KLK4,KLK15,KLK13,C19orf48,NKG7,LOC101928517,LINC01869,GPR32,C19orf84,CTU1,KLK1,ACP4,LINC01872,KLK11,KLK6,KLK5,SIGLEC7,KLKP1,ETFB,KLK2,SIGLEC17P |
| 19 | 55714086 | 56411086 | GERD | MDD | 0.44521 | 0.00229054 | positive | NLRP13,RFPL4AL1,MIR6804,SBK3,PTPRH,ZNF579,COX6B2,SNORD157,ZNF524,MIR6802,MIR6803,UBE2S,PPP6R1,NLRP11,TMEM238,ZNF580,NLRP4,KMT5C,NAT14,SSC5D,EPN1,RFPL4A,FAM71E2,MIR6805,ZNF784,ZNF581,IL11,ISOC2,BRSK1,TMEM86B,TMEM150B,ZNF865,U2AF2,RPL28,CCDC106,SHISA7,FIZ1,TMEM190,NLRP9,ZNF628,HSPBP1,SBK2 |
| 19 | 56411087 | 57143509 | GERD | MDD | 0.496575 | 0.0139629 | positive | NLRP13,ZNF444,ZNF582-AS1,ZNF71-SMIM17,LINC01864,ZNF71,ZNF470,NLRP8,ZNF582,ZFP28,ZNF583,ZSCAN5C,ZNF667,ZSCAN5B,ZNF667-AS1,ZSCAN5A,ZNF542P,EDDM13,GALP,ZNF787,ZNF471,NLRP5 |
| 20 | 3736552 | 4591847 | GERD | MDD | 0.794919 | 0.00132918 | positive | SPEF1,MAVS,MIR103B2,CENPB,SMOX,LINC01433,MIR103A2,LINC01730,AP5S1,CDC25B,PANK2,ADRA1D,C20orf27,RNF24 |
| 20 | 5379451 | 6128358 | GERD | MDD | 1 | 1.90889E-07 | positive | LINC00654,CHGB,CRLS1,LINC00658,MCM8-AS1,TRMT6,LOC643406,LRRN4,FERMT1,LINC01729,SHLD1,MCM8,GPCPD1 |
| 20 | 15962577 | 16661516 | GERD | MDD | 0.505008 | 0.00435613 | positive | MACROD2,KIF16B,LOC613266 |
| 20 | 16661517 | 17359290 | GERD | MDD | 0.830637 | 4.67506E-06 | positive | SNRPB2,PCSK2,OTOR |
| 20 | 17359291 | 18058524 | GERD | MDD | 0.625013 | 0.00829044 | positive | BFSP1,SNORD17,RRBP1,DSTN,MGME1,SNX5,OVOL2,BANF2,PCSK2 |
| 20 | 19957929 | 21682377 | GERD | MDD | 0.461634 | 0.0203852 | positive | NAA20,KIZ-AS1,CRNKL1,LINC01727,NKX2-2,LINC01726,CFAP61,KIZ,XRN2,LINC00237,INSM1,RALGAPA2,RIN2,NKX2-4 |
| 20 | 30569660 | 32484506 | GERD | MDD | 0.469418 | 0.0176588 | positive | BPIFA2,CDK5RAP1,KIF3B,E2F1,NOL4L-DT,BPIFB2,BPIFA3,DNMT3B,PXMP4,BPIFA4P,ASXL1,COMMD7,BPIFB1,MIR1825,XKR7,CBFA2T2,SUN5,ZNF341-AS1,ACTL10,MAPRE1,TM9SF4,C20orf203,ZNF341,SNTA1,PLAGL2,CHMP4B,LOC101929698,BPIFB3,EFCAB8,NECAB3,BPIFB6,C20orf144,BPIFB4,POFUT1,BPIFA1,TSPY26P,NOL4L,CCM2L,HCK |
| 20 | 32484507 | 33890060 | GERD | MDD | 0.871715 | 0.00603096 | positive | FAM83C,PROCR,MIR4755,DYNLRB1,NCOA6,ITCH,MYH7B,MIR499B,EIF2S2,MIR644A,MAP1LC3A,RALY,MMP24OS,FAM83C-AS1,GSS,EDEM2,HMGB3P1,MMP24-AS1-EDEM2,AHCY,RALY-AS1,PIGU,EIF6,TRPC4AP,TP53INP2,ACSS2,MIR499A,GGT7,MMP24,ASIP |
| 20 | 43008891 | 44072210 | GERD | MDD | 0.635405 | 0.00265401 | positive | PI3,WFDC12,KCNK15,TOMM34,KCNK15-AS1,SDC4,YWHAB,PKIG,MATN4,SEMG2,RBPJL,TP53TG5,SLPI,RIMS4,LINC01260,TTPAL,PIGT,SYS1,MIR3646,LINC01620,KCNS1,STK4,MIR6812,WFDC5,LOC105372631,HNF4A,ADA,SERINC3,DBNDD2,PABPC1L,CCN5,SYS1-DBNDD2,STK4-AS1,SEMG1,HNF4A-AS1,LINC01430 |
| 20 | 44072211 | 45673603 | GERD | MDD | 0.50573 | 0.000336413 | positive | TP53RK,ZNF335,WFDC8,SLC12A5,ZSWIM1,WFDC6,WFDC13,SPATA25,ZNF663P,EPPIN-WFDC6,WFDC2,PCIF1,MKRN7P,MMP9,UBE2C,CD40,CTSA,ACOT8,TNNC2,SLC13A3,NCOA5,ZNF334,WFDC11,CDH22,OCSTAMP,LOC105372631,SLC12A5-AS1,WFDC9,MIR3617,EPPIN,WFDC10B,SPINT4,WFDC3,SPINT3,SLC2A10,DNTTIP1,ZSWIM3,PLTP,WFDC10A,ELMO2,SLC35C2,SNX21,EYA2,NEURL2,LOC105372633 |
| 20 | 49236419 | 50653620 | GERD | MDD | 0.875725 | 4.98753E-07 | positive | RIPOR3-AS1,ATP9A,SALL4,DPM1,BCAS4,KCNG1,MIR3194,ADNP,NFATC2,LINC01429,RIPOR3,MOCS3,ADNP-AS1,PARD6B |
| 20 | 51533751 | 52411532 | GERD | MDD | 0.6255 | 0.000269336 | positive | LOC105372672,ZNF217,TSHZ2,LOC101927770 |
| 20 | 53879804 | 54743694 | GERD | MDD | 0.646079 | 0.0170329 | positive | LINC01441,CBLN4,LINC01440 |
| 20 | 55448076 | 56172843 | GERD | MDD | 0.662639 | 0.000687691 | positive | RAE1,PCK1,CTCFL,BMP7,RBM38-AS1,MIR4325,SPO11,RBM38,BMP7-AS1,MTRNR2L3 |
| 20 | 56172844 | 56951773 | GERD | MDD | 0.661425 | 0.00875757 | positive | ANKRD60,LINC01742,RAB22A,ZBP1,PPP4R1L,NKILA,C20orf85,PMEPA1 |
| 20 | 57924038 | 58780894 | GERD | MDD | 0.689527 | 0.0008373 | positive | CDH26,SYCP2,PPP1R3D,PHACTR3-AS1,C20orf197,LOC729296,MIR646HG,PHACTR3,FAM217B |
| 20 | 58780895 | 59560457 | GERD | MDD | 0.513293 | 0.00147185 | positive | LOC101928048,MIR4533,MIR646HG,MIR548AG2,MIR646 |
| 20 | 61470769 | 62179241 | GERD | MDD | 0.370347 | 0.0222646 | positive | PPDPF,SLC17A9,LINC01749,MIR124-3,LINC00029,LOC100130587,HAR1B,SRMS,BIRC7,PTK6,MIR4326,SNORA117,YTHDF1,HAR1A,GID8,DIDO1,ARFGAP1,KCNQ2,NKAIN4,DPH3P1,EEF1A2,FLJ16779,COL9A3,COL20A1,TCFL5,LINC01056,MIR3196,BHLHE23,CHRNA4 |
| 20 | 62179242 | 62963627 | GERD | MDD | 0.721906 | 1.63507E-06 | positive | RTEL1-TNFRSF6B,SAMD10,ABHD16B,ZGPAT,PRPF6,MIR6813,TCEA2,STMN3,SOX18,MIR941-5,LKAAEAR1,ZBTB46,GMEB2,SLC2A4RG,LOC112268269,TNFRSF6B,C20orf204,RGS19,MHENCR,LINC00266-1,FNDC11,TPD52L2,UCKL1-AS1,MIR647,ARFRP1,DNAJC5,RTEL1,HELZ2,ZBTB46-AS1,MIR1914,MIR941-1,MIR941-3,LIME1,PCMTD2,UCKL1,MYT1,NPBWR2,ZNF512B,MIR941-4,OPRL1,MIR941-2 |
| 21 | 16873444 | 18222037 | GERD | MDD | 0.852431 | 0.000211546 | positive | USP25,SNORD74B,MIR99AHG,MIR99A,MIR125B2,MIRLET7C |
| 21 | 18222038 | 19479024 | GERD | MDD | 0.430711 | 0.019112 | positive | BTG3-AS1,LINC01549,BTG3,C21orf91,CHODL-AS1,CHODL,CXADR,C21orf91-OT1 |
| 21 | 19479025 | 20407307 | GERD | MDD | 0.691156 | 0.00122341 | positive | MIR548X,CHODL,TMPRSS15,MIR548XHG |
| 21 | 26467827 | 27774350 | GERD | MDD | 0.837111 | 0.00031478 | positive | APP,JAM2,CYYR1-AS1,MIR155HG,ATP5PF,GABPA,MRPL39,MIR155,LINC00515,LINC00158 |
| 21 | 28616540 | 29449870 | GERD | MDD | 1 | 0.000174679 | positive | LINC00314,LINC01697,LINC00113 |
| 21 | 34383795 | 35729261 | GERD | MDD | 0.662193 | 0.00345737 | positive | IL10RB,MIR6501,LOC101928126,TMEM50B,ITSN1,DNAJC28,SLC5A3,LOC101928107,OLIG2,ATP5PO,IFNAR1,LINC01548,IL10RB-DT,CRYZL1,OLIG1,MRPS6,GART,LINC00649,LINC00945,IFNGR2,DONSON,SON,LINC00310,IFNAR2 |
| 21 | 35729262 | 37303548 | GERD | MDD | 0.471972 | 0.000656708 | positive | LINC01426,PPP1R2P2,RCAN1,RUNX1,SMIM34A,SMIM11B,FAM243B,KCNE2,MIR802,LINC00160,KCNE1,FAM243A,RUNX1-IT1,CLIC6,LOC100506403,KCNE1B,SMIM11A |
| 21 | 38046450 | 39252059 | GERD | MDD | 0.563963 | 0.00987066 | positive | KCNJ6,RIPPLY3,HLCS,DYRK1A,DSCR9,TTC3-AS1,TTC3,PIGP,VPS26C,SIM2 |
| 21 | 39252060 | 40480831 | GERD | MDD | 0.342599 | 0.0259934 | positive | KCNJ6,LINC01700,ERG,LINC01423,LOC101928398,KCNJ15,ETS2,DSCR10,DSCR4,LOC400867,DSCR8,LINC00114 |
| 21 | 43081435 | 43812791 | GERD | MDD | 0.634759 | 0.000104017 | positive | TFF1,UMODL1-AS1,LINC00111,PRDM15,TFF3,SNORA91,UMODL1,MIR6814,TMPRSS3,LINC00479,LINC00112,ZNF295-AS1,ZBTB21,ABCG1,RIPK4,C2CD2,TFF2 |
| 21 | 43812792 | 44815008 | GERD | MDD | 0.895704 | 0.00214048 | positive | LOC106780825,SLC37A1,CBS,LINC01679,MIR5692B,UBASH3A,PKNOX1,LOC101930094,FRGCA,NDUFV3,U2AF1L5,LINC01671,U2AF1,LINC01668,RSPH1,CRYAA,ERVH48-1,LINC00322,CRYAA2,TMPRSS3,WDR4,PDE9A,CBSL |
| 21 | 44815009 | 46045433 | GERD | MDD | 0.533773 | 0.017681 | positive | PWP2,MIR6070,KRTAP10-8,CSTB,LRRC3-DT,KRTAP10-6,AATBC,AGPAT3,AIRE,GATD3A,PFKL,TSPEAR-AS1,KRTAP10-4,CFAP410,RRP1,KRTAP10-5,H2BS1,LINC02575,TRAPPC10,LINC00313,GATD3B,TSPEAR-AS2,KRTAP10-7,SIK1B,TRPM2-AS,LRRC3,SIK1,LINC01678,LOC102724159,RRP1B,HSF2BP,TSPEAR,DNMT3L-AS1,ICOSLG,LINC01669,KRTAP10-3,KRTAP10-2,LOC102723996,KRTAP10-1,TRPM2,PDXK,DNMT3L,LINC00319 |
| 21 | 46045434 | 47014880 | GERD | MDD | 0.5895 | 0.00604314 | positive | LINC01424,KRTAP10-9,COL18A1-AS2,LINC01694,PTTG1IP,FAM207A,ITGB2-AS1,SSR4P1,KRTAP10-10,KRTAP12-4,LINC01547,KRTAP10-12,COL18A1-AS1,LINC00334,LINC00316,UBE2G2,KRTAP12-3,COL18A1,POFUT2,LINC00163,ADARB1,PICSAR,ITGB2,KRTAP10-11,LINC00205,KRTAP12-1,SLC19A1,TSPEAR,MIR6815,SUMO3,KRTAP12-2 |
| 22 | 17587475 | 18535225 | GERD | MDD | 0.530345 | 0.0181473 | positive | MIR3198-1,ADA2,LINC01634,CECR2,LINC00528,TMEM121B,IL17RA,BID,LINC01664,BCL2L13,SLC25A18,ATP6V1E1,HDHD5,MICAL3,MIR648,LOC101929372,HDHD5-AS1,CECR3 |
| 22 | 19635655 | 20969184 | GERD | MDD | 0.53611 | 0.00334997 | positive | TRMT2A,ZNF74,GP1BB,SEPT5-GP1BB,PI4KAP1,CCDC188,LOC107987394,FAM230J,LOC107987389,MIR1306,GNB1L,DGCR6L,TANGO2,SEPTIN5,KLHL22,SCARF2,ARVCF,ZDHHC8,COMT,GGTLC3,LOC284865,LINC00896,DGCR8,FAM230G,FAM230A,TBX1,RTN4R,MIR3618,MIR4761,RIMBP3,TMEM191B,RTL10,TXNRD2,MIR6816,MIR1286,MED15,RANBP1,SNORA77B,MIR185 |
| 22 | 25282437 | 26166934 | GERD | MDD | 0.607626 | 0.0138522 | positive | SGSM1,KIAA1671,CRYBB2P1,GRK3,LRP5L,TMEM211,IGLL3P,MIR6817,CRYBB2,KIAA1671-AS1,MYO18B,CRYBB3 |
| 22 | 38718590 | 40378783 | GERD | MDD | 0.533019 | 0.0113007 | positive | TPTEP2-CSNK1E,SNORD43,LOC100506472,RPL3,APOBEC3A_B,SUN2,APOBEC3A,FAM227A,APOBEC3C,APOBEC3B,MGAT3,DMC1,SYNGR1,JOSD1,DDX17,CBX7,TAB1,TPTEP2,CACNA1I,GTPBP1,NPTXR,TOMM22,APOBEC3D,SNORD83A,PDGFB,APOBEC3B-AS1,ATF4,APOBEC3G,APOBEC3H,RPS19BP1,LOC105373031,KDELR3,APOBEC3F,DNAL4,ENTHD1,GRAP2,KCNJ4,CBY1,CBX6,SNORD83B,SNORD139,MGAT3-AS1,MIEF1 |
| 22 | 40378784 | 41722787 | GERD | MDD | 0.911316 | 2.56667E-07 | positive | RANGAP1,MRTFA,EP300,L3MBTL2-AS1,ST13,SLC25A17,SGSM3,MRTFA-AS1,RBX1,TNRC6B,LOC114841040,XPNPEP3,SNORD140,MIR1281,MCHR1,MIR6889,CHADL,L3MBTL2,DNAJB7,MIR4766,EP300-AS1,FAM83F,ZC3H7B,ADSL |
| 22 | 41722788 | 42867927 | GERD | MDD | 0.628972 | 0.000835578 | positive | TOB2,DESI1,LINC00634,SNU13,MIR378I,NFAM1,CCDC134,CYP2D7,LINC01315,MIR33A,NAGA,LOC101929829,PHETA2,CYP2D6,WBP2NL,C22orf46,SMDT1,CSDC2,OGFRP1,SREBF2-AS1,PHF5A,PMM1,XRCC6,SHISA8,SREBF2,NDUFA6,TCF20,SEPTIN3,ACO2,NDUFA6-DT,LOC105373044,TNFRSF13C,ZC3H7B,MEI1,POLR3H,TEF,CENPM |
| 22 | 44409534 | 45170649 | GERD | MDD | 0.320763 | 0.0255297 | positive | PARVB,PRR5,RTL6,LINC01656,SHISAL1,PRR5-ARHGAP8,PARVG,LINC00229,ARHGAP8,LINC00207 |
| 22 | 49559208 | 50347996 | GERD | MDD | 0.508867 | 0.000838231 | positive | MIR3667,CRELD2,ALG12,C22orf34,BRD1,ZBED4 |
| 22 | 50347997 | 51244236 | GERD | MDD | 0.759037 | 0.000376697 | positive | TRABD,MIR12114,MAPK12,CHKB,SCO2,SBF1,MOV10L1,PIM3,TTLL8,ODF3B,CPT1B,ADM2,MLC1,LMF2,MIR6821,LOC105373100,DENND6B,MAPK11,NCAPH2,MIOX,RABL2B,TYMP,CHKB-CPT1B,PLXNB2,SHANK3,PANX2,ACR,CHKB-DT,IL17REL,SYCE3,ARSA,SELENOO,KLHDC7B,PPP6R2,HDAC10,TUBGCP6,RPL23AP82,MAPK8IP2 |
| 1 | 7711795 | 8580987 | GERD | SCZ | -0.745177 | 0.000668213 | negative | RERE-AS1,ERRFI1,SLC45A1,VAMP3,SNORD128,RERE,LINC01714,UTS2,PER3,TNFRSF9,CAMTA1,PARK7 |
| 1 | 41093876 | 41977966 | GERD | SCZ | 0.620487 | 0.00186805 | positive | NFYC,SLFNL1,RIMS3,CTPS1,FOXO6,KCNQ4,MIR30C1,MIR30E,NFYC-AS1,EDN2,HIVEP3,SLFNL1-AS1,FOXO6-AS1,CITED4,SCMH1 |
| 1 | 73992171 | 75132092 | GERD | SCZ | 0.594963 | 0.00088348 | positive | FPGT,ERICH3-AS1,FPGT-TNNI3K,LINC02238,ERICH3,LRRC53,TNNI3K,LRRIQ3 |
| 1 | 168179087 | 169521551 | GERD | SCZ | -0.794323 | 0.00185087 | negative | SFT2D2,LINC00970,CCDC181,TBX19,LINC00626,DPT,XCL1,LOC100505918,MIR557,NME7,F5,BLZF1,ANKRD36BP1,LOC101928565,XCL2,SLC19A2,LOC101928596,ATP1B1 |
| 1 | 243048004 | 244723196 | GERD | SCZ | -0.453096 | 0.000516649 | negative | SDCCAG8,LINC01347,CATSPERE,C1orf100,LINC02774,CEP170,ADSS2,ZBTB18,AKT3,MIR4677 |
| 2 | 199654970 | 201324132 | GERD | SCZ | 0.405323 | 0.000913218 | positive | C2orf69,SPATS2L,LINC01877,SATB2-AS1,MAIP1,FTCDNL1,SATB2,TYW5 |
| 2 | 229764919 | 230541092 | GERD | SCZ | -0.892964 | 0.000063224 | negative | PID1,DNER |
| 3 | 174684708 | 175671424 | GERD | SCZ | 0.9653 | 0.00181717 | positive | NAALADL2-AS1,NAALADL2-AS2,MIR4789,NAALADL2,NAALADL2-AS3 |
| 4 | 17290237 | 18753263 | GERD | SCZ | 0.543403 | 0.00107843 | positive | FAM184B,CLRN2,QDPR,DCAF16,SNORA75B,NCAPG,LAP3,MED28,LCORL |
| 5 | 91956906 | 93814604 | GERD | SCZ | 0.526815 | 0.00182197 | positive | NR2F1-AS1,POU5F2,KIAA0825,FAM172A,MIR2277,NR2F1 |
| 5 | 124109486 | 125131720 | GERD | SCZ | 0.624563 | 6.96298E-06 | positive | LOC101927421,LINC02240 |
| 5 | 132554691 | 134556570 | GERD | SCZ | 0.503386 | 0.000230745 | positive | MIR3661,UBE2B,LINC01843,MIR1289-2,PITX1,SEC24A,WSPAR,DDX46,CAMLG,SKP1,JADE2,LOC102546229,C5orf66-AS1,LOC105379183,CDKL3,CATSPER3,TCF7,VDAC1,CDKN2AIPNL,C5orf24,C5orf66,SAR1B,FSTL4,PPP2CA,C5orf15,PCBD2,TXNDC15 |
| 6 | 107309328 | 109043244 | GERD | SCZ | 0.559668 | 0.000332916 | positive | SCML4,SNX3,FOXO3,SEC63,OSTM1,NR2E1,SOBP,OSTM1-AS1,BEND3,MTRES1,AFG1L,PDSS2 |
| 7 | 81533567 | 82759283 | GERD | SCZ | 0.752313 | 7.01638E-06 | positive | CACNA2D1-AS1,CACNA2D1,PCLO |
| 8 | 9835864 | 10478851 | GERD | SCZ | -0.690207 | 0.000413376 | negative | LINCR-0001,RP1L1,PRSS55,MSRA |
| 8 | 58675118 | 59555977 | GERD | SCZ | 0.693076 | 0.00185675 | positive | NSMAF,LINC01602,SDCBP,LOC101929528,UBXN2B,FAM110B,CYP7A1 |
| 9 | 20483881 | 21674032 | GERD | SCZ | 0.855316 | 0.000256848 | positive | IFNA22P,IFNA8,MIR31,IFNA17,IFNA1,IFNA16,IFNA10,IFNA5,MIR491,IFNA14,IFNA6,SNORA30B,MIR4474,IFNW1,IFNE,MLLT3,FOCAD,MIR31HG,IFNA4,IFNA7,HACD4,IFNB1,IFNA13,IFNA21,IFNA2,KLHL9,FOCAD-AS1 |
| 9 | 121008530 | 122677719 | GERD | SCZ | 0.430871 | 0.00124982 | positive | BRINP1,LINC02578 |
| 10 | 72659224 | 73514214 | GERD | SCZ | -1 | 1.24268E-07 | negative | SLC29A3,UNC5B-AS1,LINC02622,C10orf105,CDH23-AS1,VSIR,UNC5B,CDH23 |
| 10 | 129134739 | 129831969 | GERD | SCZ | 0.538301 | 0.000744421 | positive | NPS,CLRN3,DOCK1,PTPRE,FOXI2 |
| 11 | 1061400 | 1857845 | GERD | SCZ | -0.813005 | 0.00115682 | negative | MUC2,MOB2,KRTAP5-3,KRTAP5-6,KRTAP5-5,TOLLIP,MUC5AC,SYT8,BRSK2,MUC5B,KRTAP5-4,MIR6744,KRTAP5-2,DUSP8,CTSD,FAM99A,MUC5B-AS1,KRTAP5-1,TOLLIP-AS1,IFITM10,FAM99B,KRTAP5-AS1 |
| 11 | 112755447 | 113889019 | GERD | SCZ | 0.577301 | 6.13678E-06 | positive | HTR3B,TTC12,NCAM1,NCAM1-AS1,TMPRSS5,HTR3A,ZW10,USP28,DRD2,LOC101928847,MIR4301,CLDN25,ANKK1 |
| 13 | 37499811 | 38290689 | GERD | SCZ | 0.904702 | 0.000166876 | positive | POSTN,LINC01048,ALG5,CSNK1A1L,LINC00547,SUPT20H,EXOSC8,TRPC4 |
| 13 | 53336572 | 54684856 | GERD | SCZ | -0.686827 | 0.000899523 | negative | LINC01065,MIR759,OLFM4,PCDH8,LINC00558 |
| 13 | 79489046 | 80778339 | GERD | SCZ | -0.78175 | 0.000713703 | negative | LINC01068,NDFIP2,LINC01080,NDFIP2-AS1,RBM26,LINC00382,LINC01038,RBM26-AS1 |
| 13 | 101573737 | 103050417 | GERD | SCZ | -0.577677 | 0.00131161 | negative | FGF14-IT1,MIR4705,FGF14-AS1,FGF14,NALCN-AS1,LINC00411,NALCN,MIR2681,ITGBL1,FGF14-AS2 |
| 15 | 82191078 | 84209760 | GERD | SCZ | -0.650991 | 0.000863368 | negative | LINC01583,RPS17,ACTG1P17,FSD2,C15orf40,SCARNA15,WHAMM,GOLGA6L10,HDGFL3,SNHG21,TM6SF1,HOMER2,BTBD1,EFL1,MEX3B,UBE2Q2P2,CPEB1,GOLGA6L9,AP3B2,ADAMTS7P1,LOC727751,LOC338963,SH3GL3,SAXO2,BNC1,MIR4515,RAMAC,GOLGA2P10,CPEB1-AS1,LOC102724034,GOLGA6L17P |
| 15 | 95662426 | 96864278 | GERD | SCZ | 0.488284 | 0.00165679 | positive | LINC01197,LINC00924,NR2F2-AS1,LOC105369212 |
| 16 | 5782969 | 6446081 | GERD | SCZ | 0.534963 | 0.000260208 | positive | RBFOX1 |
| 16 | 26154123 | 27443061 | GERD | SCZ | 0.553219 | 0.000528305 | positive | NSMCE1,KDM8,IL4R,C16orf82,NSMCE1-DT,IL21R |
| 16 | 72089512 | 73140781 | GERD | SCZ | 0.725858 | 9.81911E-06 | positive | HPR,TXNL4B,PMFBP1,ZFHX3,HP,HCCAT5,LINC01572,DHX38 |
| 22 | 41722788 | 42867927 | GERD | SCZ | 0.570766 | 0.00100249 | positive | TOB2,DESI1,LINC00634,SNU13,MIR378I,NFAM1,CCDC134,CYP2D7,LINC01315,MIR33A,NAGA,LOC101929829,PHETA2,CYP2D6,WBP2NL,C22orf46,SMDT1,CSDC2,OGFRP1,SREBF2-AS1,PHF5A,PMM1,XRCC6,SHISA8,SREBF2,NDUFA6,TCF20,SEPTIN3,ACO2,NDUFA6-DT,LOC105373044,TNFRSF13C,ZC3H7B,MEI1,POLR3H,TEF,CENPM |
| 1 | 38474037 | 40200950 | GERD | PTSD | 0.478675 | 0.0026107 | positive | HPCAL4,MACF1,HEYL,PABPC4,AKIRIN1,SNORA55,RHBDL2,UTP11,NT5C1A,PPIEL,GJA9,PABPC4-AS1,MYCBP,RRAGC,NDUFS5,POU3F1,OXCT2P1,GJA9-MYCBP,LINC01343,LOC105378663,BMP8A,MIR3659,MIR3659HG,LINC01685,KIAA0754 |
| 1 | 41977967 | 43512669 | GERD | PTSD | 0.681623 | 0.00480446 | positive | C1orf50,ERMAP,SVBP,GUCA2B,GUCA2A,ZNF691,P3H1,YBX1,TMEM269-DT,LOC339539,SLC2A1,PPIH,FOXJ3,CCDC30,CLDN19,PPCS,TMEM269,SLC2A1-AS1,HIVEP3,ZMYND12,RIMKLA |
| 1 | 53114187 | 54771293 | GERD | PTSD | 0.988218 | 1.45049E-05 | positive | TMEM59,CPT2,NDC1,LRRC42,PODN,SLC1A7,CYB5RL,ECHDC2,CZIB,LDLRAD1,DMRTB1,TCEANC2,LINC01771,CDCP2,MIR4781,ZYG11A,SSBP3,MAGOH,SLC25A3P1,SSBP3-AS1,YIPF1,LRP8,LOC100507564,COA7,SHISAL2A,MRPL37,SCP2,DIO1,HSPB11,GLIS1,ZYG11B |
| 1 | 54771294 | 55727134 | GERD | PTSD | 0.543882 | 0.00884289 | positive | TMEM61,LOC100507634,MROH7-TTC4,FAM151A,LEXM,ACOT11,SSBP3,PCSK9,MROH7,TTC4,MIR4422,BSND,USP24,DHCR24,PARS2,MIR4422HG,TTC22 |
| 1 | 62778734 | 64344226 | GERD | PTSD | 0.737182 | 0.000126669 | positive | FOXD3-AS1,EFCAB7,PGM1,ALG6,DLEU2L,LINC01739,DOCK7,KANK4,MIR6068,USP1,ITGB3BP,ROR1,ANGPTL3,ATG4C,FOXD3,LINC00466 |
| 1 | 72513120 | 73992170 | GERD | PTSD | 0.794871 | 0.000488785 | positive | NEGR1,LINC01360 |
| 1 | 73992171 | 75132092 | GERD | PTSD | 0.834943 | 0.000197248 | positive | FPGT,ERICH3-AS1,FPGT-TNNI3K,LINC02238,ERICH3,LRRC53,TNNI3K,LRRIQ3 |
| 1 | 87667573 | 88656689 | GERD | PTSD | 0.565936 | 0.00214437 | positive | LMO4,LINC02801,LINC01364 |
| 1 | 111134063 | 112489248 | GERD | PTSD | 0.513596 | 0.00741549 | positive | INKA2-AS1,WDR77,LOC101928718,LRIF1,KCND3,CHIAP2,KCND3-AS1,C1orf162,PIFO,DRAM2,DENND2D,DDX20,KCNA2,TMIGD3,KCND3-IT1,RAP1A,CD53,OVGP1,ADORA3,LINC01160,PGBP,ATP5PB,CHI3L2,CEPT1,CHIA,KCNA3,LOC107985184,INKA2 |
| 1 | 114664388 | 116207297 | GERD | PTSD | 0.611069 | 0.0109531 | positive | TSHB,VANGL1,DENND2C,TRIM33,NGF,AMPD1,SIKE1,SYT6,CSDE1,SYCP1,NGF-AS1,NRAS,TSPAN2,BCAS2 |
| 1 | 168179087 | 169521551 | GERD | PTSD | 0.729583 | 0.000395042 | positive | SFT2D2,LINC00970,CCDC181,TBX19,LINC00626,DPT,XCL1,LOC100505918,MIR557,NME7,F5,BLZF1,ANKRD36BP1,LOC101928565,XCL2,SLC19A2,LOC101928596,ATP1B1 |
| 1 | 204092538 | 205009623 | GERD | PTSD | 0.595198 | 0.00413963 | positive | PIK3C2B,KISS1,MDM4,LINC00628,NFASC,REN,ETNK2,PLEKHA6,PPP1R15B,SOX13,LRRN2,ERLNC1,GOLT1A |
| 1 | 213958293 | 214783872 | GERD | PTSD | 0.513003 | 0.00125617 | positive | PTPN14,PROX1-AS1,PROX1,LINC00538,CENPF,SMYD2 |
| 1 | 241033585 | 241813534 | GERD | PTSD | 0.840603 | 0.000146325 | positive | MIR3123,OPN3,FH,KMO,CHML,RGS7 |
| 2 | 15721704 | 17241115 | GERD | PTSD | 0.58625 | 0.00662854 | positive | MYCNUT,GACAT3,CYRIA,MYCN,MYCNOS,DDX1,LINC01804 |
| 2 | 21084899 | 22429640 | GERD | PTSD | 0.862951 | 0.00115122 | positive | APOB,TDRD15,LINC01822 |
| 2 | 22429641 | 23538527 | GERD | PTSD | 0.79084 | 3.62206E-05 | positive | LINC01884 |
| 2 | 23538528 | 24669224 | GERD | PTSD | 1 | 0.000334884 | positive | MFSD2B,ATAD2B,KLHL29,FAM228A,UBXN2A,SF3B6,FAM228B,PFN4,TP53I3,FKBP1B,ITSN2,WDCP |
| 2 | 49114133 | 49849185 | GERD | PTSD | 0.756129 | 0.0126701 | positive | FSHR,MIR548BA |
| 2 | 59251997 | 60775066 | GERD | PTSD | 0.58984 | 1.5863E-08 | positive | MIR4432HG,BCL11A,LINC01122,LINC01793,MIR4432 |
| 2 | 100202437 | 101556799 | GERD | PTSD | 0.686309 | 8.95356E-05 | positive | AFF3,LINC01104,LINC01849,LONRF2,NMS,CHST10,PDCL3,NPAS2 |
| 2 | 103305258 | 105120578 | GERD | PTSD | 0.737091 | 0.000498512 | positive | LINC01831,LOC100287010,SLC9A2,LINC01102,LINC01796,LINC01935,TMEM182,LINC01103,MFSD9 |
| 2 | 108417774 | 110097868 | GERD | PTSD | 0.859444 | 0.000035924 | positive | LINC01594,LINC01593,SH3RF3,MIR4266,LIMS1,LIMS1-AS1,SLC5A7,EDAR,SULT1C2P1,CCDC138,RANBP2,RGPD4-AS1,RGPD4,GACAT1,SULT1C3,SULT1C2,GCC2,MIR4265,SULT1C4,SH3RF3-AS1,GCC2-AS1 |
| 2 | 124730992 | 125741528 | GERD | PTSD | 0.866627 | 8.41343E-05 | positive | CNTNAP5,LOC107985820 |
| 2 | 138073792 | 139505819 | GERD | PTSD | 0.569397 | 0.0100372 | positive | LOC101928273,LINC01832,NXPH2,HNMT,SPOPL,THSD7B |
| 2 | 142494981 | 143259780 | GERD | PTSD | 0.518986 | 0.0098685 | positive | LRP1B |
| 2 | 144503689 | 145977920 | GERD | PTSD | 0.657908 | 0.000250829 | positive | LINC01412,GTDC1,TEX41,ZEB2-AS1,ZEB2,LOC101928386,LOC105373656,ARHGAP15 |
| 2 | 161432090 | 163607635 | GERD | PTSD | 0.76668 | 0.00148257 | positive | LOC101929512,LOC101929532,KCNH7,TANK,FAP,DPP4-DT,IFIH1,GCG,LINC01806,AHCTF1P1,PSMD14,SLC4A10,TBR1,DPP4,GCA |
| 2 | 174927564 | 176734750 | GERD | PTSD | 0.714282 | 8.91465E-05 | positive | GPR155,LINC01305,RNU6-1,H3P6,CIR1,OLA1,RNU6-7,RNU6-2,CHN1,RNU6-9,ATF2,SCRN3,RNU6-8,SP9,ATP5MC3,WIPF1,MIR933,CHRNA1 |
| 3 | 21790868 | 22648001 | GERD | PTSD | 0.892678 | 0.00170917 | positive | ZNF385D,ZNF385D-AS2 |
| 3 | 23426045 | 24270189 | GERD | PTSD | 0.672104 | 0.0095045 | positive | RPL15,UBE2E1-AS1,THRB-AS2,UBE2E2,LINC00691,THRB,NKIRAS1,UBE2E1,NR1D2 |
| 3 | 47588462 | 50387742 | GERD | PTSD | 0.862328 | 0.000069258 | positive | APEH,GNAI2,MAP4,SLC26A6,SLC25A20,MIR4443,LSMEM2,C3orf84,NPRL2,NCKIPSD,SEMA3B-AS1,SNORA94,MIR191,ZNF589,MIR5787,MIR6824,GPX1,TREX1,DALRD3,PLXNB1,CCDC51,NDUFAF3,LINC02585,RBM6,C3orf62,TMEM89,MIR5193,HYAL1,FCF1P2,CDC25A,RHOA,UCN2,ARIH2OS,RBM5,MIR4271,HYAL3,CAMP,SEMA3B,QARS1,CELSR3,QRICH1,RASSF1-AS1,NICN1,IFRD2,IMPDH2,CDHR4,NAA80,USP4,ATRIP,SHISA5,NME6,ARIH2,DHX30,SEMA3F,GNAT1,TMA7,KLHDC8B,MIR2115,MIR6890,SPINK8,IP6K2,COL7A1,RNF123,RBM5-AS1,SMARCC1,MIR4793,CCDC71,GMPPB,IP6K1,WDR6,RASSF1,UQCRC1,IHO1,UBA7,BSN-DT,TUSC2,INKA1,LAMB2,ATRIP-TREX1,PRKAR2A,MIR711,FBXW12,MON1A,TCTA,CSPG5,MIR1226,MIR425,MIR6872,MIR6823,MST1,P4HTM,BSN,SLC38A3,HYAL2,DAG1,CAMKV,AMT,ZMYND10,PRKAR2A-AS1,TRAIP,USP19,SNORD146,LAMB2P1,MST1R,PFKFB4,SEMA3F-AS1,AMIGO3 |
| 3 | 50387743 | 51953968 | GERD | PTSD | 0.779558 | 0.00263957 | positive | CYB561D2,IQCF3,MIR4787,HEMK1,IQCF6,GRM2,TEX264,IQCF5,RAD54L2,NPRL2,IQCF4P,DOCK3,RBM15B,DCAF1,CISH,CACNA2D2,IQCF1,IQCF2,MAPKAPK3,C3orf18,MANF,TMEM115,IQCF5-AS1 |
| 3 | 51953969 | 54074844 | GERD | PTSD | 0.655776 | 0.00427372 | positive | STAB1,PHF7,NISCH,MIRLET7G,MIR8064,TNNC1,DNAH1,DUSP7,PARP3,LOC101929054,SPCS1,SEMA3G,POC1A,MUSTN1,SNORD136,NT5DC2,BAP1,PBRM1,CACNA1D,GLT8D1,RPL29,ABHD14B,SNORD19C,GLYCTK,CHDH,PCBP4,ABHD14A-ACY1,ACTR8,ITIH4,ABHD14A,RRP9,STIMATE,WDR82,ALAS1,SNORD38C,PRKCD,ITIH1,RFT1,NEK4,ITIH3,GLYCTK-AS1,DCP1A,SFMBT1,TKT,IL17RB,ACY1,SNORD19,GNL3,SNORD69,ITIH4-AS1,TLR9,STIMATE-MUSTN1,SELENOK,SMIM4,MIR135A1,GPR62,TWF2,LINC00696,PPM1M,SNORD19B |
| 3 | 65326752 | 66702084 | GERD | PTSD | 0.831943 | 4.25418E-06 | positive | MAGI1-AS1,MAGI1,MAGI1-IT1,SLC25A26,LRIG1 |
| 3 | 71223282 | 72334704 | GERD | PTSD | 0.393848 | 0.0100422 | positive | LINC00877,FOXP1,MIR1284,LINC00870,EIF4E3,GPR27,PROK2,FOXP1-AS1 |
| 3 | 135288395 | 137372141 | GERD | PTSD | 0.647915 | 0.00404467 | positive | NCK1-DT,STAG1,PCCB,NCK1,SLC35G2,IL20RB,MSL2,PPP2R3A |
| 3 | 172004761 | 173415591 | GERD | PTSD | 0.544371 | 0.000555332 | positive | FNDC3B,LINC02068,ECT2,SPATA16,TNFSF10,GHSR,NLGN1,NCEH1 |
| 3 | 179372166 | 181424323 | GERD | PTSD | 0.551393 | 0.0110894 | positive | FXR1,LINC02053,PEX5L-AS2,LOC101928882,RNU6-1,DNAJC19,LOC102724604,RNU6-7,TTC14,RNU6-9,RNU6-2,USP13,CCDC39,RNU6-8,PEX5L,SOX2-OT |
| 3 | 181424324 | 183203155 | GERD | PTSD | 0.791232 | 0.0100326 | positive | LAMP3,LINC01206,SNORA63E,LINC01995,SOX2,LINC00888,DCUN1D1,SNORA63D,B3GNT5,LINC01994,MCCC1,SOX2-OT,ATP11B,MCF2L2 |
| 3 | 183203156 | 184524268 | GERD | PTSD | 0.58994 | 0.00200142 | positive | YEATS2,MAGEF1,FAM131A,ECE2,SNORD66,KLHL6,KLHL24,MAP6D1,HTR3C,LOC107986163,AP2M1,HTR3E,CHRD,HTR3D,MIR4448,THPO,CLCN2,PSMD2,EPHB3,ALG3,MIR1224,LINC02054,LINC01840,EEF1AKMT4,VWA5B2,DVL3,EIF4G1,ABCF3,EEF1AKMT4-ECE2,ABCC5,HTR3E-AS1,CAMK2N2,LINC02069,POLR2H,YEATS2-AS1,EIF2B5,KLHL6-AS1,ABCC5-AS1,PARL |
| 4 | 7135820 | 7726034 | GERD | PTSD | 0.596389 | 0.0032669 | positive | PSAPL1,SORCS2,MIR4798,MIR4274 |
| 4 | 7726035 | 8309826 | GERD | PTSD | 0.610415 | 0.00664458 | positive | MIR95,SH3TC1,SORCS2,AFAP1-AS1,ABLIM2,LOC389199,AFAP1,HTRA3 |
| 4 | 17290237 | 18753263 | GERD | PTSD | 0.733413 | 0.000171996 | positive | FAM184B,CLRN2,QDPR,DCAF16,SNORA75B,NCAPG,LAP3,MED28,LCORL |
| 4 | 37880861 | 38984838 | GERD | PTSD | 0.658868 | 0.000436852 | positive | TBC1D1,KLF3,PTTG2,TLR10,TLR6,MIR574,LINC01259,FAM114A1,TLR1,KLF3-AS1,LINC02278,LINC01258,TMEM156,LINC02513 |
| 4 | 60746710 | 61971161 | GERD | PTSD | 0.948588 | 0.00129495 | positive | MIR548AG1 |
| 4 | 61971162 | 63337954 | GERD | PTSD | 0.735785 | 0.00257255 | positive | ADGRL3,LINC02271,ADGRL3-AS1 |
| 4 | 102544804 | 104384534 | GERD | PTSD | 0.693651 | 0.00161969 | positive | LOC102723704,LINC02428,SLC9B1,NFKB1,BDH2,CISD2,UBE2D3-AS1,SLC9B2,MANBA,BANK1,UBE2D3,CENPE,LOC105377621,SLC39A8 |
| 4 | 134109979 | 135809542 | GERD | PTSD | 0.721866 | 0.00310222 | positive | PCDH10,PABPC4L,LINC02462 |
| 4 | 135809543 | 137400031 | GERD | PTSD | 0.721584 | 0.000356549 | positive | LINC02485,LINC00613 |
| 4 | 137400032 | 138537344 | GERD | PTSD | 0.644037 | 0.000283965 | positive | LINC02172,LINC02511,LINC02510,PCDH18 |
| 4 | 139553761 | 141087047 | GERD | PTSD | 0.844363 | 3.40133E-07 | positive | NDUFC1,LOC105377448,NOCT,RAB33B-AS1,MGARP,SETD7,NAA15,MGST2,MAML3,RAB33B,ELF2,LOC101927516 |
| 4 | 182946259 | 183603892 | GERD | PTSD | 0.903718 | 0.00232808 | positive | TENM3-AS1,TENM3,MIR1305 |
| 5 | 6936734 | 8079415 | GERD | PTSD | 0.486729 | 0.00163426 | positive | ADCY2,C5orf49,MTRR,FASTKD3,MIR4454,LINC02123,LOC442132 |
| 5 | 10549030 | 11850850 | GERD | PTSD | 0.643459 | 0.00618265 | positive | DAP,ANKRD33B,CTNND2 |
| 5 | 13608148 | 14910295 | GERD | PTSD | 0.846986 | 0.00183184 | positive | LOC100130744,OTULINL,MIR4637,TRIO,SNORD170,OTULIN,LOC728178,ANKH,SNORD141B,SNORD141A,DNAH5 |
| 5 | 44049879 | 45305566 | GERD | PTSD | 0.800654 | 7.33903E-05 | positive | LINC02224,FGF10,FGF10-AS1,HCN1,MRPS30,MRPS30-DT |
| 5 | 45305567 | 46388848 | GERD | PTSD | 1 | 0.00317201 | positive | HCN1 |
| 5 | 51203266 | 52139773 | GERD | PTSD | 0.847382 | 0.000122334 | positive | ITGA1,LINC02118,PELO |
| 5 | 60930754 | 62180368 | GERD | PTSD | 0.798034 | 0.000712212 | positive | LRRC70,C5orf64,IPO11,LOC101928651,DIMT1,IPO11-LRRC70,C5orf64-AS1,KIF2A |
| 5 | 65606871 | 67096191 | GERD | PTSD | 1 | 0.000473191 | positive | LINC02229,CD180,MAST4,MAST4-AS1,LOC101928858,LINC02242 |
| 5 | 102903986 | 103788460 | GERD | PTSD | 0.719061 | 0.00093073 | positive | NA |
| 5 | 107265912 | 108641784 | GERD | PTSD | 1 | 0.000049181 | positive | FBXL17,LOC285638,LINC01023,FER |
| 5 | 120960738 | 122005291 | GERD | PTSD | 0.793678 | 0.00557919 | positive | LOC100505841,FTMT,MGC32805,LOX,ZNF474,SRFBP1,LINC02201,SNCAIP,LOC101927357 |
| 5 | 124109486 | 125131720 | GERD | PTSD | 0.717692 | 0.00052295 | positive | LOC101927421,LINC02240 |
| 5 | 132554691 | 134556570 | GERD | PTSD | 0.552285 | 0.000225982 | positive | MIR3661,UBE2B,LINC01843,MIR1289-2,PITX1,SEC24A,WSPAR,DDX46,CAMLG,SKP1,JADE2,LOC102546229,C5orf66-AS1,LOC105379183,CDKL3,CATSPER3,TCF7,VDAC1,CDKN2AIPNL,C5orf24,C5orf66,SAR1B,FSTL4,PPP2CA,C5orf15,PCBD2,TXNDC15 |
| 5 | 163781269 | 165289211 | GERD | PTSD | 0.595348 | 0.00065635 | positive | LINC02143,LOC102546299 |
| 5 | 165289212 | 166863413 | GERD | PTSD | 0.500631 | 0.00409153 | positive | LOC101927908,LINC01947,TENM2 |
| 5 | 166863414 | 168342743 | GERD | PTSD | 0.606393 | 0.000221148 | positive | MIR218-2,MIR103B1,PANK3,RARS1,SLIT3,FBLL1,TENM2,WWC1,CTB-178M22.2,MIR12125,MIR103A1,SLIT3-AS2 |
| 5 | 169506967 | 170898340 | GERD | PTSD | 0.372946 | 0.0072924 | positive | MIR3912,DOCK2,RANBP17,LINC01366,KCNIP1-AS1,KCNIP1-OT1,KCNIP1,LOC100128059,FOXI1,KCNMB1,LINC01187,SNORA70J,GABRP,TLX3,NPM1,LCP2,C5orf58,FGF18 |
| 5 | 170898341 | 172285682 | GERD | PTSD | 0.498671 | 0.0100765 | positive | FBXW11,ERGIC1,LOC100288254,NEURL1B,SMIM23,UBTD2,EFCAB9,STK10,LOC101928093,MIR5003,DUSP1,SH3PXD2B,LINC01944 |
| 6 | 7808324 | 9020710 | GERD | PTSD | 0.586153 | 0.011593 | positive | SCARNA27,LOC100506207,SLC35B3,EEF1E1,BLOC1S5,HULC,EEF1E1-BLOC1S5,BLOC1S5-TXNDC5,BMP6,TXNDC5,PIP5K1P1 |
| 6 | 17391995 | 18208676 | GERD | PTSD | 0.782219 | 0.0102265 | positive | LOC105374952,NHLRC1,KDM1B,CAP2,LOC101928491,TPMT,KIF13A,NUP153,FAM8A1 |
| 6 | 23192338 | 23939306 | GERD | PTSD | 0.634032 | 0.00123816 | positive | NA |
| 6 | 24950380 | 25684629 | GERD | PTSD | 0.772601 | 0.00360311 | positive | SCGN,LOC101928663,RIPOR2,CMAHP,CARMIL1 |
| 6 | 39785908 | 40778401 | GERD | PTSD | 0.722453 | 0.000216262 | positive | LRFN2,DAAM2-AS1,DAAM2,TDRG1,LINC00951,MOCS1 |
| 6 | 40778402 | 42103738 | GERD | PTSD | 0.62829 | 0.000563758 | positive | LINC01276,USP49,MED20,APOBEC2,TREML3P,FOXP4-AS1,TREML5P,MIR4641,PRICKLE4,TFEB,FRS3,BYSL,TREML4,TSPO2,NFYA,C6orf132,TREML1,NCR2,LOC101929555,ADCY10P1,OARD1,TOMM6,PGC,TREM2,TREML2,TAF8,FOXP4,MIR10398,CCND3,UNC5CL,TREM1,MDFI |
| 6 | 42103739 | 43770626 | GERD | PTSD | 0.510585 | 0.0111215 | positive | CUL7,TJAP1,C6orf226,MRPL2,DNPH1,MEA1,GUCA1ANB,CUL9,CNPY3,RSPH9,DLK2,XPO5,KLC4,BICRAL,VEGFA,MRPS10,PPP2R5D,MIR6780B,GNMT,LOC401261,ABCC10,PRPH2,SRF,MRPS18A,POLR1C,KLHDC3,TTBK1,CNPY3-GNMT,TBCC,SLC22A7,C6orf132,RRP36,RPL7L1,MAD2L1BP,ATP6V0CP3,GTPBP2,PTK7,LRRC73,UBR2,ZNF318,GUCA1A,CRIP3,PTCRA,POLH,PEX6,TRERF1,YIPF3,GUCA1B |
| 6 | 64010287 | 65759861 | GERD | PTSD | 0.894844 | 3.09805E-05 | positive | PTP4A1,LGSN,EYS,SCAT8,PHF3 |
| 6 | 98173004 | 99678876 | GERD | PTSD | 0.627288 | 0.000510934 | positive | FBXL4,PNKY,POU3F2,MIR548AI,MIR2113 |
| 6 | 101793543 | 102637706 | GERD | PTSD | 0.928399 | 0.000076002 | positive | GRIK2 |
| 6 | 107309328 | 109043244 | GERD | PTSD | 0.640687 | 0.00786675 | positive | SCML4,SNX3,FOXO3,SEC63,OSTM1,NR2E1,SOBP,OSTM1-AS1,BEND3,MTRES1,AFG1L,PDSS2 |
| 6 | 129850179 | 130550137 | GERD | PTSD | 0.506163 | 0.0068765 | positive | SAMD3,TMEM244,ARHGAP18,L3MBTL3 |
| 6 | 143083348 | 144791455 | GERD | PTSD | 0.935697 | 5.41803E-05 | positive | LTV1,HIVEP2,SF3B5,PHACTR2,SNORA98,STX11,PLAGL1,UTRN,ADAT2,LINC01277,PEX3,ZC2HC1B,HYMAI,PHACTR2-AS1,FUCA2,AIG1 |
| 6 | 151629955 | 152376499 | GERD | PTSD | 0.651794 | 0.000532603 | positive | ARMT1,AKAP12,CCDC170,ESR1,RMND1,ZBTB2 |
| 6 | 163336362 | 164687462 | GERD | PTSD | 0.655485 | 2.55393E-06 | positive | CAHM,PACRG-AS3,DKFZp451B082,QKI,PACRG-AS1,PACRG-AS2,LOC102724152,PACRG |
| 6 | 164687463 | 165835953 | GERD | PTSD | 0.595442 | 0.0107522 | positive | C6orf118,MEAT6,PDE10A |
| 6 | 165835954 | 166853325 | GERD | PTSD | 0.430368 | 0.00298406 | positive | LOC101929297,LNCDAT,LINC00602,PDE10A,MPC1,SFT2D1,LOC729681,LINC00473,RPS6KA2,LOC100289495,TBXT,PRR18 |
| 7 | 1366973 | 2473749 | GERD | PTSD | 0.931799 | 1.93626E-08 | positive | NUDT1,LOC100128653,ELFN1,MIR4655,MRM2,CHST12,TFAMP1,INTS1,PSMG3,TMEM184A,SNX8,MAFK,ELFN1-AS1,SNORA114,EIF3B,MAD1L1,MIR6836,LOC105375303,MICALL2,PSMG3-AS1 |
| 7 | 2473750 | 3100211 | GERD | PTSD | 0.900874 | 0.000138485 | positive | BRAT1,TTYH3,CHST12,IQCE,GNA12,MIR4648,AMZ1,CARD11,LFNG,GRIFIN |
| 7 | 3100212 | 4139431 | GERD | PTSD | 0.539583 | 0.00429004 | positive | SDK1,LOC100129603 |
| 7 | 15877565 | 16739013 | GERD | PTSD | 0.701849 | 0.0107558 | positive | SOSTDC1,CRPPA-AS1,LRRC72,CRPPA,BZW2,ANKMY2 |
| 7 | 20122180 | 21390644 | GERD | PTSD | 0.695794 | 0.00847271 | positive | SP8,ITGB8,MACC1-AS1,ABCB5,LOC101927811,LOC101927769,MACC1,LOC100506098,RPL23P8,LOC101927668,LINC01162 |
| 7 | 21390645 | 22165060 | GERD | PTSD | 0.870588 | 9.59844E-05 | positive | SP4,MIR1183,RAPGEF5,DNAH11,CDCA7L |
| 7 | 43448816 | 44989571 | GERD | PTSD | 0.681353 | 0.0101193 | positive | UBE2D4,LINC00957,COA1,OGDH,H2AZ2,PURB,STK17A,CAMK2B,URGCP-MRPS24,NPC1L1,HECW1,LUARIS,MRPS24,ZMIZ2,MIR6837,BLVRA,PGAM2,URGCP,DBNL,YKT6,MIR4657,POLM,MIR6838,PPIA,AEBP1,NUDCD3,MYL7,MIR4649,POLD2,SPDYE1,RASA4CP,GCK,POLR2J4,DDX56,TMED4 |
| 7 | 77663965 | 78685009 | GERD | PTSD | 0.470414 | 0.0085318 | positive | MAGI2,MAGI2-AS2,RPL13AP17 |
| 7 | 81533567 | 82759283 | GERD | PTSD | 0.746169 | 2.56724E-05 | positive | CACNA2D1-AS1,CACNA2D1,PCLO |
| 7 | 92530166 | 93691725 | GERD | PTSD | 0.746967 | 0.00104571 | positive | MIR489,TFPI2-DT,CALCR,MIR653,SAMD9,GNG11,SAMD9L,VPS50,TFPI2,GNGT1,MIR4652,HEPACAM2,BET1,CDK6-AS1 |
| 7 | 113339387 | 115321301 | GERD | PTSD | 0.768622 | 5.45482E-06 | positive | MIR3666,LINC01392,LINC01393,PPP1R3A,FOXP2,SNORA25B,MDFIC |
| 7 | 115321302 | 116835336 | GERD | PTSD | 0.580768 | 0.0031481 | positive | ST7,CAV1,MET,CAV2,TFEC,COMETT,ST7-AS1,LOC102724434,MIR6132,ST7-OT3,TES,ST7-OT4,ST7-AS2,CAPZA2 |
| 7 | 126640873 | 127903738 | GERD | PTSD | 0.628565 | 0.00967271 | positive | LRRC4,LEP,MIR592,GCC1,ZNF800,MIR593,SND1-IT1,MIR129-1,SND1,LOC100506682,PAX4,FSCN3,ARF5,GRM8-AS1,GRM8 |
| 7 | 136113468 | 137657459 | GERD | PTSD | 0.782661 | 0.010012 | positive | LOC349160,CHRM2,CREB3L2,MIR490,CREB3L2-AS1,DGKI,PTN |
| 8 | 19488889 | 20135628 | GERD | PTSD | 0.700773 | 0.00882962 | positive | LPL,LZTS1-AS1,ATP6V1B2,INTS10,CSGALNACT1,LZTS1,SLC18A1 |
| 8 | 33982538 | 34792220 | GERD | PTSD | 0.642759 | 0.00988264 | positive | LINC01288 |
| 8 | 60476249 | 61324889 | GERD | PTSD | 0.915747 | 0.00398112 | positive | CA8,LINC01301 |
| 8 | 66018205 | 66960438 | GERD | PTSD | 0.630392 | 0.00176089 | positive | PDE7A,MTFR1,LINC00251,LINC01299,DNAJC5B,ARMC1 |
| 8 | 92876621 | 94999066 | GERD | PTSD | 0.937604 | 5.15103E-08 | positive | C8orf87,LOC102724710,MIR378D2,RUNX1T1,LINC00535,FLJ46284,TMEM67,RBM12B,RBM12B-AS1,PDP1,CIBAR1,MIR8084,TRIQK |
| 8 | 101448440 | 102548680 | GERD | PTSD | 0.832735 | 0.00211067 | positive | YWHAZ,ZNF706,SNX31,NACA4P,PABPC1,ANKRD46,FLJ42969,ZNNT1,GRHL2,MIR7705 |
| 8 | 119423299 | 120216587 | GERD | PTSD | 0.705639 | 0.00516109 | positive | LOC101927513,SAMD12-AS1,SAMD12,TNFRSF11B,COLEC10 |
| 8 | 136220343 | 137527617 | GERD | PTSD | 1 | 4.72627E-05 | positive | KHDRBS3,LINC01591 |
| 8 | 142611890 | 143611461 | GERD | PTSD | 0.514732 | 0.000499831 | positive | MIR1302-7,LINC00051,ADGRB1,MIR4472-1,TSNARE1,MIR4539 |
| 8 | 143611462 | 144934288 | GERD | PTSD | 0.634993 | 0.00382313 | positive | PYCR3,LOC101928087,THEM6,CDC42P3,ZC3H3,LY6L,TIGD5,GML,SLURP1,LY6H,LY6E-DT,MAPK15,LYNX1,SNORD149,LY6K,FAM83H,RHPN1-AS1,SLURP2,CCDC166,LNCOC1,MIR6845,MROH6,LINC02878,LYPD2,GFUS,LY6E,CYP11B2,EEF1D,ZFP41,GSDMD,LOC105375800,LOC101928160,NAPRT,LOC100310756,ZNF696,MINCR,C8orf31,SCRIB,IQANK1,ZNF623,TOP1MT,PUF60,LY6D,GPIHBP1,PSCA,JRK,ADGRB1,ARC,GLI4,MIR937,NRBP2,CYP11B1,MAFA-AS1,MIR4664,LYNX1-SLURP2,MAFA,ZNF707,RHPN1 |
| 9 | 8262304 | 9170120 | GERD | PTSD | 0.42981 | 0.00935786 | positive | PTPRD-AS1,PTPRD |
| 9 | 14244748 | 14902924 | GERD | PTSD | 0.619436 | 0.00111566 | positive | ZDHHC21,FREM1,NFIB,CER1 |
| 9 | 36749274 | 37687112 | GERD | PTSD | 0.658656 | 0.00317707 | positive | TOMM5,GRHPR,MIR4476,FBXO10,EBLN3P,PAX5,MIR4540,MIR4475,POLR1E,FRMPD1,ZCCHC7,ZBTB5 |
| 9 | 119890465 | 121008529 | GERD | PTSD | 0.817723 | 0.000241577 | positive | SNORA70C,TLR4,ASTN2,LOC101928797 |
| 9 | 121008530 | 122677719 | GERD | PTSD | 0.980767 | 8.93754E-06 | positive | BRINP1,LINC02578 |
| 9 | 132193799 | 132999452 | GERD | PTSD | 0.595914 | 0.00461769 | positive | TOR1A,TOR1B,GPRACR,MIR6855,NCS1,PTGES,ASB6,C9orf78,FNBP1,NTMT1,C9orf50,LINC00963,USP20,GPR107,PRRX2,PRRX2-AS1 |
| 9 | 134141937 | 135135887 | GERD | PTSD | 0.65955 | 0.00881157 | positive | MED27,UCK1,SNORD62A,FAM78A,PLPP7,POMT1,NTNG2,RAPGEF1,PRRC2B,PRRT1B,SNORD62B |
| 9 | 137593528 | 138202112 | GERD | PTSD | 0.626272 | 0.000236907 | positive | MIR3689B,LOC401557,MIR3689A,MIR3689C,MIR3689E,FCN2,MIR3689D1,LOC101448202,MIR3689F,COL5A1,FCN1,OLFM1,MIR3689D2 |
| 10 | 8771921 | 10081279 | GERD | PTSD | 0.623519 | 0.00535673 | positive | LOC101928272,LINC02676,LINC00709 |
| 10 | 11856925 | 12581571 | GERD | PTSD | 0.686345 | 0.00146748 | positive | CDC123,MIR548AK,UPF2,CAMK1D,SEC61A2,PROSER2,NUDT5,PROSER2-AS1,DHTKD1 |
| 10 | 62626466 | 64069687 | GERD | PTSD | 0.892102 | 0.000623438 | positive | LINC00845,ARID5B,TMEM26-AS1,RHOBTB1,CABCOCO1,LOC107984236,TMEM26,RTKN2 |
| 10 | 104206838 | 106142283 | GERD | PTSD | 1 | 0.000259323 | positive | MIR609,WBP1L,ITPRIP,CFAP58,CFAP43,LOC107984265,GSTO2,ACTR1A,PCGF6,CYP17A1,NEURL1-AS1,INA,TAF5,COL17A1,RPARP-AS1,CNNM2,C10orf95,NEURL1,GSTO1,ARL3,CFAP58-DT,TRIM8,MFSD13A,SUFU,CALHM1,CALHM2,BORCS7,SFR1,SLK,CALHM3,ATP5MD,NT5C2,SFXN2,PDCD11,AS3MT,SH3PXD2A-AS1,MIR4482,SH3PXD2A,MIR1307,RPEL1,STN1,BORCS7-ASMT,ITPRIP-AS1,MIR936 |
| 10 | 106142284 | 107877787 | GERD | PTSD | 0.608646 | 0.00730077 | positive | LINC02627,CFAP58,SORCS3-AS1,SORCS3,LINC02620 |
| 10 | 123856185 | 124894142 | GERD | PTSD | 0.512067 | 0.0125368 | positive | DMBT1L1,ACADSB,FAM24B-CUZD1,ARMS2,C10orf88,MIR3941,HTRA1,DMBT1,FAM24A,CUZD1,FAM24B,TACC2,PLEKHA1,C10orf88B,IKZF5,BTBD16,C10orf120,PSTK,LOC112577516 |
| 10 | 125798320 | 126875816 | GERD | PTSD | 0.464548 | 0.0129855 | positive | ABRAXAS2,OAT,ZRANB1,CTBP2,MIR4296,LHPP,EEF1AKMT2,NKX1-2,FAM53B,CHST15,FAM53B-AS1 |
| 11 | 2477450 | 3362508 | GERD | PTSD | -0.65163 | 0.0122548 | negative | MRGPRG,OSBPL5,SLC22A18AS,CDKN1C,KCNQ1,PHLDA2,CARS1-AS1,MRGPRE,SLC22A18,KCNQ1OT1,CARS1,SNORA54,KCNQ1DN,KCNQ1-AS1,NAP1L4,MRGPRG-AS1 |
| 11 | 11632651 | 12470449 | GERD | PTSD | 0.599736 | 0.0101375 | positive | MIR6124,DKK3,PARVA,MICALCL,GALNT18,MIR4299,MICAL2,LINC02547,MIR8070,USP47 |
| 11 | 18740268 | 19780784 | GERD | PTSD | 0.730065 | 0.00754309 | positive | E2F8,CSRP3,LOC100126784,NAV2,PTPN5,ZDHHC13,MRGPRX1,MIR4486,NAV2-AS5,IGSF22,MRGPRX2,NAV2-AS4 |
| 11 | 28577719 | 29530460 | GERD | PTSD | 0.761812 | 0.00535897 | positive | NA |
| 11 | 30570130 | 32065771 | GERD | PTSD | 0.846018 | 6.75671E-05 | positive | MPPED2,ELP4,PAX6,PAX6-AS1,IMMP1L,LOC100506675,DNAJC24,DCDC1,PAUPAR |
| 11 | 56623495 | 57675986 | GERD | PTSD | 0.873882 | 0.00113626 | positive | PRG3,SELENOH,YPEL4,P2RX3,SLC43A1,CTNND1,TIMM10,BTBD18,OR5AK4P,PRG2,CLP1,UBE2L6,RTN4RL2,OR5AK2,APLNR,LINC02735,MED19,SMTNL1,SSRP1,SLC43A3,TMX2-CTNND1,MIR130A,LRRC55,SERPING1,TNKS1BP1,ZDHHC5,TMX2 |
| 11 | 57675987 | 58452708 | GERD | PTSD | 0.722903 | 0.00220115 | positive | ZFP91,CNTF,OR5B21,OR1S1,ZFP91-CNTF,OR9I1,OR5B2,OR9Q1,OR5B3,OR6Q1,OR5B12,OR10W1,OR9Q2,LPXN,OR10Q1,OR1S2,OR5B17 |
| 11 | 112755447 | 113889019 | GERD | PTSD | 0.955375 | 7.64435E-10 | positive | HTR3B,TTC12,NCAM1,NCAM1-AS1,TMPRSS5,HTR3A,ZW10,USP28,DRD2,LOC101928847,MIR4301,CLDN25,ANKK1 |
| 11 | 131424362 | 132584401 | GERD | PTSD | 0.693085 | 0.00513903 | positive | NTM-AS1,NTM,NTM-IT,OPCML |
| 11 | 133521643 | 134351064 | GERD | PTSD | 0.805977 | 0.000022499 | positive | SNORD153,LINC02743,IGSF9B,THYN1,GLB1L3,VPS26B,B3GAT1-DT,MIR4697,JAM3,ACAD8,GLB1L2,LINC02731,SPATA19,NCAPD3,B3GAT1 |
| 12 | 60317 | 1078397 | GERD | PTSD | 0.611315 | 0.00625888 | positive | SLC6A13,KDM5A,IQSEC3,WNK1,LOC574538,CCDC77,RAD52,SLC6A12-AS1,LOC102723544,LOC105369595,NINJ2,B4GALNT3,FAM138D,NINJ2-AS1,SLC6A12,WASH8P |
| 12 | 16136743 | 17374895 | GERD | PTSD | 0.62098 | 0.00138877 | positive | SLC15A5,LMO3,SKP1P2,DERA,MGST1 |
| 12 | 17374896 | 18470023 | GERD | PTSD | 0.742594 | 0.00179212 | positive | LINC02378,PIK3C2G,MIR3974,RERGL |
| 12 | 23923799 | 25058714 | GERD | PTSD | 0.414635 | 0.0108608 | positive | SOX5,BCAT1,MIR920,LINC00477,SOX5-AS1 |
| 12 | 106964978 | 108183188 | GERD | PTSD | 0.863078 | 0.00111906 | positive | PWP1,CRY1,PRDM4,LOC100505978,BTBD11,TMEM263,RFX4,MTERF2,ASCL4,LOC101929162,RIC8B,LOC100287944 |
| 12 | 119301062 | 120567740 | GERD | PTSD | 0.610755 | 0.0021021 | positive | RAB35,GCN1,PRKAB1,HSPB8,SRRM4,LINC00934,TMEM233,CCDC60,CIT,MIR1178,LOC105370024,BICDL1 |
| 12 | 121817510 | 123396634 | GERD | PTSD | 0.609167 | 0.0029644 | positive | MIR9902-2,RSRC2,CLIP1,ZCCHC8,DIABLO,LRRC43,TMEM120B,CLIP1-AS1,CCDC62,HCAR3,VPS37B,KNTC1,KDM2B,LOC100506691,MIR9902-1,B3GNT4,IL31,RNF34,VPS33A,MORN3,ORAI1,DENR,HCAR2,PSMD9,MIR7107,LINC01089,RHOF,MIR548AQ,BCL7A,HIP1R,HCAR1,HPD,CFAP251,LOC101593348,MLXIP,SETD1B |
| 12 | 123396635 | 124843768 | GERD | PTSD | 0.673301 | 0.000215209 | positive | MIR4304,C12orf65,ZNF664-RFLNA,SNORA9B,CDK2AP1,DNAH10,ZNF664,ABCB9,RFLNA,TMED2-DT,PITPNM2-AS1,NCOR2,ARL6IP4,SBNO1-AS1,EIF2B1,TMED2,RILPL1,MIR6880,OGFOD2,RILPL2,MPHOSPH9,PITPNM2,TCTN2,CCDC92,MIR3908,GTF2H3,ATP6V0A2,SBNO1,KMT5A,MIR8072,SNRNP35,DDX55 |
| 12 | 129170297 | 129887418 | GERD | PTSD | 0.504874 | 0.00637605 | positive | TMEM132D-AS2,SLC15A4,TMEM132C,TMEM132D-AS1,GLT1D1,TMEM132D,LOC100128276 |
| 13 | 32986687 | 34722391 | GERD | PTSD | 0.79782 | 0.000882872 | positive | N4BP2L2,MICOS10P1,N4BP2L2-IT2,LINC00423,PDS5B,N4BP2L1,RFC3,KL,STARD13,STARD13-AS,LINC02344 |
| 13 | 35744569 | 36579413 | GERD | PTSD | 0.635376 | 0.00799908 | positive | DCLK1,LINC00445,NBEA,MAB21L1 |
| 13 | 49381728 | 50419896 | GERD | PTSD | 0.82966 | 0.00505104 | positive | RCBTB1,SETDB2,ARL11,PHF11,SETDB2-PHF11,CAB39L,FNDC3A,LOC105370203,MLNR,KPNA3,EBPL,CDADC1 |
| 13 | 58245844 | 59751795 | GERD | PTSD | 0.856852 | 0.000201516 | positive | LINC00374,PCDH17,LINC02338 |
| 13 | 66382287 | 67718879 | GERD | PTSD | 0.599117 | 0.00623765 | positive | PCDH9,PCDH9-AS2,LINC01052,PCDH9-AS4,MIR548X2,MIR4704,PCDH9-AS3 |
| 13 | 100470869 | 101573736 | GERD | PTSD | 0.762591 | 0.000193959 | positive | TMTC4,ZIC2,PCCA-DT,CLYBL,GGACT,NALCN-AS1,PCCA-AS1,LOC101927437,LINC00554,PCCA,ZIC5 |
| 13 | 101573737 | 103050417 | GERD | PTSD | 0.717511 | 0.000733466 | positive | FGF14-IT1,MIR4705,FGF14-AS1,FGF14,NALCN-AS1,LINC00411,NALCN,MIR2681,ITGBL1,FGF14-AS2 |
| 13 | 109813577 | 110995432 | GERD | PTSD | 0.544911 | 0.0119113 | positive | COL4A1,LINC00676,MYO16,COL4A2,LINC00396,MIR8073,IRS2,LINC00399,MYO16-AS1,LOC105370361 |
| 14 | 29029225 | 30831154 | GERD | PTSD | 0.456035 | 0.00134798 | positive | LINC01551,LINC02282,PRKD1,FOXG1-AS1,FOXG1 |
| 14 | 33591114 | 34695195 | GERD | PTSD | 0.596683 | 1.02187E-05 | positive | EGLN3,NPAS3,SNORA89 |
| 14 | 46748239 | 48177483 | GERD | PTSD | 0.529301 | 0.0128515 | positive | MDGA2,LINC00871,RPL10L |
| 14 | 68976913 | 69794542 | GERD | PTSD | 0.874091 | 6.77036E-06 | positive | ACTN1,ACTN1-AS1,EXD2,ZFP36L1,LOC100996664,DCAF5,RAD51B,GALNT16 |
| 14 | 71140427 | 72665319 | GERD | PTSD | 0.578428 | 0.0115683 | positive | TTC9,LOC145474,SNORD56B,LINC01269,PCNX1,MAP3K9,SIPA1L1,RGS6 |
| 14 | 102004378 | 103617472 | GERD | PTSD | 0.894194 | 0.000204114 | positive | CINP,TNFAIP2,MIR1247,DIO3,ANKRD9,LINC00677,RCOR1,PPP2R5C,LINC00239,EXOC3L4,TECPR2,WDR20,MIR4309,CDC42BPB,MOK,LINC02323,TRAF3,DYNC1H1,LBHD2,ZNF839,HSP90AA1,DIO3OS,AMN |
| 15 | 37962916 | 39238840 | GERD | PTSD | 0.650313 | 1.00305E-05 | positive | LINC02345,LINC01852,FAM98B,TMCO5A,RASGRP1,LINC02694,SPRED1 |
| 15 | 56322589 | 57641164 | GERD | PTSD | 0.594184 | 0.00929352 | positive | TCF12,SNORD13D,ZNF280D,LINC01413,TEX9,MNS1,LINC00926,LOC145783,RFX7 |
| 15 | 69089816 | 70767983 | GERD | PTSD | 0.44273 | 0.00417669 | positive | EWSAT1,TLE3,PAQR5,PCAT29,GLCE,RPLP1,LOC145694,NOX5,KIF23,ANP32A-IT1,MIR4312,LINC00593,ANP32A,DRAIC,MIR629,SPESP1 |
| 15 | 94565522 | 95662425 | GERD | PTSD | 0.763248 | 0.000637004 | positive | MCTP2,LINC01581,LOC440311 |
| 15 | 95662426 | 96864278 | GERD | PTSD | 0.532133 | 0.00190664 | positive | LINC01197,LINC00924,NR2F2-AS1,LOC105369212 |
| 16 | 60169 | 1049079 | GERD | PTSD | 0.720374 | 1.15619E-06 | positive | CEROX1,WFIKKN1,RPUSD1,LOC100287175,LUC7L,PGAP6,DDX11L10,WDR24,HAGHL,JMJD8,ARHGDIG,POLR3K,MIR662,LINC02867,NME4,SNRNP25,NHLRC4,CHTF18,MIR3176,MIR6859-4,METTL26,ANTKMT,RHOT2,HBM,HBA2,HBA1,STUB1,CIAO3,HBZ,HBQ1,PRR35,RHBDF1,FBXL16,MPG,METRN,PIGQ,CAPN15,LOC105371038,DECR2,RAB11FIP3,RGS11,LINC00235,NPRL3,GNG13,CCDC78,PRR25,MSLN,MIR5587,PDIA2,RAB40C,MIR6859-2,FAM234A,AXIN1,SOX8,MRPL28,LMF1,LMF1-AS1,MCRIP2,WASIR2,RHBDL1,MIR6859-1,LOC100134368,MIR6859-3,WDR90 |
| 16 | 1049080 | 1996492 | GERD | PTSD | 0.613055 | 0.00178694 | positive | MAPK8IP3,MIR3177,CLCN7,LOC105371046,TMEM204,TPSAB1,FAHD1,C16orf91,TPSG1,CACNA1H,JPT2,IFT140,MRPS34,RPL3L,HS3ST6,TELO2,GNPTG,UNKL,PERCC1,UBE2I,CCDC154,PTX4,NME3,IGFALS,LINC02124,SSTR5-AS1,NUBP2,C1QTNF8,BAIAP3,EME2,LINC00254,MEIOB,SPSB3,TPSB2,SSTR5,CRAMP1,TPSD1,MSRB1,TSR3,HAGH |
| 16 | 1996493 | 3379996 | GERD | PTSD | 0.583807 | 0.00309906 | positive | ZNF598,ELOB,MIR6511B2,SNHG9,TRAF7,PGP,MIR3180-5,RPS2,ERVK13-1,BICDL2,TSC2,MIR6767,NPW,LOC100128770,RNPS1,LOC101929613,CASKIN1,CLDN6,E4F1,CCNF,MIR4516,PRSS33,ZNF75A,ZG16B,FLYWCH1,AMDHD2,SYNGR3,NOXO1,RNF151,KREMEN2,MEFV,SNORD60,MLST8,BRICD5,ZNF213-AS1,NTHL1,SLC9A3R2,FLJ42627,FLYWCH2,MIR6768,TBL3,ZNF200,NDUFB10,CASP16P,MIR940,TBC1D24,OR1F2P,ZNF205,SNORA78,RAB26,ABCA17P,TIGD7,LINC00921,ABCA3,TNFRSF12A,PRSS30P,MIR6511B1,LOC106660606,ZSCAN10,SNORA64,LOC652276,ECI1,PRSS22,ZNF213,THOC6,MIR4717,LOC729652,OR1F1,MMP25,MIR3677,CEMP1,PRSS27,SNHG19,ATP6V0C,SNORA3C,IL32,PRSS21,ZNF263,LINC00514,DNASE1L2,GFER,TEDC2,PKD1,CLDN9,KCTD5,MIR1225,LOC105371049,SRRM2-AS1,MMP25-AS1,RPL3L,ZNF205-AS1,SNORA10,PRSS41,SRRM2,PAQR4,PDPK1,HCFC1R1,NTN3,MIR3178,PKMYT1 |
| 16 | 3379997 | 4816145 | GERD | PTSD | 0.515421 | 0.00582407 | positive | LINC01569,ZNF597,ZNF174,ADCY9,CORO7-PAM16,GLIS2-AS1,ANKS3,LINC02861,MGRN1,DNAJA3,VASN,MTRNR2L4,CLUAP1,NLRC3,ZSCAN32,PAM16,SLX4,UBALD1,NAA60,DNASE1,CDIP1,GLIS2,TFAP4,NUDT16L1,C16orf90,CORO7,MIR6126,NMRAL1,SRL,C16orf96,C16orf71,CREBBP,HMOX2,ZNF500,OR2C1,TRAP1,MIR6769A |
| 16 | 5782969 | 6446081 | GERD | PTSD | 0.790367 | 2.37852E-05 | positive | RBFOX1 |
| 16 | 6446082 | 7051429 | GERD | PTSD | 0.643006 | 0.00921007 | positive | RBFOX1 |
| 16 | 9104470 | 9754796 | GERD | PTSD | 0.960573 | 0.000279874 | positive | LINC01195,LINC02177,C16orf72,LINC01177 |
| 16 | 13893408 | 15921108 | GERD | PTSD | 0.905093 | 0.000262206 | positive | LINC02185,MIR6506,NPIPA3,MIR6511B2,RRN3,PKD1P3-NPIPA1,MRTFB,MIR1972-2,MIR6511A1,MIR6511B1,MIR3180-2,MARF1,NPIPA1,MIR193BHG,MIR3180-1,NPIPA5,LOC100505915,LOC100288162,MIR6511A4,MIR6511A2,MIR3670-2,MIR193B,PLA2G10,PKD1P6-NPIPP1,MIR6511A3,NTAN1,MIR6770-2,MPV17L,MIR3179-2,MIR3180-3,NPIPA2,PDXDC1,PARN,ABCC6P2,MIR3179-4,MIR484,BMERB1,NDE1,MIR3670-3,MIR6770-3,MIR3179-1,MIR6770-1,MIR3670-1,NOMO1,LINC02186,MIR1972-1,MIR3670-4,MIR365A,MIR3180-4,ERCC4,LINC02130,BFAR,MYH11,MIR3179-3 |
| 16 | 25235252 | 26154122 | GERD | PTSD | 0.5868 | 0.000613487 | positive | HS3ST4,AQP8,MIR548W,ZKSCAN2 |
| 16 | 49010762 | 50467033 | GERD | PTSD | 0.457548 | 0.0043344 | positive | TENT4B,LINC02178,CBLN1,ADAM3B,ADCY7,ZNF423,CNEP1R1,HEATR3,C16orf78,BRD7,MIR6771 |
| 16 | 59885500 | 61092844 | GERD | PTSD | 0.709599 | 1.17219E-05 | positive | LINC02141,MIR4426 |
| 16 | 82310205 | 83077869 | GERD | PTSD | 0.622562 | 4.90013E-05 | positive | LOC101928417,MIR8058,CDH13,LOC101928446 |
| 16 | 85146806 | 86058597 | GERD | PTSD | 0.932039 | 0.000507791 | positive | MIR5093,LINC00311,EMC8,GSE1,MIR1910,LINC02139,GINS2,C16orf74,COX4I1,IRF8,MIR6774,LOC101928557,MIR12128 |
| 17 | 36116885 | 37361178 | GERD | PTSD | 0.573186 | 0.0124098 | positive | ARL5C,SRCIN1,FBXO47,TBC1D3H,SNORA21,LINC02079,RPL23,TBC1D3K,TBC1D3E,TBC1D3L,CISD3,RDM1P5,LRRC37A11P,TBC1D3,ARHGAP23,LOC105371763,LOC100287808,C17orf98,MLLT6,SOCS7,PLXDC1,LINC00672,LOC101929950,SNORA21B,RPL19,NPEPPSP1,TBC1D3J,TBC1D3D,MIR6779,CACNB1,EPOP,TBC1D3C,MRPL45,MIR4734,TBC1D3G,MIR4726,CWC25,PCGF2,GPR179,LASP1,PSMB3,TBC1D3F,TBC1D3I,MIR4727,YWHAEP7,PIP4K2B |
| 17 | 76596288 | 77412786 | GERD | PTSD | 0.49242 | 0.0053011 | positive | SCAT1,CYTH1,C1QTNF1-AS1,LGALS3BP,ENGASE,CANT1,USP36,C1QTNF1,TIMP2,RBFOX3,CEP295NL |
| 18 | 5575813 | 6756496 | GERD | PTSD | 0.40708 | 0.00989745 | positive | LINC01387,L3MBTL4,L3MBTL4-AS1,TMEM200C,LOC101927168,MIR3976,MIR4317,EPB41L3,MIR3976HG,ARHGAP28 |
| 18 | 21622717 | 22746153 | GERD | PTSD | -0.638364 | 0.00227158 | negative | MIR320C2,TTC39C,LINC01915,CABYR,HRH4,LOC105372028,ZNF521,LINC01894,IMPACT,OSBPL1A |
| 18 | 41425455 | 42974165 | GERD | PTSD | 0.549089 | 6.72247E-05 | positive | SLC14A2,MIR4319,LINC01478,SETBP1,LINC01601 |
| 18 | 50372824 | 51060986 | GERD | PTSD | 1 | 1.21332E-05 | positive | DCC,MIR4528 |
| 18 | 52512524 | 53762996 | GERD | PTSD | 0.477557 | 0.00235703 | positive | LINC01539,TCF4,CCDC68,LINC01905,TCF4-AS1,RAB27B,LINC01415,MIR4529,LINC01929,LOC642484,LINC01416 |
| 19 | 1515921 | 2253174 | GERD | PTSD | 0.544737 | 0.00549837 | positive | MEX3D,JSRP1,ABHD17A,MBD3,SCAMP4,ATP8B3,AP3D1,BTBD2,PLEKHJ1,MIR1909,TCF3,MIR4321,CSNK1G2-AS1,PLK5,ONECUT3,LOC100288123,MIR6789,DOT1L,UQCR11,MOB3A,MKNK2,MIR1227,AMH,KLF16,IZUMO4,ADAT3,SF3A2,CSNK1G2,REXO1 |
| 19 | 56411087 | 57143509 | GERD | PTSD | 0.670747 | 0.00968874 | positive | NLRP13,ZNF444,ZNF582-AS1,ZNF71-SMIM17,LINC01864,ZNF71,ZNF470,NLRP8,ZNF582,ZFP28,ZNF583,ZSCAN5C,ZNF667,ZSCAN5B,ZNF667-AS1,ZSCAN5A,ZNF542P,EDDM13,GALP,ZNF787,ZNF471,NLRP5 |
| 20 | 15962577 | 16661516 | GERD | PTSD | 0.571313 | 0.00139967 | positive | MACROD2,KIF16B,LOC613266 |
| 20 | 17359291 | 18058524 | GERD | PTSD | 0.653551 | 0.00367717 | positive | BFSP1,SNORD17,RRBP1,DSTN,MGME1,SNX5,OVOL2,BANF2,PCSK2 |
| 20 | 49236419 | 50653620 | GERD | PTSD | 0.587181 | 0.00385331 | positive | RIPOR3-AS1,ATP9A,SALL4,DPM1,BCAS4,KCNG1,MIR3194,ADNP,NFATC2,LINC01429,RIPOR3,MOCS3,ADNP-AS1,PARD6B |
| 20 | 55448076 | 56172843 | GERD | PTSD | 0.496544 | 0.0106444 | positive | RAE1,PCK1,CTCFL,BMP7,RBM38-AS1,MIR4325,SPO11,RBM38,BMP7-AS1,MTRNR2L3 |
| 20 | 58780895 | 59560457 | GERD | PTSD | 0.77393 | 8.26247E-05 | positive | LOC101928048,MIR4533,MIR646HG,MIR548AG2,MIR646 |
| 20 | 62179242 | 62963627 | GERD | PTSD | 0.945376 | 1.9018E-06 | positive | RTEL1-TNFRSF6B,SAMD10,ABHD16B,ZGPAT,PRPF6,MIR6813,TCEA2,STMN3,SOX18,MIR941-5,LKAAEAR1,ZBTB46,GMEB2,SLC2A4RG,LOC112268269,TNFRSF6B,C20orf204,RGS19,MHENCR,LINC00266-1,FNDC11,TPD52L2,UCKL1-AS1,MIR647,ARFRP1,DNAJC5,RTEL1,HELZ2,ZBTB46-AS1,MIR1914,MIR941-1,MIR941-3,LIME1,PCMTD2,UCKL1,MYT1,NPBWR2,ZNF512B,MIR941-4,OPRL1,MIR941-2 |
| 21 | 24055942 | 25247986 | GERD | PTSD | 0.777073 | 0.00328174 | positive | D21S2088E,MIR6130 |
| 21 | 27774351 | 28616539 | GERD | PTSD | 0.840637 | 0.000710959 | positive | CYYR1-AS1,ADAMTS5,ADAMTS1,CYYR1,MIR4759 |
| 21 | 43081435 | 43812791 | GERD | PTSD | 0.599149 | 0.00849539 | positive | TFF1,UMODL1-AS1,LINC00111,PRDM15,TFF3,SNORA91,UMODL1,MIR6814,TMPRSS3,LINC00479,LINC00112,ZNF295-AS1,ZBTB21,ABCG1,RIPK4,C2CD2,TFF2 |
| 22 | 33501726 | 34361897 | GERD | PTSD | 0.724724 | 0.00589915 | positive | LARGE-AS1,LARGE1,LINC01640,SNORA50B,MIR4764 |
| 22 | 38718590 | 40378783 | GERD | PTSD | 0.749986 | 0.000828877 | positive | TPTEP2-CSNK1E,SNORD43,LOC100506472,RPL3,APOBEC3A_B,SUN2,APOBEC3A,FAM227A,APOBEC3C,APOBEC3B,MGAT3,DMC1,SYNGR1,JOSD1,DDX17,CBX7,TAB1,TPTEP2,CACNA1I,GTPBP1,NPTXR,TOMM22,APOBEC3D,SNORD83A,PDGFB,APOBEC3B-AS1,ATF4,APOBEC3G,APOBEC3H,RPS19BP1,LOC105373031,KDELR3,APOBEC3F,DNAL4,ENTHD1,GRAP2,KCNJ4,CBY1,CBX6,SNORD83B,SNORD139,MGAT3-AS1,MIEF1 |
| 1 | 2215496 | 2983519 | GERD | OCD | 0.837318 | 0.000888978 | positive | PRXL2B,PANK4,PLCH2,RER1,TNFRSF14,ACTRT2,MMEL1,TNFRSF14-AS1,PRDM16-DT,MORN1,TTC34,LOC100996583,SKI,LOC100129534,HES5,PEX10 |
| 1 | 36855140 | 38474036 | GERD | OCD | 0.743165 | 0.000457023 | positive | MIR6732,SNIP1,MRPS15,ZC3H12A,MANEAL,FHL3,YRDC,CSF3R,INPP5B,C1orf122,CDCA8,MIR5581,MEAF6,OSCP1,C1orf109,SF3A3,DNALI1,MTF1,GRIK3,LINC01137,EPHA10,GNL2,LSM10,MIR4255,SNORA63C,RSPO1 |
| 1 | 73992171 | 75132092 | GERD | OCD | 0.964896 | 0.000747471 | positive | FPGT,ERICH3-AS1,FPGT-TNNI3K,LINC02238,ERICH3,LRRC53,TNNI3K,LRRIQ3 |
| 1 | 87667573 | 88656689 | GERD | OCD | 0.8315 | 0.000325929 | positive | LMO4,LINC02801,LINC01364 |
| 2 | 59251997 | 60775066 | GERD | OCD | 0.471315 | 0.000365584 | positive | MIR4432HG,BCL11A,LINC01122,LINC01793,MIR4432 |
| 3 | 125992651 | 127495173 | GERD | OCD | 0.75845 | 0.000136414 | positive | ABTB1,CFAP100,LINC01471,CHST13,TPRA1,LINC02016,KLF15,MCM2,NUP210P1,MGLL,MIR7976,MIR6825,UROC1,PLXNA1,PODXL2,C3orf56,ZXDC,CCDC37-DT,CHCHD6,C3orf22,TXNRD3,PRR20G |
| 4 | 97540788 | 98445515 | GERD | OCD | 1 | 0.000456822 | positive | LINC02267,STPG2-AS1 |
| 6 | 150635316 | 151629954 | GERD | OCD | 0.660848 | 0.000294733 | positive | LOC102723831,MIR12131,AKAP12,MTHFD1L,PLEKHG1,IYD |
| 7 | 3100212 | 4139431 | GERD | OCD | 1 | 3.47584E-07 | positive | SDK1,LOC100129603 |
| 9 | 137593528 | 138202112 | GERD | OCD | 0.553028 | 0.000638097 | positive | MIR3689B,LOC401557,MIR3689A,MIR3689C,MIR3689E,FCN2,MIR3689D1,LOC101448202,MIR3689F,COL5A1,FCN1,OLFM1,MIR3689D2 |
| 10 | 33655459 | 35178224 | GERD | OCD | 0.698596 | 0.000303958 | positive | PARD3-AS1,PARD3,LINC00838 |
| 12 | 43072680 | 43980531 | GERD | OCD | -1 | 0.000294439 | negative | LINC02461,ADAMTS20,LINC02450,LINC02451 |
| 12 | 61737954 | 63055936 | GERD | OCD | 0.648447 | 0.000513883 | positive | PPM1H,MIRLET7I,TAFA2,LINC01465,USP15,MIR6125,MON2 |
| 14 | 29029225 | 30831154 | GERD | OCD | 0.551304 | 0.000375116 | positive | LINC01551,LINC02282,PRKD1,FOXG1-AS1,FOXG1 |
| 16 | 5782969 | 6446081 | GERD | OCD | 0.606018 | 0.000574178 | positive | RBFOX1 |
| 16 | 86058598 | 86748867 | GERD | OCD | 0.453705 | 0.000179825 | positive | LINC01081,FOXC2,FOXL1,LINC02135,LINC00917,LINC01082,FOXC2-AS1,FOXF1,FLJ30679,FENDRR,MTHFSD |
| 17 | 47516225 | 48722590 | GERD | OCD | 0.602013 | 1.12262E-05 | positive | LOC100288866,DLX3,MIR6165,PICART1,CHAD,MRPL27,EPN3,LINC02075,NGFR,SGCA,PDK2,MYCBPAP,RSAD1,DLX4,SPOP,PPP1R9B,FLJ45513,SAMD14,SLC35B1,NXPH3,EME1,ITGA3,TAC4,LINC01969,CACNA1G-AS1,CACNA1G,FAM117A,TMEM92-AS1,KAT7,ABCC3,LRRC59,COL1A1,H1-9P,TMEM92,SPATA20,ACSF2,XYLT2,LOC105371824 |
| 18 | 5575813 | 6756496 | GERD | OCD | 0.554133 | 0.00095946 | positive | LINC01387,L3MBTL4,L3MBTL4-AS1,TMEM200C,LOC101927168,MIR3976,MIR4317,EPB41L3,MIR3976HG,ARHGAP28 |
| 18 | 55809316 | 56832312 | GERD | OCD | 0.814953 | 0.000016597 | positive | ALPK2,LINC01926,SEC11C,MALT1,OACYLP,ZNF532,LOC101927322,NEDD4L,SNORA108,MIR122,MIR3591 |
| 21 | 43081435 | 43812791 | GERD | OCD | 0.658736 | 0.00023698 | positive | TFF1,UMODL1-AS1,LINC00111,PRDM15,TFF3,SNORA91,UMODL1,MIR6814,TMPRSS3,LINC00479,LINC00112,ZNF295-AS1,ZBTB21,ABCG1,RIPK4,C2CD2,TFF2 |
| 1 | 6136815 | 7711794 | GERD | ANX | 0.998436 | 2.92603E-06 | positive | KCNAB2,KLHL21,ZBTB48,PHF13,HES2,ACOT7,MIR4252,RNF207,PLEKHG5,TNFRSF25,NOL9,THAP3,CAMTA1-DT,CAMTA1-AS2,CAMTA1,HES3,LINC00337,CHD5,ESPN,DNAJC11,LINC01672,RPL22,ICMT,RNF207-AS1,TAS1R1,GPR153 |
| 1 | 7711795 | 8580987 | GERD | ANX | 0.544397 | 0.0090458 | positive | RERE-AS1,ERRFI1,SLC45A1,VAMP3,SNORD128,RERE,LINC01714,UTS2,PER3,TNFRSF9,CAMTA1,PARK7 |
| 1 | 36855140 | 38474036 | GERD | ANX | 0.473246 | 0.00100061 | positive | MIR6732,SNIP1,MRPS15,ZC3H12A,MANEAL,FHL3,YRDC,CSF3R,INPP5B,C1orf122,CDCA8,MIR5581,MEAF6,OSCP1,C1orf109,SF3A3,DNALI1,MTF1,GRIK3,LINC01137,EPHA10,GNL2,LSM10,MIR4255,SNORA63C,RSPO1 |
| 1 | 38474037 | 40200950 | GERD | ANX | 0.437734 | 0.00248747 | positive | HPCAL4,MACF1,HEYL,PABPC4,AKIRIN1,SNORA55,RHBDL2,UTP11,NT5C1A,PPIEL,GJA9,PABPC4-AS1,MYCBP,RRAGC,NDUFS5,POU3F1,OXCT2P1,GJA9-MYCBP,LINC01343,LOC105378663,BMP8A,MIR3659,MIR3659HG,LINC01685,KIAA0754 |
| 1 | 41093876 | 41977966 | GERD | ANX | 0.683379 | 0.00167421 | positive | NFYC,SLFNL1,RIMS3,CTPS1,FOXO6,KCNQ4,MIR30C1,MIR30E,NFYC-AS1,EDN2,HIVEP3,SLFNL1-AS1,FOXO6-AS1,CITED4,SCMH1 |
| 1 | 41977967 | 43512669 | GERD | ANX | 0.95253 | 2.30045E-05 | positive | C1orf50,ERMAP,SVBP,GUCA2B,GUCA2A,ZNF691,P3H1,YBX1,TMEM269-DT,LOC339539,SLC2A1,PPIH,FOXJ3,CCDC30,CLDN19,PPCS,TMEM269,SLC2A1-AS1,HIVEP3,ZMYND12,RIMKLA |
| 1 | 53114187 | 54771293 | GERD | ANX | 0.495998 | 0.00273149 | positive | TMEM59,CPT2,NDC1,LRRC42,PODN,SLC1A7,CYB5RL,ECHDC2,CZIB,LDLRAD1,DMRTB1,TCEANC2,LINC01771,CDCP2,MIR4781,ZYG11A,SSBP3,MAGOH,SLC25A3P1,SSBP3-AS1,YIPF1,LRP8,LOC100507564,COA7,SHISAL2A,MRPL37,SCP2,DIO1,HSPB11,GLIS1,ZYG11B |
| 1 | 65894185 | 66778015 | GERD | ANX | 0.701264 | 0.00126297 | positive | LEPR,LEPROT,PDE4B,PDE4B-AS1 |
| 1 | 72513120 | 73992170 | GERD | ANX | 0.666775 | 0.00038985 | positive | NEGR1,LINC01360 |
| 1 | 73992171 | 75132092 | GERD | ANX | 0.680616 | 0.00118555 | positive | FPGT,ERICH3-AS1,FPGT-TNNI3K,LINC02238,ERICH3,LRRC53,TNNI3K,LRRIQ3 |
| 1 | 82123369 | 83461215 | GERD | ANX | 0.708103 | 0.00170284 | positive | LINC01361,ADGRL2,LINC01362 |
| 1 | 96147577 | 97721186 | GERD | ANX | 0.927967 | 7.44892E-05 | positive | LINC02790,DPYD-AS1,DPYD,PTBP2,LINC01787 |
| 1 | 176449097 | 177746472 | GERD | ANX | 0.763612 | 0.000215378 | positive | MIR488,BRINP2,LINC01645,LINC01741,ASTN1,PAPPA2 |
| 1 | 196179111 | 197935821 | GERD | ANX | 0.829459 | 0.00208857 | positive | F13B,C1orf53,CFHR5,ZBTB41,ASPM,CFHR1,CFH,MIR4735,KCNT2,CRB1,LHX9,CFHR3,CFHR2,DENND1B,CFHR4 |
| 2 | 1875061 | 2680063 | GERD | ANX | 0.751444 | 0.000109647 | positive | MYT1L-AS1,MYT1L |
| 2 | 12732507 | 13625450 | GERD | ANX | 1 | 0.000126315 | positive | MIR3125,TRIB2,LOC100506474 |
| 2 | 21084899 | 22429640 | GERD | ANX | 0.837342 | 8.24349E-05 | positive | APOB,TDRD15,LINC01822 |
| 2 | 22429641 | 23538527 | GERD | ANX | 0.744451 | 0.000671656 | positive | LINC01884 |
| 2 | 26894103 | 28819510 | GERD | ANX | 0.550327 | 0.00653787 | positive | PLB1,KRTCAP3,LOC100505716,MAPRE3-AS1,PREB,FOSL2,KCNK3,LINC01460,CGREF1,ZNF512,ATRAID,GTF3C2,SLC30A3,DNAJC5G,FNDC4,IFT172,UCN,ZNF513,ABHD1,MRPL33,CCDC121,CENPA,BABAM2-AS1,FLJ31356,TCF23,TMEM214,OST4,PRR30,MIR4263,DPYSL5,AGBL5-AS1,SUPT7L,LOC105374378,TRIM54,CAD,BABAM2,GTF3C2-AS1,KHK,EIF2B4,SLC35F6,PPM1G,C2orf16,RBKS,SNX17,NRBP1,AGBL5,GPN1,FTH1P3,GCKR,MAPRE3,SLC4A1AP,EMILIN1,SLC5A6,LOC100505736,MPV17 |
| 2 | 59251997 | 60775066 | GERD | ANX | 0.475933 | 1.87225E-06 | positive | MIR4432HG,BCL11A,LINC01122,LINC01793,MIR4432 |
| 2 | 64696090 | 65938002 | GERD | ANX | 0.585888 | 0.00562825 | positive | LOC101927402,LINC02245,SLC1A4,CEP68,LINC02579,LINC01800,SERTAD2,SPRED2,RAB1A,AFTPH,LINC01805,ACTR2,MIR4434 |
| 2 | 76649031 | 77372830 | GERD | ANX | 0.643106 | 0.00563045 | positive | LRRTM4,LRRTM4-AS1 |
| 2 | 103305258 | 105120578 | GERD | ANX | 0.634677 | 0.000502212 | positive | LINC01831,LOC100287010,SLC9A2,LINC01102,LINC01796,LINC01935,TMEM182,LINC01103,MFSD9 |
| 2 | 124730992 | 125741528 | GERD | ANX | 0.747666 | 0.000219225 | positive | CNTNAP5,LOC107985820 |
| 2 | 127895645 | 129486897 | GERD | ANX | 0.58835 | 0.00856636 | positive | SAP130,PROC,AMMECR1L,MIR4783,LIMS2,MAP3K2,LOC105373609,GPR17,CYP27C1,IWS1,POLR2D,ERCC3,HS6ST1,UGGT1,MYO7B,WDR33,SFT2D3 |
| 2 | 143259781 | 144503688 | GERD | ANX | 0.597681 | 0.000602376 | positive | KYNU,ARHGAP15 |
| 2 | 155207970 | 156130947 | GERD | ANX | 0.681016 | 0.00392223 | positive | KCNJ3,LOC100144595,GALNT13 |
| 2 | 165917789 | 167488576 | GERD | ANX | 0.899153 | 0.00216962 | positive | SCN2A,CSRNP3,SCN1A-AS1,LOC100506124,TTC21B,SCN1A,LOC102724058,SCN3A,GALNT3,SCN7A,TTC21B-AS1,SCN9A |
| 2 | 169224446 | 170155561 | GERD | ANX | 0.546936 | 0.00950578 | positive | LRP2,CERS6,SPC25,MIR4774,ABCB11,G6PC2,NOSTRIN,DHRS9,CERS6-AS1 |
| 2 | 193033983 | 195300250 | GERD | ANX | 0.789886 | 0.000249351 | positive | PCGEM1,LINC01821,TMEFF2 |
| 2 | 199654970 | 201324132 | GERD | ANX | 0.928878 | 0.000189959 | positive | C2orf69,SPATS2L,LINC01877,SATB2-AS1,MAIP1,FTCDNL1,SATB2,TYW5 |
| 2 | 220771553 | 221445191 | GERD | ANX | 1 | 5.10666E-05 | positive | NA |
| 2 | 227557842 | 228812485 | GERD | ANX | 0.746197 | 3.39476E-05 | positive | MIR5703,SLC19A3,C2orf83,MFF,AGFG1,RHBDD1,CCL20,LOC729968,IRS1,DAW1,TM4SF20,COL4A4,COL4A3,MFF-DT |
| 2 | 239634351 | 240325600 | GERD | ANX | 0.783717 | 0.00551082 | positive | LOC100287387,MIR4440,LINC01940,TWIST2,HDAC4,MIR4269,MIR4441,MIR2467,MGC16025,HDAC4-AS1 |
| 3 | 2140012 | 3128152 | GERD | ANX | 0.579183 | 0.000318733 | positive | IL5RA,CNTN4-AS1,CNTN4,CNTN4-AS2 |
| 3 | 8664893 | 9970731 | GERD | ANX | 0.869903 | 1.94452E-05 | positive | JAGN1,BRPF1,TADA3,THUMPD3-AS1,RPUSD3,SRGAP3,THUMPD3,IL17RE,SSUH2,TTLL3,RAD18,CIDEC,MTMR14,ARPC4,OXTR,IL17RC,CPNE9,SRGAP3-AS2,LHFPL4,SRGAP3-AS4,SETD5,ARPC4-TTLL3,CAV3,CAMK1,SRGAP3-AS3,OGG1 |
| 3 | 20754843 | 21790867 | GERD | ANX | 0.501361 | 0.00830199 | positive | ZNF385D,VENTXP7,ZNF385D-AS1 |
| 3 | 35009560 | 35913033 | GERD | ANX | 0.885334 | 0.00088416 | positive | LOC101928135,ARPP21,MIR128-2 |
| 3 | 47588462 | 50387742 | GERD | ANX | 0.831086 | 2.17833E-05 | positive | APEH,GNAI2,MAP4,SLC26A6,SLC25A20,MIR4443,LSMEM2,C3orf84,NPRL2,NCKIPSD,SEMA3B-AS1,SNORA94,MIR191,ZNF589,MIR5787,MIR6824,GPX1,TREX1,DALRD3,PLXNB1,CCDC51,NDUFAF3,LINC02585,RBM6,C3orf62,TMEM89,MIR5193,HYAL1,FCF1P2,CDC25A,RHOA,UCN2,ARIH2OS,RBM5,MIR4271,HYAL3,CAMP,SEMA3B,QARS1,CELSR3,QRICH1,RASSF1-AS1,NICN1,IFRD2,IMPDH2,CDHR4,NAA80,USP4,ATRIP,SHISA5,NME6,ARIH2,DHX30,SEMA3F,GNAT1,TMA7,KLHDC8B,MIR2115,MIR6890,SPINK8,IP6K2,COL7A1,RNF123,RBM5-AS1,SMARCC1,MIR4793,CCDC71,GMPPB,IP6K1,WDR6,RASSF1,UQCRC1,IHO1,UBA7,BSN-DT,TUSC2,INKA1,LAMB2,ATRIP-TREX1,PRKAR2A,MIR711,FBXW12,MON1A,TCTA,CSPG5,MIR1226,MIR425,MIR6872,MIR6823,MST1,P4HTM,BSN,SLC38A3,HYAL2,DAG1,CAMKV,AMT,ZMYND10,PRKAR2A-AS1,TRAIP,USP19,SNORD146,LAMB2P1,MST1R,PFKFB4,SEMA3F-AS1,AMIGO3 |
| 3 | 56954879 | 58557718 | GERD | ANX | 0.754283 | 0.0024004 | positive | RPP14,DNAH12,LOC105377102,ARF4-AS1,PXK,FLNB-AS1,SLMAP,ABHD6,ARHGEF3-AS1,DNASE1L3,PDHB,FLNB,KCTD6,ASB14,HTD2,SPATA12,IL17RD,DENND6A-DT,PDE12,ARHGEF3,HESX1,FAM107A,APPL1,ACOX2,ARF4,DENND6A |
| 3 | 65326752 | 66702084 | GERD | ANX | 0.467755 | 0.00165867 | positive | MAGI1-AS1,MAGI1,MAGI1-IT1,SLC25A26,LRIG1 |
| 3 | 68319771 | 69784969 | GERD | ANX | 0.566981 | 0.00295128 | positive | EOGT,UBA3,TAFA4,MIR3136,LMOD3,ARL6IP5,TMF1,FRMD4B,LOC105377146,TAFA1 |
| 3 | 69784970 | 71223281 | GERD | ANX | 0.906608 | 1.13316E-06 | positive | FOXP1,MDFIC2,MITF,SAMMSON |
| 3 | 71223282 | 72334704 | GERD | ANX | 0.37709 | 0.00650645 | positive | LINC00877,FOXP1,MIR1284,LINC00870,EIF4E3,GPR27,PROK2,FOXP1-AS1 |
| 3 | 76157471 | 77507832 | GERD | ANX | 0.596002 | 0.00274233 | positive | ROBO2 |
| 3 | 112408514 | 113657665 | GERD | ANX | 1 | 0.000299933 | positive | SPICE1,NAA50,USF3,CD200R1,LINC02044,MIR9900,NEPRO,GTPBP8,CFAP44-AS1,CFAP44,ATP6V1A,BOC,CD200R1L-AS1,LINC02042,MIR8076,GRAMD1C,MIR4446,SIDT1,CD200R1L |
| 3 | 118086930 | 118903053 | GERD | ANX | 0.970277 | 0.00128866 | positive | IGSF11,UPK1B,IGSF11-AS1,LOC105374060,LOC101926968,TEX55 |
| 3 | 149998412 | 151131307 | GERD | ANX | 0.858409 | 0.00235729 | positive | P2RY12,TSC22D2,EIF2A,MINDY4B,CLRN1-AS1,MED12L,LINC01214,ERICH6-AS1,GPR171,ERICH6,LOC101928105,P2RY13,SERP1,P2RY14,GPR87,CLRN1,SIAH2,SELENOT |
| 3 | 187939200 | 189451805 | GERD | ANX | 0.53536 | 0.00471769 | positive | MIR28,TPRG1-AS1,TPRG1-AS2,TPRG1,TP63,LPP,LPP-AS1 |
| 4 | 60746710 | 61971161 | GERD | ANX | 0.5089 | 0.00821516 | positive | MIR548AG1 |
| 4 | 61971162 | 63337954 | GERD | ANX | 0.435143 | 0.00703311 | positive | ADGRL3,LINC02271,ADGRL3-AS1 |
| 4 | 137400032 | 138537344 | GERD | ANX | 0.547066 | 0.0010761 | positive | LINC02172,LINC02511,LINC02510,PCDH18 |
| 4 | 177129679 | 178314527 | GERD | ANX | 0.583776 | 0.00877561 | positive | VEGFC,LINC02509,ASB5,NEIL3,SPCS3 |
| 4 | 184338585 | 185483262 | GERD | ANX | 0.494641 | 0.00170201 | positive | CDKN2AIP,RWDD4,ING2,LINC02363,LINC02362,TRAPPC11,ENPP6,LOC389247,IRF2,STOX2,LINC02427 |
| 5 | 3677596 | 4636542 | GERD | ANX | 0.774156 | 0.0016112 | positive | NA |
| 5 | 6936734 | 8079415 | GERD | ANX | 0.486865 | 0.00513501 | positive | ADCY2,C5orf49,MTRR,FASTKD3,MIR4454,LINC02123,LOC442132 |
| 5 | 25722098 | 26938178 | GERD | ANX | 0.654316 | 0.00693964 | positive | CDH9 |
| 5 | 60930754 | 62180368 | GERD | ANX | 0.90011 | 0.000369815 | positive | LRRC70,C5orf64,IPO11,LOC101928651,DIMT1,IPO11-LRRC70,C5orf64-AS1,KIF2A |
| 5 | 65606871 | 67096191 | GERD | ANX | 0.765842 | 0.00245089 | positive | LINC02229,CD180,MAST4,MAST4-AS1,LOC101928858,LINC02242 |
| 5 | 82720249 | 84094797 | GERD | ANX | 0.429784 | 0.00738097 | positive | EDIL3,HAPLN1,VCAN-AS1,VCAN |
| 5 | 102903986 | 103788460 | GERD | ANX | 0.540784 | 0.00339942 | positive | NA |
| 5 | 119664174 | 120371857 | GERD | ANX | 0.854612 | 0.000492555 | positive | PRR16 |
| 5 | 132554691 | 134556570 | GERD | ANX | 0.77781 | 2.03605E-07 | positive | MIR3661,UBE2B,LINC01843,MIR1289-2,PITX1,SEC24A,WSPAR,DDX46,CAMLG,SKP1,JADE2,LOC102546229,C5orf66-AS1,LOC105379183,CDKL3,CATSPER3,TCF7,VDAC1,CDKN2AIPNL,C5orf24,C5orf66,SAR1B,FSTL4,PPP2CA,C5orf15,PCBD2,TXNDC15 |
| 5 | 145319097 | 147180171 | GERD | ANX | 0.55612 | 0.00550569 | positive | LARS1,PPP2R2B,PLAC8L1,TCERG1,STK32A,RBM27,JAKMIP2-AS1,JAKMIP2,GPR151,PPP2R2B-IT1,SH3RF2,DPYSL3,POU4F3,STK32A-AS1 |
| 5 | 165289212 | 166863413 | GERD | ANX | 0.435218 | 0.000249644 | positive | LOC101927908,LINC01947,TENM2 |
| 5 | 168342744 | 169506966 | GERD | ANX | 0.547739 | 0.00266597 | positive | SPDL1,DOCK2,MIR585,SLIT3,SLIT3-AS1,INSYN2B,MIR378E |
| 6 | 63979 | 1380679 | GERD | ANX | 0.420066 | 0.00410588 | positive | LINC01622,LINC00266-3,IRF4,HUS1B,FOXQ1,LOC285766,EXOC2,DUSP22,LOC101927691 |
| 6 | 15069422 | 16566882 | GERD | ANX | -0.449945 | 0.00722706 | negative | JARID2,JARID2-AS1,ATXN1,DTNBP1,MYLIP,MIR4639,GMPR |
| 6 | 16566883 | 17391994 | GERD | ANX | 0.769318 | 0.0011959 | positive | ATXN1-AS1,RBM24,STMND1,ATXN1 |
| 6 | 22199714 | 23192337 | GERD | ANX | 0.883005 | 0.00472276 | positive | HDGFL1,LOC105374972,PRL |
| 6 | 40778402 | 42103738 | GERD | ANX | 0.558212 | 1.33521E-05 | positive | LINC01276,USP49,MED20,APOBEC2,TREML3P,FOXP4-AS1,TREML5P,MIR4641,PRICKLE4,TFEB,FRS3,BYSL,TREML4,TSPO2,NFYA,C6orf132,TREML1,NCR2,LOC101929555,ADCY10P1,OARD1,TOMM6,PGC,TREM2,TREML2,TAF8,FOXP4,MIR10398,CCND3,UNC5CL,TREM1,MDFI |
| 6 | 42103739 | 43770626 | GERD | ANX | 0.438958 | 0.00894291 | positive | CUL7,TJAP1,C6orf226,MRPL2,DNPH1,MEA1,GUCA1ANB,CUL9,CNPY3,RSPH9,DLK2,XPO5,KLC4,BICRAL,VEGFA,MRPS10,PPP2R5D,MIR6780B,GNMT,LOC401261,ABCC10,PRPH2,SRF,MRPS18A,POLR1C,KLHDC3,TTBK1,CNPY3-GNMT,TBCC,SLC22A7,C6orf132,RRP36,RPL7L1,MAD2L1BP,ATP6V0CP3,GTPBP2,PTK7,LRRC73,UBR2,ZNF318,GUCA1A,CRIP3,PTCRA,POLH,PEX6,TRERF1,YIPF3,GUCA1B |
| 6 | 64010287 | 65759861 | GERD | ANX | 0.858498 | 0.00106957 | positive | PTP4A1,LGSN,EYS,SCAT8,PHF3 |
| 6 | 93696826 | 95176337 | GERD | ANX | 0.558873 | 0.00291997 | positive | TSG1,EPHA7 |
| 6 | 98173004 | 99678876 | GERD | ANX | 0.697324 | 5.58623E-05 | positive | FBXL4,PNKY,POU3F2,MIR548AI,MIR2113 |
| 6 | 101793543 | 102637706 | GERD | ANX | 0.706262 | 0.00751581 | positive | GRIK2 |
| 6 | 107309328 | 109043244 | GERD | ANX | 0.900349 | 1.12931E-06 | positive | SCML4,SNX3,FOXO3,SEC63,OSTM1,NR2E1,SOBP,OSTM1-AS1,BEND3,MTRES1,AFG1L,PDSS2 |
| 6 | 143083348 | 144791455 | GERD | ANX | 0.480387 | 0.000693161 | positive | LTV1,HIVEP2,SF3B5,PHACTR2,SNORA98,STX11,PLAGL1,UTRN,ADAT2,LINC01277,PEX3,ZC2HC1B,HYMAI,PHACTR2-AS1,FUCA2,AIG1 |
| 6 | 155523958 | 156762740 | GERD | ANX | 0.56875 | 0.00675783 | positive | CLDN20,NOX3,MIR1202,LOC105378068,SNORD28B,TFB1M,TIAM2 |
| 6 | 165835954 | 166853325 | GERD | ANX | 0.435985 | 0.00441311 | positive | LOC101929297,LNCDAT,LINC00602,PDE10A,MPC1,SFT2D1,LOC729681,LINC00473,RPS6KA2,LOC100289495,TBXT,PRR18 |
| 7 | 1366973 | 2473749 | GERD | ANX | 0.52148 | 0.00161308 | positive | NUDT1,LOC100128653,ELFN1,MIR4655,MRM2,CHST12,TFAMP1,INTS1,PSMG3,TMEM184A,SNX8,MAFK,ELFN1-AS1,SNORA114,EIF3B,MAD1L1,MIR6836,LOC105375303,MICALL2,PSMG3-AS1 |
| 7 | 2473750 | 3100211 | GERD | ANX | 0.658189 | 5.84007E-05 | positive | BRAT1,TTYH3,CHST12,IQCE,GNA12,MIR4648,AMZ1,CARD11,LFNG,GRIFIN |
| 7 | 27351287 | 28890886 | GERD | ANX | 0.593208 | 0.00479937 | positive | TAX1BP1,CREB5,JAZF1-AS1,HIBADH,TSL,JAZF1 |
| 7 | 28890887 | 30175113 | GERD | ANX | 0.544795 | 0.0009804 | positive | PRR15,DPY19L2P3,FKBP14,LOC101928168,WIPF3,CHN2,MTURN,TRIL,CPVL,LOC100506497,LOC646762,ZNRF2P2,SCRN1,MIR550A3,PLEKHA8,CHN2-AS1 |
| 7 | 31730555 | 32460228 | GERD | ANX | 0.992276 | 2.38977E-05 | positive | PDE1C,PPP1R17 |
| 7 | 43448816 | 44989571 | GERD | ANX | 0.714476 | 0.00215547 | positive | UBE2D4,LINC00957,COA1,OGDH,H2AZ2,PURB,STK17A,CAMK2B,URGCP-MRPS24,NPC1L1,HECW1,LUARIS,MRPS24,ZMIZ2,MIR6837,BLVRA,PGAM2,URGCP,DBNL,YKT6,MIR4657,POLM,MIR6838,PPIA,AEBP1,NUDCD3,MYL7,MIR4649,POLD2,SPDYE1,RASA4CP,GCK,POLR2J4,DDX56,TMED4 |
| 7 | 77663965 | 78685009 | GERD | ANX | 0.547349 | 5.92143E-05 | positive | MAGI2,MAGI2-AS2,RPL13AP17 |
| 7 | 81533567 | 82759283 | GERD | ANX | 0.444096 | 0.00632682 | positive | CACNA2D1-AS1,CACNA2D1,PCLO |
| 7 | 113339387 | 115321301 | GERD | ANX | 0.681729 | 0.000137445 | positive | MIR3666,LINC01392,LINC01393,PPP1R3A,FOXP2,SNORA25B,MDFIC |
| 7 | 125389587 | 126640872 | GERD | ANX | 0.591991 | 0.00620619 | positive | GRM8 |
| 7 | 133807217 | 134943221 | GERD | ANX | 0.712369 | 0.00126004 | positive | WDR91,AKR1B1,MIR6509,STRA8,CALD1,AKR1B15,LRGUK,CYREN,TMEM140,AKR1B10,SLC35B4,AGBL3,BPGM |
| 7 | 136113468 | 137657459 | GERD | ANX | 0.713085 | 0.000436251 | positive | LOC349160,CHRM2,CREB3L2,MIR490,CREB3L2-AS1,DGKI,PTN |
| 8 | 4893121 | 5566269 | GERD | ANX | 0.527346 | 0.0089781 | positive | NA |
| 8 | 16988031 | 17492672 | GERD | ANX | 0.693462 | 0.00419878 | positive | VPS37A,ZDHHC2,PDGFRL,MTMR7,SLC7A2,CNOT7 |
| 8 | 58675118 | 59555977 | GERD | ANX | 0.669509 | 0.00146365 | positive | NSMAF,LINC01602,SDCBP,LOC101929528,UBXN2B,FAM110B,CYP7A1 |
| 8 | 59555978 | 60476248 | GERD | ANX | 0.723406 | 0.000400562 | positive | TOX,NSMAF,LOC100505501 |
| 8 | 73838028 | 75011699 | GERD | ANX | 0.682729 | 0.000236406 | positive | RDH10,KCNB2,SBSPON,TMEM70,RPL7,C8orf89,TERF1,LY96,STAU2,RDH10-AS1,STAU2-AS1,ELOC,UBE2W,LINC01617 |
| 8 | 92876621 | 94999066 | GERD | ANX | 0.888557 | 7.25667E-08 | positive | C8orf87,LOC102724710,MIR378D2,RUNX1T1,LINC00535,FLJ46284,TMEM67,RBM12B,RBM12B-AS1,PDP1,CIBAR1,MIR8084,TRIQK |
| 8 | 139404632 | 140355988 | GERD | ANX | 0.565885 | 0.00349988 | positive | COL22A1,FAM135B |
| 8 | 140355989 | 141527881 | GERD | ANX | 0.736996 | 0.00009883 | positive | TRAPPC9,CHRAC1,KCNK9,PEG13 |
| 8 | 141527882 | 142611889 | GERD | ANX | 0.561033 | 0.00525926 | positive | AGO2,SLC45A4,PTK2,DENND3,GPR20,LINC01300,PTP4A3,LOC105375787,MIR151A,MROH5 |
| 8 | 142611890 | 143611461 | GERD | ANX | 0.441889 | 0.00512214 | positive | MIR1302-7,LINC00051,ADGRB1,MIR4472-1,TSNARE1,MIR4539 |
| 9 | 4025356 | 4596614 | GERD | ANX | 0.51424 | 0.00714921 | positive | SLC1A1,SPATA6L,GLIS3 |
| 9 | 19463293 | 20483880 | GERD | ANX | 0.584338 | 0.00372568 | positive | MLLT3,MIR4473,SLC24A2 |
| 9 | 34704545 | 36749273 | GERD | ANX | 0.887015 | 7.42212E-05 | positive | TMEM8B,RNF38,GLIPR2,SPAAR,OR13J1,SIT1,OR2S2,TLN1,MIR4667,GNE,ARHGEF39,RMRP,HRCT1,FAM205A,MIR6852,RUSC2,DNAJB5,CLTA,CD72,C9orf131,VCP,STOML2,FAM166B,ATP8B5P,FAM205BP,PHF24,FAM205C,MELK,FANCG,PIGO,GBA2,FAM214B,LOC101926948,UNC13B,CA9,CCDC107,NPR2,TESK1,RECK,CCIN,RGP1,CCL21,TPM2,FAM221B,CREB3,MSMP,SPAG8,DNAJB5-DT,MIR6853,HINT2 |
| 9 | 121008530 | 122677719 | GERD | ANX | 0.434107 | 0.000720509 | positive | BRINP1,LINC02578 |
| 9 | 126092932 | 127184582 | GERD | ANX | 0.537975 | 0.00353124 | positive | PSMB7,LOC613206,MIR7150,LHX2,LOC100505588,NEK6,MIR601,DENND1A,CRB2,LOC100129034 |
| 9 | 136770927 | 137593527 | GERD | ANX | 0.802903 | 0.00460247 | positive | MIR4669,WDR5,BRD3OS,COL5A1,LINC02247,COL5A1-AS1,RXRA,LOC100130548,VAV2,BRD3,RNU6ATAC |
| 10 | 7659873 | 8771920 | GERD | ANX | 0.554706 | 0.00038462 | positive | ATP5F1C,ITIH5,GATA3,TAF3,KIN,GATA3-AS1,LINC00708,ITIH2,LOC105376398 |
| 10 | 14820756 | 15946677 | GERD | ANX | -0.782788 | 0.00158538 | negative | PPIAP30,HSPA14,RPP38-DT,CDNF,FAM171A1,OLAH,ITGA8,DCLRE1CP1,MINDY3,SUV39H2,MEIG1,ACBD7,RPP38,DCLRE1C,LOC100421372,NMT2,ACBD7-DCLRE1CP1 |
| 10 | 33655459 | 35178224 | GERD | ANX | 0.568 | 0.00146279 | positive | PARD3-AS1,PARD3,LINC00838 |
| 10 | 106142284 | 107877787 | GERD | ANX | 0.826294 | 1.15765E-07 | positive | LINC02627,CFAP58,SORCS3-AS1,SORCS3,LINC02620 |
| 10 | 114255955 | 115588903 | GERD | ANX | 0.461808 | 0.00237443 | positive | MIR4295,SNORA87,MIR4483,HABP2,PLEKHS1,TCF7L2,CASP7,NRAP,LOC103344931,VTI1A |
| 10 | 122803507 | 123856184 | GERD | ANX | 0.59673 | 0.00300518 | positive | ATE1-AS1,TACC2,ATE1,NSMCE4A,FGFR2 |
| 11 | 30570130 | 32065771 | GERD | ANX | 0.65975 | 0.000629805 | positive | MPPED2,ELP4,PAX6,PAX6-AS1,IMMP1L,LOC100506675,DNAJC24,DCDC1,PAUPAR |
| 11 | 32065772 | 32830422 | GERD | ANX | 0.589037 | 0.00802855 | positive | WT1,RCN1,SNORA88,CCDC73,EIF3M,WT1-AS |
| 11 | 77719871 | 78912252 | GERD | ANX | 0.64676 | 0.00147506 | positive | NDUFC2-KCTD14,LINC02728,TENM4,GAB2,LOC101928896,KCTD21-AS1,KCTD14,NDUFC2,THRSP,KCTD21,ALG8,NARS2,USP35 |
| 11 | 111117781 | 112755446 | GERD | ANX | 0.768676 | 0.001506 | positive | HSPB2,COLCA2,HOATZ,BTG4,MIR34C,C11orf52,MIR34B,ALG9,BCO2,IL18,NKAPD1,CRYAB,PLET1,COLCA1,DIXDC1,LINC02762,C11orf53,POU2AF1,SDHD,SIK2,HSPB2-C11orf52,PTS,LOC100132078,FDXACB1,DLAT,LINC02763,LAYN,LOC100132686,TIMM8B,MIR4491,PPP2R1B,TEX12,C11orf1,LOC728196,PIH1D2,LINC02764 |
| 11 | 112755447 | 113889019 | GERD | ANX | 0.829662 | 1.5664E-09 | positive | HTR3B,TTC12,NCAM1,NCAM1-AS1,TMPRSS5,HTR3A,ZW10,USP28,DRD2,LOC101928847,MIR4301,CLDN25,ANKK1 |
| 11 | 121208187 | 122589224 | GERD | ANX | 0.669286 | 0.00238468 | positive | MIRLET7A2,LOC105369535,BLID,MIR125B1,UBASH3B,MIR100,MIR10526,SORL1,MIR100HG |
| 11 | 126304241 | 127306871 | GERD | ANX | 0.506764 | 0.00279052 | positive | LINC02712,LOC101929473,KIRREL3-AS3,KIRREL3,MIR3167,KIRREL3-AS2,LOC101929427 |
| 11 | 133521643 | 134351064 | GERD | ANX | 0.522076 | 0.00709134 | positive | SNORD153,LINC02743,IGSF9B,THYN1,GLB1L3,VPS26B,B3GAT1-DT,MIR4697,JAM3,ACAD8,GLB1L2,LINC02731,SPATA19,NCAPD3,B3GAT1 |
| 12 | 61737954 | 63055936 | GERD | ANX | 0.964987 | 1.48529E-07 | positive | PPM1H,MIRLET7I,TAFA2,LINC01465,USP15,MIR6125,MON2 |
| 12 | 77800464 | 79315178 | GERD | ANX | 0.584972 | 0.00224428 | positive | SYT1,NAV3,LINC02424 |
| 12 | 102966634 | 104140831 | GERD | ANX | 0.781399 | 0.00162917 | positive | STAB2,LOC101929058,LOC105369945,PAH,ASCL1,LINC00485,C12orf42,LINC02401 |
| 12 | 110111515 | 111592381 | GERD | ANX | 0.676572 | 0.00832336 | positive | CCDC63,ANAPC7,LINC01405,MIR4497,PPTC7,FAM222A,GPN3,GIT2,VPS29,RAD9B,ATP2A2,C12orf76,FAM216A,TCHP,MYL2,LOC105369980,IFT81,CUX2,TRPV4,TCTN1,ANKRD13A,FAM222A-AS1,GLTP,HVCN1,ARPC3,PPP1CC |
| 12 | 111592382 | 113947983 | GERD | ANX | 0.490923 | 0.00950645 | positive | MIR6762,OAS1,DDX54,LHX5-AS1,CFAP73,ATXN2,TRAFD1,SH2B3,PTPN11,RPL6,IQCD,MIR6761,MIR3657,HECTD4,ATXN2-AS,RPH3A,MIR7106,MIR1302-1,ACAD10,MAPKAPK5,DTX1,PHETA1,TMEM116,PLBD2,SDS,OAS2,RITA1,OAS3,ALDH2,MAPKAPK5-AS1,CUX2,BRAP,TPCN1,ERP29,MIR6861,RASAL1,NAA25,MIR6760,SLC8B1,SDSL,LHX5 |
| 12 | 120567741 | 121817509 | GERD | ANX | 0.620086 | 0.00414529 | positive | NRAV,PXN-AS1,RNF10,MIR4498,ACADS,RNU4-1,HNF1A,P2RX4,SPPL3,ANAPC5,UNC119B,RNU4-2,CABP1,HNF1A-AS1,P2RX7,SIRT4,POP5,MLEC,TRIAP1,PLA2G1B,COX6A1,RPLP0,MIR4700,CAMKK2,XLOC_009911,GCN1,OASL,GATC,C12orf43,COQ5,SRSF9,MSI1,DYNLL1,PXN |
| 12 | 123396635 | 124843768 | GERD | ANX | 0.796441 | 0.00224739 | positive | MIR4304,C12orf65,ZNF664-RFLNA,SNORA9B,CDK2AP1,DNAH10,ZNF664,ABCB9,RFLNA,TMED2-DT,PITPNM2-AS1,NCOR2,ARL6IP4,SBNO1-AS1,EIF2B1,TMED2,RILPL1,MIR6880,OGFOD2,RILPL2,MPHOSPH9,PITPNM2,TCTN2,CCDC92,MIR3908,GTF2H3,ATP6V0A2,SBNO1,KMT5A,MIR8072,SNRNP35,DDX55 |
| 13 | 31433219 | 32986686 | GERD | ANX | 0.765023 | 7.15078E-06 | positive | RXFP2,BRCA2,ZAR1L,TEX26-AS1,MEDAG,TEX26,N4BP2L1,LINC00545,FRY-AS1,B3GLCT,HSPH1,FRY,EEF1DP3 |
| 13 | 32986687 | 34722391 | GERD | ANX | 0.664962 | 0.000123726 | positive | N4BP2L2,MICOS10P1,N4BP2L2-IT2,LINC00423,PDS5B,N4BP2L1,RFC3,KL,STARD13,STARD13-AS,LINC02344 |
| 13 | 42950158 | 43931931 | GERD | ANX | -0.666592 | 0.00137345 | negative | TNFSF11,LINC02341,FAM216B,EPSTI1,LINC01050,LINC00428,LINC00400,ENOX1,DNAJC15 |
| 13 | 59751796 | 61268155 | GERD | ANX | 0.680062 | 3.68904E-05 | positive | DIAPH3-AS2,LINC00434,TDRD3,LINC00378,DIAPH3,DIAPH3-AS1 |
| 13 | 66382287 | 67718879 | GERD | ANX | 0.444207 | 0.00599675 | positive | PCDH9,PCDH9-AS2,LINC01052,PCDH9-AS4,MIR548X2,MIR4704,PCDH9-AS3 |
| 13 | 67718880 | 69004702 | GERD | ANX | 1 | 0.000430819 | positive | PCDH9,LINC00364 |
| 13 | 91764205 | 93285142 | GERD | ANX | 0.622344 | 0.0056513 | positive | LINC00379,GPC5,GPC5-AS2,MIR17HG,MIR19A,MIR18A,MIR17,MIR548AS,MIR19B1,MIR92A1,MIR20A |
| 13 | 107037865 | 108521978 | GERD | ANX | 0.516198 | 0.00593242 | positive | EFNB2,SNORD31B,LINC00551,ARGLU1,FAM155A,MIR1267,LINC00443,FAM155A-IT1 |
| 13 | 110995433 | 111621244 | GERD | ANX | 0.567276 | 0.00158899 | positive | RAB20,COL4A2-AS1,COL4A2,LINC00567,ING1,ANKRD10,PRECSIT,COL4A2-AS2,CARS2,NAXD,LOC105370362,LINC00431 |
| 14 | 29029225 | 30831154 | GERD | ANX | 0.486102 | 0.0011244 | positive | LINC01551,LINC02282,PRKD1,FOXG1-AS1,FOXG1 |
| 14 | 41614834 | 42562550 | GERD | ANX | 0.587353 | 0.00226694 | positive | LRFN5 |
| 14 | 46748239 | 48177483 | GERD | ANX | 0.781622 | 0.000155663 | positive | MDGA2,LINC00871,RPL10L |
| 14 | 74245429 | 75699501 | GERD | ANX | 0.597974 | 0.00193854 | positive | ENTPD5,YLPM1,SYNDIG1L,FCF1,COQ6,ISCA2,LIN52,LTBP2,MLH3,ZC2HC1C,BBOF1,PTGR2,LINC02274,EIF2B2,NEK9,LOC100506476,ZNF410,DLST,PROX2,ALDH6A1,ACYP1,ABCD4,AREL1,PGF,NPC2,FAM161B,TMED10,MIR4709,VSX2,RPS6KL1,MIDEAS,VRTN |
| 15 | 26392948 | 27563172 | GERD | ANX | 0.477337 | 0.00444854 | positive | GABRG3,GABRA5,GABRG3-AS1,GABRB3 |
| 15 | 36583328 | 37962915 | GERD | ANX | 0.522016 | 0.000858552 | positive | CDIN1,MEIS2,LOC145845,CSNK1A1P1,MIR8063 |
| 15 | 40604781 | 42332685 | GERD | ANX | 0.633264 | 0.000131806 | positive | VPS18,GCHFR,PPP1R14D,TYRO3,RPAP1,CHST14,PHGR1,NUSAP1,INO80,DNAJC17,JMJD7,RHOV,MRPL42P5,DISP2,EXD1,RMDN3,CHP1,C15orf62,OIP5,RPUSD2,CCDC32,DLL4,EHD4,IVD,KNL1,BAHD1,LOC105370943,OIP5-AS1,ITPKA,RAD51,CCDC9B,EHD4-AS1,MIR4310,NDUFAF1,SPINT1-AS1,INAFM2,RTF1,RAD51-AS1,MGA,MAPKBP1,PLA2G4B,MIR626,SPTBN5,PLA2G4E,KNSTRN,JMJD7-PLA2G4B,ZFYVE19,SPINT1,LOC105370792,CHAC1,LTK,PLA2G4E-AS1 |
| 15 | 63218979 | 64339945 | GERD | ANX | 0.961386 | 3.69772E-05 | positive | RAB8B,RPS27L,FBXL22,TPM1-AS,TPM1,HERC1,LOC101928988,DAPK2,USP3-AS1,APH1B,MIR422A,LACTB,CA12,LINC02568,USP3 |
| 15 | 67396521 | 69089815 | GERD | ANX | 0.646096 | 0.00748028 | positive | RNU6-9,MAP2K5,FEM1B,CORO2B,AAGAB,ANP32A,CLN6,C15orf61,CALML4,RNU6-1,SKOR1,SMAD3,IQCH-AS1,IQCH,LOC101929076,RNU6-7,ITGA11,RNU6-2,RNU6-8,PIAS1 |
| 15 | 69089816 | 70767983 | GERD | ANX | 0.458898 | 0.000316414 | positive | EWSAT1,TLE3,PAQR5,PCAT29,GLCE,RPLP1,LOC145694,NOX5,KIF23,ANP32A-IT1,MIR4312,LINC00593,ANP32A,DRAIC,MIR629,SPESP1 |
| 15 | 89385188 | 90632718 | GERD | ANX | 0.497646 | 0.00426088 | positive | MFGE8,MIR6766,ZNF710-AS1,ACAN,ANPEP,LINC00928,IDH2,FANCI,ARPIN,ARPIN-AP3S2,RLBP1,MIR9-3,MIR9-3HG,PLIN1,PEX11A,ZNF710,MIR3174,HAPLN3,WDR93,ABHD2,MESP2,AP3S2,POLG,TICRR,MIR5094,MIR5009,RHCG,MESP1,KIF7,LOC105371031 |
| 15 | 96864279 | 98025684 | GERD | ANX | 0.464515 | 0.00191504 | positive | NR2F2-AS1,NR2F2,LINC02254,SPATA8-AS1,MIR1469,SPATA8 |
| 15 | 98025685 | 98828261 | GERD | ANX | 0.543706 | 0.00332316 | positive | LOC101927310,LINC02254,LINC00923,LINC02251,LINC01582,ARRDC4 |
| 16 | 60169 | 1049079 | GERD | ANX | 0.587753 | 0.00360737 | positive | CEROX1,WFIKKN1,RPUSD1,LOC100287175,LUC7L,PGAP6,DDX11L10,WDR24,HAGHL,JMJD8,ARHGDIG,POLR3K,MIR662,LINC02867,NME4,SNRNP25,NHLRC4,CHTF18,MIR3176,MIR6859-4,METTL26,ANTKMT,RHOT2,HBM,HBA2,HBA1,STUB1,CIAO3,HBZ,HBQ1,PRR35,RHBDF1,FBXL16,MPG,METRN,PIGQ,CAPN15,LOC105371038,DECR2,RAB11FIP3,RGS11,LINC00235,NPRL3,GNG13,CCDC78,PRR25,MSLN,MIR5587,PDIA2,RAB40C,MIR6859-2,FAM234A,AXIN1,SOX8,MRPL28,LMF1,LMF1-AS1,MCRIP2,WASIR2,RHBDL1,MIR6859-1,LOC100134368,MIR6859-3,WDR90 |
| 16 | 5782969 | 6446081 | GERD | ANX | 0.386516 | 0.00201836 | positive | RBFOX1 |
| 16 | 6446082 | 7051429 | GERD | ANX | 0.55595 | 0.005856 | positive | RBFOX1 |
| 16 | 25235252 | 26154122 | GERD | ANX | 0.410942 | 0.00837396 | positive | HS3ST4,AQP8,MIR548W,ZKSCAN2 |
| 16 | 76635468 | 77270665 | GERD | ANX | 1 | 1.69194E-06 | positive | MON1B,LINC02125,SYCE1L,MIR4719 |
| 16 | 81437862 | 82310204 | GERD | ANX | 0.548866 | 0.00292387 | positive | HSD17B2,MIR7854,CMIP,MPHOSPH6,MIR6504,SDR42E1,LOC100129617,PLCG2 |
| 16 | 83778372 | 84440154 | GERD | ANX | 0.624462 | 6.60795E-05 | positive | HSBP1,ADAD2,OSGIN1,LOC102724163,SLC38A8,ATP2C2,TAF1C,CDH13,WFDC1,KCNG4,LOC654780,DNAAF1,NECAB2,HSDL1,MBTPS1,MLYCD |
| 16 | 89106305 | 90292787 | GERD | ANX | 0.531682 | 0.00509789 | positive | ACSF3,SPATA33,LINC00304,LOC100287036,PRDM7,GAS8,SNORA119,VPS9D1,SPATA2L,VPS9D1-AS1,URAHP,CPNE7,ZNF778,LOC101927817,ANKRD11,LINC02138,MC1R,CDK10,DEF8,CENPBD1,FAM157B,CDH15,GAS8-AS1,CHMP1A,AFG3L1P,TUBB3,SLC22A31,FANCA,SPIRE2,LOC105371414,SNORD68,TCF25,DPEP1,SPG7,ZNF276,DBNDD1,FAM157C,RPL13 |
| 17 | 5573785 | 6305078 | GERD | ANX | 0.667433 | 0.00567747 | positive | LOC339166,WSCD1 |
| 17 | 48722591 | 49909298 | GERD | ANX | 0.70546 | 0.00415308 | positive | LINC02073,ABCC3,TOB1,LINC02071,SPAG9,UTP18,ANKRD40CL,TOB1-AS1,ANKRD40,LUC7L3,WFIKKN2,MIR8059,CA10,NME1,LINC02072,NME1-NME2,MBTD1,NME2 |
| 17 | 60453121 | 62112373 | GERD | ANX | 1 | 0.000531688 | positive | DCAF7,PRR29,ICAM2,PSMC5,ACE,SMARCD2,DDX42,TANC2,CCDC47,SCN4A,LIMD2,KCNH6,CSHL1,TCAM1P,CYB561,METTL2A,MAP3K3,STRADA,LOC105371855,MIR633,MARCHF10,LOC729683,TACO1,FTSJ3,EFCAB3,MRC2,CSH2,CD79B,GH2,MARCHF10-DT,GH1,CSH1,TLK2,PRR29-AS1 |
| 17 | 64438388 | 65582327 | GERD | ANX | 0.645662 | 0.00772165 | positive | PRKCA,PSMD12,CACNG5,PITPNC1,CACNG4,HELZ,LOC101928021,MIR548AA2,CACNG1,MIR548D2,MIR634 |
| 17 | 79333775 | 80459149 | GERD | ANX | 0.509876 | 0.00954273 | positive | RAC3,MRPL12,FAAP100,BAHCC1,LRRC45,RFNG,HGS,LOC100130370,CCDC57,MCRIP1,TEX19,MIR3186,DUS1L,MIR6786,NOTUM,MAFG,ANAPC11,ARHGDIA,CENPX,SNORD134,PPP1R27,P4HB,MAFG-DT,ACTG1,SIRT7,SECTM1,CD7,HEXD,DCXR,UTS2R,MIR4740,NARF,MYADML2,LINC01970,PCYT2,TSPAN10,OGFOD3,ARL16,NPLOC4,CSNK1D,GCGR,PDE6G,OXLD1,NPB,PYCR1,FSCN2,FASN,DCXR-DT,CCDC137,SLC16A3,ASPSCR1,ALYREF,MIR6787,SLC25A10,CYBC1,GPS1 |
| 18 | 49790997 | 50372823 | GERD | ANX | 0.721704 | 0.00690518 | positive | DCC |
| 18 | 52512524 | 53762996 | GERD | ANX | 0.41001 | 0.0022984 | positive | LINC01539,TCF4,CCDC68,LINC01905,TCF4-AS1,RAB27B,LINC01415,MIR4529,LINC01929,LOC642484,LINC01416 |
| 19 | 1515921 | 2253174 | GERD | ANX | 0.639289 | 0.000793495 | positive | MEX3D,JSRP1,ABHD17A,MBD3,SCAMP4,ATP8B3,AP3D1,BTBD2,PLEKHJ1,MIR1909,TCF3,MIR4321,CSNK1G2-AS1,PLK5,ONECUT3,LOC100288123,MIR6789,DOT1L,UQCR11,MOB3A,MKNK2,MIR1227,AMH,KLF16,IZUMO4,ADAT3,SF3A2,CSNK1G2,REXO1 |
| 19 | 11681979 | 13213186 | GERD | ANX | 0.601832 | 0.00553772 | positive | PRDX2,MIR6794,DAND5,FARSA,MAN2B1,KLF1,RNASEH2A,ZNF491,FBXW9,SYCE2,NFIX,HOOK2,LYL1,MAST1,CALR,MIR5695,ZNF136,MIR6515,JUNB,ACP5,ZNF563,BEST2,ZNF627,SNORD135,WDR83OS,ZNF564,ZNF439,ZNF442,FARSA-AS1,ZNF799,ZNF44,ZNF709,ZNF791,RTBDN,ZNF490,ZNF823,ZNF433-AS1,ZNF844,ZNF20,DHPS,ZNF763,ZNF443,RAD23A,WDR83,GCDH,ZNF625-ZNF20,MIR5684,LOC100289333,ZNF878,HNRNPA1P10,ZNF440,ZNF625,TRIR,ZNF441,ZNF788P,GET3,SNORD41,DNASE2,GADD45GIP1,ZNF833P,ZNF700,ZNF69,TNPO2,ZNF433,GNG14 |
| 19 | 17750519 | 18504867 | GERD | ANX | 0.654371 | 1.63593E-05 | positive | RAB3A,IQCN,ARRDC2,CCDC124,LSM4,FCHO1,IL12RB1,JAK3,MAP1S,RPL18A,PIK3R2,IFI30,LOC102725254,UNC13A,LOC729966,MPV17L2,JUND,MIR3188,SLC5A5,SNORA68,PDE4C,GDF15,INSL3,B3GNT3,MAST3,MIR3189,LRRC25,PGPEP1,KCNN1 |
| 19 | 31763009 | 33028099 | GERD | ANX | 0.513573 | 0.000889653 | positive | ZNF507,DPY19L3,LINC01782,LOC400684,TSHZ3,LINC01533,THEG5 |
| 19 | 34633275 | 35649343 | GERD | ANX | 0.680964 | 0.00440283 | positive | HPN,PDCD2L,ZNF30-AS1,ZNF807P,SCGB1B2P,SCGB2B3P,ZNF30,LINC00904,HPN-AS1,MIR6887,LINC01838,GARRE1,LINC01801,FXYD1,ZNF181,SCN1B,LGI4,FXYD5,FXYD3,LSM14A,UBA2,GPI,SCGB2B2,ZNF792,ZNF302,FXYD7,ZNF599,GRAMD1A,WTIP |
| 20 | 9279551 | 10279603 | GERD | ANX | 0.77054 | 0.000104341 | positive | SNAP25,LAMP5-AS1,PLCB4,ANKEF1,SNAP25-AS1,PARAL1,LAMP5,PAK5 |
| 20 | 16661517 | 17359290 | GERD | ANX | 0.759863 | 0.000235148 | positive | SNRPB2,PCSK2,OTOR |
| 20 | 19957929 | 21682377 | GERD | ANX | 0.669054 | 9.51981E-05 | positive | NAA20,KIZ-AS1,CRNKL1,LINC01727,NKX2-2,LINC01726,CFAP61,KIZ,XRN2,LINC00237,INSM1,RALGAPA2,RIN2,NKX2-4 |
| 20 | 41351515 | 42185671 | GERD | ANX | 0.572204 | 0.00157395 | positive | L3MBTL1,SRSF6,PTPRT-AS1,PTPRT |
| 20 | 44072211 | 45673603 | GERD | ANX | 0.515055 | 0.000477906 | positive | TP53RK,ZNF335,WFDC8,SLC12A5,ZSWIM1,WFDC6,WFDC13,SPATA25,ZNF663P,EPPIN-WFDC6,WFDC2,PCIF1,MKRN7P,MMP9,UBE2C,CD40,CTSA,ACOT8,TNNC2,SLC13A3,NCOA5,ZNF334,WFDC11,CDH22,OCSTAMP,LOC105372631,SLC12A5-AS1,WFDC9,MIR3617,EPPIN,WFDC10B,SPINT4,WFDC3,SPINT3,SLC2A10,DNTTIP1,ZSWIM3,PLTP,WFDC10A,ELMO2,SLC35C2,SNX21,EYA2,NEURL2,LOC105372633 |
| 20 | 51533751 | 52411532 | GERD | ANX | 0.698033 | 0.000021904 | positive | LOC105372672,ZNF217,TSHZ2,LOC101927770 |
| 20 | 58780895 | 59560457 | GERD | ANX | 0.644434 | 0.000362853 | positive | LOC101928048,MIR4533,MIR646HG,MIR548AG2,MIR646 |
| 21 | 16873444 | 18222037 | GERD | ANX | 0.707507 | 0.00382137 | positive | USP25,SNORD74B,MIR99AHG,MIR99A,MIR125B2,MIRLET7C |
| 21 | 27774351 | 28616539 | GERD | ANX | 0.688589 | 0.00435463 | positive | CYYR1-AS1,ADAMTS5,ADAMTS1,CYYR1,MIR4759 |
| 21 | 35729262 | 37303548 | GERD | ANX | 0.466631 | 0.00198181 | positive | LINC01426,PPP1R2P2,RCAN1,RUNX1,SMIM34A,SMIM11B,FAM243B,KCNE2,MIR802,LINC00160,KCNE1,FAM243A,RUNX1-IT1,CLIC6,LOC100506403,KCNE1B,SMIM11A |
| 21 | 39252060 | 40480831 | GERD | ANX | 0.610481 | 7.59858E-05 | positive | KCNJ6,LINC01700,ERG,LINC01423,LOC101928398,KCNJ15,ETS2,DSCR10,DSCR4,LOC400867,DSCR8,LINC00114 |
| 22 | 38718590 | 40378783 | GERD | ANX | 0.828253 | 0.00283763 | positive | TPTEP2-CSNK1E,SNORD43,LOC100506472,RPL3,APOBEC3A_B,SUN2,APOBEC3A,FAM227A,APOBEC3C,APOBEC3B,MGAT3,DMC1,SYNGR1,JOSD1,DDX17,CBX7,TAB1,TPTEP2,CACNA1I,GTPBP1,NPTXR,TOMM22,APOBEC3D,SNORD83A,PDGFB,APOBEC3B-AS1,ATF4,APOBEC3G,APOBEC3H,RPS19BP1,LOC105373031,KDELR3,APOBEC3F,DNAL4,ENTHD1,GRAP2,KCNJ4,CBY1,CBX6,SNORD83B,SNORD139,MGAT3-AS1,MIEF1 |
| 22 | 40378784 | 41722787 | GERD | ANX | 0.929466 | 6.87798E-05 | positive | RANGAP1,MRTFA,EP300,L3MBTL2-AS1,ST13,SLC25A17,SGSM3,MRTFA-AS1,RBX1,TNRC6B,LOC114841040,XPNPEP3,SNORD140,MIR1281,MCHR1,MIR6889,CHADL,L3MBTL2,DNAJB7,MIR4766,EP300-AS1,FAM83F,ZC3H7B,ADSL |
| 22 | 41722788 | 42867927 | GERD | ANX | 0.721432 | 0.000263767 | positive | TOB2,DESI1,LINC00634,SNU13,MIR378I,NFAM1,CCDC134,CYP2D7,LINC01315,MIR33A,NAGA,LOC101929829,PHETA2,CYP2D6,WBP2NL,C22orf46,SMDT1,CSDC2,OGFRP1,SREBF2-AS1,PHF5A,PMM1,XRCC6,SHISA8,SREBF2,NDUFA6,TCF20,SEPTIN3,ACO2,NDUFA6-DT,LOC105373044,TNFRSF13C,ZC3H7B,MEI1,POLR3H,TEF,CENPM |
| 22 | 46351049 | 47299625 | GERD | ANX | 0.903992 | 0.00465101 | positive | TTC38,CDPF1,GRAMD4,PPARA,CELSR1,TBC1D22A,TRMU,MIRLET7A3,LOC642648,CERK,LOC730668,PKDREJ,PRR34-AS1,GTSE1,WNT7B,MIRLET7B,PRR34,MIR4763,MIR3619,MIRLET7BHG,LINC00899,GTSE1-DT |
| 4 | 22835186 | 23858350 | GERD | AN | 0.885375 | 0.000353394 | positive | PPARGC1A |
| 5 | 1206610 | 2106885 | GERD | AN | 0.751965 | 7.46718E-06 | positive | NDUFS6,IRX4,LINC01511,MIR4457,TERT,LPCAT1,MIR6075,MIR4277,SLC6A18,CLPTM1L,SDHAP3,LINC02116,LOC728613,CTD-2194D22.4,MRPL36,SLC6A19,SLC6A3 |
| 5 | 79005159 | 80448225 | GERD | AN | -0.922653 | 1.38119E-05 | negative | CRSP8P,MSH3,ANKRD34B,THBS4,LINC01337,DHFR,RASGRF2-AS1,ZFYVE16,CMYA5,MTX3,MTRNR2L2,RASGRF2,LINC01455,FAM151B,SPZ1,THBS4-AS1,LOC644936,SERINC5 |
| 9 | 14244748 | 14902924 | GERD | AN | -0.77512 | 5.77132E-05 | negative | ZDHHC21,FREM1,NFIB,CER1 |
| 16 | 58508979 | 59885499 | GERD | AN | 0.854313 | 0.000331069 | positive | SLC38A7,SNORA46,SNORA50A,APOOP5,NDRG4,SETD6,GOT2,CNOT1 |
| 18 | 52512524 | 53762996 | GERD | AN | 0.892002 | 4.00337E-05 | positive | LINC01539,TCF4,CCDC68,LINC01905,TCF4-AS1,RAB27B,LINC01415,MIR4529,LINC01929,LOC642484,LINC01416 |
| 6 | 33194976 | 33864262 | GERD | ADHD | 1 | 7.07512E-06 | positive | TAPBP,PFDN6,VPS52,MIR1234,DAXX,SMIM40,LINC00336,WDR46,B3GALT4,ZBTB9,BAK1,CUTA,MIR6873,LINC01016,KIFC1,MIR5004,PHF1,SYNGAP1,IP6K3,MIR6834,RGL2,RPS18,MIR3934,UQCC2,LEMD2,ITPR3,GGNBP1,ZBTB22,LOC101929188,HCG25,MLN |
| 6 | 27261036 | 28666364 | GERD | ANX | 1 | 2.98499E-10 | positive | H2AC13,ZSCAN31,LOC100131289,H3C12,H2BC13,ZKSCAN8,ZKSCAN4,ZSCAN16,POM121L2,GPX6,H2AC15,ZSCAN16-AS1,OR2B6,VN1R10P,ZNF184,ZSCAN26,H2BC14,H1-5,ZSCAN12P1,OR1F12,H2AC17,H3C11,ZNF391,LINC01012,H2AC14,ZNF165,ZBED9,H4C12,OR2B2,LINC00533,ZSCAN9,H3C10,H2AC16,ZNF192P1,NKAPL,GPX5,H2BC15,PGBD1,ZNF204P,TOB2P1,ZKSCAN3,H4C11,ZSCAN12,H2BC17,H4C13,ZSCAN23 |
| 6 | 27261036 | 28666364 | GERD | BIP | 0.95478 | 1.84372E-09 | positive | H2AC13,ZSCAN31,LOC100131289,H3C12,H2BC13,ZKSCAN8,ZKSCAN4,ZSCAN16,POM121L2,GPX6,H2AC15,ZSCAN16-AS1,OR2B6,VN1R10P,ZNF184,ZSCAN26,H2BC14,H1-5,ZSCAN12P1,OR1F12,H2AC17,H3C11,ZNF391,LINC01012,H2AC14,ZNF165,ZBED9,H4C12,OR2B2,LINC00533,ZSCAN9,H3C10,H2AC16,ZNF192P1,NKAPL,GPX5,H2BC15,PGBD1,ZNF204P,TOB2P1,ZKSCAN3,H4C11,ZSCAN12,H2BC17,H4C13,ZSCAN23 |
| 6 | 27261036 | 28666364 | GERD | MDD | 0.950073 | 1.85665E-13 | positive | H2AC13,ZSCAN31,LOC100131289,H3C12,H2BC13,ZKSCAN8,ZKSCAN4,ZSCAN16,POM121L2,GPX6,H2AC15,ZSCAN16-AS1,OR2B6,VN1R10P,ZNF184,ZSCAN26,H2BC14,H1-5,ZSCAN12P1,OR1F12,H2AC17,H3C11,ZNF391,LINC01012,H2AC14,ZNF165,ZBED9,H4C12,OR2B2,LINC00533,ZSCAN9,H3C10,H2AC16,ZNF192P1,NKAPL,GPX5,H2BC15,PGBD1,ZNF204P,TOB2P1,ZKSCAN3,H4C11,ZSCAN12,H2BC17,H4C13,ZSCAN23 |
| 6 | 26396201 | 27261035 | GERD | OCD | 1 | 0.000382836 | positive | H2BC12,LOC100270746,HCG11,MIR3143,BTN1A1,BTN2A1,H2AC12,PRSS16,BTN3A1,ABT1,GUSBP2,BTN2A3P,BTN3A3,LOC285819,H2BC11,H2AC11,LOC105374988,H4C9,ZNF322,HMGN4,LINC00240 |
| 6 | 27261036 | 28666364 | GERD | PTSD | 1 | 1.51483E-10 | positive | H2AC13,ZSCAN31,LOC100131289,H3C12,H2BC13,ZKSCAN8,ZKSCAN4,ZSCAN16,POM121L2,GPX6,H2AC15,ZSCAN16-AS1,OR2B6,VN1R10P,ZNF184,ZSCAN26,H2BC14,H1-5,ZSCAN12P1,OR1F12,H2AC17,H3C11,ZNF391,LINC01012,H2AC14,ZNF165,ZBED9,H4C12,OR2B2,LINC00533,ZSCAN9,H3C10,H2AC16,ZNF192P1,NKAPL,GPX5,H2BC15,PGBD1,ZNF204P,TOB2P1,ZKSCAN3,H4C11,ZSCAN12,H2BC17,H4C13,ZSCAN23 |
| 6 | 27261036 | 28666364 | GERD | SCZ | 0.748267 | 9.71863E-11 | positive | H2AC13,ZSCAN31,LOC100131289,H3C12,H2BC13,ZKSCAN8,ZKSCAN4,ZSCAN16,POM121L2,GPX6,H2AC15,ZSCAN16-AS1,OR2B6,VN1R10P,ZNF184,ZSCAN26,H2BC14,H1-5,ZSCAN12P1,OR1F12,H2AC17,H3C11,ZNF391,LINC01012,H2AC14,ZNF165,ZBED9,H4C12,OR2B2,LINC00533,ZSCAN9,H3C10,H2AC16,ZNF192P1,NKAPL,GPX5,H2BC15,PGBD1,ZNF204P,TOB2P1,ZKSCAN3,H4C11,ZSCAN12,H2BC17,H4C13,ZSCAN23 |

Note:

chr: chromosome;

start: start position of the genomic locus;

stop: end position of the genomic locus;

phen1/phen2: trait pair analyzed;

rho: local genetic correlation estimate;

*p*: *P*-value for local genetic correlation;

direction: direction of correlation (positive or negative);

genes: genes located within the locus.

**Table S7.** Novel pleiotropic loci identified through conditional FDR analysis between GERD and psychiatric disorders.

| **Trait1** | **Trait2** | **SNP** | **CHR** | **BP** | **A1** | **A2** | **condfdr** | **conjfdr** | **beta1** | **beta2** | **pval1** | **pval2** | **annotation** | **Gene** | **Function** | **Exonic Function** | **CADD** | **cytoBand** |
| --- | --- | --- | --- | --- | --- | --- | --- | --- | --- | --- | --- | --- | --- | --- | --- | --- | --- | --- |
| GERD | ADHD | rs56227503 | 1 | 8610837 | T | C | 0.012707314 | 0.045637572 | 0.0228 | -0.044599955 | 1.331E-07 | 0.0002234 | new | RERE | intronic | . | . | 1p36.23 |
| GERD | ADHD | rs3766823 | 1 | 32197257 | A | G | 0.019708824 | 0.027584504 | 0.0229 | -0.049200748 | 0.000001 | 0.00004975 | new | ADGRB2 | intronic | . | . | 1p35.2 |
| GERD | ADHD | rs4656024 | 1 | 88724815 | T | G | 0.045278973 | 0.045278973 | -0.0146 | -0.039697627 | 0.00003291 | 0.00002539 | new | PKN2-AS1 | intergenic | . | . | 1p22.2 |
| GERD | ADHD | rs10737305 | 1 | 174923045 | C | G | 0.002381243 | 0.03929106 | 0.0228 | 0.036196915 | 4.22E-11 | 0.000148 | known | RABGAP1L-AS1 | ncRNA_intronic | . | . | 1q25.1 |
| GERD | ADHD | rs861585 | 1 | 184737925 | A | G | 0.026357237 | 0.042965356 | 0.0177 | -0.03509882 | 0.000002503 | 0.0001754 | new | EDEM3 | intergenic | . | . | 1q25.3 |
| GERD | ADHD | rs57537843 | 2 | 22558973 | A | G | 0.029672168 | 0.040616574 | 0.0154 | -0.03630098 | 0.00000447 | 0.0001356 | new | LINC01884 | intergenic | . | . | 2p24.1 |
| GERD | ADHD | rs34439217 | 2 | 36656809 | A | T | 0.036591073 | 0.036591073 | -0.0216 | 0.055103526 | 0.00001541 | 0.00004527 | new | CRIM1 | intronic | . | . | 2p22.2 |
| GERD | ADHD | rs11126769 | 2 | 80713142 | T | C | 0.017221167 | 0.041798488 | 0.018 | 0.039903173 | 5.331E-07 | 0.0001752 | new | CTNNA2 | intronic | . | . | 2p12 |
| GERD | ADHD | rs7607172 | 2 | 100492135 | T | C | 0.000726202 | 0.022111371 | -0.025 | -0.039395929 | 2.296E-12 | 0.0000272 | known | AFF3 | intronic | . | . | 2q11.2 |
| GERD | ADHD | rs3754757 | 2 | 173835127 | A | G | 0.016295183 | 0.039844167 | -0.0183 | 0.037295785 | 4.789E-07 | 0.0001538 | new | RAPGEF4 | intronic | . | . | 2q31.1 |
| GERD | ADHD | rs6704768 | 2 | 233592501 | A | G | 0.008173104 | 0.015409678 | 0.0194 | -0.041395075 | 2.791E-08 | 0.000009165 | known | GIGYF2 | intronic | . | . | 2q37.1 |
| GERD | ADHD | rs56403804 | 3 | 43368021 | A | G | 0.019638558 | 0.019638558 | 0.0253 | -0.066898637 | 9.874E-07 | 9.971E-07 | known | SNRK | intronic | . | . | 3p22.1 |
| GERD | ADHD | rs73094902 | 3 | 71603361 | T | C | 0.028648753 | 0.028648753 | -0.016 | 0.04449524 | 0.000004633 | 0.000004232 | new | FOXP1 | intronic | . | . | 3p13 |
| GERD | ADHD | rs7426688 | 3 | 77240830 | T | C | 0.036745547 | 0.036745547 | -0.0184 | 0.050598053 | 0.0000157 | 0.00000357 | new | ROBO2 | intronic | . | . | 3p12.3 |
| GERD | ADHD | rs73163783 | 3 | 117602144 | T | C | 0.039251289 | 0.039251289 | -0.0169 | -0.047301273 | 0.00001945 | 0.0000795 | new | LINC02024 | intergenic | . | . | 3q13.32 |
| GERD | ADHD | rs7689432 | 4 | 3000317 | A | T | 0.0170924 | 0.03361562 | -0.018 | 0.043796765 | 5.874E-07 | 0.00009296 | new | GRK4 | intronic | . | . | 4p16.3 |
| GERD | ADHD | rs28645467 | 4 | 80905740 | T | C | 0.013153849 | 0.023755169 | 0.0172 | 0.038797547 | 2.473E-07 | 0.00003233 | new | ANTXR2 | intronic | . | . | 4q21.21 |
| GERD | ADHD | rs72678864 | 4 | 112422145 | A | G | 0.031101057 | 0.031101057 | -0.02 | 0.084901653 | 0.000007106 | 5.35E-10 | new* | MIR297 | intergenic | . | . | 4q25 |
| GERD | ADHD | rs28676930 | 4 | 176758546 | A | G | 0.039642047 | 0.039642047 | -0.0162 | 0.038999535 | 0.0000194 | 0.00008659 | new | GPM6A | intronic | . | . | 4q34.2 |
| GERD | ADHD | rs10941692 | 5 | 45099778 | T | C | 0.00300974 | 0.042639491 | 0.0283 | 0.045298361 | 1.878E-10 | 0.0001853 | known | HCN1 | intergenic | . | . | 5p12 |
| GERD | ADHD | rs288151 | 5 | 107377338 | T | C | 0.01976945 | 0.01976945 | -0.0222 | 0.050797672 | 0.000001011 | 0.00001978 | new | FBXL17 | intronic | . | . | 5q21.3 |
| GERD | ADHD | rs10947428 | 6 | 33647058 | T | C | 0.003796592 | 0.041381698 | -0.027 | -0.043001451 | 4.857E-10 | 0.0001704 | known | ITPR3 | intronic | . | . | 6p21.31 |
| GERD | ADHD | rs9489931 | 6 | 98303699 | A | G | 0.042551495 | 0.042551495 | -0.0146 | -0.039198313 | 0.00002681 | 0.00002951 | new | LOC101927314 | intergenic | . | . | 6q16.1 |
| GERD | ADHD | rs7807014 | 7 | 1939021 | T | C | 0.000702028 | 0.015410029 | 0.0316 | 0.053701891 | 2.131E-12 | 0.000009169 | known | MAD1L1 | intronic | . | . | 7p22.3 |
| GERD | ADHD | rs73035954 | 7 | 3563804 | T | C | 0.006232314 | 0.039528811 | -0.0233 | 0.041899813 | 1.007E-08 | 0.0001506 | known | SDK1 | intronic | . | . | 7p22.2 |
| GERD | ADHD | rs2158220 | 7 | 82410969 | T | C | 0.01294835 | 0.030744392 | -0.0185 | -0.037203549 | 2.308E-07 | 0.00007356 | new | PCLO | intronic | . | . | 7q21.11 |
| GERD | ADHD | rs8180817 | 7 | 114047542 | C | G | 0.00588593 | 0.00588593 | -0.0202 | 0.059598213 | 5.658E-09 | 4.681E-10 | known | NONE | intergenic | . | . | 7q31.1 |
| GERD | ADHD | rs11774014 | 8 | 12698174 | A | C | 0.005912665 | 0.038856304 | -0.0349 | -0.058996573 | 6.484E-09 | 0.0001427 | known | LINC00681 | intergenic | . | . | 8p23.1 |
| GERD | ADHD | rs10818077 | 9 | 120505417 | C | G | 0.008054917 | 0.030713919 | 0.0195 | 0.036997089 | 2.64E-08 | 0.00007328 | known | TLR4 | intergenic | . | . | 9q33.1 |
| GERD | ADHD | rs11596214 | 10 | 106453832 | A | G | 0.006116967 | 0.006116967 | -0.0208 | 0.052801157 | 9.627E-09 | 3.167E-08 | known | SORCS3 | intronic | . | . | 10q25.1 |
| GERD | ADHD | rs7092307 | 10 | 134956451 | T | C | 0.005776928 | 0.027864175 | -0.0241 | -0.04780465 | 3.427E-09 | 0.00005143 | known | ADGRA1 | intergenic | . | . | 10q26.3 |
| GERD | ADHD | rs981895 | 11 | 24594148 | T | C | 0.042494753 | 0.042494753 | 0.0143 | 0.038403071 | 0.00002587 | 0.00008243 | new | LUZP2 | intronic | . | . | 11p14.3 |
| GERD | ADHD | rs12419325 | 11 | 28580338 | T | G | 0.036986163 | 0.036986163 | 0.0198 | 0.072199724 | 0.00001606 | 5.525E-09 | new* | MIR8068 | intergenic | . | . | 11p14.1 |
| GERD | ADHD | rs1943354 | 11 | 89921664 | T | G | 0.025520692 | 0.030798124 | 0.0167 | -0.037701867 | 0.000002735 | 0.00006254 | new | NAALAD2 | intronic | . | . | 11q14.3 |
| GERD | ADHD | rs10789931 | 11 | 112842773 | T | C | 0.006026068 | 0.043242708 | 0.0299 | -0.052905119 | 7.989E-09 | 0.0001929 | known | NCAM1 | intronic | . | . | 11q23.2 |
| GERD | ADHD | rs7933981 | 11 | 113438068 | A | G | 0.046277638 | 0.046277638 | -0.0138 | 0.036100466 | 0.0000306 | 0.000146 | new | DRD2 | intergenic | . | . | 11q23.2 |
| GERD | ADHD | rs11223560 | 11 | 133561399 | A | G | 0.035443517 | 0.035443517 | -0.0159 | 0.040901991 | 0.0000133 | 0.0000219 | new | LINC02743 | intergenic | . | . | 11q25 |
| GERD | ADHD | rs10846378 | 12 | 16743434 | T | G | 0.025546167 | 0.030193322 | 0.0168 | 0.039201482 | 0.000002746 | 0.00005755 | new | LMO3 | intronic | . | . | 12p12.3 |
| GERD | ADHD | rs773110 | 12 | 56375137 | C | G | 0.004807984 | 0.018948142 | 0.0221 | 0.043595744 | 1.096E-09 | 0.00001886 | known | RAB5B | intronic | . | . | 12q13.2 |
| GERD | ADHD | rs9537777 | 13 | 58261917 | A | G | 0.015653748 | 0.025207304 | 0.022 | 0.044896881 | 4.885E-07 | 0.00003763 | new | PCDH17 | intronic | . | . | 13q21.1 |
| GERD | ADHD | rs34500631 | 14 | 46777783 | A | C | 0.0246198 | 0.0246198 | 0.0203 | 0.057900916 | 0.000002373 | 0.00001547 | new | LINC00871 | ncRNA_intronic | . | . | 14q21.2 |
| GERD | ADHD | rs4906269 | 14 | 103362017 | A | C | 0.005810606 | 0.015199138 | -0.0276 | -0.056697344 | 3.994E-09 | 0.000005734 | known | TRAF3 | intronic | . | . | 14q32.32 |
| GERD | ADHD | rs2733330 | 15 | 57551789 | A | G | 0.001832097 | 0.036428437 | 0.0276 | 0.044600447 | 1.538E-11 | 0.0001164 | known | TCF12 | intronic | . | . | 15q21.3 |
| GERD | ADHD | rs4966009 | 15 | 99203496 | T | C | 0.024955402 | 0.038573217 | 0.0162 | -0.036498023 | 0.000002222 | 0.000131 | new | IGF1R | intronic | . | . | 15q26.3 |
| GERD | ADHD | rs9941176 | 16 | 963843 | A | G | 0.046591544 | 0.046591544 | -0.0196 | -0.067797158 | 0.00003632 | 0.000004159 | new | LMF1 | intronic | . | . | 16p13.3 |
| GERD | ADHD | rs1162202 | 16 | 61966703 | T | C | 0.029943736 | 0.029943736 | -0.018 | 0.061396089 | 0.000005852 | 1.922E-09 | new* | CDH8 | intronic | . | . | 16q21 |
| GERD | ADHD | rs35322724 | 16 | 77137324 | A | C | 0.014120384 | 0.029477125 | 0.0174 | -0.037504587 | 3.304E-07 | 0.00006277 | new | MON1B | intergenic | . | . | 16q23.1 |
| GERD | ADHD | rs2447091 | 17 | 2296014 | T | C | 0.025818119 | 0.043253976 | -0.0174 | -0.03740077 | 0.000002281 | 0.0001825 | new | MNT | intronic | . | . | 17p13.3 |
| GERD | ADHD | rs62098049 | 18 | 50904666 | A | G | 0.007676219 | 0.029568227 | 0.0202 | -0.037203549 | 2.203E-08 | 0.00006349 | known | DCC | intronic | . | . | 18q21.2 |
| GERD | ADHD | rs11081564 | 18 | 77551586 | A | G | 0.033284849 | 0.033284849 | -0.0179 | -0.049599992 | 0.00001008 | 0.000004276 | new | CTDP1 | intergenic | . | . | 18q23 |
| GERD | ADHD | rs8105094 | 19 | 19374061 | T | C | 0.002990785 | 0.025102494 | 0.0307 | -0.048497207 | 1.189E-10 | 0.00003722 | known | HAPLN4 | upstream | . | . | 19p13.11 |
| GERD | ADHD | rs6044120 | 20 | 16545951 | T | C | 0.04437133 | 0.044406529 | -0.0149 | -0.035896644 | 0.00002658 | 0.0001461 | new | KIF16B | intronic | . | . | 20p12.1 |
| GERD | ADHD | rs4599194 | 20 | 21017279 | A | G | 0.035102278 | 0.035102278 | 0.0204 | 0.061104507 | 0.00001273 | 3.609E-08 | new* | LINC00237 | intergenic | . | . | 20p11.23 |
| GERD | ADHD | rs6017596 | 20 | 35868219 | T | C | 0.042518718 | 0.044258841 | 0.0148 | -0.03630098 | 0.00002208 | 0.0001469 | new | RPN2 | intronic | . | . | 20q11.23 |
| GERD | ADHD | rs2838762 | 21 | 46487831 | A | C | 0.031856799 | 0.031856799 | 0.0153 | 0.043701046 | 0.000008066 | 0.000004758 | new | SSR4P1 | intergenic | . | . | 21q22.3 |
| GERD | ANX | rs632051 | 1 | 5291879 | A | C | 0.048633167 | 0.048633167 | -0.0175 | -0.02499991 | 0.00009662 | 0.00001668 | new | AJAP1 | intergenic | . | . | 1p36.32 |
| GERD | ANX | rs2401137 | 1 | 8693324 | A | T | 0.022634143 | 0.023407573 | -0.0164 | -0.02799832 | 0.000002091 | 0.00005439 | new | RERE | intronic | . | . | 1p36.23 |
| GERD | ANX | rs4147222 | 1 | 37176422 | A | G | 0.016461143 | 0.016461143 | -0.0202 | -0.021795812 | 8.587E-08 | 0.0000199 | new | GRIK3 | intergenic | . | . | 1p34.3 |
| GERD | ANX | rs12039463 | 1 | 41116336 | A | C | 0.043630632 | 0.043630632 | -0.0155 | -0.020202707 | 0.00003696 | 0.00007321 | new | RIMS3 | intronic | . | . | 1p34.2 |
| GERD | ANX | rs2503185 | 1 | 66461401 | A | G | 0.01384832 | 0.04052722 | 0.0219 | -0.01789924 | 7.154E-10 | 0.0001594 | known | PDE4B | intronic | . | . | 1p31.3 |
| GERD | ANX | rs12082973 | 1 | 69254610 | A | G | 0.034223342 | 0.034223342 | -0.0267 | -0.030799472 | 0.0000134 | 0.00006852 | new | DEPDC1-AS1 | intergenic | . | . | 1p31.2 |
| GERD | ANX | rs61765555 | 1 | 73957815 | T | C | 0.017493092 | 0.017493092 | -0.0218 | -0.027196495 | 1.048E-07 | 0.000001997 | new | LINC01360 | intergenic | . | . | 1p31.1 |
| GERD | ANX | rs2209458 | 1 | 76862648 | A | G | 0.040048018 | 0.040048018 | -0.0154 | -0.020702832 | 0.00004075 | 0.00003217 | new | ST6GALNAC3 | intronic | . | . | 1p31.1 |
| GERD | ANX | rs10874123 | 1 | 80831434 | A | G | 0.022045866 | 0.033693903 | 0.0198 | 0.020400488 | 3.842E-07 | 0.0001108 | new | LINC01781 | intergenic | . | . | 1p31.1 |
| GERD | ANX | rs11162017 | 1 | 87902311 | A | G | 0.033905272 | 0.033905272 | -0.0161 | -0.019998647 | 0.00001305 | 0.00006806 | new | LINC01364 | intergenic | . | . | 1p22.3 |
| GERD | ANX | rs1200161 | 1 | 169117533 | A | G | 0.019405314 | 0.019405314 | 0.0188 | -0.021898021 | 0.00000046 | 0.00001765 | new | NME7 | intronic | . | . | 1q24.2 |
| GERD | ANX | rs1670770 | 1 | 191202747 | T | C | 0.019404717 | 0.021545979 | 0.0187 | -0.021795812 | 2.323E-07 | 0.00004638 | new | LINC01680 | intergenic | . | . | 1q31.2 |
| GERD | ANX | rs7597000 | 2 | 2315021 | T | G | 0.043678756 | 0.043678756 | -0.0146 | 0.022201708 | 0.00006322 | 0.00001878 | new | MYT1L | intronic | . | . | 2p25.3 |
| GERD | ANX | rs67446571 | 2 | 22554648 | A | G | 0.023599366 | 0.023599366 | -0.0158 | 0.029704423 | 0.000003402 | 8.964E-10 | new* | LINC01884 | intergenic | . | . | 2p24.1 |
| GERD | ANX | rs13016779 | 2 | 48831266 | T | G | 0.047291741 | 0.047291741 | 0.0157 | -0.025902594 | 0.00008751 | 0.00001109 | new | STON1-GTF2A1L | intronic | . | . | 2p16.3 |
| GERD | ANX | rs10188396 | 2 | 60450992 | T | C | 0.020355156 | 0.020355156 | -0.0181 | 0.023403978 | 9.433E-07 | 0.000004393 | new | MIR4432HG | intergenic | . | . | 2p16.1 |
| GERD | ANX | rs6737943 | 2 | 105517422 | C | G | 0.021153946 | 0.021153946 | -0.0255 | 0.03329937 | 0.000001366 | 0.00001329 | new | LINC01159 | intergenic | . | . | 2q12.1 |
| GERD | ANX | rs17269661 | 2 | 109822524 | A | G | 0.016461143 | 0.016461143 | -0.0195 | -0.025102447 | 6.532E-08 | 0.000000739 | new | SH3RF3 | intronic | . | . | 2q12.3 |
| GERD | ANX | rs1186066 | 2 | 125024950 | T | C | 0.042321219 | 0.042321219 | -0.0158 | 0.021996298 | 0.00005533 | 0.000002787 | new | CNTNAP5 | intronic | . | . | 2q14.3 |
| GERD | ANX | rs35125029 | 2 | 144261657 | A | G | 0.019405314 | 0.019405314 | -0.018 | -0.023698607 | 2.911E-07 | 0.000001786 | new | ARHGAP15 | intronic | . | . | 2q22.3 |
| GERD | ANX | rs11678980 | 2 | 162101261 | A | G | 0.028963181 | 0.035441404 | 0.0166 | 0.020096702 | 0.000003229 | 0.0001226 | new | LINC01806 | ncRNA_exonic | . | . | 2q24.2 |
| GERD | ANX | rs1367858 | 2 | 201160771 | T | C | 0.023274436 | 0.028586151 | 0.0176 | 0.01949866 | 0.000001314 | 0.0000803 | new | SPATS2L | intergenic | . | . | 2q33.1 |
| GERD | ANX | rs11678860 | 2 | 212676106 | A | C | 0.045836897 | 0.045836897 | 0.0145 | 0.02059643 | 0.00007813 | 0.00004421 | new | ERBB4 | intronic | . | . | 2q34 |
| GERD | ANX | rs1083522 | 2 | 233745521 | T | C | 0.026861707 | 0.030129996 | -0.0174 | -0.020896826 | 0.000003212 | 0.00008905 | new | NGEF | intronic | . | . | 2q37.1 |
| GERD | ANX | rs1995175 | 3 | 23535928 | A | G | 0.026283999 | 0.026283999 | 0.018 | -0.02290022 | 0.000007261 | 0.00003344 | new | UBE2E2 | intronic | . | . | 3p24.3 |
| GERD | ANX | rs7642054 | 3 | 29726964 | T | C | 0.033653953 | 0.033653953 | 0.0153 | 0.025297308 | 0.00001711 | 0.00001413 | new | RBMS3 | intronic | . | . | 3p24.1 |
| GERD | ANX | rs6809116 | 3 | 44712097 | A | C | 0.024302847 | 0.024302847 | 0.0174 | -0.024098039 | 0.000003932 | 0.000001459 | new | ZKSCAN7-AS1 | ncRNA_intronic | . | . | 3p21.31 |
| GERD | ANX | rs352140 | 3 | 52256697 | T | C | 0.0229037 | 0.0229037 | -0.0156 | -0.02220471 | 0.000002948 | 0.000002926 | new | TLR9 | exonic | synonymous SNV | . | 3p21.2 |
| GERD | ANX | rs1905803 | 3 | 67984169 | A | C | 0.023482598 | 0.026008286 | -0.0375 | 0.044198687 | 0.000001904 | 0.00006741 | new | SUCLG2-AS1 | ncRNA_intronic | . | . | 3p14.1 |
| GERD | ANX | rs331164 | 3 | 78911097 | T | C | 0.025143221 | 0.025143221 | 0.0162 | -0.021703833 | 0.000004935 | 0.00003105 | new | ROBO1 | intronic | . | . | 3p12.3 |
| GERD | ANX | rs9808901 | 3 | 87702109 | T | C | 0.030151437 | 0.035314158 | -0.0162 | 0.019204409 | 0.000003913 | 0.0001217 | new | HTR1F | intergenic | . | . | 3p11.2 |
| GERD | ANX | rs144169510 | 3 | 109309963 | A | T | 0.048455327 | 0.048455327 | 0.02 | -0.028594967 | 0.00003062 | 0.0001339 | new | MIR4445 | intergenic | . | . | 3q13.13 |
| GERD | ANX | rs34242063 | 3 | 111170361 | T | C | 0.038323614 | 0.038323614 | -0.0209 | 0.027595712 | 0.00003034 | 0.00004096 | new | CD96 | intergenic | . | . | 3q13.13 |
| GERD | ANX | rs1585084 | 3 | 117616044 | T | C | 0.03072192 | 0.03072192 | -0.0171 | -0.023596214 | 0.00001261 | 0.000002676 | new | LINC02024 | intergenic | . | . | 3q13.32 |
| GERD | ANX | rs7619391 | 3 | 131541988 | A | G | 0.045252836 | 0.045252836 | -0.0244 | 0.035897891 | 0.00007378 | 0.000006447 | new | CPNE4 | intronic | . | . | 3q22.1 |
| GERD | ANX | rs2140364 | 3 | 167218543 | T | C | 0.047200132 | 0.047200132 | 0.0145 | -0.019804827 | 0.00008692 | 0.00003953 | new | WDR49 | intronic | . | . | 3q26.1 |
| GERD | ANX | rs39654 | 3 | 173095123 | A | G | 0.021263068 | 0.038628013 | -0.0184 | 0.019096496 | 0.000000125 | 0.0001458 | new | NLGN1 | intergenic | . | . | 3q26.31 |
| GERD | ANX | rs57010886 | 4 | 3283996 | T | G | 0.018201717 | 0.036212815 | -0.0284 | 0.029801491 | 5.064E-08 | 0.0001282 | new | MSANTD1 | intergenic | . | . | 4p16.3 |
| GERD | ANX | rs1609993 | 4 | 103514658 | T | C | 0.030418242 | 0.043445239 | 0.032 | -0.038003055 | 0.000002466 | 0.0001828 | new | NFKB1 | exonic | synonymous SNV | . | 4q24 |
| GERD | ANX | rs1510719 | 4 | 140938116 | T | C | 0.00206078 | 0.011070075 | 0.0244 | -0.021499466 | 5.022E-12 | 0.000009524 | known | MAML3 | intronic | . | . | 4q31.1 |
| GERD | ANX | rs4502841 | 5 | 30863015 | A | G | 0.037104021 | 0.049513169 | 0.016 | 0.017201207 | 0.000005367 | 0.0002597 | new | CDH6 | intergenic | . | . | 5p13.3 |
| GERD | ANX | rs7737552 | 5 | 61478134 | C | G | 0.016461143 | 0.01716726 | 0.0257 | 0.0299956 | 7.407E-08 | 0.0000287 | new | KIF2A | intergenic | . | . | 5q12.1 |
| GERD | ANX | rs391236 | 5 | 102678073 | A | G | 0.021809794 | 0.021809794 | 0.0182 | -0.023504069 | 0.000001894 | 0.00001138 | new | MACIR | intergenic | . | . | 5q21.1 |
| GERD | ANX | rs288177 | 5 | 107351694 | T | C | 0.034780354 | 0.034780354 | 0.0155 | -0.026703383 | 0.00001934 | 0.000000108 | new | FBXL17 | intronic | . | . | 5q21.3 |
| GERD | ANX | rs13153752 | 5 | 120096907 | A | G | 0.023467661 | 0.023467661 | 0.0166 | -0.029294937 | 0.000003311 | 7.954E-10 | new* | PRR16 | intergenic | . | . | 5q23.1 |
| GERD | ANX | rs55724060 | 5 | 166135549 | A | G | 0.036405824 | 0.036405824 | 0.0251 | -0.031800313 | 0.00002378 | 0.0000148 | new | NONE | intergenic | . | . | 5q34 |
| GERD | ANX | rs28752874 | 6 | 31302755 | T | C | 3.48871E-06 | 0.031975177 | 0.0284 | 0.019704583 | 1.141E-15 | 0.0001003 | known | HLA-B | intergenic | . | . | 6p21.33 |
| GERD | ANX | rs12198480 | 6 | 84342600 | T | C | 0.024614051 | 0.024614051 | 0.0178 | 0.026096505 | 0.000004192 | 0.000000218 | new | SNAP91 | intronic | . | . | 6q14.2 |
| GERD | ANX | rs3757323 | 6 | 152202007 | T | C | 0.014942859 | 0.035720868 | 0.0225 | -0.022695608 | 9.579E-10 | 0.0001246 | known | ESR1 | intronic | . | . | 6q25.1 |
| GERD | ANX | rs76450081 | 7 | 7264082 | A | G | 0.034909378 | 0.034909378 | -0.0202 | 0.027595712 | 0.00001966 | 0.00001888 | new | C1GALT1 | intronic | . | . | 7p22.1 |
| GERD | ANX | rs10272806 | 7 | 68394474 | A | G | 0.027669264 | 0.027669264 | -0.0164 | 0.020498464 | 0.000006232 | 0.00006339 | new | CT66 | intergenic | . | . | 7q11.22 |
| GERD | ANX | rs11770348 | 7 | 69920528 | A | T | 0.036981985 | 0.040091995 | 0.0185 | 0.026203665 | 0.000008813 | 0.0001504 | new | AUTS2 | intronic | . | . | 7q11.22 |
| GERD | ANX | rs2189246 | 7 | 82444372 | A | G | 0.019194964 | 0.019194964 | 0.0185 | 0.025404554 | 1.421E-07 | 6.482E-08 | new | PCLO | intronic | . | . | 7q21.11 |
| GERD | ANX | rs7778502 | 7 | 109101305 | T | C | 0.043605868 | 0.043605868 | -0.0146 | -0.020202707 | 0.00005655 | 0.00005285 | new | EIF3IP1 | intergenic | . | . | 7q31.1 |
| GERD | ANX | rs2894699 | 7 | 114059156 | T | C | 0.020698215 | 0.048797966 | -0.02 | -0.018102873 | 7.429E-09 | 0.0002484 | known | FOXP2 | intronic | . | . | 7q31.1 |
| GERD | ANX | rs12537384 | 7 | 115025708 | T | C | 0.02021652 | 0.025009959 | -0.0174 | 0.019302502 | 0.000000493 | 0.00006208 | new | LINC01392 | intergenic | . | . | 7q31.2 |
| GERD | ANX | rs6952845 | 7 | 135088308 | A | C | 0.026402525 | 0.026402525 | -0.0157 | 0.020802128 | 0.000007373 | 0.000009921 | new | CNOT4 | intronic | . | . | 7q33 |
| GERD | ANX | rs10238991 | 7 | 146672902 | A | G | 0.019405314 | 0.019405314 | 0.0189 | -0.020304753 | 2.578E-07 | 0.00002656 | new | CNTNAP2 | intronic | . | . | 7q35 |
| GERD | ANX | rs2622540 | 8 | 56320967 | A | G | 0.037598242 | 0.037598242 | -0.0152 | 0.02180063 | 0.00002767 | 0.000023 | new | XKR4 | intronic | . | . | 8q12.1 |
| GERD | ANX | rs16938729 | 8 | 74702487 | A | T | 0.0242904 | 0.048110464 | -0.028 | 0.029102387 | 1.313E-07 | 0.000238 | new | UBE2W | UTR3 | . | . | 8q21.11 |
| GERD | ANX | rs785957 | 8 | 84226239 | A | G | 0.043306648 | 0.043306648 | -0.0153 | 0.020802128 | 0.00005736 | 0.00005065 | new | LINC01419 | intergenic | . | . | 8q21.13 |
| GERD | ANX | rs13262595 | 8 | 143316970 | A | G | 0.035226844 | 0.035226844 | 0.0145 | -0.027196495 | 0.00002047 | 8.467E-09 | new* | TSNARE1 | intronic | . | . | 8q24.3 |
| GERD | ANX | rs10810098 | 9 | 14161670 | T | C | 0.025930156 | 0.025930156 | 0.0184 | -0.02200024 | 0.000006617 | 0.00003758 | new | NFIB | intronic | . | . | 9p23 |
| GERD | ANX | rs10739220 | 9 | 109054417 | T | C | 0.032342205 | 0.049469774 | -0.0199 | -0.020896826 | 0.000002672 | 0.000259 | new | MIR8081 | intergenic | . | . | 9q31.2 |
| GERD | ANX | rs11789671 | 9 | 120504614 | A | G | 0.016461143 | 0.022965266 | -0.0187 | -0.019203207 | 8.153E-08 | 0.00005244 | new | TLR4 | intergenic | . | . | 9q33.1 |
| GERD | ANX | rs1333904 | 9 | 122674412 | T | C | 0.023571546 | 0.025182469 | 0.0171 | 0.021595137 | 0.000002303 | 0.00006297 | new | LINC01613 | intergenic | . | . | 9q33.2 |
| GERD | ANX | rs656322 | 9 | 127834277 | A | G | 0.016461143 | 0.021571296 | -0.0188 | -0.02110107 | 0.000000031 | 0.0000465 | known | SCAI | intronic | . | . | 9q33.3 |
| GERD | ANX | rs10906187 | 10 | 12673010 | A | G | 0.033568816 | 0.034322717 | -0.0153 | 0.019900662 | 0.000008159 | 0.000112 | new | CAMK1D | intronic | . | . | 10p13 |
| GERD | ANX | rs12414777 | 10 | 104697781 | T | C | 0.018036376 | 0.018036376 | -0.0237 | -0.028101165 | 1.155E-07 | 0.000004897 | new | CNNM2 | intronic | . | . | 10q24.32 |
| GERD | ANX | rs4537697 | 10 | 106418969 | A | G | 0.016461143 | 0.016461143 | -0.0192 | -0.020100672 | 7.112E-08 | 0.00002017 | new | SORCS3 | intronic | . | . | 10q25.1 |
| GERD | ANX | rs11017001 | 10 | 131683760 | A | C | 0.019539307 | 0.019539307 | 0.0216 | 0.027897228 | 6.578E-07 | 0.000009075 | new | EBF3 | intronic | . | . | 10q26.3 |
| GERD | ANX | rs7083196 | 10 | 134954751 | T | C | 0.020674498 | 0.020674498 | -0.0168 | 0.021595137 | 0.000001091 | 0.00002257 | new | ADGRA1 | intergenic | . | . | 10q26.3 |
| GERD | ANX | rs4440995 | 11 | 31838128 | A | G | 0.011338742 | 0.011338742 | 0.029 | 0.030403106 | 1.871E-10 | 5.549E-07 | known | PAX6-AS1 | ncRNA_exonic | . | . | 11p13 |
| GERD | ANX | rs7128164 | 11 | 32319139 | C | G | 0.022945508 | 0.037990599 | -0.0184 | -0.02220471 | 5.723E-07 | 0.0001415 | new | WT1 | intergenic | . | . | 11p13 |
| GERD | ANX | rs2917531 | 11 | 89261646 | T | C | 0.037049691 | 0.045395322 | 0.0157 | -0.017695649 | 0.000006632 | 0.000201 | new | NOX4 | intronic | . | . | 11q14.3 |
| GERD | ANX | rs12787545 | 11 | 90012546 | A | G | 0.022182747 | 0.034366303 | -0.0188 | 0.018399683 | 2.856E-07 | 0.0001152 | new | DISC1FP1 | ncRNA_intronic | . | . | 11q14.3 |
| GERD | ANX | rs1487707 | 11 | 104362993 | A | G | 0.028206149 | 0.028206149 | -0.0162 | 0.020400488 | 0.000009306 | 0.00004129 | new | LINC02552 | intergenic | . | . | 11q22.3 |
| GERD | ANX | rs4938017 | 11 | 113303929 | T | C | 0.019405314 | 0.019405314 | 0.0174 | 0.022201708 | 5.181E-07 | 0.00000881 | new | DRD2 | intronic | . | . | 11q23.2 |
| GERD | ANX | rs73034035 | 12 | 215121 | A | G | 0.016461143 | 0.021870943 | 0.0214 | 0.020400488 | 1.857E-08 | 0.00004791 | known | IQSEC3 | intronic | . | . | 12p13.33 |
| GERD | ANX | rs1479406 | 12 | 15387543 | T | C | 0.019056023 | 0.024277751 | 0.0182 | 0.020802128 | 1.383E-07 | 0.00005844 | new | RERG | intergenic | . | . | 12p12.3 |
| GERD | ANX | rs28380723 | 12 | 121396646 | T | C | 0.037428562 | 0.037428562 | 0.0148 | -0.020702832 | 0.00002708 | 0.00001856 | new | HNF1A-AS1 | intergenic | . | . | 12q24.31 |
| GERD | ANX | rs2088714 | 13 | 53809246 | A | C | 0.035149636 | 0.035149636 | -0.0146 | -0.021795812 | 0.00002027 | 0.0000127 | new | LINC01065 | intergenic | . | . | 13q14.3 |
| GERD | ANX | rs9537780 | 13 | 58271352 | A | T | 0.024606253 | 0.03093447 | 0.0206 | -0.024897384 | 0.000001599 | 0.00009398 | new | PCDH17 | intronic | . | . | 13q21.1 |
| GERD | ANX | rs10507643 | 13 | 60513202 | C | G | 0.023178501 | 0.025479059 | 0.0171 | -0.024702613 | 0.000001826 | 0.00006453 | new | DIAPH3 | intronic | . | . | 13q21.2 |
| GERD | ANX | rs9571745 | 13 | 67843475 | A | C | 0.02213977 | 0.02213977 | -0.0164 | 0.025696986 | 0.000002267 | 8.777E-07 | new | PCDH9 | intergenic | . | . | 13q21.32 |
| GERD | ANX | rs9517313 | 13 | 99105892 | C | G | 0.041320185 | 0.041320185 | 0.0163 | 0.024302293 | 0.00005006 | 0.00004785 | new | FARP1 | UTR3 | . | . | 13q32.2 |
| GERD | ANX | rs10132715 | 14 | 42182103 | A | G | 0.016461143 | 0.016461143 | -0.0185 | 0.02399969 | 1.592E-08 | 9.855E-07 | known | LRFN5 | intronic | . | . | 14q21.1 |
| GERD | ANX | rs2415953 | 14 | 46798259 | A | G | 0.03277824 | 0.03277824 | -0.015 | 0.019998687 | 0.00001562 | 0.00004343 | new | LINC00871 | ncRNA_intronic | . | . | 14q21.2 |
| GERD | ANX | rs719732 | 14 | 75119057 | A | G | 0.019405314 | 0.019405314 | -0.0208 | 0.025404554 | 2.846E-07 | 0.000009392 | new | AREL1 | intergenic | . | . | 14q24.3 |
| GERD | ANX | rs8013266 | 14 | 103326780 | T | C | 0.01740184 | 0.031918294 | -0.0255 | 0.023599332 | 8.288E-09 | 0.00009997 | known | TRAF3 | intronic | . | . | 14q32.32 |
| GERD | ANX | rs3803358 | 15 | 40662773 | T | C | 0.048439622 | 0.048439622 | -0.0222 | -0.027299257 | 0.00003718 | 0.0001184 | new | DISP2 | UTR3 | . | . | 15q15.1 |
| GERD | ANX | rs9920305 | 15 | 69955799 | T | C | 0.0354764 | 0.0354764 | -0.016 | -0.022695608 | 0.00002113 | 0.00001054 | new | PCAT29 | ncRNA_intronic | . | . | 15q23 |
| GERD | ANX | rs58352128 | 16 | 6115246 | A | G | 0.020441811 | 0.020441811 | -0.0169 | -0.020498669 | 9.801E-07 | 0.00001441 | new | RBFOX1 | intronic | . | . | 16p13.3 |
| GERD | ANX | rs7498798 | 16 | 73097029 | A | G | 0.000213458 | 0.041417037 | -0.0271 | 0.019900662 | 1.703E-13 | 0.0001662 | known | ZFHX3 | intergenic | . | . | 16q22.3 |
| GERD | ANX | rs35449523 | 16 | 77122295 | C | G | 0.015207985 | 0.034396255 | -0.0216 | 0.021399391 | 1.4E-09 | 0.0001154 | known | MON1B | intergenic | . | . | 16q23.1 |
| GERD | ANX | rs75664430 | 17 | 8064779 | C | G | 0.016461143 | 0.016461143 | 0.0214 | -0.027104018 | 8.194E-08 | 0.0000157 | new | VAMP2 | intronic | . | . | 17p13.1 |
| GERD | ANX | rs11651809 | 17 | 43255681 | C | G | 0.040180702 | 0.040180702 | -0.0164 | 0.023696995 | 0.00004169 | 0.00003872 | new | LOC105371795 | intergenic | . | . | 17q21.31 |
| GERD | ANX | rs3809770 | 17 | 47047596 | A | G | 0.027998567 | 0.048411545 | 0.0178 | -0.018296362 | 0.000001049 | 0.0002425 | new | GIP | intergenic | . | . | 17q21.32 |
| GERD | ANX | rs7223643 | 17 | 65830282 | A | G | 0.022631016 | 0.022631016 | -0.0206 | 0.026602982 | 0.000002787 | 0.00001073 | new | BPTF | intronic | . | . | 17q24.2 |
| GERD | ANX | rs3110690 | 18 | 27919922 | A | G | 0.035071324 | 0.035071324 | -0.0186 | -0.021601645 | 0.00001136 | 0.00009232 | new | MIR302F | intergenic | . | . | 18q12.1 |
| GERD | ANX | rs11665070 | 18 | 35152563 | A | G | 0.01136447 | 0.017024018 | -0.0239 | -0.021295141 | 2.023E-10 | 0.00002817 | known | CELF4 | intergenic | . | . | 18q12.2 |
| GERD | ANX | rs17411339 | 18 | 50807359 | A | G | 0.016461143 | 0.016461143 | -0.0194 | 0.022299507 | 7.857E-08 | 0.000003346 | new | DCC | intronic | . | . | 18q21.2 |
| GERD | ANX | rs4468713 | 18 | 53104019 | T | C | 0.013965433 | 0.044591233 | -0.0214 | -0.018296362 | 7.381E-10 | 0.0001929 | known | TCF4 | intronic | . | . | 18q21.2 |
| GERD | ANX | rs66723169 | 18 | 57808978 | A | C | 0.016461143 | 0.016461143 | -0.0238 | -0.024805119 | 1.522E-08 | 0.00001562 | known | MC4R | intergenic | . | . | 18q21.32 |
| GERD | ANX | rs12985909 | 19 | 18439383 | T | C | 0.000503629 | 0.039737821 | -0.0248 | -0.018102873 | 5.502E-13 | 0.0001536 | known | LSM4 | intergenic | . | . | 19p13.11 |
| GERD | ANX | rs12973240 | 19 | 32038701 | A | G | 0.018310223 | 0.018310223 | -0.0195 | 0.024399887 | 1.213E-07 | 0.000002767 | new | THEG5 | intergenic | . | . | 19q12 |
| GERD | ANX | rs2295762 | 20 | 31025535 | A | G | 0.044411261 | 0.044411261 | -0.0137 | 0.019302502 | 0.00005061 | 0.00006418 | new | ASXL1 | UTR3 | . | . | 20q11.21 |
| GERD | ANX | rs4470425 | 22 | 40716488 | A | C | 0.014388663 | 0.042369559 | 0.0235 | 0.019704583 | 8.263E-10 | 0.0001738 | known | TNRC6B | intronic | . | . | 22q13.1 |
| GERD | ANX | rs11090045 | 22 | 41753603 | A | G | 0.016461143 | 0.016461143 | 0.0212 | 0.024399887 | 2.958E-08 | 0.00000726 | known | ZC3H7B | UTR3 | . | . | 22q13.2 |
| GERD | MDD | rs632051 | 1 | 5291879 | A | C | 0.042921922 | 0.042921922 | -0.0175 | 0.016001294 | 0.00009662 | 0.000009244 | new | AJAP1 | intergenic | . | . | 1p36.32 |
| GERD | MDD | rs11121211 | 1 | 8714239 | T | C | 0.008860788 | 0.020116072 | 0.0248 | 0.016099701 | 1.107E-07 | 0.00003291 | new | RERE | intronic | . | . | 1p36.23 |
| GERD | MDD | rs3766823 | 1 | 32197257 | A | G | 0.016459409 | 0.028910923 | 0.0229 | 0.014297305 | 0.000001 | 0.00008524 | new | ADGRB2 | intronic | . | . | 1p35.2 |
| GERD | MDD | rs60906999 | 1 | 33770060 | A | G | 0.034688752 | 0.034688752 | -0.0147 | 0.013301146 | 0.00004579 | 0.000006904 | new | A3GALT2 | intergenic | . | . | 1p35.1 |
| GERD | MDD | rs6677366 | 1 | 37187504 | A | G | 0.004940021 | 0.004940021 | 0.021 | 0.022103899 | 4.278E-08 | 3.153E-12 | known | GRIK3 | intergenic | . | . | 1p34.3 |
| GERD | MDD | rs11587148 | 1 | 50994712 | T | C | 0.023114156 | 0.033332164 | -0.0178 | 0.012195334 | 0.000002682 | 0.0001189 | new | FAF1 | intronic | . | . | 1p32.3 |
| GERD | MDD | rs543192 | 1 | 65724871 | A | G | 0.02966377 | 0.040440175 | 0.0186 | -0.012599036 | 0.000005296 | 0.0001733 | new | DNAJC6 | intergenic | . | . | 1p31.3 |
| GERD | MDD | rs2503185 | 1 | 66461401 | A | G | 0.002267811 | 0.003350382 | 0.0219 | -0.014697481 | 7.154E-10 | 2.595E-07 | known | PDE4B | intronic | . | . | 1p31.3 |
| GERD | MDD | rs1325266 | 1 | 67160602 | A | G | 0.043030925 | 0.043030925 | -0.0129 | 0.012096541 | 0.00007167 | 0.00001909 | new | SGIP1 | intronic | . | . | 1p31.3 |
| GERD | MDD | rs12082973 | 1 | 69254610 | A | G | 0.025924782 | 0.025924782 | -0.0267 | -0.020396604 | 0.0000134 | 0.000009378 | new | DEPDC1-AS1 | intergenic | . | . | 1p31.2 |
| GERD | MDD | rs61765555 | 1 | 73957815 | T | C | 0.006534327 | 0.006534327 | -0.0218 | -0.021795812 | 1.048E-07 | 3.146E-11 | new* | LINC01360 | intergenic | . | . | 1p31.1 |
| GERD | MDD | rs34959106 | 1 | 80020959 | A | G | 0.044313142 | 0.044313142 | 0.0146 | 0.012699025 | 0.00004619 | 0.00004917 | new | ADGRL4 | intergenic | . | . | 1p31.1 |
| GERD | MDD | rs12084568 | 1 | 80800862 | T | C | 0.007802693 | 0.007802693 | 0.0202 | -0.019101273 | 1.761E-07 | 4.388E-08 | new* | LINC01781 | intergenic | . | . | 1p31.1 |
| GERD | MDD | rs4655961 | 1 | 87889822 | A | G | 0.018889757 | 0.018889757 | -0.0172 | -0.014596005 | 0.000006677 | 0.000003273 | new | LINC01364 | intergenic | . | . | 1p22.3 |
| GERD | MDD | rs7529408 | 1 | 88825811 | T | C | 0.027284527 | 0.042344182 | 0.0219 | -0.013399372 | 0.000003535 | 0.0002155 | new | PKN2-AS1 | intergenic | . | . | 1p22.2 |
| GERD | MDD | rs7553296 | 1 | 90671625 | A | C | 0.03122381 | 0.03122381 | 0.0146 | 0.013202463 | 0.00003456 | 0.000005303 | new | ZNF326 | intergenic | . | . | 1p22.2 |
| GERD | MDD | rs61787782 | 1 | 98310239 | A | G | 0.012062107 | 0.040806731 | -0.0206 | 0.012303994 | 1.391E-07 | 0.000202 | known | DPYD | intronic | . | . | 1p21.3 |
| GERD | MDD | rs1521185 | 1 | 151764049 | A | G | 0.026945521 | 0.026945521 | -0.021 | -0.017797439 | 0.00001753 | 0.000006702 | new | TDRKH-AS1 | ncRNA_exonic | . | . | 1q21.3 |
| GERD | MDD | rs333423 | 1 | 174635695 | A | G | 0.001019974 | 0.006930177 | 0.0273 | -0.015001969 | 1.216E-12 | 0.000001677 | known | RABGAP1L | intronic | . | . | 1q25.1 |
| GERD | MDD | rs10920936 | 1 | 191377664 | T | G | 0.00572824 | 0.009451978 | 0.0195 | 0.01339982 | 6.705E-08 | 0.000003957 | new | LINC01680 | intergenic | . | . | 1q31.2 |
| GERD | MDD | rs17621768 | 1 | 197509624 | A | G | 0.024027609 | 0.038395167 | -0.0168 | 0.012096541 | 0.000002432 | 0.0001713 | new | DENND1B | intronic | . | . | 1q31.3 |
| GERD | MDD | rs6428459 | 1 | 198366112 | T | G | 0.031629437 | 0.031629437 | -0.0169 | 0.015499263 | 0.000042 | 0.000003005 | new | NEK7 | intergenic | . | . | 1q31.3 |
| GERD | MDD | rs12131852 | 1 | 214433323 | C | G | 0.013015024 | 0.013015024 | 0.0179 | -0.020304753 | 0.000001626 | 8.091E-11 | new* | SMYD2 | intergenic | . | . | 1q32.3 |
| GERD | MDD | rs708777 | 1 | 226933722 | A | G | 0.042490716 | 0.042490716 | -0.0151 | 0.013103769 | 0.00008098 | 0.00001354 | new | ITPKB | intergenic | . | . | 1q42.12 |
| GERD | MDD | rs2756224 | 1 | 241791117 | T | C | 0.027576062 | 0.027576062 | 0.0166 | -0.01360209 | 0.00001883 | 0.000006926 | new | OPN3 | intronic | . | . | 1q43 |
| GERD | MDD | rs9724830 | 1 | 243251189 | T | C | 0.009508731 | 0.023729564 | 0.0218 | 0.013103769 | 1.223E-07 | 0.00005111 | new | LINC01347 | ncRNA_exonic | . | . | 1q43 |
| GERD | MDD | rs67446571 | 2 | 22554648 | A | G | 0.015622027 | 0.015622027 | -0.0158 | 0.02350166 | 0.000003402 | 2.462E-16 | new* | LINC01884 | intergenic | . | . | 2p24.1 |
| GERD | MDD | rs56009471 | 2 | 23890084 | A | C | 0.033495219 | 0.033495219 | -0.0188 | 0.019302502 | 0.00005254 | 7.843E-07 | new | KLHL29 | intronic | . | . | 2p24.1 |
| GERD | MDD | rs72889466 | 2 | 51056712 | A | C | 0.024037796 | 0.025193993 | 0.0194 | 0.013902905 | 0.000004546 | 0.00005241 | new | NRXN1 | intronic | . | . | 2p16.3 |
| GERD | MDD | rs11691352 | 2 | 59949495 | A | G | 0.029435704 | 0.030173892 | -0.0175 | -0.012204169 | 0.000008192 | 0.00007189 | new | LINC01793 | intergenic | . | . | 2p16.1 |
| GERD | MDD | rs10188396 | 2 | 60450992 | T | C | 0.011137418 | 0.011137418 | -0.0181 | -0.014900463 | 9.433E-07 | 7.733E-07 | new | MIR4432HG | intergenic | . | . | 2p16.1 |
| GERD | MDD | rs1519104 | 2 | 66680892 | A | G | 0.015655047 | 0.03960737 | -0.0189 | 0.011800104 | 0.000000521 | 0.0001864 | new | MEIS1 | intronic | . | . | 2p14 |
| GERD | MDD | rs7851 | 2 | 73478461 | A | T | 0.011138912 | 0.015953103 | -0.0189 | 0.013804281 | 5.148E-07 | 0.00001761 | new | CCT7 | exonic | synonymous SNV | . | 2p13.2 |
| GERD | MDD | rs7607172 | 2 | 100492135 | T | C | 0.001080338 | 0.025147234 | -0.025 | 0.011800104 | 2.296E-12 | 0.00005931 | known | AFF3 | intronic | . | . | 2q11.2 |
| GERD | MDD | rs4851300 | 2 | 100958804 | A | C | 0.013960945 | 0.016465004 | -0.0185 | -0.012902885 | 0.000001176 | 0.00001918 | new | LONRF2 | intergenic | . | . | 2q11.2 |
| GERD | MDD | rs11691266 | 2 | 109829904 | A | G | 0.005629585 | 0.018137993 | 0.0198 | -0.012700309 | 2.781E-08 | 0.00002482 | known | SH3RF3 | intronic | . | . | 2q12.3 |
| GERD | MDD | rs7581524 | 2 | 113234316 | T | C | 0.001178409 | 0.019272413 | 0.0257 | 0.013103769 | 1.523E-11 | 0.00002918 | known | TTL | intergenic | . | . | 2q13 |
| GERD | MDD | rs1186066 | 2 | 125024950 | T | C | 0.033926583 | 0.033926583 | -0.0158 | 0.016503072 | 0.00005533 | 3.025E-09 | new* | CNTNAP5 | intronic | . | . | 2q14.3 |
| GERD | MDD | rs10209281 | 2 | 138307575 | T | C | 0.031289462 | 0.031289462 | 0.0141 | 0.013202463 | 0.00004032 | 0.000003137 | new | THSD7B | intronic | . | . | 2q22.1 |
| GERD | MDD | rs28396876 | 2 | 144487090 | A | G | 0.002024156 | 0.002318566 | 0.022 | -0.015205013 | 2.093E-10 | 9.076E-08 | known | ARHGAP15 | intronic | . | . | 2q22.3 |
| GERD | MDD | rs7595938 | 2 | 148840271 | A | G | 0.010509797 | 0.013437109 | 0.0183 | 0.012797758 | 5.201E-07 | 0.00001114 | new | MBD5 | intronic | . | . | 2q23.1 |
| GERD | MDD | rs10185435 | 2 | 155287778 | T | C | 0.043763771 | 0.043763771 | -0.0208 | -0.017695649 | 0.00006584 | 0.00002561 | new | GALNT13 | intronic | . | . | 2q24.1 |
| GERD | MDD | rs17755617 | 2 | 161935424 | A | C | 0.019739871 | 0.019739871 | -0.0171 | -0.018398214 | 0.000007886 | 2.485E-09 | new* | TANK | intergenic | . | . | 2q24.2 |
| GERD | MDD | rs3754757 | 2 | 173835127 | A | G | 0.009467023 | 0.009467023 | -0.0183 | -0.014504686 | 4.789E-07 | 7.419E-07 | new | RAPGEF4 | intronic | . | . | 2q31.1 |
| GERD | MDD | rs6744754 | 2 | 175177999 | T | C | 0.040372374 | 0.040372374 | 0.0157 | -0.020600746 | 0.0001102 | 2.006E-09 | new* | LINC01305 | intergenic | . | . | 2q31.1 |
| GERD | MDD | rs9287989 | 2 | 176717741 | T | C | 0.00279578 | 0.025615577 | 0.0216 | 0.011701272 | 2.838E-10 | 0.00006205 | known | LNPK | intergenic | . | . | 2q31.1 |
| GERD | MDD | rs1598794 | 2 | 194006187 | T | G | 0.000722484 | 0.035508983 | 0.0259 | -0.01069701 | 6.184E-14 | 0.0001416 | known | PCGEM1 | intergenic | . | . | 2q32.3 |
| GERD | MDD | rs55677233 | 2 | 201168695 | T | C | 0.005933778 | 0.013498925 | -0.0189 | 0.012303994 | 5.623E-08 | 0.00001128 | new | SPATS2L | intergenic | . | . | 2q33.1 |
| GERD | MDD | rs7603172 | 2 | 208020941 | A | G | 0.028035262 | 0.028035262 | 0.0149 | 0.01949866 | 0.00002614 | 1.337E-10 | new* | KLF7 | intronic | . | . | 2q33.3 |
| GERD | MDD | rs11897810 | 2 | 208756744 | T | C | 0.04085383 | 0.04085383 | 0.0168 | 0.013202463 | 0.00004334 | 0.00003073 | new | PLEKHM3 | intronic | . | . | 2q33.3 |
| GERD | MDD | rs11678860 | 2 | 212676106 | A | C | 0.036805117 | 0.036805117 | 0.0145 | 0.014799939 | 0.00007813 | 8.777E-07 | new | ERBB4 | intronic | . | . | 2q34 |
| GERD | MDD | rs3770161 | 2 | 222334358 | T | C | 0.035739551 | 0.035979714 | -0.0143 | -0.010899181 | 0.00001303 | 0.00009887 | new | EPHA4 | intronic | . | . | 2q36.1 |
| GERD | MDD | rs6704768 | 2 | 233592501 | A | G | 0.004384859 | 0.008403106 | 0.0194 | -0.013004189 | 2.791E-08 | 0.000002829 | known | GIGYF2 | intronic | . | . | 2q37.1 |
| GERD | MDD | rs9881798 | 3 | 16846967 | A | C | 0.014382338 | 0.014382338 | -0.0173 | 0.01660143 | 0.000002422 | 3.296E-09 | new* | PLCL2 | intergenic | . | . | 3p24.3 |
| GERD | MDD | rs2683532 | 3 | 24265142 | A | G | 0.048251001 | 0.048251001 | -0.0162 | 0.012096541 | 0.00006276 | 0.00005748 | new | THRB | intronic | . | . | 3p24.2 |
| GERD | MDD | rs7644506 | 3 | 29723935 | T | C | 0.02110068 | 0.02110068 | -0.0158 | -0.014200349 | 0.000009571 | 0.000003054 | new | RBMS3 | intronic | . | . | 3p24.1 |
| GERD | MDD | rs56403804 | 3 | 43368021 | A | G | 0.012113408 | 0.012531059 | 0.0253 | 0.017702384 | 9.874E-07 | 0.000009276 | known | SNRK | intronic | . | . | 3p22.1 |
| GERD | MDD | rs9825080 | 3 | 44251270 | T | C | 0.005877674 | 0.005877674 | -0.0218 | -0.016098895 | 7.295E-08 | 9.879E-07 | new | TOPAZ1 | intergenic | . | . | 3p21.31 |
| GERD | MDD | rs3774751 | 3 | 50209053 | T | G | 0.002267811 | 0.002267811 | -0.0208 | -0.015499499 | 8.271E-10 | 3.545E-08 | known | SEMA3F | intronic | . | . | 3p21.31 |
| GERD | MDD | rs352140 | 3 | 52256697 | T | C | 0.015012945 | 0.015012945 | -0.0156 | 0.014799939 | 0.000002948 | 4.218E-07 | new | TLR9 | exonic | synonymous SNV | . | 3p21.2 |
| GERD | MDD | rs4974157 | 3 | 56015670 | A | G | 0.03621235 | 0.03621235 | -0.0167 | 0.012699025 | 0.00003077 | 0.00002263 | new | ERC2 | intronic | . | . | 3p14.3 |
| GERD | MDD | rs12715593 | 3 | 65178981 | T | C | 0.016509718 | 0.016509718 | -0.0161 | -0.012801592 | 0.000002109 | 0.00001777 | new | MAGI1 | intergenic | . | . | 3p14.1 |
| GERD | MDD | rs12638072 | 3 | 65654480 | A | G | 0.023073088 | 0.032529911 | 0.0175 | 0.011800104 | 0.000002788 | 0.0001114 | new | MAGI1 | intronic | . | . | 3p14.1 |
| GERD | MDD | rs7610856 | 3 | 71579022 | A | C | 0.025830296 | 0.025830296 | -0.015 | -0.012295278 | 0.00001078 | 0.00001597 | new | FOXP1 | intronic | . | . | 3p13 |
| GERD | MDD | rs1441958 | 3 | 77333352 | A | C | 0.022534381 | 0.025194002 | 0.0168 | 0.011997738 | 0.000003554 | 0.00005558 | new | ROBO2 | intronic | . | . | 3p12.3 |
| GERD | MDD | rs9631522 | 3 | 78562318 | T | C | 0.040436769 | 0.040436769 | -0.0155 | -0.013703465 | 0.00003507 | 0.00003912 | new | ROBO1 | intergenic | . | . | 3p12.3 |
| GERD | MDD | rs324028 | 3 | 113893343 | A | G | 0.034701426 | 0.034701426 | 0.0154 | -0.013298028 | 0.00001814 | 0.000041 | new | DRD3 | intronic | . | . | 3q13.31 |
| GERD | MDD | rs7427162 | 3 | 117586681 | T | C | 0.021933619 | 0.021933619 | -0.0174 | 0.016896448 | 0.00001074 | 1.255E-07 | new | LINC02024 | intergenic | . | . | 3q13.32 |
| GERD | MDD | rs2140364 | 3 | 167218543 | T | C | 0.037695434 | 0.037695434 | 0.0145 | -0.015001969 | 0.00008692 | 3.998E-07 | new | WDR49 | intronic | . | . | 3q26.1 |
| GERD | MDD | rs247975 | 3 | 173107443 | T | C | 0.004399982 | 0.013971547 | -0.0194 | -0.01239652 | 1.911E-08 | 0.00001241 | known | NLGN1 | intergenic | . | . | 3q26.31 |
| GERD | MDD | rs13075342 | 3 | 177675295 | C | G | 0.006059219 | 0.009938873 | 0.023 | -0.015804232 | 8.036E-08 | 0.00000458 | new | LINC02015 | intergenic | . | . | 3q26.32 |
| GERD | MDD | rs1479175 | 3 | 181208277 | A | T | 0.036870047 | 0.036870047 | 0.0169 | -0.017502275 | 0.00007874 | 0.000002808 | new | SOX2-OT | ncRNA_intronic | . | . | 3q26.33 |
| GERD | MDD | rs3845942 | 3 | 193421825 | A | G | 0.025223234 | 0.025223234 | -0.0157 | 0.015203834 | 0.00001731 | 1.787E-07 | new | OPA1 | intergenic | . | . | 3q29 |
| GERD | MDD | rs2051559 | 4 | 3298800 | T | C | 0.002614118 | 0.007315862 | -0.0304 | 0.01949866 | 5.112E-09 | 0.00000194 | known | RGS12 | intergenic | . | . | 4p16.3 |
| GERD | MDD | rs11947190 | 4 | 23476360 | T | C | 0.00972057 | 0.02948222 | -0.0197 | -0.012001734 | 1.098E-07 | 0.00008921 | new | PPARGC1A | intergenic | . | . | 4p15.2 |
| GERD | MDD | rs10024502 | 4 | 61461481 | T | G | 0.028752587 | 0.048249675 | -0.0184 | 0.012303994 | 0.000003217 | 0.0002948 | new | MIR548AG1 | intergenic | . | . | 4q13.1 |
| GERD | MDD | rs72643078 | 4 | 61825207 | A | G | 0.046970296 | 0.046970296 | 0.0145 | 0.012699025 | 0.00009508 | 0.0000225 | new | MIR548AG1 | intergenic | . | . | 4q13.1 |
| GERD | MDD | rs1122336 | 4 | 62911559 | T | C | 0.035930805 | 0.035930805 | 0.0215 | 0.020900064 | 0.00007036 | 0.000001599 | new | ADGRL3 | intronic | . | . | 4q13.1 |
| GERD | MDD | rs1031035 | 4 | 102223084 | T | C | 0.0020903 | 0.006594077 | -0.0242 | 0.014898465 | 2.297E-10 | 0.000001477 | known | PPP3CA | intronic | . | . | 4q24 |
| GERD | MDD | rs11933762 | 4 | 119307126 | A | G | 0.002917955 | 0.010217119 | -0.0276 | 0.017604134 | 7.535E-09 | 0.000004979 | known | PRSS12 | intergenic | . | . | 4q26 |
| GERD | MDD | rs1445842 | 4 | 138601778 | T | C | 0.004991966 | 0.010897491 | -0.0253 | -0.017604046 | 4.294E-08 | 0.000006055 | known | LINC02172 | intergenic | . | . | 4q28.3 |
| GERD | MDD | rs1510719 | 4 | 140938116 | T | C | 0.001109136 | 0.001109136 | 0.0244 | -0.017095295 | 5.022E-12 | 2.583E-09 | known | MAML3 | intronic | . | . | 4q31.1 |
| GERD | MDD | rs7655694 | 4 | 164519194 | T | C | 0.044406738 | 0.044406738 | 0.0137 | 0.014297305 | 0.0001529 | 0.000001608 | new | MARCHF1 | intronic | . | . | 4q32.3 |
| GERD | MDD | rs17206022 | 5 | 7333533 | T | G | 0.014156379 | 0.014156379 | -0.0165 | -0.013298028 | 0.000002199 | 0.000003767 | new | LINC02123 | intergenic | . | . | 5p15.31 |
| GERD | MDD | rs4502841 | 5 | 30863015 | A | G | 0.019107991 | 0.019107991 | 0.016 | -0.012902885 | 0.000005367 | 0.000007805 | new | CDH6 | intergenic | . | . | 5p13.3 |
| GERD | MDD | rs1482698 | 5 | 44539453 | C | G | 0.043413809 | 0.043413809 | 0.0139 | 0.014799939 | 0.0001432 | 0.000001669 | new | LINC02224 | intergenic | . | . | 5p12 |
| GERD | MDD | rs62369904 | 5 | 45092933 | A | T | 0.002267811 | 0.009699218 | 0.0272 | -0.017400515 | 5.847E-10 | 0.000004262 | known | HCN1 | intergenic | . | . | 5p12 |
| GERD | MDD | rs4481304 | 5 | 50759415 | A | G | 0.017411234 | 0.031643703 | 0.0171 | 0.011602431 | 0.000001074 | 0.0001056 | new | ISL1 | intergenic | . | . | 5q11.2 |
| GERD | MDD | rs7727098 | 5 | 51574444 | A | G | 0.015757791 | 0.015757791 | -0.0152 | -0.013896105 | 0.000003503 | 0.000001137 | new | LINC02118 | intergenic | . | . | 5q11.2 |
| GERD | MDD | rs58962658 | 5 | 52789117 | T | C | 0.045537818 | 0.045537818 | 0.0177 | -0.015397942 | 0.0001095 | 0.00001182 | new | FST | intergenic | . | . | 5q11.2 |
| GERD | MDD | rs10054434 | 5 | 61534535 | C | G | 0.003681428 | 0.003681428 | 0.0277 | -0.024702613 | 1.625E-08 | 1.47E-08 | known | KIF2A | intergenic | . | . | 5q12.1 |
| GERD | MDD | rs3846617 | 5 | 102969219 | A | G | 0.037817917 | 0.037817917 | 0.0148 | 0.012303994 | 0.00003226 | 0.00002833 | new | NUDT12 | intergenic | . | . | 5q21.2 |
| GERD | MDD | rs1357716 | 5 | 105207305 | A | T | 0.037427903 | 0.037427903 | 0.0232 | -0.023002542 | 0.00008418 | 6.234E-07 | new | RAB9BP1 | intergenic | . | . | 5q21.3 |
| GERD | MDD | rs288178 | 5 | 107351517 | T | C | 0.037126943 | 0.037126943 | 0.0155 | -0.012599036 | 0.00002101 | 0.0000491 | new | FBXL17 | intronic | . | . | 5q21.3 |
| GERD | MDD | rs77744641 | 5 | 119971968 | A | G | 0.009502537 | 0.009502537 | -0.0237 | 0.022504855 | 4.868E-07 | 6.108E-09 | new* | PRR16 | intronic | . | . | 5q23.1 |
| GERD | MDD | rs79122663 | 5 | 137617438 | C | G | 0.012381669 | 0.018067051 | 0.0166 | 0.012402767 | 6.735E-07 | 0.00002457 | new | CDC25C | intergenic | . | . | 5q31.2 |
| GERD | MDD | rs11167642 | 5 | 153222233 | T | C | 0.047874957 | 0.047874957 | 0.0178 | -0.022695608 | 0.0001922 | 4.947E-10 | new* | GRIA1 | intergenic | . | . | 5q33.2 |
| GERD | MDD | rs72823607 | 5 | 166137119 | T | G | 0.039528008 | 0.039528008 | -0.0255 | -0.017695649 | 0.00003376 | 0.00003581 | new | NONE | intergenic | . | . | 5q34 |
| GERD | MDD | rs12524341 | 6 | 14729525 | T | C | 0.01548566 | 0.025907294 | -0.0237 | -0.017502275 | 9.257E-07 | 0.00006382 | new | LINC01108 | intergenic | . | . | 6p23 |
| GERD | MDD | rs72829857 | 6 | 16966052 | A | G | 0.036387778 | 0.036387778 | 0.0187 | -0.017095295 | 0.00007432 | 2.653E-07 | new | STMND1 | intergenic | . | . | 6p22.3 |
| GERD | MDD | rs28367822 | 6 | 31305405 | A | G | 1.26367E-05 | 0.027878243 | -0.0285 | 0.012195334 | 5.338E-16 | 0.00007717 | known | HLA-B | intergenic | . | . | 6p21.33 |
| GERD | MDD | rs4714291 | 6 | 40003632 | A | G | 0.033135329 | 0.033135329 | -0.0158 | -0.020100672 | 0.00005032 | 3.656E-10 | new* | MOCS1 | intergenic | . | . | 6p21.2 |
| GERD | MDD | rs11756556 | 6 | 64812713 | T | C | 0.001762614 | 0.011737599 | 0.0253 | -0.014900463 | 1.414E-10 | 0.000007616 | known | EYS | intronic | . | . | 6q12 |
| GERD | MDD | rs12198480 | 6 | 84342600 | T | C | 0.016753057 | 0.016753057 | 0.0178 | 0.013804281 | 0.000004192 | 0.00000395 | new | SNAP91 | intronic | . | . | 6q14.2 |
| GERD | MDD | rs13198331 | 6 | 101632027 | T | G | 0.011064464 | 0.02939288 | -0.0283 | -0.018703831 | 1.614E-07 | 0.00008859 | new | GRIK2 | intergenic | . | . | 6q16.3 |
| GERD | MDD | rs221618 | 6 | 105472320 | A | G | 0.035431129 | 0.035431129 | 0.0132 | -0.013298028 | 0.00006627 | 0.000003377 | new | LIN28B | intronic | . | . | 6q16.3 |
| GERD | MDD | rs9389524 | 6 | 137924466 | T | C | 0.030712892 | 0.030712892 | 0.0179 | -0.015397942 | 0.00002944 | 0.000006496 | new | LOC102723649 | intergenic | . | . | 6q23.3 |
| GERD | MDD | rs198683 | 6 | 143200547 | T | C | 0.001251831 | 0.035634718 | -0.0232 | 0.010504633 | 7.132E-12 | 0.0001428 | known | HIVEP2 | intronic | . | . | 6q24.2 |
| GERD | MDD | rs6942134 | 6 | 147952023 | C | G | 0.003293421 | 0.025934887 | -0.022 | 0.011800104 | 3.025E-09 | 0.00006399 | known | SAMD5 | intergenic | . | . | 6q24.3 |
| GERD | MDD | rs4870060 | 6 | 152231404 | A | G | 0.003003222 | 0.003003222 | -0.0213 | 0.028499989 | 8.368E-09 | 1.991E-21 | known | ESR1 | intronic | . | . | 6q25.1 |
| GERD | MDD | rs2764650 | 6 | 164621949 | T | C | 0.009466603 | 0.011070529 | -0.0178 | -0.012801592 | 3.973E-07 | 0.000006348 | new | LOC102724152 | intergenic | . | . | 6q27 |
| GERD | MDD | rs6927228 | 6 | 166420900 | T | C | 0.019648993 | 0.044399526 | 0.0182 | -0.011202515 | 9.258E-07 | 0.000245 | new | LINC00602 | intergenic | . | . | 6q27 |
| GERD | MDD | rs11766944 | 7 | 1888051 | A | G | 0.001072261 | 0.001072261 | -0.0301 | -0.030902605 | 1.917E-12 | 3.389E-19 | known | MAD1L1 | intronic | . | . | 7p22.3 |
| GERD | MDD | rs10227151 | 7 | 3599352 | C | G | 0.005034232 | 0.005034232 | -0.0223 | -0.022296735 | 4.515E-08 | 1.539E-10 | known | SDK1 | intronic | . | . | 7p22.2 |
| GERD | MDD | rs7802254 | 7 | 8697172 | C | G | 0.001102968 | 0.001865253 | 0.0262 | 0.017102908 | 4.662E-12 | 3.312E-08 | known | NXPH1 | intronic | . | . | 7p21.3 |
| GERD | MDD | rs62447639 | 7 | 12486803 | T | C | 0.029198596 | 0.029198596 | 0.0218 | -0.018398214 | 0.00002777 | 0.000004605 | new | VWDE | intergenic | . | . | 7p21.3 |
| GERD | MDD | rs13438525 | 7 | 21575459 | T | C | 0.03528445 | 0.03528445 | 0.0159 | -0.011698157 | 0.00001409 | 0.00007684 | new | DNAH11 | intergenic | . | . | 7p15.3 |
| GERD | MDD | rs73079438 | 7 | 27548192 | T | C | 0.042319087 | 0.042319087 | -0.0208 | -0.017003748 | 0.0000411 | 0.00004222 | new | HIBADH | intergenic | . | . | 7p15.2 |
| GERD | MDD | rs1424297 | 7 | 37043363 | T | C | 0.049723727 | 0.049723727 | 0.0184 | 0.018703985 | 0.0002171 | 0.000002162 | new | ELMO1-AS1 | ncRNA_intronic | . | . | 7p14.2 |
| GERD | MDD | rs79039482 | 7 | 68381673 | A | T | 0.032661693 | 0.032661693 | 0.0158 | 0.014297305 | 0.00002392 | 0.00001637 | new | CT66 | intergenic | . | . | 7q11.22 |
| GERD | MDD | rs10275122 | 7 | 69897826 | A | G | 0.009809909 | 0.009809909 | -0.0214 | 0.0161981 | 5.427E-07 | 0.00000393 | new | AUTS2 | intronic | . | . | 7q11.22 |
| GERD | MDD | rs10954666 | 7 | 81829434 | A | G | 0.02810474 | 0.02810474 | -0.0157 | -0.012295278 | 0.00001233 | 0.00002137 | new | CACNA2D1 | intronic | . | . | 7q21.11 |
| GERD | MDD | rs2189246 | 7 | 82444372 | A | G | 0.007297198 | 0.007297198 | 0.0185 | -0.018897437 | 1.421E-07 | 9.312E-12 | new* | PCLO | intronic | . | . | 7q21.11 |
| GERD | MDD | rs4729021 | 7 | 91758422 | A | C | 0.033691316 | 0.033691316 | -0.0143 | -0.011202515 | 0.00001364 | 0.00005731 | new | CYP51A1 | intronic | . | . | 7q21.2 |
| GERD | MDD | rs9641468 | 7 | 111401160 | T | C | 0.034040603 | 0.034040603 | -0.0153 | -0.013298028 | 0.00002886 | 0.00001597 | new | DOCK4 | intronic | . | . | 7q31.1 |
| GERD | MDD | rs12536335 | 7 | 114043159 | A | G | 0.002887837 | 0.002887837 | 0.0199 | -0.021795812 | 7.261E-09 | 7.946E-14 | known | NONE | intergenic | . | . | 7q31.1 |
| GERD | MDD | rs12537384 | 7 | 115025708 | T | C | 0.010319586 | 0.013177172 | -0.0174 | 0.013103769 | 0.000000493 | 0.00001057 | new | LINC01392 | intergenic | . | . | 7q31.2 |
| GERD | MDD | rs11770163 | 7 | 116417848 | C | G | 0.004536037 | 0.03144798 | 0.0205 | 0.011503579 | 7.744E-09 | 0.000104 | known | MET | intronic | . | . | 7q31.2 |
| GERD | MDD | rs1362000 | 7 | 126401715 | T | C | 0.002267811 | 0.009024632 | 0.0214 | 0.013804281 | 1.915E-09 | 0.000003477 | known | GRM8 | intronic | . | . | 7q31.33 |
| GERD | MDD | rs6952845 | 7 | 135088308 | A | C | 0.019396214 | 0.019396214 | -0.0157 | 0.015400797 | 0.000007373 | 2.847E-08 | new* | CNOT4 | intronic | . | . | 7q33 |
| GERD | MDD | rs79453306 | 7 | 140173170 | A | G | 0.037842175 | 0.037842175 | 0.0162 | -0.018897437 | 0.00008833 | 4.467E-09 | new* | MKRN1 | intronic | . | . | 7q34 |
| GERD | MDD | rs13233877 | 7 | 146681505 | A | G | 0.004031998 | 0.022611487 | -0.0218 | -0.012902885 | 6.687E-09 | 0.00004471 | known | CNTNAP2 | intronic | . | . | 7q35 |
| GERD | MDD | rs11774014 | 8 | 12698174 | A | C | 0.002795821 | 0.009449453 | -0.0349 | 0.022895877 | 6.484E-09 | 0.000003954 | known | LINC00681 | intergenic | . | . | 8p23.1 |
| GERD | MDD | rs10954826 | 8 | 31949764 | T | C | 0.023661161 | 0.026389433 | -0.017 | -0.011900531 | 0.000004039 | 0.00006058 | new | NRG1-IT1 | ncRNA_intronic | . | . | 8p12 |
| GERD | MDD | rs2658899 | 8 | 56325058 | T | C | 0.026546171 | 0.026546171 | 0.0153 | -0.014900463 | 0.0000211 | 8.046E-07 | new | XKR4 | intronic | . | . | 8q12.1 |
| GERD | MDD | rs2197803 | 8 | 59858040 | A | C | 0.030352379 | 0.033514343 | -0.0162 | 0.011503579 | 0.000008031 | 0.00009511 | new | TOX | intronic | . | . | 8q12.1 |
| GERD | MDD | rs1511031 | 8 | 60720759 | A | C | 0.022788304 | 0.032444492 | -0.0165 | -0.011202515 | 0.000002638 | 0.0001109 | new | CA8 | intergenic | . | . | 8q12.1 |
| GERD | MDD | rs10104523 | 8 | 73889570 | T | C | 0.005958203 | 0.006128269 | -0.0182 | -0.013500726 | 7.632E-08 | 0.000001217 | new | TERF1 | intergenic | . | . | 8q13.3 |
| GERD | MDD | rs10104596 | 8 | 87754626 | A | G | 0.03431585 | 0.03431585 | 0.0159 | 0.011602431 | 0.00001312 | 0.00007235 | new | CNGB3 | intronic | . | . | 8q21.3 |
| GERD | MDD | rs7841546 | 8 | 130103252 | A | G | 0.030419599 | 0.030419599 | -0.0163 | 0.015095488 | 0.00003632 | 0.000001163 | new | LINC00976 | intergenic | . | . | 8q24.21 |
| GERD | MDD | rs13263652 | 8 | 139346461 | T | C | 0.045652573 | 0.045652573 | 0.0153 | -0.014596005 | 0.000166 | 0.000001235 | new | FAM135B | intronic | . | . | 8q24.23 |
| GERD | MDD | rs13262595 | 8 | 143316970 | A | G | 0.026893015 | 0.026893015 | 0.0145 | 0.012896482 | 0.00002047 | 0.0000044 | new | TSNARE1 | intronic | . | . | 8q24.3 |
| GERD | MDD | rs12004755 | 9 | 14159091 | A | G | 0.018091369 | 0.019948625 | 0.0244 | 0.016503072 | 0.000002099 | 0.00003193 | new | NFIB | intronic | . | . | 9p23 |
| GERD | MDD | rs3793577 | 9 | 23737627 | A | G | 0.005983492 | 0.0071642 | -0.0185 | -0.014200349 | 7.741E-08 | 0.000001832 | new | ELAVL2 | intronic | . | . | 9p21.3 |
| GERD | MDD | rs13285448 | 9 | 31318709 | A | T | 0.021865991 | 0.021865991 | 0.0196 | -0.018296362 | 0.00001064 | 4.259E-07 | new | LINC01243 | intergenic | . | . | 9p21.1 |
| GERD | MDD | rs68026871 | 9 | 37479198 | T | G | 0.018002024 | 0.04959283 | -0.0274 | -0.015103486 | 5.297E-07 | 0.0003183 | new | POLR1E | intergenic | . | . | 9p13.2 |
| GERD | MDD | rs1328302 | 9 | 91835655 | T | C | 0.013189287 | 0.026818636 | 0.0278 | 0.019704583 | 4.924E-07 | 0.00006968 | new | SHC3 | intergenic | . | . | 9q22.2 |
| GERD | MDD | rs12349299 | 9 | 96174764 | T | C | 0.006973889 | 0.006973889 | -0.0283 | 0.023902056 | 1.249E-07 | 2.689E-08 | new* | FAM120AOS | intergenic | . | . | 9q22.31 |
| GERD | MDD | rs1484375 | 9 | 109067561 | A | G | 0.005396507 | 0.005396507 | 0.0231 | 0.017800625 | 5.555E-08 | 7.975E-08 | new | MIR8081 | intergenic | . | . | 9q31.2 |
| GERD | MDD | rs9696053 | 9 | 113429129 | T | G | 0.019937986 | 0.019937986 | 0.0165 | -0.014200349 | 0.00000517 | 0.00001244 | new | MUSK | intergenic | . | . | 9q31.3 |
| GERD | MDD | rs10817886 | 9 | 119170305 | T | C | 0.036891162 | 0.036891162 | -0.0161 | -0.016302161 | 0.00006289 | 0.000005986 | new | PAPPA | intergenic | . | . | 9q33.1 |
| GERD | MDD | rs10817972 | 9 | 119768572 | A | G | 0.044459994 | 0.044459994 | 0.0228 | 0.018802127 | 0.00007577 | 0.00002264 | new | ASTN2 | intronic | . | . | 9q33.1 |
| GERD | MDD | rs4837504 | 9 | 120521428 | A | G | 0.004329218 | 0.01103517 | -0.0196 | 0.012699025 | 2.535E-08 | 0.000006287 | known | TLR4 | intergenic | . | . | 9q33.1 |
| GERD | MDD | rs7032155 | 9 | 122672771 | A | C | 0.012473755 | 0.012473755 | 0.017 | -0.020896826 | 0.00000141 | 9.289E-13 | new* | LINC01613 | intergenic | . | . | 9q33.2 |
| GERD | MDD | rs740917 | 9 | 132954602 | A | G | 0.01358428 | 0.01358428 | -0.0173 | 0.013695783 | 0.000001485 | 0.000008409 | new | NCS1 | intronic | . | . | 9q34.11 |
| GERD | MDD | rs10906187 | 10 | 12673010 | A | G | 0.019947497 | 0.019947497 | -0.0153 | 0.015400797 | 0.000008159 | 2.168E-07 | new | CAMK1D | intronic | . | . | 10p13 |
| GERD | MDD | rs10828248 | 10 | 21824619 | A | G | 0.002511368 | 0.013432664 | -0.0219 | 0.013695783 | 8.924E-10 | 0.00001113 | known | MLLT10 | intronic | . | . | 10p12.31 |
| GERD | MDD | rs4746339 | 10 | 77656509 | A | T | 0.012217591 | 0.012217591 | -0.0194 | -0.016495303 | 0.000001318 | 2.125E-07 | new | LRMDA | intronic | . | . | 10q22.2 |
| GERD | MDD | rs10748872 | 10 | 83797900 | T | C | 0.041661836 | 0.047887125 | 0.0143 | 0.010702523 | 0.00001544 | 0.0002046 | new | NRG3 | intronic | . | . | 10q23.1 |
| GERD | MDD | rs535945 | 10 | 84501786 | A | G | 0.022665852 | 0.022665852 | 0.0161 | 0.012303994 | 0.000007482 | 0.00001517 | new | NRG3 | intronic | . | . | 10q23.1 |
| GERD | MDD | rs10736071 | 10 | 94675198 | T | G | 0.017811416 | 0.017811416 | -0.0175 | -0.014798967 | 0.000004681 | 0.000005707 | new | EXOC6 | intronic | . | . | 10q23.33 |
| GERD | MDD | rs10786736 | 10 | 104849116 | C | G | 0.002434738 | 0.0118787 | -0.0344 | -0.020498669 | 3.207E-09 | 0.000007915 | known | CNNM2 | UTR3 | . | . | 10q24.32 |
| GERD | MDD | rs11596214 | 10 | 106453832 | A | G | 0.00312819 | 0.00312819 | -0.0208 | -0.024098039 | 9.627E-09 | 6.583E-15 | known | SORCS3 | intronic | . | . | 10q25.1 |
| GERD | MDD | rs761777 | 10 | 134938075 | A | G | 0.002267811 | 0.002267811 | -0.0251 | 0.019998687 | 8.348E-10 | 1.032E-08 | known | ADGRA1 | intronic | . | . | 10q26.3 |
| GERD | MDD | rs12295734 | 11 | 13292281 | C | G | 0.025100495 | 0.042103268 | 0.0178 | -0.011900531 | 0.000002446 | 0.0002182 | new | ARNTL | intergenic | . | . | 11p15.2 |
| GERD | MDD | rs11023926 | 11 | 16373714 | A | C | 0.027755691 | 0.027755691 | 0.0205 | 0.019704583 | 0.00002511 | 0.000002673 | new | SOX6 | intronic | . | . | 11p15.1 |
| GERD | MDD | rs1509591 | 11 | 24593694 | A | G | 0.026782545 | 0.026782545 | 0.0143 | -0.014301785 | 0.00002183 | 8.245E-07 | new | LUZP2 | intronic | . | . | 11p14.3 |
| GERD | MDD | rs12419325 | 11 | 28580338 | T | G | 0.024723267 | 0.024723267 | 0.0198 | -0.023104874 | 0.00001606 | 3.44E-09 | new* | MIR8068 | intergenic | . | . | 11p14.1 |
| GERD | MDD | rs1415118 | 11 | 29863909 | T | G | 0.032084821 | 0.032084821 | -0.0143 | -0.013095371 | 0.00004436 | 0.00000345 | new | LINC01616 | intergenic | . | . | 11p14.1 |
| GERD | MDD | rs66743582 | 11 | 30603413 | T | C | 0.046058062 | 0.046058062 | 0.0138 | 0.013202463 | 0.0001705 | 0.000003072 | new | MPPED2 | intronic | . | . | 11p14.1 |
| GERD | MDD | rs4440995 | 11 | 31838128 | A | G | 0.001944423 | 0.002847118 | 0.029 | 0.018802127 | 1.871E-10 | 1.683E-07 | known | PAX6-AS1 | ncRNA_exonic | . | . | 11p13 |
| GERD | MDD | rs764669 | 11 | 32302799 | T | C | 0.017136692 | 0.036187816 | -0.0181 | -0.010899181 | 8.463E-07 | 0.0001482 | new | WT1 | intergenic | . | . | 11p13 |
| GERD | MDD | rs7928395 | 11 | 38581040 | T | C | 0.030144858 | 0.030144858 | -0.0148 | 0.011898926 | 0.00001467 | 0.00002396 | new | LINC02759 | intergenic | . | . | 11p12 |
| GERD | MDD | rs1187952 | 11 | 63790496 | A | G | 0.035868314 | 0.035868314 | 0.0147 | 0.013902905 | 0.00004545 | 0.000009513 | new | MACROD1 | intronic | . | . | 11q13.1 |
| GERD | MDD | rs1783541 | 11 | 65294799 | T | C | 0.012127239 | 0.012127239 | 0.0207 | 0.017102908 | 0.000001287 | 0.000001691 | new | SCYL1 | intronic | . | . | 11q13.1 |
| GERD | MDD | rs67446678 | 11 | 68227466 | T | C | 0.041334739 | 0.045251627 | 0.0217 | 0.016099701 | 0.00001653 | 0.0001712 | new | PPP6R3 | upstream | . | . | 11q13.2 |
| GERD | MDD | rs1432058 | 11 | 83242712 | T | C | 0.033663306 | 0.033663306 | -0.0131 | -0.014098924 | 0.00005361 | 0.000001063 | new | DLG2 | intronic | . | . | 11q14.1 |
| GERD | MDD | rs11234641 | 11 | 86120586 | T | C | 0.01591916 | 0.045963247 | -0.0219 | -0.012599036 | 3.539E-07 | 0.0002651 | new | CCDC81 | intronic | . | . | 11q14.2 |
| GERD | MDD | rs61901970 | 11 | 88318056 | T | C | 0.028545618 | 0.028545618 | 0.0185 | -0.019998647 | 0.00002813 | 1.513E-07 | new | GRM5 | intronic | . | . | 11q14.3 |
| GERD | MDD | rs72966168 | 11 | 90465045 | A | G | 0.045172928 | 0.045172928 | -0.0229 | -0.020304753 | 0.0000528 | 0.00004687 | new | DISC1FP1 | ncRNA_intronic | . | . | 11q14.3 |
| GERD | MDD | rs2513198 | 11 | 104458757 | T | C | 0.032980451 | 0.032980451 | -0.0148 | -0.012102946 | 0.00001934 | 0.00002588 | new | LINC02552 | ncRNA_intronic | . | . | 11q22.3 |
| GERD | MDD | rs4533068 | 11 | 112833618 | A | G | 0.002744059 | 0.002744059 | 0.0309 | 0.025297308 | 6.084E-09 | 4.62E-09 | known | LOC101928847 | ncRNA_intronic | . | . | 11q23.2 |
| GERD | MDD | rs6590700 | 11 | 133315491 | T | C | 0.041395142 | 0.041395142 | 0.0158 | -0.015702644 | 0.0001205 | 0.00000222 | new | OPCML | intronic | . | . | 11q25 |
| GERD | MDD | rs73034035 | 12 | 215121 | A | G | 0.004530249 | 0.015023809 | 0.0214 | 0.013301146 | 1.857E-08 | 0.00001508 | known | IQSEC3 | intronic | . | . | 12p13.33 |
| GERD | MDD | rs10846167 | 12 | 15515945 | T | C | 0.038828801 | 0.040286989 | 0.0157 | 0.011800104 | 0.00001536 | 0.0001278 | new | PTPRO | intronic | . | . | 12p12.3 |
| GERD | MDD | rs10846378 | 12 | 16743434 | T | G | 0.015582475 | 0.015582475 | 0.0168 | 0.013695783 | 0.000002746 | 0.000006598 | new | LMO3 | intronic | . | . | 12p12.3 |
| GERD | MDD | rs6581420 | 12 | 62115165 | T | C | 0.020998438 | 0.032225245 | -0.0165 | 0.010702523 | 0.000001846 | 0.0001105 | new | TAFA2 | intronic | . | . | 12q14.1 |
| GERD | MDD | rs11174358 | 12 | 62561299 | A | G | 0.011511507 | 0.011511507 | 0.0286 | 0.024204689 | 0.000001073 | 1.758E-07 | new | TAFA2 | intronic | . | . | 12q14.1 |
| GERD | MDD | rs6539788 | 12 | 84226327 | T | G | 0.022174252 | 0.022174252 | -0.016 | -0.011799339 | 0.000005917 | 0.00002031 | new | TMTC2 | intergenic | . | . | 12q21.31 |
| GERD | MDD | rs12367037 | 12 | 121303820 | A | G | 0.017141224 | 0.017141224 | 0.016 | 0.015597721 | 0.000004719 | 4.601E-07 | new | SPPL3 | intronic | . | . | 12q24.31 |
| GERD | MDD | rs35655684 | 13 | 31800257 | A | C | 0.033600618 | 0.033600618 | -0.0188 | -0.014301785 | 0.0000132 | 0.00006047 | new | B3GLCT | intronic | . | . | 13q12.3 |
| GERD | MDD | rs7984963 | 13 | 53812390 | A | G | 0.025364336 | 0.025364336 | -0.0146 | 0.015804449 | 0.00001768 | 1.859E-08 | new* | LINC01065 | intergenic | . | . | 13q14.3 |
| GERD | MDD | rs13378714 | 13 | 55070587 | A | G | 0.032043368 | 0.032043368 | -0.017 | -0.013500726 | 0.00003623 | 0.000005963 | new | MIR1297 | intergenic | . | . | 13q14.3 |
| GERD | MDD | rs1962922 | 13 | 58717996 | C | G | 0.00833961 | 0.00833961 | 0.0226 | 0.01949866 | 2.361E-07 | 9.144E-08 | new | LINC02338 | intergenic | . | . | 13q21.1 |
| GERD | MDD | rs9538483 | 13 | 60219426 | A | T | 0.003757197 | 0.028331005 | 0.0337 | 0.019998687 | 4.618E-09 | 0.00008061 | known | DIAPH3 | intergenic | . | . | 13q21.2 |
| GERD | MDD | rs9538607 | 13 | 60807867 | T | C | 0.021616952 | 0.021616952 | 0.0156 | 0.012896482 | 0.000005614 | 0.00001884 | new | LINC00434 | ncRNA_intronic | . | . | 13q21.2 |
| GERD | MDD | rs9540720 | 13 | 66922705 | A | G | 0.007775463 | 0.007775463 | -0.0185 | 0.014100124 | 1.735E-07 | 0.000001868 | new | PCDH9 | intronic | . | . | 13q21.32 |
| GERD | MDD | rs9571745 | 13 | 67843475 | A | C | 0.016728257 | 0.016728257 | -0.0164 | -0.012801592 | 0.000002267 | 0.00001733 | new | PCDH9 | intergenic | . | . | 13q21.32 |
| GERD | MDD | rs9318683 | 13 | 80849370 | A | G | 0.002789363 | 0.016585685 | -0.0212 | -0.012001734 | 1.692E-09 | 0.00001957 | known | SPRY2 | intergenic | . | . | 13q31.1 |
| GERD | MDD | rs4462482 | 13 | 85261917 | T | C | 0.038234023 | 0.038234023 | -0.0153 | -0.014504686 | 0.00009141 | 0.000001436 | new | LINC00333 | intergenic | . | . | 13q31.1 |
| GERD | MDD | rs4559792 | 13 | 92867011 | A | T | 0.043696756 | 0.043696756 | -0.0166 | 0.015499263 | 0.0001459 | 0.000003339 | new | GPC5 | intronic | . | . | 13q31.3 |
| GERD | MDD | rs9517313 | 13 | 99105892 | C | G | 0.033195262 | 0.033195262 | 0.0163 | 0.014297305 | 0.00005006 | 0.000003623 | new | FARP1 | UTR3 | . | . | 13q32.2 |
| GERD | MDD | rs2806306 | 13 | 100453482 | A | G | 0.033471253 | 0.046169335 | -0.017 | -0.011000282 | 0.000006858 | 0.0002288 | new | LOC101927437 | ncRNA_intronic | . | . | 13q32.3 |
| GERD | MDD | rs2231810 | 14 | 23826819 | T | C | 0.041359965 | 0.041359965 | 0.0227 | 0.018998382 | 0.00006249 | 0.00001781 | new | EFS | exonic | synonymous SNV | . | 14q11.2 |
| GERD | MDD | rs10132715 | 14 | 42182103 | A | G | 0.003653773 | 0.003653773 | -0.0185 | 0.015902876 | 1.592E-08 | 1.259E-08 | known | LRFN5 | intronic | . | . | 14q21.1 |
| GERD | MDD | rs57536190 | 14 | 49915830 | A | G | 0.043187737 | 0.043187737 | 0.0169 | 0.01400152 | 0.00005293 | 0.00003407 | new | RPS29 | intergenic | . | . | 14q21.3 |
| GERD | MDD | rs2274794 | 14 | 57283934 | T | C | 0.010755467 | 0.010755467 | -0.0184 | -0.014301785 | 7.927E-07 | 0.000004092 | new | OTX2-AS1 | ncRNA_intronic | . | . | 14q22.3 |
| GERD | MDD | rs61741172 | 14 | 69521345 | A | G | 0.011137061 | 0.013999396 | -0.0187 | -0.01360209 | 6.395E-07 | 0.00001248 | new | DCAF5 | exonic | synonymous SNV | . | 14q24.1 |
| GERD | MDD | rs7350725 | 14 | 72216675 | A | C | 0.030092619 | 0.038495771 | -0.016 | 0.011503579 | 0.000006108 | 0.0001466 | new | SIPA1L1 | intergenic | . | . | 14q24.2 |
| GERD | MDD | rs719732 | 14 | 75119057 | A | G | 0.008664867 | 0.008664867 | -0.0208 | -0.01789924 | 2.846E-07 | 1.751E-07 | new | AREL1 | intergenic | . | . | 14q24.3 |
| GERD | MDD | rs10129979 | 14 | 93953629 | A | G | 0.040834108 | 0.040834108 | -0.0153 | -0.012001734 | 0.00003883 | 0.00003622 | new | UNC79 | intronic | . | . | 14q32.12 |
| GERD | MDD | rs4906267 | 14 | 103305077 | T | G | 0.002811959 | 0.00459668 | 0.0271 | 0.018497855 | 6.614E-09 | 5.833E-07 | known | TRAF3 | intronic | . | . | 14q32.32 |
| GERD | MDD | rs638182 | 15 | 37658915 | T | C | 0.049073745 | 0.049073745 | 0.0121 | 0.014602857 | 0.000208 | 5.948E-07 | new | MEIS2 | intergenic | . | . | 15q14 |
| GERD | MDD | rs8035957 | 15 | 38838264 | T | C | 0.046137881 | 0.046137881 | -0.016 | 0.020096702 | 0.0001714 | 1.247E-10 | new* | RASGRP1 | intronic | . | . | 15q14 |
| GERD | MDD | rs76277863 | 15 | 40649863 | A | G | 0.040349145 | 0.040349145 | 0.0222 | -0.01939691 | 0.00004709 | 0.00002455 | new | DISP2 | upstream | . | . | 15q15.1 |
| GERD | MDD | rs34604744 | 15 | 51847442 | A | C | 0.026864453 | 0.032597408 | -0.0173 | 0.014198719 | 0.00000502 | 0.0000999 | new | DMXL2 | intronic | . | . | 15q21.2 |
| GERD | MDD | rs4776729 | 15 | 66399197 | A | G | 0.019640427 | 0.019640427 | -0.0225 | 0.022504855 | 0.000007734 | 7.82E-08 | new | MEGF11 | intronic | . | . | 15q22.31 |
| GERD | MDD | rs1452388 | 15 | 74139078 | A | G | 0.017463112 | 0.017463112 | -0.0218 | 0.017004599 | 0.000005049 | 0.000001299 | new | TBC1D21 | intergenic | . | . | 15q24.1 |
| GERD | MDD | rs919592 | 15 | 86926176 | T | C | 0.049197291 | 0.049197291 | -0.0134 | -0.014798967 | 0.0002097 | 0.000002362 | new | AGBL1 | intronic | . | . | 15q25.3 |
| GERD | MDD | rs150304 | 15 | 89949210 | A | G | 0.003441009 | 0.023551032 | 0.0202 | -0.011495825 | 3.925E-09 | 0.00005003 | known | LOC105371031 | ncRNA_intronic | . | . | 15q26.1 |
| GERD | MDD | rs12438687 | 15 | 96955776 | T | C | 0.003581603 | 0.003581603 | 0.0301 | 0.026797712 | 1.509E-08 | 5.259E-10 | known | NR2F2 | intergenic | . | . | 15q26.2 |
| GERD | MDD | rs12929878 | 16 | 412447 | A | G | 0.042654546 | 0.042654546 | 0.0164 | 0.012402767 | 0.00003105 | 0.00006935 | new | MRPL28 | intergenic | . | . | 16p13.3 |
| GERD | MDD | rs12921271 | 16 | 6639512 | A | G | 0.045995259 | 0.045995259 | -0.0135 | -0.012902885 | 0.0001036 | 0.00001519 | new | RBFOX1 | intronic | . | . | 16p13.3 |
| GERD | MDD | rs4433823 | 16 | 30433730 | A | G | 0.04793011 | 0.04793011 | 0.022 | 0.023296517 | 0.0001929 | 1.109E-07 | new | DCTPP1 | intergenic | . | . | 16p11.2 |
| GERD | MDD | rs11645288 | 16 | 51172677 | A | G | 0.034293552 | 0.034293552 | 0.0195 | 0.018703985 | 0.00005782 | 4.156E-08 | new* | SALL1 | exonic | synonymous SNV | . | 16q12.1 |
| GERD | MDD | rs62046253 | 16 | 60644621 | T | C | 0.002267811 | 0.003070995 | -0.022 | -0.015205013 | 1.99E-09 | 2.072E-07 | known | MIR4426 | intergenic | . | . | 16q21 |
| GERD | MDD | rs30895 | 16 | 63047663 | A | G | 0.009548422 | 0.009548422 | -0.0224 | 0.017800625 | 4.972E-07 | 0.000003386 | new | CDH8 | intergenic | . | . | 16q21 |
| GERD | MDD | rs2917692 | 16 | 64298023 | T | C | 0.003141144 | 0.02447939 | -0.0202 | 0.011997738 | 2.19E-09 | 0.00005561 | known | NONE | intergenic | . | . | 16q21 |
| GERD | MDD | rs739414 | 16 | 73097956 | T | C | 0.000944336 | 0.022236972 | -0.0302 | -0.01360209 | 1.928E-13 | 0.00004275 | known | ZFHX3 | intergenic | . | . | 16q22.3 |
| GERD | MDD | rs636425 | 16 | 77125714 | A | C | 0.002267811 | 0.005851616 | -0.0215 | 0.015203834 | 1.734E-09 | 0.00000108 | known | MON1B | intergenic | . | . | 16q23.1 |
| GERD | MDD | rs11150495 | 16 | 82868235 | A | T | 0.006321803 | 0.00958153 | -0.0182 | -0.013896105 | 9.312E-08 | 0.000004114 | new | CDH13 | intronic | . | . | 16q23.3 |
| GERD | MDD | rs1051322 | 17 | 1946503 | C | G | 0.028309131 | 0.028309131 | -0.0169 | 0.015804449 | 0.00001868 | 0.000008867 | new | DPH1 | UTR3 | . | . | 17p13.3 |
| GERD | MDD | rs150491398 | 17 | 8074039 | T | C | 0.005578642 | 0.005578642 | -0.0215 | -0.017695649 | 6.165E-08 | 1.129E-07 | new | TMEM107 | intergenic | . | . | 17p13.1 |
| GERD | MDD | rs17688916 | 17 | 43778680 | A | T | 0.02564401 | 0.02564401 | -0.017 | 0.015804449 | 0.000008879 | 0.00002426 | new | LINC02210-CRHR1 | intronic | . | . | 17q21.31 |
| GERD | MDD | rs62074125 | 17 | 44852612 | A | C | 0.019878869 | 0.019878869 | -0.0179 | -0.014900463 | 0.0000035 | 0.00002607 | new | WNT3 | intronic | . | . | 17q21.31 |
| GERD | MDD | rs9901869 | 17 | 45575206 | A | G | 0.025483668 | 0.034544453 | 0.0154 | 0.011701272 | 0.000003748 | 0.0001242 | new | MRPL45P2 | intergenic | . | . | 17q21.32 |
| GERD | MDD | rs11653507 | 17 | 65892064 | A | T | 0.009898515 | 0.009898515 | -0.0213 | 0.018399683 | 5.842E-07 | 9.243E-08 | new | BPTF | intronic | . | . | 17q24.2 |
| GERD | MDD | rs7503779 | 17 | 78920172 | A | G | 0.001810305 | 0.041522763 | -0.0239 | -0.011404788 | 5.697E-11 | 0.0002119 | known | RPTOR | intronic | . | . | 17q25.3 |
| GERD | MDD | rs619294 | 18 | 29819131 | A | G | 0.0429675 | 0.0429675 | 0.0148 | -0.013500726 | 0.00006145 | 0.00002514 | new | MEP1B | intergenic | . | . | 18q12.1 |
| GERD | MDD | rs11665070 | 18 | 35152563 | A | G | 0.001999966 | 0.001999966 | -0.0239 | 0.02180063 | 2.023E-10 | 2.302E-12 | known | CELF4 | intergenic | . | . | 18q12.2 |
| GERD | MDD | rs117763335 | 18 | 42260348 | T | C | 0.033528475 | 0.033528475 | -0.0169 | -0.017095295 | 0.00005275 | 6.282E-07 | new | SETBP1 | UTR5 | . | . | 18q12.3 |
| GERD | MDD | rs1026186 | 18 | 42741410 | A | G | 0.025391755 | 0.032359691 | -0.0165 | -0.011900531 | 0.000004072 | 0.0001027 | new | SLC14A2 | intergenic | . | . | 18q12.3 |
| GERD | MDD | rs11663217 | 18 | 50526533 | A | G | 0.003506534 | 0.003506534 | -0.0209 | 0.022201708 | 1.411E-08 | 1.061E-13 | known | DCC | intronic | . | . | 18q21.2 |
| GERD | MDD | rs4801102 | 18 | 52522841 | T | C | 0.025665534 | 0.038754714 | 0.0158 | 0.011098187 | 0.000003257 | 0.0001727 | new | RAB27B | intronic | . | . | 18q21.2 |
| GERD | MDD | rs7228159 | 18 | 53104253 | A | T | 0.001080338 | 0.00188856 | 0.0246 | 0.0161981 | 2.885E-12 | 3.437E-08 | known | TCF4 | intronic | . | . | 18q21.2 |
| GERD | MDD | rs12606645 | 18 | 53475163 | A | G | 0.038526698 | 0.038526698 | 0.014 | 0.015696168 | 0.00009378 | 1.565E-07 | new | LINC01415 | intergenic | . | . | 18q21.2 |
| GERD | MDD | rs8091998 | 18 | 77561978 | T | G | 0.022972896 | 0.022972896 | 0.0185 | 0.01660143 | 0.0000124 | 0.000003379 | new | CTDP1 | intergenic | . | . | 18q23 |
| GERD | MDD | rs3803916 | 19 | 18669987 | A | G | 0.000251166 | 0.039242794 | -0.036 | -0.015601066 | 1.053E-14 | 0.0001819 | known | KXD1 | intronic | . | . | 19p13.11 |
| GERD | MDD | rs12151059 | 19 | 30759421 | T | C | 0.031018278 | 0.031018278 | -0.0154 | -0.013703465 | 0.0000306 | 0.000006521 | new | ZNF536 | intronic | . | . | 19q12 |
| GERD | MDD | rs2111530 | 19 | 31891006 | A | G | 0.020006568 | 0.020006568 | -0.0149 | 0.017496048 | 0.000008226 | 3.375E-09 | new* | TSHZ3 | intergenic | . | . | 19q12 |
| GERD | MDD | rs11084707 | 19 | 33612873 | T | C | 0.032410829 | 0.032410829 | 0.0145 | 0.013301146 | 0.0000334 | 0.000007934 | new | GPATCH1 | intronic | . | . | 19q13.11 |
| GERD | MDD | rs6044120 | 20 | 16545951 | T | C | 0.03083292 | 0.03083292 | -0.0149 | -0.012801592 | 0.00002658 | 0.000008352 | new | KIF16B | intronic | . | . | 20p12.1 |
| GERD | MDD | rs6141302 | 20 | 30984239 | T | G | 0.028349999 | 0.028349999 | 0.014 | 0.013804281 | 0.00002735 | 0.000002046 | new | ASXL1 | intronic | . | . | 20q11.21 |
| GERD | MDD | rs6030812 | 20 | 42001011 | A | G | 0.045698164 | 0.045698164 | -0.0159 | 0.018301501 | 0.0001665 | 3.975E-07 | new | SRSF6 | intergenic | . | . | 20q13.11 |
| GERD | MDD | rs4809370 | 20 | 62470872 | T | C | 0.005455821 | 0.028988957 | -0.0205 | -0.011404788 | 1.549E-08 | 0.00008584 | known | ZBTB46 | intergenic | . | . | 20q13.33 |
| GERD | MDD | rs2824419 | 21 | 19025739 | T | C | 0.04636615 | 0.04636615 | 0.0154 | -0.017502275 | 0.000174 | 0.000001108 | new | BTG3-AS1 | intergenic | . | . | 21q21.1 |
| GERD | MDD | rs62211552 | 21 | 35888237 | T | C | 0.025676764 | 0.025676764 | 0.0186 | -0.014697481 | 0.000007743 | 0.00003277 | new | RCAN1 | downstream | . | . | 21q22.12 |
| GERD | MDD | rs8133276 | 21 | 45126629 | T | C | 0.031718397 | 0.031718397 | -0.0142 | -0.013298028 | 0.00003872 | 0.00000451 | new | RRP1B | intergenic | . | . | 21q22.3 |
| GERD | MDD | rs2838762 | 21 | 46487831 | A | C | 0.021484349 | 0.021484349 | 0.0153 | 0.013103769 | 0.000008066 | 0.000008382 | new | SSR4P1 | intergenic | . | . | 21q22.3 |
| GERD | MDD | rs861825 | 22 | 22046318 | T | C | 0.031494681 | 0.038426422 | 0.0198 | 0.013695783 | 0.000007474 | 0.0001387 | new | PPIL2 | intronic | . | . | 22q11.21 |
| GERD | MDD | rs4470425 | 22 | 40716488 | A | C | 0.002267811 | 0.00388542 | 0.0235 | 0.01530232 | 8.263E-10 | 3.818E-07 | known | TNRC6B | intronic | . | . | 22q13.1 |
| GERD | MDD | rs11090045 | 22 | 41753603 | A | G | 0.004453299 | 0.004453299 | 0.0212 | 0.016099701 | 2.958E-08 | 1.293E-07 | known | ZC3H7B | UTR3 | . | . | 22q13.2 |
| GERD | MDD | rs11090844 | 22 | 48874685 | A | G | 0.037365421 | 0.037365421 | -0.0161 | 0.012600282 | 0.00004488 | 0.00001454 | new | TAFA5 | intergenic | . | . | 22q13.32 |
| GERD | MDD | rs11703673 | 22 | 50652427 | T | C | 0.026756079 | 0.036631971 | 0.0164 | 0.011602431 | 0.000004121 | 0.0001419 | new | SELENOO | intronic | . | . | 22q13.33 |
| GERD | PTSD | rs1889853 | 1 | 8415290 | A | C | 0.010488354 | 0.026140967 | 0.0217 | 0.0197 | 4.762E-07 | 0.0001995 | new | RERE | intronic | . | . | 1p36.23 |
| GERD | PTSD | rs3766823 | 1 | 32197257 | A | G | 0.012143798 | 0.028156046 | 0.0229 | 0.0203 | 0.000001 | 0.0002352 | new | ADGRB2 | intronic | . | . | 1p35.2 |
| GERD | PTSD | rs1395455 | 1 | 37185190 | A | G | 0.006121603 | 0.011634371 | 0.0204 | 0.019 | 5.236E-08 | 0.00003611 | new | GRIK3 | intergenic | . | . | 1p34.3 |
| GERD | PTSD | rs6662282 | 1 | 38595864 | A | G | 0.04395226 | 0.04395226 | -0.0162 | -0.0169 | 0.00009273 | 0.000175 | new | MIR3659HG | intergenic | . | . | 1p34.3 |
| GERD | PTSD | rs11587148 | 1 | 50994712 | T | C | 0.018697459 | 0.037130424 | -0.0178 | -0.0173 | 0.000002682 | 0.0003774 | new | FAF1 | intronic | . | . | 1p32.3 |
| GERD | PTSD | rs2503185 | 1 | 66461401 | A | G | 0.004652931 | 0.023822175 | 0.0219 | 0.016 | 7.154E-10 | 0.000164 | known | PDE4B | intronic | . | . | 1p31.3 |
| GERD | PTSD | rs61765555 | 1 | 73957815 | T | C | 0.00711072 | 0.007603289 | -0.0218 | -0.0218 | 1.048E-07 | 0.0000153 | new | LINC01360 | intergenic | . | . | 1p31.1 |
| GERD | PTSD | rs514070 | 1 | 88718155 | A | G | 0.017556609 | 0.034991426 | -0.0202 | -0.0187 | 0.000002337 | 0.0003401 | new | PKN2-AS1 | intergenic | . | . | 1p22.2 |
| GERD | PTSD | rs197419 | 1 | 112315450 | A | C | 0.019916605 | 0.020559013 | -0.016 | -0.0173 | 0.000008935 | 0.00008361 | new | KCND3 | UTR3 | . | . | 1p13.2 |
| GERD | PTSD | rs10919095 | 1 | 169180857 | T | C | 0.011768662 | 0.015180236 | -0.0173 | -0.0168 | 0.000001924 | 0.00005866 | new | NME7 | intronic | . | . | 1q24.2 |
| GERD | PTSD | rs10920936 | 1 | 191377664 | T | G | 0.006446289 | 0.006446289 | 0.0195 | 0.025 | 6.705E-08 | 1.244E-08 | new* | LINC01680 | intergenic | . | . | 1q31.2 |
| GERD | PTSD | rs12131852 | 1 | 214433323 | C | G | 0.017425638 | 0.042208971 | 0.0179 | 0.0164 | 0.000001626 | 0.0005711 | new | SMYD2 | intergenic | . | . | 1q32.3 |
| GERD | PTSD | rs67446571 | 2 | 22554648 | A | G | 0.01428936 | 0.01428936 | -0.0158 | -0.0282 | 0.000003402 | 7.95E-11 | new* | LINC01884 | intergenic | . | . | 2p24.1 |
| GERD | PTSD | rs786404 | 2 | 44710780 | T | C | 0.030382598 | 0.030382598 | -0.0197 | -0.0234 | 0.00004845 | 0.00005299 | new | CAMKMT | intronic | . | . | 2p21 |
| GERD | PTSD | rs11691352 | 2 | 59949495 | A | G | 0.019709704 | 0.021607353 | -0.0175 | -0.0176 | 0.000008192 | 0.00009569 | new | LINC01793 | intergenic | . | . | 2p16.1 |
| GERD | PTSD | rs359258 | 2 | 60486157 | T | C | 0.010852828 | 0.012867074 | -0.018 | -0.0179 | 0.000001544 | 0.00004397 | new | MIR4432HG | intergenic | . | . | 2p16.1 |
| GERD | PTSD | rs6753453 | 2 | 100500887 | A | C | 0.001030235 | 0.015713667 | -0.0246 | -0.0171 | 4.932E-12 | 0.00006508 | known | AFF3 | intronic | . | . | 2q11.2 |
| GERD | PTSD | rs4851300 | 2 | 100958804 | A | C | 0.013571863 | 0.031895289 | -0.0185 | -0.0162 | 0.000001176 | 0.0003022 | new | LONRF2 | intergenic | . | . | 2q11.2 |
| GERD | PTSD | rs11678150 | 2 | 109824790 | A | T | 0.006287508 | 0.014122665 | -0.0197 | -0.0182 | 6.104E-08 | 0.00005262 | new | SH3RF3 | intronic | . | . | 2q12.3 |
| GERD | PTSD | rs2312459 | 2 | 113001329 | A | G | 0.013421016 | 0.035882506 | -0.0179 | -0.0165 | 0.000000925 | 0.0003955 | new | ZC3H8 | intronic | . | . | 2q13 |
| GERD | PTSD | rs2163249 | 2 | 124980406 | T | C | 0.034312087 | 0.034312087 | 0.0141 | 0.0189 | 0.00007621 | 0.000006654 | new | CNTNAP5 | intronic | . | . | 2q14.3 |
| GERD | PTSD | rs6735244 | 2 | 138286223 | A | G | 0.025976641 | 0.026643039 | -0.0145 | -0.0163 | 0.00001687 | 0.0001271 | new | THSD7B | intronic | . | . | 2q22.1 |
| GERD | PTSD | rs35125029 | 2 | 144261657 | A | G | 0.007981723 | 0.007981723 | -0.018 | -0.0221 | 2.911E-07 | 6.958E-07 | new | ARHGAP15 | intronic | . | . | 2q22.3 |
| GERD | PTSD | rs16844866 | 2 | 161063407 | T | C | 0.046926117 | 0.046926117 | -0.0177 | -0.0207 | 0.0001172 | 0.0001632 | new | ITGB6 | intergenic | . | . | 2q24.2 |
| GERD | PTSD | rs17755617 | 2 | 161935424 | A | C | 0.01785408 | 0.01785408 | -0.0171 | -0.0222 | 0.000007886 | 0.000002165 | new | TANK | intergenic | . | . | 2q24.2 |
| GERD | PTSD | rs9287989 | 2 | 176717741 | T | C | 0.003255614 | 0.013104641 | 0.0216 | 0.0173 | 2.838E-10 | 0.00004549 | known | LNPK | intergenic | . | . | 2q31.1 |
| GERD | PTSD | rs9881798 | 3 | 16846967 | A | C | 0.01279734 | 0.01279734 | -0.0173 | -0.02 | 0.000002422 | 0.000002532 | new | PLCL2 | intergenic | . | . | 3p24.3 |
| GERD | PTSD | rs17075317 | 3 | 43250443 | A | G | 0.008580318 | 0.013782298 | 0.0227 | 0.0222 | 6.778E-07 | 0.00005012 | known | SNRK | intergenic | . | . | 3p22.1 |
| GERD | PTSD | rs1858828 | 3 | 50242387 | T | G | 0.003372912 | 0.014277751 | 0.0214 | 0.0177 | 3.491E-10 | 0.0000538 | known | SLC38A3 | upstream | . | . | 3p21.31 |
| GERD | PTSD | rs352140 | 3 | 52256697 | T | C | 0.014544349 | 0.018827991 | -0.0156 | -0.0167 | 0.000002948 | 0.00008081 | new | TLR9 | exonic | synonymous SNV | . | 3p21.2 |
| GERD | PTSD | rs73094902 | 3 | 71603361 | T | C | 0.015616271 | 0.015616271 | -0.016 | -0.0188 | 0.000004633 | 0.00003445 | new | FOXP1 | intronic | . | . | 3p13 |
| GERD | PTSD | rs13319705 | 3 | 104693252 | T | C | 0.015091422 | 0.015091422 | -0.0173 | -0.0194 | 0.000004092 | 0.00002205 | new | ALCAM | intergenic | . | . | 3q13.11 |
| GERD | PTSD | rs9824903 | 3 | 117621537 | T | C | 0.028435148 | 0.032759569 | -0.0172 | -0.0168 | 0.0000177 | 0.0002094 | new | LINC02024 | intergenic | . | . | 3q13.32 |
| GERD | PTSD | rs73876550 | 3 | 163775972 | T | C | 0.031189563 | 0.031189563 | -0.018 | -0.0238 | 0.00005349 | 0.00001002 | new | MIR1263 | intergenic | . | . | 3q26.1 |
| GERD | PTSD | rs1479175 | 3 | 181208277 | A | T | 0.036153028 | 0.036153028 | 0.0169 | 0.0223 | 0.00007874 | 0.00007539 | new | SOX2-OT | ncRNA_intronic | . | . | 3q26.33 |
| GERD | PTSD | rs10027061 | 4 | 61467972 | A | G | 0.022791714 | 0.047768463 | -0.0184 | -0.0174 | 0.000003451 | 0.0006521 | new | MIR548AG1 | intergenic | . | . | 4q13.1 |
| GERD | PTSD | rs35041220 | 4 | 67856704 | T | C | 0.019511203 | 0.037642562 | -0.0161 | -0.0163 | 0.000003091 | 0.0003807 | new | LOC101927237 | intergenic | . | . | 4q13.2 |
| GERD | PTSD | rs114725911 | 4 | 80766564 | A | G | 0.037513885 | 0.037513885 | 0.0178 | 0.0229 | 0.0000519 | 0.0001795 | new | PCAT4 | ncRNA_intronic | . | . | 4q21.21 |
| GERD | PTSD | rs62319060 | 4 | 96145415 | T | C | 0.017357355 | 0.043002479 | -0.0178 | -0.0152 | 0.000001547 | 0.000594 | new | UNC5C | intronic | . | . | 4q22.3 |
| GERD | PTSD | rs4864365 | 4 | 137733154 | A | G | 0.021995685 | 0.029336228 | -0.0158 | -0.0169 | 0.000008104 | 0.0001976 | new | LINC02511 | ncRNA_intronic | . | . | 4q28.3 |
| GERD | PTSD | rs6840410 | 4 | 138651769 | A | G | 0.015049656 | 0.049473098 | -0.0256 | -0.0214 | 5.334E-07 | 0.0008007 | new | LINC02172 | intergenic | . | . | 4q28.3 |
| GERD | PTSD | rs1510719 | 4 | 140938116 | T | C | 0.001002019 | 0.001002019 | 0.0244 | 0.0239 | 5.022E-12 | 4.27E-08 | known | MAML3 | intronic | . | . | 4q31.1 |
| GERD | PTSD | rs7655694 | 4 | 164519194 | T | C | 0.043917928 | 0.043917928 | 0.0137 | 0.019 | 0.0001529 | 0.00001692 | new | MARCHF1 | intronic | . | . | 4q32.3 |
| GERD | PTSD | rs17206022 | 5 | 7333533 | T | G | 0.016832153 | 0.033178093 | -0.0165 | -0.0158 | 0.000002199 | 0.0003072 | new | LINC02123 | intergenic | . | . | 5p15.31 |
| GERD | PTSD | rs4502841 | 5 | 30863015 | A | G | 0.016185185 | 0.016185185 | 0.016 | 0.0209 | 0.000005367 | 7.409E-07 | new | CDH6 | intergenic | . | . | 5p13.3 |
| GERD | PTSD | rs1482698 | 5 | 44539453 | C | G | 0.042744452 | 0.042744452 | 0.0139 | 0.0196 | 0.0001432 | 0.00001294 | new | LINC02224 | intergenic | . | . | 5p12 |
| GERD | PTSD | rs10941692 | 5 | 45099778 | T | C | 0.003165027 | 0.014834756 | 0.0283 | 0.0213 | 1.878E-10 | 0.00005826 | known | HCN1 | intergenic | . | . | 5p12 |
| GERD | PTSD | rs16879569 | 5 | 51573487 | C | G | 0.015810629 | 0.020580632 | -0.0152 | -0.0169 | 0.000003594 | 0.00009554 | new | LINC02118 | intergenic | . | . | 5q11.2 |
| GERD | PTSD | rs987960 | 5 | 61469379 | T | C | 0.006121603 | 0.013873773 | -0.027 | -0.0245 | 3.897E-08 | 0.00005078 | known | KIF2A | intergenic | . | . | 5q12.1 |
| GERD | PTSD | rs288194 | 5 | 107318454 | T | C | 0.013347675 | 0.021034297 | -0.016 | -0.0163 | 0.00000196 | 0.0001186 | new | FBXL17 | intronic | . | . | 5q21.3 |
| GERD | PTSD | rs17480154 | 5 | 123122388 | T | C | 0.045612182 | 0.048400913 | 0.0202 | 0.0228 | 0.00006886 | 0.0003688 | new | CSNK1G3 | intergenic | . | . | 5q23.2 |
| GERD | PTSD | rs13169282 | 5 | 144256718 | A | G | 0.033543966 | 0.040585462 | -0.0168 | 0.0186 | 0.00002439 | 0.0003157 | new | KCTD16 | intergenic | . | . | 5q31.3 |
| GERD | PTSD | rs6556570 | 5 | 155812596 | C | G | 0.036323136 | 0.036323136 | -0.0156 | -0.0239 | 0.00009078 | 2.273E-07 | new | SGCD | intronic | . | . | 5q33.3 |
| GERD | PTSD | rs6900444 | 6 | 31241182 | T | C | 3.01704E-05 | 0.003817744 | 0.0272 | 0.0201 | 1.564E-14 | 0.000003066 | known | HLA-C | intergenic | . | . | 6p21.33 |
| GERD | PTSD | rs1337898 | 6 | 58053857 | T | C | 0.03565152 | 0.0383418 | 0.0145 | 0.0166 | 0.00003495 | 0.0002564 | new | GUSBP4 | intergenic | . | . | 6p11.2 |
| GERD | PTSD | rs9351261 | 6 | 65171419 | T | C | 0.004821538 | 0.009680117 | 0.0224 | 0.0197 | 2.962E-09 | 0.00002508 | known | EYS | intronic | . | . | 6q12 |
| GERD | PTSD | rs1933720 | 6 | 98297832 | T | C | 0.007152277 | 0.048269218 | 0.0228 | 0.0156 | 1.957E-09 | 0.0007596 | known | LOC101927314 | intergenic | . | . | 6q16.1 |
| GERD | PTSD | rs17657328 | 6 | 101614312 | T | C | 0.032559513 | 0.047465802 | 0.0159 | 0.015 | 0.0000162 | 0.0005148 | new | GRIK2 | intergenic | . | . | 6q16.3 |
| GERD | PTSD | rs4870060 | 6 | 152231404 | A | G | 0.004927723 | 0.004927723 | -0.0213 | -0.028 | 8.368E-09 | 3.205E-10 | known | ESR1 | intronic | . | . | 6q25.1 |
| GERD | PTSD | rs11766944 | 7 | 1888051 | A | G | 0.00073966 | 0.00073966 | -0.0301 | -0.0348 | 1.917E-12 | 1.722E-11 | known | MAD1L1 | intronic | . | . | 7p22.3 |
| GERD | PTSD | rs73035954 | 7 | 3563804 | T | C | 0.005181743 | 0.008500506 | -0.0233 | -0.0207 | 1.007E-08 | 0.00001915 | known | SDK1 | intronic | . | . | 7p22.2 |
| GERD | PTSD | rs1922525 | 7 | 70766925 | A | T | 0.042667568 | 0.044799544 | 0.015 | -0.0161 | 0.00005927 | 0.0003151 | new | GALNT17 | intronic | . | . | 7q11.22 |
| GERD | PTSD | rs2189246 | 7 | 82444372 | A | G | 0.007438804 | 0.007438804 | 0.0185 | 0.0227 | 1.421E-07 | 6.442E-08 | new | PCLO | intronic | . | . | 7q21.11 |
| GERD | PTSD | rs4729021 | 7 | 91758422 | A | C | 0.028580295 | 0.040099632 | -0.0143 | -0.015 | 0.00001364 | 0.0003623 | new | CYP51A1 | intronic | . | . | 7q21.2 |
| GERD | PTSD | rs12536335 | 7 | 114043159 | A | G | 0.004821538 | 0.004821538 | 0.0199 | 0.0284 | 7.261E-09 | 1.154E-10 | known | NONE | intergenic | . | . | 7q31.1 |
| GERD | PTSD | rs12537384 | 7 | 115025708 | T | C | 0.012477387 | 0.038431824 | -0.0174 | -0.0151 | 0.000000493 | 0.0004712 | new | LINC01392 | intergenic | . | . | 7q31.2 |
| GERD | PTSD | rs11770163 | 7 | 116417848 | C | G | 0.004821538 | 0.004821538 | 0.0205 | 0.0243 | 7.744E-09 | 1.247E-07 | known | MET | intronic | . | . | 7q31.2 |
| GERD | PTSD | rs13225829 | 7 | 126403584 | C | G | 0.006074504 | 0.038505801 | -0.0214 | -0.0154 | 6.797E-10 | 0.0004736 | known | GRM8 | intronic | . | . | 7q31.33 |
| GERD | PTSD | rs6952845 | 7 | 135088308 | A | C | 0.026438036 | 0.046524256 | -0.0157 | -0.0145 | 0.000007373 | 0.0005723 | new | CNOT4 | intronic | . | . | 7q33 |
| GERD | PTSD | rs79453306 | 7 | 140173170 | A | G | 0.035960767 | 0.035960767 | 0.0162 | 0.0216 | 0.00008833 | 0.00001684 | new | MKRN1 | intronic | . | . | 7q34 |
| GERD | PTSD | rs579905 | 8 | 60889903 | C | G | 0.031889704 | 0.039384997 | -0.0152 | -0.0158 | 0.00002097 | 0.0003055 | new | CA8 | intergenic | . | . | 8q12.1 |
| GERD | PTSD | rs9324542 | 8 | 142227854 | A | T | 0.023360185 | 0.023360185 | 0.0152 | 0.0184 | 0.00001748 | 0.00003062 | new | SLC45A4 | intronic | . | . | 8q24.3 |
| GERD | PTSD | rs13262595 | 8 | 143316970 | A | G | 0.024494206 | 0.024494206 | 0.0145 | 0.0207 | 0.00002047 | 0.000001009 | new | TSNARE1 | intronic | . | . | 8q24.3 |
| GERD | PTSD | rs7034703 | 9 | 14461188 | C | G | 0.03994435 | 0.03994435 | -0.0141 | -0.0202 | 0.0001193 | 0.000002686 | new | NFIB | intronic | . | . | 9p22.3 |
| GERD | PTSD | rs7470094 | 9 | 31233264 | T | C | 0.015078813 | 0.034546412 | 0.0193 | 0.017 | 0.000001433 | 0.0003608 | new | LINC01243 | intergenic | . | . | 9p21.1 |
| GERD | PTSD | rs10983764 | 9 | 120506307 | C | G | 0.008268453 | 0.026202156 | -0.0188 | -0.0158 | 5.757E-08 | 0.0002005 | new | TLR4 | intergenic | . | . | 9q33.1 |
| GERD | PTSD | rs7032155 | 9 | 122672771 | A | C | 0.010511374 | 0.010511374 | 0.017 | 0.0211 | 0.00000141 | 0.000002909 | new | LINC01613 | intergenic | . | . | 9q33.2 |
| GERD | PTSD | rs78897470 | 9 | 126349963 | T | G | 0.016513705 | 0.030132982 | -0.0235 | -0.0231 | 0.000002412 | 0.0002463 | new | DENND1A | intronic | . | . | 9q33.3 |
| GERD | PTSD | rs870811 | 9 | 132969084 | A | C | 0.013920639 | 0.013920639 | 0.0164 | 0.0191 | 0.000003125 | 0.000009394 | new | NCS1 | intronic | . | . | 9q34.11 |
| GERD | PTSD | rs2183271 | 10 | 21957229 | T | C | 0.005406961 | 0.01629988 | -0.0213 | -0.0194 | 1.02E-08 | 0.00006986 | known | MLLT10 | intronic | . | . | 10p12.31 |
| GERD | PTSD | rs1359033 | 10 | 77636083 | T | C | 0.01122604 | 0.018326299 | -0.0191 | -0.0188 | 0.000001332 | 0.00008937 | new | LRMDA | intronic | . | . | 10q22.2 |
| GERD | PTSD | rs2806446 | 10 | 134948249 | A | G | 0.004855869 | 0.028352282 | 0.0247 | 0.0181 | 6.733E-10 | 0.000239 | known | ADGRA1 | intergenic | . | . | 10q26.3 |
| GERD | PTSD | rs12419325 | 11 | 28580338 | T | G | 0.027185481 | 0.030937453 | 0.0198 | 0.0214 | 0.00001606 | 0.0001869 | new | MIR8068 | intergenic | . | . | 11p14.1 |
| GERD | PTSD | rs4440995 | 11 | 31838128 | A | G | 0.003164929 | 0.00376163 | 0.029 | 0.0255 | 1.871E-10 | 0.000002837 | known | PAX6-AS1 | ncRNA_exonic | . | . | 11p13 |
| GERD | PTSD | rs61901965 | 11 | 88287811 | A | G | 0.027743529 | 0.027743529 | 0.0189 | 0.0242 | 0.00003081 | 0.00006783 | new | GRM5 | intronic | . | . | 11q14.2 |
| GERD | PTSD | rs10789931 | 11 | 112842773 | T | C | 0.004864142 | 0.004864142 | 0.0299 | 0.0284 | 7.989E-09 | 0.00000319 | known | NCAM1 | intronic | . | . | 11q23.2 |
| GERD | PTSD | rs1866767 | 11 | 134274763 | T | C | 0.011658526 | 0.032134345 | 0.0269 | 0.0233 | 5.352E-07 | 0.000307 | new | B3GAT1 | intronic | . | . | 11q25 |
| GERD | PTSD | rs2160515 | 12 | 16753965 | A | G | 0.017780588 | 0.017780588 | 0.0162 | 0.0219 | 0.000007793 | 0.000001092 | new | LMO3 | intronic | . | . | 12p12.3 |
| GERD | PTSD | rs7953894 | 12 | 123404966 | A | C | 0.006628407 | 0.023518874 | 0.0211 | 0.0169 | 1.019E-08 | 0.0001591 | known | ABCB9 | downstream | . | . | 12q24.31 |
| GERD | PTSD | rs4883696 | 13 | 54028549 | C | G | 0.027078341 | 0.027078341 | 0.0149 | 0.0201 | 0.0000306 | 0.000002918 | new | LINC01065 | intergenic | . | . | 13q14.3 |
| GERD | PTSD | rs1348675 | 13 | 58712638 | A | G | 0.012030928 | 0.040940263 | 0.0215 | 0.0175 | 2.624E-07 | 0.0005363 | new | LINC02338 | intergenic | . | . | 13q21.1 |
| GERD | PTSD | rs12435216 | 14 | 40534511 | A | C | 0.020824914 | 0.046422963 | 0.0159 | 0.0162 | 0.000002549 | 0.0006393 | new | FBXO33 | intergenic | . | . | 14q21.1 |
| GERD | PTSD | rs10141927 | 14 | 42169777 | T | G | 0.016816939 | 0.018543653 | 0.0151 | 0.0203 | 0.000005417 | 0.00007156 | new | LRFN5 | intronic | . | . | 14q21.1 |
| GERD | PTSD | rs8010469 | 14 | 58643368 | A | G | 0.038582099 | 0.038582099 | 0.0155 | 0.0169 | 0.00005894 | 0.0001721 | new | ACTR10 | intergenic | . | . | 14q23.1 |
| GERD | PTSD | rs1275420 | 14 | 69620920 | A | G | 0.009889337 | 0.011960849 | 0.0185 | 0.0188 | 0.000001195 | 0.0000381 | new | DCAF5 | intergenic | . | . | 14q24.1 |
| GERD | PTSD | rs56330734 | 14 | 103292150 | A | G | 0.004821538 | 0.004821538 | 0.0272 | 0.0296 | 7.321E-09 | 1.731E-07 | known | TRAF3 | intronic | . | . | 14q32.32 |
| GERD | PTSD | rs113726301 | 16 | 6345251 | A | G | 0.026880327 | 0.027075032 | -0.018 | -0.0191 | 0.00001873 | 0.0001277 | new | RBFOX1 | intronic | . | . | 16p13.3 |
| GERD | PTSD | rs4624174 | 16 | 6633822 | T | C | 0.043978398 | 0.043978398 | 0.0129 | 0.0168 | 0.0001441 | 0.00006896 | new | RBFOX1 | intronic | . | . | 16p13.3 |
| GERD | PTSD | rs11861013 | 16 | 9163309 | C | G | 0.014281144 | 0.036123354 | 0.0228 | 0.0201 | 0.000001153 | 0.0004021 | new | C16orf72 | intergenic | . | . | 16p13.2 |
| GERD | PTSD | rs11645288 | 16 | 51172677 | A | G | 0.031875738 | 0.031875738 | 0.0195 | 0.021 | 0.00005782 | 0.00004296 | new | SALL1 | exonic | synonymous SNV | . | 16q12.1 |
| GERD | PTSD | rs62046243 | 16 | 60627007 | A | G | 0.004821538 | 0.011212249 | -0.0211 | -0.0182 | 4.913E-09 | 0.00003369 | known | MIR4426 | intergenic | . | . | 16q21 |
| GERD | PTSD | rs62043630 | 16 | 77469711 | A | G | 0.004826392 | 0.028463757 | 0.0212 | 0.0163 | 4.366E-10 | 0.000241 | known | ADAMTS18 | upstream | . | . | 16q23.1 |
| GERD | PTSD | rs11651453 | 17 | 8103695 | A | G | 0.010505987 | 0.030926409 | 0.0215 | 0.0187 | 1.979E-07 | 0.0002835 | new | AURKB | intergenic | . | . | 17p13.1 |
| GERD | PTSD | rs55797987 | 17 | 27962197 | A | G | 0.048393186 | 0.048393186 | -0.013 | 0.017 | 0.000143 | 0.0001328 | new | SSH2 | intronic | . | . | 17q11.2 |
| GERD | PTSD | rs199531 | 17 | 44830414 | T | C | 0.010650145 | 0.010650145 | 0.0182 | -0.0233 | 0.000001463 | 0.000008317 | new | NSF | intronic | . | . | 17q21.31 |
| GERD | PTSD | rs11867396 | 17 | 61697037 | T | C | 0.014940753 | 0.017741982 | 0.0186 | 0.0195 | 0.000003599 | 0.00006853 | new | MAP3K3 | intergenic | . | . | 17q23.3 |
| GERD | PTSD | rs7223643 | 17 | 65830282 | A | G | 0.013423816 | 0.013423816 | -0.0206 | -0.0257 | 0.000002787 | 0.000001944 | new | BPTF | intronic | . | . | 17q24.2 |
| GERD | PTSD | rs9964724 | 18 | 35159124 | T | C | 0.0035499 | 0.013688199 | -0.0233 | -0.0185 | 7.043E-10 | 0.00004945 | known | CELF4 | intergenic | . | . | 18q12.2 |
| GERD | PTSD | rs269982 | 18 | 42785329 | A | G | 0.008765284 | 0.008765284 | -0.0183 | -0.0227 | 7.491E-07 | 0.000002305 | new | SLC14A2 | intronic | . | . | 18q12.3 |
| GERD | PTSD | rs62097900 | 18 | 50526808 | T | C | 0.005748065 | 0.011632685 | 0.0209 | 0.0176 | 1.544E-08 | 0.0000361 | known | DCC | intronic | . | . | 18q21.2 |
| GERD | PTSD | rs7228159 | 18 | 53104253 | A | T | 0.000840951 | 0.012662209 | 0.0246 | 0.0184 | 2.885E-12 | 0.0000427 | known | TCF4 | intronic | . | . | 18q21.2 |
| GERD | PTSD | rs12606645 | 18 | 53475163 | A | G | 0.036753823 | 0.036753823 | 0.014 | -0.0186 | 0.00009378 | 0.00003975 | new | LINC01415 | intergenic | . | . | 18q21.2 |
| GERD | PTSD | rs7408312 | 19 | 18438507 | T | G | 0.001134266 | 0.010816805 | -0.0244 | -0.0186 | 7.553E-12 | 0.00003157 | known | LSM4 | intergenic | . | . | 19p13.11 |
| GERD | PTSD | rs12151059 | 19 | 30759421 | T | C | 0.037731245 | 0.046901956 | -0.0154 | -0.0159 | 0.0000306 | 0.0004264 | new | ZNF536 | intronic | . | . | 19q12 |
| GERD | PTSD | rs2111530 | 19 | 31891006 | A | G | 0.027341382 | 0.047063338 | -0.0149 | -0.0149 | 0.000008226 | 0.0005769 | new | TSHZ3 | intergenic | . | . | 19q12 |
| GERD | PTSD | rs817311 | 20 | 62580728 | A | G | 0.01014919 | 0.046781435 | 0.0202 | 0.0168 | 6.08E-08 | 0.0007117 | new | UCKL1 | intronic | . | . | 20q13.33 |
| GERD | PTSD | rs11090045 | 22 | 41753603 | A | G | 0.008361059 | 0.030220152 | 0.0212 | 0.0176 | 2.958E-08 | 0.0002706 | known | ZC3H7B | UTR3 | . | . | 22q13.2 |

Note:

Trait1/Trait2: trait pair analyzed;

SNP: lead single-nucleotide polymorphism;

CHR: chromosome;

BP: base-pair position;

A1/A2: effect allele and non-effect allele;

condfdr: conditional false discovery rate for GERD given the conditioning trait;

conjfdr: conjunctional false discovery rate;

beta1/beta2: effect size estimates for Trait1 and Trait2;

pval1/pval2: association P-values for Trait1 and Trait2;

annotation: locus novelty status (new, not previously reported in any GERD GWAS; new*, previously reported in psychiatric disorder GWAS but newly identified as GERD-associated);

Gene: nearest gene;

Function: functional annotation relative to RefSeq gene (e.g., intronic, intergenic);

Exonic Function: exonic functional consequence (e.g., synonymous, nonsynonymous);

CADD: Combined Annotation Dependent Depletion (CADD) Phred-scaled score;

cytoBand: cytogenetic band location.

Table S8. Genome-wide significant loci identified by Genomic SEM analysis of GERD and F1 factor traits.

| SNP | CHR | BP | A1 | A2 | Z_Estimate | Pval_Estimate | GERD_z | GERD_pval | ADHD_z | ADHD_pval | ANX_z | ANX_pval | MDD_z | MDD_pval | PTSD_z | PTSD_pval | BMI_z | BMI_pval | annotation | Gene | Function | Exonic Function | CADD | cytoBand | confdr_repeat |
| --- | --- | --- | --- | --- | --- | --- | --- | --- | --- | --- | --- | --- | --- | --- | --- | --- | --- | --- | --- | --- | --- | --- | --- | --- | --- |
| rs301806 | 1 | 8482078 | C | T | 6.17420527 | 6.64971E-10 | -4.002487426 | 0.00006268 | 2.681970105 | 0.007319 | -2.518966064 | 0.01177 | 6.318566943 | 2.64E-10 | -3.458297873 | 0.0005436 | 4.112333624 | 3.9168E-05 | new* | RERE | intronic | . | . | 1p36.23 | Y |
| rs3766823 | 1 | 32197257 | G | A | -7.10266595 | 1.22373E-12 | 4.891638476 | 0.000001 | -4.056798348 | 0.00004975 | 2.821363389 | 0.004782 | 3.929168773 | 0.00008524 | 3.677858901 | 0.0002352 | 4.788987304 | 1.67625E-06 | new | ADGRB2 | intronic | . | . | 1p35.2 | Y |
| rs4361958 | 1 | 33796554 | G | A | -6.299461887 | 2.98681E-10 | 3.979707886 | 0.000069 | -2.387140513 | 0.01698 | 3.283298399 | 0.001026 | 4.426127119 | 0.000009594 | 2.461908289 | 0.01382 | 3.282223571 | 0.001029919 | new | PHC2 | intronic | . | . | 1p35.1 | Y |
| rs1002656 | 1 | 37192741 | C | T | 8.192447111 | 2.55968E-16 | -5.344753864 | 9.054E-08 | 1.52363466 | 0.1276 | -4.229510834 | 0.00002342 | 7.39392892 | 1.426E-13 | -4.552587936 | 0.000005299 | -0.441652444 | 0.658740728 | new* | GRIK3 | intergenic | . | . | 1p34.3 | Y |
| rs2503185 | 1 | 66461401 | A | G | 8.049438113 | 8.3175E-16 | 6.162644492 | 7.154E-10 | 2.468448967 | 0.01357 | -3.775948384 | 0.0001594 | -5.150711578 | 2.595E-07 | 3.768852134 | 0.000164 | -6.36931103 | 1.89879E-10 | known | PDE4B | intronic | . | . | 1p31.3 | Y |
| rs2590942 | 1 | 72885281 | T | G | 11.54656348 | 7.68315E-31 | 5.1024976 | 3.352E-07 | 1.683491415 | 0.09228 | -5.094209149 | 3.502E-07 | -8.209536152 | 2.41E-16 | 3.997888655 | 0.00006391 | -13.89160242 | 7.12239E-44 | new* | NEGR1 | intergenic | . | . | 1p31.1 |  |
| rs12080711 | 1 | 80814326 | A | G | -7.816653093 | 5.42463E-15 | -5.087926293 | 0.000000362 | -1.055492802 | 0.2912 | 3.759578306 | 0.0001702 | 6.297543197 | 3.024E-10 | -2.91789765 | 0.003524 | 5.733031127 | 9.86515E-09 | new* | LINC01781 | intergenic | . | . | 1p31.1 | Y |
| rs197419 | 1 | 112315450 | A | C | -7.078773431 | 1.45436E-12 | -4.441461343 | 0.000008935 | -3.075003618 | 0.002105 | 2.569654455 | 0.01018 | 3.943622393 | 0.00008026 | -3.933809666 | 0.00008361 | 6.913830853 | 4.71738E-12 | new* | KCND3 | UTR3 | . | . | 1p13.2 | Y |
| rs3790514 | 1 | 151744168 | T | C | 6.142886882 | 8.1035E-10 | 4.030155358 | 0.00005574 | 2.350067636 | 0.01877 | -3.124182221 | 0.001783 | -4.488875874 | 0.00000716 | 2.596386592 | 0.009421 | -2.428958143 | 0.01514228 | new | TDRKH | UTR3 | . | . | 1q21.3 | Y |
| rs6691053 | 1 | 173868955 | C | T | 6.273458818 | 3.53114E-10 | -4.291462727 | 0.00001775 | 2.623153546 | 0.008712 | -0.756749421 | 0.4492 | -6.267368684 | 3.672E-10 | -1.269676183 | 0.2042 | -4.777526276 | 1.77465E-06 | new* | ZBTB37 | UTR3 | . | . | 1q25.1 | Y |
| rs1760376 | 1 | 191210269 | A | C | -6.628406544 | 3.3933E-11 | -5.151435595 | 2.585E-07 | -1.179246588 | 0.2383 | 3.684446677 | 0.0002292 | 4.474333576 | 0.000007665 | -4.763037907 | 0.000001907 | 1.173149646 | 0.240735801 | new | LINC01680 | intergenic | . | . | 1q31.2 | Y |
| rs12614708 | 2 | 22568238 | C | A | -10.65149656 | 1.71584E-26 | 4.492415085 | 0.000007042 | -3.838873671 | 0.0001236 | 6.165885026 | 7.009E-10 | 8.209536152 | 2.179E-16 | 6.712132876 | 1.918E-11 | 2.694372093 | 0.007052139 | new* | LINC01884 | intergenic | . | . | 2p24.1 | Y |
| rs3842981 | 2 | 50976459 | T | G | 5.725687423 | 1.03016E-08 | 4.121814403 | 0.00003759 | 1.4121697 | 0.1579 | 0.315476356 | 0.7524 | 5.73077976 | 9.997E-09 | 2.137467332 | 0.03256 | -4.248383001 | 2.15319E-05 | new* | NRXN1 | intronic | . | . | 2p16.3 | Y |
| rs359233 | 2 | 60470926 | A | G | -8.004702053 | 1.19756E-15 | -4.522200661 | 0.00000612 | -3.024675523 | 0.002489 | 4.565375937 | 0.000004986 | -5.309212346 | 1.101E-07 | -3.734886321 | 0.0001878 | 5.442642851 | 5.24958E-08 | new | MIR4432HG | intergenic | . | . | 2p16.1 | Y |
| rs11887574 | 2 | 100109251 | C | A | -6.748531507 | 1.49349E-11 | 6.079178269 | 1.208E-09 | -3.345683594 | 0.0008208 | 2.240014216 | 0.02509 | -4.33920359 | 0.0000143 | 2.42165525 | 0.01545 | 3.184576033 | 0.001449662 | known | REV1 | intergenic | . | . | 2q11.2 |  |
| rs12105749 | 2 | 100500674 | C | T | 6.92888534 | 4.24169E-12 | -6.885191127 | 5.771E-12 | 4.251124378 | 0.00002127 | -2.626728851 | 0.008621 | -4.728037789 | 0.000002267 | -4.001545841 | 0.00006293 | -1.320872784 | 0.186543786 | known | AFF3 | intronic | . | . | 2q11.2 | Y |
| rs6739285 | 2 | 105357317 | C | A | -5.95896995 | 2.53833E-09 | 4.073157686 | 0.00004638 | -2.121854004 | 0.03385 | 4.657482258 | 0.000003201 | 3.004978068 | 0.002656 | 2.644465695 | 0.008182 | 4.008018156 | 6.12304E-05 | new | LINC01114 | intergenic | . | . | 2q12.1 | Y |
| rs11691266 | 2 | 109829904 | A | G | 6.711022842 | 1.93265E-11 | 5.554693057 | 2.781E-08 | 1.556454728 | 0.1196 | -4.97726201 | 6.449E-07 | -4.216430258 | 0.00002482 | 3.916134665 | 0.00008998 | -1.105103965 | 0.269114543 | known | SH3RF3 | intronic | . | . | 2q12.3 | Y |
| rs7581524 | 2 | 113234316 | C | T | -5.792299193 | 6.94293E-09 | 6.745691417 | 1.523E-11 | -2.162106086 | 0.03061 | 2.728827201 | 0.006356 | 4.179773177 | 0.00002918 | 2.670936298 | 0.007564 | 1.641872599 | 0.10061641 | known | TTL | intergenic | . | . | 2q13 | Y |
| rs6430030 | 2 | 144166523 | C | T | 8.083890838 | 6.27325E-16 | -4.753323289 | 0.000002001 | 2.739034003 | 0.006162 | -4.162777279 | 0.00003144 | 5.642806849 | 1.673E-08 | -5.200643233 | 1.986E-07 | -2.425713095 | 0.015278343 | new* | ARHGAP15 | intronic | . | . | 2q22.3 | Y |
| rs17755617 | 2 | 161935424 | C | A | 7.846490043 | 4.27843E-15 | -4.468255719 | 0.000007886 | 2.682519005 | 0.007307 | -2.835388422 | 0.004577 | -5.962439286 | 2.485E-09 | -4.737379041 | 0.000002165 | -2.305649518 | 0.021130224 | new* | TANK | intergenic | . | . | 2q24.2 | Y |
| rs13026466 | 2 | 173811426 | T | G | 7.487944132 | 6.99608E-14 | 4.305910884 | 0.00001663 | 4.440069095 | 0.000008993 | -2.247000424 | 0.02464 | -5.763760756 | 8.226E-09 | 2.980264063 | 0.00288 | -3.225225028 | 0.001258737 | new* | RAPGEF4 | intronic | . | . | 2q31.1 | Y |
| rs1374369 | 2 | 176721984 | G | A | 6.030999279 | 1.62949E-09 | -5.975987883 | 2.287E-09 | 1.655515376 | 0.09782 | -3.378169831 | 0.0007297 | -3.797097869 | 0.0001464 | -4.052561043 | 0.00005066 | -1.005655284 | 0.31458142 | known | LNPK | intergenic | . | . | 2q31.1 | Y |
| rs12995239 | 2 | 193817686 | A | G | -6.372153817 | 1.86392E-10 | -7.050793769 | 1.779E-12 | -1.718310474 | 0.08574 | 2.924509093 | 0.00345 | -4.573737512 | 0.000004791 | -2.133057617 | 0.03292 | 3.15379288 | 0.001611634 | known | PCGEM1 | intergenic | . | . | 2q32.3 | Y |
| rs7558609 | 2 | 233700379 | A | G | 5.987788121 | 2.12714E-09 | 4.09597241 | 0.00004204 | 4.556229631 | 0.000005208 | -2.273434651 | 0.023 | 5.901780224 | 3.596E-09 | 2.337561637 | 0.01941 | 2.049237322 | 0.040438914 | new* | GIGYF2 | intronic | . | . | 2q37.1 | Y |
| rs1440512 | 3 | 29732862 | C | T | -6.507394543 | 7.64654E-11 | 3.966094191 | 0.00007306 | -2.397220035 | 0.01652 | 4.033124927 | 0.00005504 | -4.775905946 | 0.000001789 | 2.142416268 | 0.03216 | 3.654430584 | 0.000257753 | new | RBMS3 | intronic | . | . | 3p24.1 | Y |
| rs6802567 | 3 | 43537085 | T | C | -7.468057467 | 8.13873E-14 | -4.234756002 | 0.00002288 | -5.385086046 | 7.241E-08 | 2.340072438 | 0.01928 | 5.251580012 | 1.508E-07 | -6.167655361 | 6.931E-10 | -0.625659545 | 0.531538274 | new* | ANO10 | intronic | . | . | 3p22.1 | Y |
| rs2271961 | 3 | 49878113 | T | C | -10.27606626 | 9.03399E-25 | -5.681395672 | 1.336E-08 | -3.704869012 | 0.0002115 | 4.433489558 | 0.000009272 | 5.62086837 | 0.000000019 | -4.84175096 | 0.000001287 | 11.20027178 | 4.06485E-29 | known | TRAIP | intronic | . | . | 3p21.31 | Y |
| rs1010553 | 3 | 52540773 | T | C | -6.287019192 | 3.23619E-10 | -4.232507698 | 0.00002311 | -1.664512776 | 0.09601 | 5.868257094 | 4.404E-09 | 6.742299091 | 1.559E-11 | -4.130674765 | 0.00003617 | -5.822454954 | 5.79894E-09 | new* | STAB1 | exonic | synonymous SNV | . | 3p21.1 | Y |
| rs1586070 | 3 | 65172403 | C | T | 6.678220795 | 2.4186E-11 | -3.969379621 | 0.00007206 | 2.266022593 | 0.02345 | -2.993940107 | 0.002754 | 4.303527463 | 0.00001681 | -3.495260093 | 0.0004736 | -4.726171303 | 2.28793E-06 | new | MAGI1 | intergenic | . | . | 3p14.1 | Y |
| rs4683981 | 3 | 77240101 | A | G | 6.118623875 | 9.43869E-10 | 4.190940917 | 0.00002778 | 4.525580321 | 0.000006023 | -2.089864203 | 0.03663 | -6.002084726 | 1.948E-09 | 2.43069246 | 0.01507 | 0.086781016 | 0.930845578 | new* | ROBO2 | intronic | . | . | 3p12.3 | Y |
| rs488029 | 3 | 173119378 | G | A | -7.858879288 | 3.87586E-15 | 5.386382222 | 7.189E-08 | -1.803816999 | 0.07126 | 3.463801181 | 0.0005326 | -4.566134532 | 0.000004968 | 3.470185001 | 0.0005201 | 8.086426058 | 6.1441E-16 | new* | NLGN1 | intronic | . | . | 3q26.31 | Y |
| rs12506821 | 4 | 3284560 | T | C | -6.797527853 | 1.0643E-11 | -5.442730497 | 5.247E-08 | -0.866164424 | 0.3864 | 3.341756693 | 0.0008325 | 4.28166512 | 0.00001855 | -3.034418326 | 0.00241 | 6.196404 | 5.77677E-10 | new* | MSANTD1 | intergenic | . | . | 4p16.3 | Y |
| rs1031034 | 4 | 102223386 | C | A | -7.576166415 | 3.55915E-14 | 5.766301594 | 8.103E-09 | -1.993013215 | 0.04626 | 3.551666303 | 0.0003828 | 5.109917218 | 3.223E-07 | 2.896126561 | 0.003778 | 4.987396418 | 6.11984E-07 | known | PPP3CA | intronic | . | . | 4q24 | Y |
| rs233816 | 4 | 103206555 | G | T | -6.79038685 | 1.11833E-11 | 4.944652012 | 7.628E-07 | -1.459963884 | 0.1443 | 2.893973719 | 0.003804 | 2.604754914 | 0.009194 | 2.883646302 | 0.003931 | 9.956766598 | 2.35601E-23 | new* | SLC39A8 | intronic | . | . | 4q24 |  |
| rs4863720 | 4 | 140923030 | T | C | -9.620198398 | 6.57043E-22 | -4.422356923 | 0.000009763 | -3.330339993 | 0.0008674 | 4.619373746 | 0.000003849 | 7.84181248 | 4.458E-15 | -5.343517548 | 9.116E-08 | 4.632644374 | 3.61024E-06 | new* | MAML3 | intronic | . | . | 4q31.1 | Y |
| rs10941692 | 5 | 45099778 | T | C | 6.650039448 | 2.93014E-11 | 6.370999708 | 1.878E-10 | 3.738257645 | 0.0001853 | -0.305874765 | 0.7597 | -4.254731518 | 0.00002093 | 4.019749719 | 0.00005826 | -3.034237214 | 0.002411447 | known | HCN1 | intergenic | . | . | 5p12 | Y |
| rs918634 | 5 | 50860339 | A | G | -5.469880094 | 4.5034E-08 | -4.008483143 | 0.00006111 | -0.385860481 | 0.6996 | 0.919756538 | 0.3577 | 4.608902549 | 0.000004048 | -4.044611325 | 0.00005241 | 1.387188607 | 0.165384259 | new | ISL1 | intergenic | . | . | 5q11.2 | Y |
| rs16879569 | 5 | 51573487 | G | C | 5.988496609 | 2.1179E-09 | -4.633577571 | 0.000003594 | -0.226001826 | 0.8212 | -3.067856671 | 0.002156 | -4.483205663 | 0.000007353 | -3.90164805 | 0.00009554 | -1.253739472 | 0.209936716 | new | LINC02118 | intergenic | . | . | 5q11.2 | Y |
| rs288178 | 5 | 107351517 | T | C | 8.690361507 | 3.61288E-18 | 4.253877787 | 0.00002101 | 4.156471831 | 0.00003232 | -5.339350797 | 9.328E-08 | -4.059870215 | 0.0000491 | 4.737379041 | 0.000002165 | -7.579893323 | 3.45839E-14 | new* | FBXL17 | intronic | . | . | 5q21.3 | Y |
| rs10056247 | 5 | 133898136 | C | T | 6.484832367 | 8.88305E-11 | -3.970141368 | 0.00007183 | 2.008030977 | 0.04464 | -3.110137929 | 0.00187 | -3.850244437 | 0.000118 | -2.984545457 | 0.00284 | -6.732469132 | 1.66808E-11 | new* | JADE2 | intronic | . | . | 5q31.1 |  |
| rs2563263 | 5 | 140137644 | C | T | 5.731015005 | 9.98314E-09 | -4.20026734 | 0.00002666 | 0.370916131 | 0.7107 | -3.420657289 | 0.0006247 | -3.144976803 | 0.001661 | -3.438151459 | 0.0005857 | -3.016538411 | 0.002556788 | new | PCDHA1 | intergenic | . | . | 5q31.3 |  |
| rs29267 | 6 | 29575003 | G | A | -9.014538733 | 1.97701E-19 | 6.051391486 | 1.436E-09 | -1.036862377 | 0.2998 | 4.827151156 | 0.000001385 | 6.490169104 | 8.574E-11 | 5.552445845 | 2.817E-08 | 3.911169445 | 9.18503E-05 | known | GABBR1 | intronic | . | . | 6p22.1 |  |
| rs397228 | 6 | 64626592 | C | T | -7.074206028 | 1.50307E-12 | 4.703291907 | 0.00000256 | 0.252700062 | 0.8005 | 2.225763901 | 0.02603 | -6.050050992 | 1.448E-09 | 4.349095784 | 0.00001367 | 3.097075313 | 0.001954402 | new* | EYS | intronic | . | . | 6q12 | Y |
| rs607416 | 6 | 65085180 | C | T | -5.968086391 | 2.40052E-09 | 6.131307049 | 8.716E-10 | -0.554699819 | 0.5791 | 1.219907348 | 0.2225 | -5.445190401 | 5.175E-08 | 3.104656684 | 0.001905 | 0.243335339 | 0.807745631 | known | EYS | intronic | . | . | 6q12 | Y |
| rs1933720 | 6 | 98297832 | T | C | 5.87887091 | 4.13074E-09 | 6.001336451 | 1.957E-09 | 0.980377119 | 0.3269 | -3.564104853 | 0.0003651 | -1.573081276 | 0.1157 | 3.36711139 | 0.0007596 | -6.440843663 | 1.18811E-10 | known | LOC101927314 | intergenic | . | . | 6q16.1 | Y |
| rs9373363 | 6 | 143150043 | A | G | 7.123773045 | 1.05012E-12 | 5.395429055 | 6.836E-08 | 1.24508638 | 0.2131 | -5.679486135 | 1.351E-08 | -5.107992251 | 3.256E-07 | 3.030437157 | 0.002442 | -2.987355793 | 0.00281402 | new* | HIVEP2 | intronic | . | . | 6q24.2 | Y |
| rs11755113 | 6 | 147964336 | T | C | 7.062892228 | 1.63072E-12 | 5.362825824 | 8.193E-08 | 2.604196284 | 0.009209 | -3.85400757 | 0.0001162 | 5.154285353 | 2.546E-07 | 2.629422598 | 0.008553 | -2.984168833 | 0.002843498 | new | SAMD5 | intergenic | . | . | 6q24.3 | Y |
| rs6557171 | 6 | 152234593 | T | C | 10.36291875 | 3.65631E-25 | 5.309046861 | 1.102E-07 | 2.875412473 | 0.004035 | -3.266983541 | 0.001087 | 50 | 1.407E-20 | 6.6222804 | 3.537E-11 | -1.3137738 | 0.188922337 | new* | ESR1 | intronic | . | . | 6q25.1 | Y |
| rs11764337 | 7 | 1892309 | C | T | 11.43014568 | 2.95612E-30 | -6.942727819 | 3.846E-12 | 3.600504771 | 0.0003176 | -2.832746282 | 0.004615 | -50 | 6.675E-19 | -6.665258579 | 2.642E-11 | -5.90983248 | 3.42456E-09 | known | MAD1L1 | intronic | . | . | 7p22.3 | Y |
| rs2341447 | 7 | 3703188 | G | C | 7.622691302 | 2.48441E-14 | -5.041705915 | 4.614E-07 | 3.337124566 | 0.0008465 | -1.627645855 | 0.1036 | -6.231801999 | 4.611E-10 | -4.546212083 | 0.000005462 | -2.351103407 | 0.018717831 | new* | SDK1 | intronic | . | . | 7p22.2 | Y |
| rs10275122 | 7 | 69897826 | A | G | -6.085554259 | 1.16089E-09 | -5.010566644 | 5.427E-07 | -0.512358686 | 0.6084 | 2.560904666 | 0.01044 | 4.615050365 | 0.00000393 | -2.669430481 | 0.007598 | 4.301109778 | 1.69945E-05 | new | AUTS2 | intronic | . | . | 7q11.22 | Y |
| rs2715147 | 7 | 82448405 | C | T | -8.993362086 | 2.39781E-19 | 5.029017267 | 0.000000493 | -3.413093081 | 0.0006423 | 5.510311119 | 3.582E-08 | 6.92834487 | 4.258E-12 | 5.226017472 | 1.732E-07 | 2.465621304 | 0.013677585 | new* | PCLO | intronic | . | . | 7q21.11 | Y |
| rs2894699 | 7 | 114059156 | C | T | 9.559809499 | 1.17974E-21 | -5.78092728 | 7.429E-09 | 6.156564298 | 7.434E-10 | -3.663903915 | 0.0002484 | -7.637172105 | 2.21E-14 | -6.605491422 | 3.962E-11 | -0.028208631 | 0.977495753 | known | FOXP2 | intronic | . | . | 7q31.1 | Y |
| rs1621 | 7 | 116437606 | G | A | 6.775179425 | 1.24252E-11 | -5.633684116 | 1.764E-08 | 3.219978972 | 0.001282 | -1.574811332 | 0.1153 | 3.88270703 | 0.0001033 | -5.26148056 | 1.429E-07 | -3.026744785 | 0.002472026 | known | MET | UTR3 | . | . | 7q31.2 | Y |
| rs10487459 | 7 | 126378255 | A | G | -6.524242439 | 6.83461E-11 | -5.087873898 | 3.621E-07 | -1.556034032 | 0.1197 | 3.903478441 | 0.00009482 | 4.447711471 | 0.000008679 | -4.019628429 | 0.00005829 | 1.684595216 | 0.092066695 | known | GRM8 | intronic | . | . | 7q31.33 | Y |
| rs10243212 | 7 | 146673884 | A | G | 5.747418033 | 9.06165E-09 | 5.141330289 | 2.728E-07 | 3.031056056 | 0.002437 | -4.198153676 | 0.00002691 | -4.345592852 | 0.00001389 | 2.084111789 | 0.03715 | -0.131356188 | 0.895493545 | new | CNTNAP2 | intronic | . | . | 7q35 | Y |
| rs11786924 | 8 | 12686438 | T | G | -5.984375074 | 2.17222E-09 | -4.292968533 | 0.00001763 | -4.325480113 | 0.00001522 | 2.923338337 | 0.003463 | 4.625109807 | 0.000003744 | -2.846952125 | 0.004414 | -0.712759387 | 0.475994661 | new | LINC00681 | intergenic | . | . | 8p23.1 | Y |
| rs7844647 | 8 | 34503776 | T | C | 6.749574997 | 1.48279E-11 | 4.141262098 | 0.00003454 | 2.983684793 | 0.002848 | -1.370598984 | 0.1705 | -5.020303135 | 5.159E-07 | 2.357872593 | 0.01838 | -5.935845343 | 2.92335E-09 | new* | LINC01288 | intergenic | . | . | 8p12 |  |
| rs3863241 | 8 | 73890335 | C | T | -7.39532484 | 1.41063E-13 | 5.190349252 | 2.099E-07 | -2.499410835 | 0.01244 | 3.435207623 | 0.0005921 | -5.019152269 | 0.000000519 | 2.696511396 | 0.007007 | 5.183190076 | 2.18123E-07 | new | TERF1 | intergenic | . | . | 8q13.3 | Y |
| rs11784932 | 8 | 130095478 | C | A | -5.797547728 | 6.72916E-09 | 3.89591177 | 0.00009783 | -1.935491588 | 0.05293 | 1.573081276 | 0.1157 | 4.728935559 | 0.000002257 | 2.124240957 | 0.03365 | 2.731839109 | 0.006298189 | new | LINC00976 | intergenic | . | . | 8q24.21 | Y |
| rs10098073 | 8 | 143309504 | C | A | -5.54804797 | 2.88877E-08 | 4.013204406 | 0.0000599 | 0.644579073 | 0.5192 | 6.022352503 | 1.719E-09 | 4.612842124 | 0.000003972 | 4.761247411 | 0.000001924 | -3.850830415 | 0.000117718 | new* | TSNARE1 | intronic | . | . | 8q24.3 | Y |
| rs10810098 | 9 | 14161670 | T | C | 6.343096973 | 2.25192E-10 | 4.505650246 | 0.000006617 | 2.804721963 | 0.005036 | -4.121875681 | 0.00003758 | 4.613785759 | 0.000003954 | 1.977634095 | 0.04797 | -2.805026128 | 0.00503125 | new | NFIB | intronic | . | . | 9p23 | Y |
| rs10119773 | 9 | 23736400 | A | G | -5.952094767 | 2.64732E-09 | -5.358275616 | 8.402E-08 | -3.103262175 | 0.001914 | 0.994457883 | 0.32 | -5.43259618 | 5.554E-08 | -3.25876073 | 0.001119 | 0.045473232 | 0.963730111 | new | ELAVL2 | intronic | . | . | 9p21.3 | Y |
| rs7024334 | 9 | 109072075 | T | G | 6.788814746 | 1.13059E-11 | 5.395507863 | 6.833E-08 | 1.445918722 | 0.1482 | -2.947070554 | 0.003208 | 5.347433849 | 8.921E-08 | 1.24182808 | 0.2143 | -6.9056711 | 4.99666E-12 | new* | MIR8081 | intergenic | . | . | 9q31.2 | Y |
| rs4837502 | 9 | 120517844 | G | A | 7.369151254 | 1.71718E-13 | -4.879046374 | 0.000001066 | 3.63899887 | 0.0002737 | -3.332595292 | 0.0008604 | 5.130678266 | 2.887E-07 | -3.412838624 | 0.0006429 | -3.694396465 | 0.00022041 | new | TLR4 | intergenic | . | . | 9q33.1 | Y |
| rs1333908 | 9 | 122673299 | T | G | -9.023701998 | 1.81838E-19 | -4.620454669 | 0.000003829 | -3.132541375 | 0.001733 | 4.060012923 | 0.00004907 | -7.207900282 | 5.683E-13 | -4.804420953 | 0.000001552 | 3.54353754 | 0.000394797 | new* | LINC01613 | intergenic | . | . | 9q33.2 | Y |
| rs10818846 | 9 | 126346105 | T | C | -6.542128745 | 6.06493E-11 | -4.66308765 | 0.000003115 | -1.984052389 | 0.04725 | 1.994659769 | 0.04608 | 3.981749009 | 0.00006841 | -3.585448039 | 0.0003365 | 4.769783779 | 1.84424E-06 | new | DENND1A | intronic | . | . | 9q33.3 | Y |
| rs10760344 | 9 | 127026282 | G | T | -6.333342592 | 2.39906E-10 | 5.004731321 | 5.594E-07 | -2.696083865 | 0.007016 | 3.074578837 | 0.002108 | 3.237457267 | 0.001206 | 1.940491386 | 0.05232 | 5.390943013 | 7.00889E-08 | new | NEK6 | intronic | . | . | 9q33.3 |  |
| rs589292 | 9 | 127818276 | C | T | 7.298740262 | 2.90474E-13 | -4.644077092 | 0.000003416 | 3.087570351 | 0.002018 | -3.659406559 | 0.0002528 | -6.433133208 | 1.25E-10 | -4.337519124 | 0.00001441 | 1.371236054 | 0.170301382 | new* | SCAI | exonic | nonsynonymous SNV | 24.5 | 9q33.3 | Y |
| rs10828248 | 10 | 21824619 | A | G | -7.606555069 | 2.81498E-14 | -6.127554565 | 8.924E-10 | -2.074639051 | 0.03802 | 1.166283109 | 0.2435 | 4.393968403 | 0.00001113 | -3.61469622 | 0.0003007 | 9.775077771 | 1.44047E-22 | known | MLLT10 | intronic | . | . | 10p12.31 | Y |
| rs942093 | 10 | 77626488 | C | G | -7.678626332 | 1.60804E-14 | -4.724837005 | 0.000002303 | -2.695894009 | 0.00702 | 2.223828279 | 0.02616 | -5.344893963 | 9.047E-08 | -3.731689303 | 0.0001902 | 5.866021724 | 4.46375E-09 | new* | LRMDA | intronic | . | . | 10q22.2 | Y |
| rs11599236 | 10 | 106454672 | T | C | 9.765858289 | 1.5777E-22 | 5.715783029 | 1.092E-08 | 5.239284916 | 1.612E-07 | -6.478205309 | 9.282E-11 | -8.125890665 | 4.929E-16 | 2.455215892 | 0.01408 | -3.70665545 | 0.000210014 | known | SORCS3 | intronic | . | . | 10q25.1 | Y |
| rs12768641 | 10 | 134966154 | A | G | -7.367293216 | 1.74127E-13 | -5.660196235 | 1.512E-08 | -3.977785766 | 0.00006956 | 2.076583685 | 0.03784 | 5.73979719 | 9.479E-09 | -3.399375179 | 0.0006754 | 3.111500063 | 0.001861394 | known | KNDC1 | intergenic | . | . | 10q26.3 | Y |
| rs7942007 | 11 | 31835418 | A | G | -6.674424268 | 2.48205E-11 | -6.025392228 | 1.687E-09 | -2.097379436 | 0.03596 | 5.165108104 | 2.403E-07 | 5.523559352 | 3.322E-08 | -5.055217743 | 4.299E-07 | -4.437882047 | 9.08484E-06 | known | PAX6 | intronic | . | . | 11p13 | Y |
| rs10896012 | 11 | 65278461 | T | C | -5.919326147 | 3.23263E-09 | -4.748234534 | 0.000002052 | -1.462880795 | 0.1435 | 1.786922595 | 0.07395 | 4.931788772 | 8.148E-07 | -0.354318773 | 0.7231 | 6.748698524 | 1.49177E-11 | new* | TALAM1 | intergenic | . | . | 11q13.1 | Y |
| rs3758711 | 11 | 76495560 | C | T | 5.633761043 | 1.76321E-08 | -4.638645499 | 0.000003507 | 2.244659498 | 0.02479 | -1.534527228 | 0.1249 | -3.149947151 | 0.001633 | -1.765677319 | 0.07745 | -5.661168901 | 1.50345E-08 | new* | TSKU | intronic | . | . | 11q13.5 |  |
| rs719182 | 11 | 88289382 | G | A | 6.389836346 | 1.66063E-10 | -4.111674397 | 0.00003928 | 0.675276675 | 0.4995 | -3.029942872 | 0.002446 | -5.452702599 | 4.961E-08 | -4.038299867 | 0.00005384 | -1.598641966 | 0.109900179 | new* | GRM5 | intronic | . | . | 11q14.2 | Y |
| rs7130186 | 11 | 104466550 | C | A | 5.738196918 | 9.56898E-09 | -4.292214414 | 0.00001769 | 2.08653708 | 0.03693 | -4.132973518 | 0.00003581 | -3.966420814 | 0.00007296 | -3.045233146 | 0.002325 | -0.675336465 | 0.499462022 | new | LINC02552 | ncRNA_intronic | . | . | 11q22.3 | Y |
| rs4938021 | 11 | 113364803 | C | T | -9.864744089 | 5.91852E-23 | 4.354781026 | 0.00001332 | -1.917219908 | 0.05521 | 4.985474285 | 6.181E-07 | -50 | 3.313E-27 | 4.621541018 | 0.000003809 | 2.571436487 | 0.010127759 | new* | DRD2 | intergenic | . | . | 11q23.2 | Y |
| rs12578517 | 12 | 15407185 | T | C | -5.467332622 | 4.56858E-08 | -5.298590549 | 1.167E-07 | -1.264083662 | 0.2062 | 3.650333535 | 0.0002619 | 2.831846914 | 0.004628 | -2.251877727 | 0.02433 | 3.502986151 | 0.000460073 | new | RERG | intergenic | . | . | 12p12.3 | Y |
| rs2058799 | 12 | 16743217 | A | G | -5.876531895 | 4.18951E-09 | -4.213000953 | 0.0000252 | -3.801194857 | 0.000144 | 0.774897888 | 0.4384 | -4.286757525 | 0.00001813 | -4.095257183 | 0.00004217 | 1.842832637 | 0.06535345 | new | LMO3 | intronic | . | . | 12p12.3 | Y |
| rs2970839 | 12 | 19254829 | G | A | -5.662244323 | 1.49406E-08 | 4.06230756 | 0.00004859 | 0.394651636 | 0.6931 | 2.157714961 | 0.03095 | -4.417779662 | 0.000009972 | 2.468976808 | 0.01355 | 5.353587555 | 8.62273E-08 | new | PLEKHA5 | intergenic | . | . | 12p12.3 |  |
| rs773107 | 12 | 56369506 | A | G | 5.484496165 | 4.14649E-08 | 6.014358893 | 1.806E-09 | 4.243663522 | 0.00002199 | -0.565549596 | 0.5717 | -2.940862297 | 0.003273 | 1.666720473 | 0.09557 | -6.168256257 | 6.90472E-10 | known | RAB5B | intronic | . | . | 12q13.2 | Y |
| rs11174461 | 12 | 62797683 | G | A | -6.644009001 | 3.05264E-11 | 3.920487554 | 0.00008837 | -3.448169369 | 0.0005644 | 3.037941742 | 0.002382 | 5.264462622 | 1.406E-07 | 2.309605587 | 0.02091 | 3.602746934 | 0.000314872 | new | USP15 | intronic | . | . | 12q14.1 | Y |
| rs12308738 | 12 | 84220855 | G | T | -5.63475731 | 1.75305E-08 | 4.494175541 | 0.000006984 | -2.065454674 | 0.03888 | 3.452168662 | 0.0005561 | -3.913653669 | 0.00009091 | 2.530500262 | 0.01139 | 2.267794296 | 0.023341747 | new | TMTC2 | intergenic | . | . | 12q21.31 | Y |
| rs8001047 | 13 | 33376884 | G | A | 5.76682546 | 8.07786E-09 | -4.955620019 | 0.000000721 | 2.774833354 | 0.005523 | -2.84208173 | 0.004482 | -2.217777602 | 0.02657 | -1.801716632 | 0.07159 | -7.954414138 | 1.79981E-15 | new* | LINC00423 | intergenic | . | . | 13q13.1 |  |
| rs1334297 | 13 | 58335375 | G | A | 7.651207098 | 1.99101E-14 | -4.506937487 | 0.000006577 | 4.393578308 | 0.00001115 | -2.561570802 | 0.01042 | 5.115506603 | 3.129E-07 | -3.325721493 | 0.0008819 | -5.300839094 | 1.15272E-07 | new | PCDH17 | intergenic | . | . | 13q21.1 | Y |
| rs7995254 | 13 | 67603745 | T | C | -6.668480554 | 2.58465E-11 | -3.979329164 | 0.00006911 | -2.111026898 | 0.03477 | 2.65110035 | 0.008023 | 6.213128294 | 5.194E-10 | -2.267986984 | 0.02333 | 2.909644527 | 0.0036184 | new* | PCDH9 | intronic | . | . | 13q21.32 |  |
| rs1950829 | 14 | 42097937 | A | G | 8.154475506 | 3.50698E-16 | 4.086091939 | 0.00004387 | 2.677698107 | 0.007413 | -5.326542258 | 1.001E-07 | 7.808807497 | 5.757E-15 | 3.889140182 | 0.0001006 | 0.88705576 | 0.37504888 | new* | LRFN5 | intronic | . | . | 14q21.1 | Y |
| rs7140574 | 14 | 69751282 | G | A | -6.62976106 | 3.36231E-11 | 4.360441974 | 0.00001298 | -1.052217004 | 0.2927 | 2.208204799 | 0.02723 | 3.541340148 | 0.0003981 | 5.362385563 | 8.213E-08 | 4.66239297 | 3.12554E-06 | new | GALNT16 | intronic | . | . | 14q24.1 | Y |
| rs4583163 | 14 | 71296910 | T | C | 5.499952356 | 3.79894E-08 | 3.908112481 | 0.00009302 | 1.5280505 | 0.1265 | -3.874599956 | 0.0001068 | -3.101719766 | 0.001924 | 2.903514401 | 0.00369 | -3.53352347 | 0.00041006 | new | MAP3K9 | intergenic | . | . | 14q24.2 |  |
| rs17094893 | 14 | 93994870 | A | G | -5.90333733 | 3.56221E-09 | -4.023498293 | 0.00005734 | -2.037399739 | 0.04161 | 0.847709095 | 0.3966 | -3.781158892 | 0.0001561 | -0.928593098 | 0.3531 | 9.025800535 | 1.78386E-19 | new* | UNC79 | intronic | . | . | 14q32.12 | Y |
| rs7143963 | 14 | 103304425 | C | T | -9.610520081 | 7.21835E-22 | 5.782839808 | 7.345E-09 | -4.544981804 | 0.000005494 | 3.437136152 | 0.0005879 | 4.997959999 | 5.794E-07 | 5.484687909 | 4.142E-08 | 8.907534657 | 5.21799E-19 | known | TRAF3 | intronic | . | . | 14q32.32 | Y |
| rs4776729 | 15 | 66399197 | A | G | -6.070133552 | 1.27804E-09 | -4.472418169 | 0.000007734 | -2.322426162 | 0.02021 | 3.115553565 | 0.001836 | 5.371233205 | 7.82E-08 | -2.716280284 | 0.006602 | 0.588542235 | 0.556168394 | new | MEGF11 | intronic | . | . | 15q22.31 | Y |
| rs9302725 | 16 | 60716005 | T | G | 6.345997327 | 2.20989E-10 | 4.699891305 | 0.000002603 | 0.210471107 | 0.8333 | -1.378658729 | 0.168 | 6.208625583 | 5.345E-10 | 4.635195672 | 0.000003566 | -0.56539505 | 0.571805091 | new* | MIR4426 | intergenic | . | . | 16q21 | Y |
| rs511048 | 16 | 63056804 | A | G | -5.884576772 | 3.99074E-09 | -5.015375463 | 5.293E-07 | -1.775047769 | 0.07589 | 1.559830254 | 0.1188 | 4.57078526 | 0.000004859 | -1.78107622 | 0.0749 | 4.159837968 | 3.18473E-05 | new | CDH8 | intergenic | . | . | 16q21 | Y |
| rs2917688 | 16 | 64297174 | G | A | 5.873671128 | 4.26249E-09 | -6.037108739 | 1.569E-09 | 1.027893346 | 0.304 | -1.373812176 | 0.1695 | 3.880831378 | 0.0001041 | -1.605426393 | 0.1084 | -5.090716946 | 3.56712E-07 | known | NONE | intergenic | . | . | 16q21 | Y |
| rs285019 | 16 | 77099450 | G | A | -7.121421513 | 1.0682E-12 | 4.265337775 | 0.00001996 | -4.049213715 | 0.00005139 | 5.640152118 | 1.699E-08 | -4.594831066 | 0.000004331 | 3.323830953 | 0.0008879 | 1.189783577 | 0.234131466 | new* | MON1B | intergenic | . | . | 16q23.1 | Y |
| rs9933518 | 16 | 82867755 | A | G | 6.024275699 | 1.69869E-09 | 5.314586148 | 1.069E-07 | 1.20138988 | 0.2296 | -3.40452184 | 0.0006628 | -4.870884097 | 0.000001111 | 2.278614353 | 0.02269 | -1.544808931 | 0.122392486 | new | CDH13 | intronic | . | . | 16q23.3 | Y |
| rs8078510 | 17 | 47045862 | G | A | 6.184625298 | 6.22502E-10 | -5.489707455 | 4.026E-08 | 1.630480421 | 0.103 | -2.891260439 | 0.003837 | -2.932264521 | 0.003365 | -2.175669265 | 0.02958 | -7.114073072 | 1.12667E-12 | known | GIP | intronic | . | . | 17q21.32 | Y |
| rs12601661 | 17 | 50296779 | A | C | -6.194057848 | 5.86347E-10 | -7.153002593 | 8.49E-13 | -3.267504869 | 0.001085 | 1.836660703 | 0.06626 | 3.639281317 | 0.0002734 | -3.310799016 | 0.0009303 | 2.027064893 | 0.042655777 | known | CA10 | intergenic | . | . | 17q22 |  |
| rs12602912 | 17 | 65870073 | C | T | -9.300011526 | 1.4043E-20 | 4.243154079 | 0.00002204 | -3.25498088 | 0.001134 | 4.326350687 | 0.00001516 | 6.223559654 | 4.86E-10 | 4.718920027 | 0.000002371 | 7.859073936 | 3.86984E-15 | new* | BPTF | intronic | . | . | 17q24.2 | Y |
| rs9964724 | 18 | 35159124 | C | T | 7.785227079 | 6.95881E-15 | -6.165119346 | 7.043E-10 | 3.087717664 | 0.002017 | -3.958981711 | 0.00007527 | 6.950765488 | 3.633E-12 | -4.058211376 | 0.00004945 | 0.421434143 | 0.673438092 | known | CELF4 | intergenic | . | . | 18q12.2 | Y |
| rs1431196 | 18 | 50832102 | A | G | -10.40539438 | 2.34284E-25 | -5.546827189 | 2.909E-08 | -4.014151577 | 0.00005966 | 4.583708932 | 0.000004568 | 50 | 5.486E-20 | -5.974427203 | 2.309E-09 | 1.14725362 | 0.251276816 | known | DCC | intronic | . | . | 18q21.2 | Y |
| rs732779 | 18 | 53123172 | A | G | -7.677126198 | 1.62697E-14 | -5.44392217 | 5.212E-08 | -2.353252344 | 0.01861 | 4.321460219 | 0.0000155 | 7.745136553 | 9.579E-15 | -4.690078691 | 0.000002731 | -2.035896327 | 0.041760767 | new* | TCF4-AS1 | ncRNA_intronic | . | . | 18q21.2 | Y |
| rs17724992 | 19 | 18454825 | A | G | 8.745281681 | 2.2246E-18 | 6.89910821 | 5.233E-12 | 2.429971744 | 0.0151 | -4.738223453 | 0.000002156 | -4.485392013 | 0.000007278 | 3.554776857 | 0.0003783 | -9.198150679 | 3.64163E-20 | known | PGPEP1 | intronic | . | . | 19p13.11 | Y |
| rs6082289 | 20 | 21019305 | T | C | 6.201491895 | 5.59304E-10 | 3.956392831 | 0.00007609 | 5.962834162 | 2.479E-09 | -3.903325408 | 0.00009488 | 3.798626777 | 0.0001455 | 3.38973585 | 0.0006996 | -0.002698529 | 0.997846888 | new* | LINC00237 | intergenic | . | . | 20p11.23 | Y |
| rs8129315 | 21 | 46498863 | G | A | 8.360910178 | 6.22362E-17 | -4.402521465 | 0.0000107 | 3.612208757 | 0.0003036 | -2.175001553 | 0.02963 | -6.467238677 | 9.981E-11 | -3.34349562 | 0.0008273 | -6.687208059 | 2.27468E-11 | new* | ADARB1 | intronic | . | . | 21q22.3 | Y |
| rs5758264 | 22 | 41616284 | T | G | -8.100417257 | 5.4771E-16 | -3.913866144 | 0.00009083 | -1.144056997 | 0.2526 | 4.998526728 | 5.777E-07 | 7.425500116 | 1.124E-13 | -3.967140877 | 0.00007274 | 1.972905883 | 0.048506286 | new* | L3MBTL2 | intronic | . | . | 22q13.2 |  |

Note:

SNP: lead single-nucleotide polymorphism;

CHR: chromosome;

BP: base-pair position;

A1/A2: effect allele and non-effect allele;

Z_Estimate: Z-statistic from Genomic SEM common factor GWAS;

Pval_Estimate: P-value from Genomic SEM common factor GWAS;

GERD_z/GERD_pval: Z-statistic and P-value from GERD GWAS;

ADHD_z/ADHD_pval: Z-statistic and P-value from ADHD GWAS;

ANX_z/ANX_pval: Z-statistic and P-value from ANX GWAS;

MDD_z/MDD_pval: Z-statistic and P-value from MDD GWAS;

PTSD_z/PTSD_pval: Z-statistic and P-value from PTSD GWAS;

BMI_z/BMI_pval: Z-statistic and P-value from BMI GWAS;

annotation: locus novelty status (new, not previously reported in any GERD GWAS; new*, previously reported in psychiatric disorder GWAS but newly identified as GERD-associated);

Gene: nearest gene;

Function: functional annotation relative to RefSeq gene;

Exonic Function: exonic functional consequence;

CADD: Combined Annotation Dependent Depletion Phred-scaled score;

cytoBand: cytogenetic band location;

confdr_repeat: whether the locus was also identified by condFDR analysis (Y, yes; N, no).

**Table S9.** TDEP cell-type enrichment analysis of GERD-associated genetic signals in the human brain.

| **Cluster** | **P1** | **P2** | **Top Brain Regions** | **Anatomical Detail** | **Supercluster** | **Neurotransmitter** |
| --- | --- | --- | --- | --- | --- | --- |
| 266 | 1.97082E-08 | 0.003308302 | Cerebral cortex: 72.8%, Amygdala: 17.1%, Hippocampus: 5.1% | Cerebral cortex (Cx) - Anterior parahippocampal gyrus, posterior part (APH) - Medial entorhinal cortex - MEC | LAMP5-LHX6 and Chandelier | GABA |
| 163 | 1.86057E-07 | 0.001886296 | Amygdala: 84.9%, Hippocampus: 8.9%, Cerebral cortex: 6.2% | Amygdaloid complex (AMY) - basolateral nuclear group (BLN) - basolateral nucleus (basal nucleus) - BL | Amygdala excitatory | VGLUT1 VGLUT2 |
| 405 | 1.93782E-07 | 0.000698062 | Cerebral cortex: 94.4%, Amygdala: 2.5%, Basal forebrain: 1.2% | Cerebral cortex (Cx) - Gyrus rectus (ReG) - Medial orbitofrontal cortex - A14 | Miscellaneous | VGLUT1 |
| 263 | 2.68781E-07 | 0.04126772 | Cerebral cortex: 94.5%, Basal forebrain: 2.1%, Hippocampus: 1.8% | Cerebral cortex (Cx) - Precentral gyrus (PrCG) - Primary motor cortex - M1C | MGE interneuron | GABA |
| 287 | 3.24553E-07 | 0.034935298 | Cerebral cortex: 70.6%, Hippocampus: 13.8%, Amygdala: 10.2% | Cerebral cortex (Cx) - Anterior parahippocampal gyrus, posterior part (APH) - Medial entorhinal cortex - MEC | CGE interneuron | GABA |
| 265 | 3.78439E-07 | 0.020629425 | Cerebral cortex: 84.5%, Amygdala: 5.7%, Basal forebrain: 4.7% | Cerebral cortex (Cx) - Precentral gyrus (PrCG) - Primary motor cortex - M1C | LAMP5-LHX6 and Chandelier | GABA |
| 270 | 4.55828E-07 | 0.00652638 | Cerebral cortex: 43.7%, Hippocampus: 25.7%, Amygdala: 22.8% | Cerebral cortex (Cx) - Anterior parahippocampal gyrus, posterior part (APH) - Medial entorhinal cortex - MEC | LAMP5-LHX6 and Chandelier | GABA |
| 142 | 1.44963E-06 | 0.00168077 | Cerebral cortex: 95.4%, Basal forebrain: 2.0%, Hippocampus: 1.4% | Cerebral cortex (Cx) - Middle Temporal Gyrus - MTG | Deep-layer intratelencephalic | VGLUT1 VGLUT2 |
| 137 | 1.54057E-06 | 0.003129541 | Amygdala: 54.6%, Cerebral cortex: 36.2%, Hypothalamus: 8.6% | Amygdaloid complex (AMY) - Corticomedial nuclear group (CMN) - anterior cortical nucleus - CoA | Deep-layer intratelencephalic | VGLUT1 VGLUT2 |
| 406 | 1.54121E-06 | 0.001391653 | Cerebral cortex: 41.9%, Hypothalamus: 27.8%, Amygdala: 27.3% | Paleocortex (PalCx) - Anterior Olfactory Nucleus - AON | Amygdala excitatory | VGLUT1 VGLUT2 |
| 288 | 1.66797E-06 | 0.026461893 | Cerebral cortex: 53.6%, Amygdala: 17.5%, Hippocampus: 15.0% | Cerebral cortex (Cx) - Precentral gyrus (PrCG) - Primary motor cortex - M1C | CGE interneuron | GABA |
| 238 | 1.7929E-06 | 0.001578726 | Amygdala: 57.4%, Cerebral cortex: 20.2%, Basal forebrain: 8.4% | Amygdaloid complex (AMY) - basolateral nuclear group (BLN) - basolateral nucleus (basal nucleus) - BL | Splatter | GABA |
| 241 | 1.99912E-06 | 0.006745021 | Cerebral cortex: 92.3%, Amygdala: 3.0%, Hippocampus: 2.3% | Cerebral cortex (Cx) - Precentral gyrus (PrCG) - Primary motor cortex - M1C | MGE interneuron | GABA |
| 171 | 2.07631E-06 | 0.000624752 | Hippocampus: 68.6%, Hypothalamus: 21.7%, Cerebral cortex: 6.4% | Head of hippocampus (HiH) - CA1 | Miscellaneous | VGLUT1 VGLUT2 |
| 160 | 2.27431E-06 | 0.000487567 | Amygdala: 80.8%, Cerebral cortex: 17.6%, Hippocampus: 1.3% | Amygdaloid complex (AMY) - corticomedial nuclear group - CMN | Amygdala excitatory | VGLUT1 VGLUT2 |
| 274 | 2.59382E-06 | 0.032634733 | Amygdala: 44.9%, Cerebral cortex: 33.4%, Hippocampus: 7.3% | Amygdaloid complex (AMY) - basolateral nuclear group (BLN) - basolateral nucleus (basal nucleus) - BL | LAMP5-LHX6 and Chandelier | GABA |
| 253 | 2.66807E-06 | 0.115415138 | Cerebral cortex: 85.8%, Hippocampus: 8.0%, Basal forebrain: 3.6% | Cerebral cortex (Cx) - Precentral gyrus (PrCG) - Primary motor cortex - M1C | MGE interneuron | GABA VGLUT3 |
| 237 | 2.85441E-06 | 0.017753948 | Hippocampus: 61.2%, Cerebral cortex: 21.0%, Amygdala: 16.5% | Tail of Hippocampus (HiT) - Caudal Hippocampus - CA1-CA3 | MGE interneuron | GABA |
| 428 | 3.5539E-06 | 0.005541425 | Amygdala: 56.6%, Hypothalamus: 19.3%, Thalamus: 14.0% | Extended amygdala (EXA) - Bed nucleus of stria terminalis and nearby - BNST | Medium spiny neuron | GABA |
| 243 | 4.37051E-06 | 0.010827119 | Cerebral cortex: 90.8%, Hippocampus: 4.6%, Basal forebrain: 1.8% | Cerebral cortex (Cx) - Precentral gyrus (PrCG) - Primary motor cortex - M1C | MGE interneuron |  |
| 99 | 4.78987E-06 | 2.82458E-05 | Cerebral cortex: 75.6%, Basal forebrain: 10.3%, Amygdala: 7.8% | Cerebral cortex (Cx) - Middle Temporal Gyrus - MTG | Deep-layer corticothalamic and 6b | VGLUT1 |
| 140 | 5.13389E-06 | 0.000284363 | Cerebral cortex: 95.2%, Basal forebrain: 1.9%, Hippocampus: 1.5% | Cerebral cortex (Cx) - Middle Temporal Gyrus - MTG | Deep-layer intratelencephalic | VGLUT1 VGLUT2 |
| 84 | 6.13857E-06 | 0.001209652 | Cerebral cortex: 92.0%, Basal forebrain: 3.2%, Hypothalamus: 1.9% | Cerebral cortex (Cx) - Precentral gyrus (PrCG) - Primary motor cortex - M1C | Deep-layer near-projecting | VGLUT1 |
| 293 | 6.37342E-06 | 0.182101588 | Cerebral cortex: 86.1%, Hippocampus: 7.4%, Amygdala: 3.1% | Cerebral cortex (Cx) - Middle Temporal Gyrus - MTG | CGE interneuron | GABA |
| 267 | 7.16741E-06 | 0.029674322 | Amygdala: 42.8%, Hippocampus: 27.1%, Cerebral cortex: 25.1% | Amygdaloid complex (AMY) - corticomedial nuclear group - CMN | LAMP5-LHX6 and Chandelier | GABA |
| 268 | 7.56173E-06 | 0.01277914 | Hippocampus: 80.6%, Cerebral cortex: 13.8%, Amygdala: 1.9% | Tail of Hippocampus (HiT) - Caudal Hippocampus - CA4-DGC | LAMP5-LHX6 and Chandelier | GABA VGLUT3 |
| 100 | 7.8921E-06 | 0.000139891 | Cerebral cortex: 80.0%, Basal forebrain: 11.2%, Hippocampus: 6.5% | Cerebral cortex (Cx) - Frontal agranular insular cortex - FI | Deep-layer corticothalamic and 6b | VGLUT1 |
| 306 | 8.13758E-06 | 0.052735235 | Cerebellum: 88.0%, Pons: 4.7%, Midbrain: 3.7% | Cerebellum (CB) - Cerebellar Vermis - CBV | Cerebellar inhibitory | GABA |
| 308 | 1.00035E-05 | 0.141986682 | Cerebellum: 70.3%, Medulla: 12.1%, Pons: 11.7% | Cerebellum (CB) - Cerebellar Vermis - CBV | Cerebellar inhibitory | GABA |
| 128 | 1.01143E-05 | 0.000716359 | Cerebral cortex: 90.2%, Basal forebrain: 4.2%, Amygdala: 3.4% | Cerebral cortex (Cx) - Temporal pole (TP) - Temporopolar area - A38 | Upper-layer intratelencephalic | VGLUT1 VGLUT2 |
| 307 | 1.141E-05 | 0.058470884 | Cerebellum: 91.6%, Pons: 3.4%, Medulla: 3.3% | Cerebellum (CB) - Cerebellar Vermis - CBV | Cerebellar inhibitory | GABA |
| 139 | 1.19601E-05 | 0.007314057 | Cerebral cortex: 96.4%, Basal forebrain: 1.2%, Hippocampus: 1.2% | Cerebral cortex (Cx) - Middle Temporal Gyrus - MTG | Upper-layer intratelencephalic | VGLUT1 VGLUT2 |
| 254 | 1.22067E-05 | 0.033250326 | Cerebral cortex: 81.1%, Amygdala: 8.4%, Basal forebrain: 6.1% | Cerebral cortex (Cx) - Precentral gyrus (PrCG) - Primary motor cortex - M1C | MGE interneuron | GABA |
| 275 | 1.25235E-05 | 0.012412005 | Cerebral cortex: 84.6%, Basal forebrain: 5.9%, Hippocampus: 3.1% | Cerebral cortex (Cx) - Precentral gyrus (PrCG) - Primary motor cortex - M1C | LAMP5-LHX6 and Chandelier | GABA |
| 279 | 1.39701E-05 | 0.034494139 | Cerebral cortex: 78.0%, Amygdala: 10.4%, Hippocampus: 8.1% | Cerebral cortex (Cx) - Anterior parahippocampal gyrus, posterior part (APH) - Medial entorhinal cortex - MEC | CGE interneuron | GABA VGLUT3 |
| 101 | 1.54476E-05 | 1.43112E-05 | Cerebral cortex: 63.0%, Amygdala: 27.0%, Basal forebrain: 5.9% | Paleocortex (PalCx) - Anterior Olfactory Nucleus - AON | Deep-layer corticothalamic and 6b | VGLUT1 |
| 290 | 1.78817E-05 | 0.085192095 | Cerebral cortex: 82.1%, Hippocampus: 7.9%, Amygdala: 5.8% | Cerebral cortex (Cx) - Precentral gyrus (PrCG) - Primary motor cortex - M1C | CGE interneuron | GABA |
| 145 | 1.84935E-05 | 0.005977553 | Cerebral cortex: 96.2%, Basal forebrain: 1.4%, Amygdala: 1.2% | Cerebral cortex (Cx) - Middle Temporal Gyrus - MTG | Deep-layer intratelencephalic | VGLUT1 |
| 92 | 1.85951E-05 | 0.000222962 | Cerebral cortex: 89.4%, Basal forebrain: 4.7%, Hippocampus: 3.3% | Cerebral cortex (Cx) - Middle Temporal Gyrus - MTG | Deep-layer near-projecting | VGLUT1 |
| 201 | 1.92792E-05 | 0.03747494 | Hippocampus: 99.4%, Thalamus: 0.5%, Midbrain: 0.1% | Tail of Hippocampus (HiT) - Caudal Hippocampus - CA4-DGC | Hippocampal dentate gyrus | VGLUT1 |
| 165 | 1.95696E-05 | 0.0154649 | Cerebral cortex: 66.8%, Amygdala: 18.2%, Hippocampus: 15.0% | Cerebral cortex (Cx) - Anterior parahippocampal gyrus (AG) - Lateral entorhinal cortex - LEC | Miscellaneous | VGLUT1 VGLUT2 |
| 155 | 2.10578E-05 | 0.009650185 | Amygdala: 87.5%, Cerebral cortex: 6.8%, Hippocampus: 5.7% | Amygdaloid complex (AMY) - basolateral nuclear group (BLN) - basolateral nucleus (basal nucleus) - BL | Amygdala excitatory | VGLUT1 VGLUT2 |
| 294 | 2.20142E-05 | 0.033338101 | Cerebral cortex: 70.3%, Hippocampus: 15.1%, Amygdala: 8.5% | Cerebral cortex (Cx) - Anterior parahippocampal gyrus, posterior part (APH) - Medial entorhinal cortex - MEC | CGE interneuron | GABA |
| 249 | 2.44619E-05 | 0.014985088 | Cerebral cortex: 78.8%, Hippocampus: 12.5%, Amygdala: 5.0% | Cerebral cortex (Cx) - Lingual gyrus (LiG) - Primary Visual Cortex - V1C | MGE interneuron | GABA |
| 178 | 2.52975E-05 | 0.007019731 | Hippocampus: 68.2%, Midbrain: 20.5%, Cerebral cortex: 10.8% | Head of hippocampus (HiH) - Tail of Hippocampus (HiT) - Subicular cortex - Sub | Miscellaneous | VGLUT1 VGLUT2 |
| 141 | 2.88324E-05 | 0.002231986 | Cerebral cortex: 99.2%, Hippocampus: 0.5%, Basal forebrain: 0.2% | Cerebral cortex (Cx) - Precentral gyrus (PrCG) - Primary motor cortex - M1C | Deep-layer intratelencephalic | VGLUT1 VGLUT2 |
| 260 | 2.89908E-05 | 0.027797578 | Cerebral cortex: 89.3%, Amygdala: 3.4%, Hippocampus: 3.3% | Cerebral cortex (Cx) - Middle Temporal Gyrus - MTG | MGE interneuron | GABA |
| 250 | 2.9401E-05 | 0.01453418 | Cerebral cortex: 61.8%, Hippocampus: 23.3%, Amygdala: 10.2% | Cerebral cortex (Cx) - Anterior parahippocampal gyrus, posterior part (APH) - Medial entorhinal cortex - MEC | MGE interneuron | GABA |
| 256 | 3.11757E-05 | 0.032537616 | Cerebral cortex: 67.2%, Amygdala: 13.0%, Hippocampus: 12.1% | Cerebral cortex (Cx) - Precentral gyrus (PrCG) - Primary motor cortex - M1C | MGE interneuron | GABA |
| 262 | 3.31503E-05 | 0.042061444 | Cerebral cortex: 95.5%, Hippocampus: 1.4%, Basal forebrain: 1.4% | Cerebral cortex (Cx) - Precentral gyrus (PrCG) - Primary motor cortex - M1C | MGE interneuron | GABA |
| 261 | 3.54193E-05 | 0.036419751 | Cerebral cortex: 86.4%, Amygdala: 4.6%, Basal forebrain: 4.2% | Cerebral cortex (Cx) - Anterior parahippocampal gyrus, posterior part (APH) - Medial entorhinal cortex - MEC | MGE interneuron | GABA |
| 273 | 3.55726E-05 | 0.022765516 | Cerebral cortex: 67.5%, Amygdala: 16.1%, Basal forebrain: 7.8% | Cerebral cortex (Cx) - Anterior parahippocampal gyrus, posterior part (APH) - Medial entorhinal cortex - MEC | LAMP5-LHX6 and Chandelier | GABA |
| 282 | 3.67385E-05 | 0.021653475 | Cerebral cortex: 70.5%, Amygdala: 13.0%, Hippocampus: 10.6% | Paleocortex (PalCx) - Piriform cortex - Pir | CGE interneuron | GABA |
| 278 | 3.81569E-05 | 0.006556143 | Cerebral cortex: 77.4%, Amygdala: 13.3%, Hippocampus: 6.1% | Cerebral cortex (Cx) - Precentral gyrus (PrCG) - Primary motor cortex - M1C | CGE interneuron | GABA VGLUT3 |
| 159 | 4.19888E-05 | 0.002951842 | Amygdala: 68.1%, Cerebral cortex: 25.8%, Hippocampus: 6.0% | Amygdaloid complex (AMY) - corticomedial nuclear group - CMN | Amygdala excitatory | VGLUT1 VGLUT2 |
| 297 | 4.66033E-05 | 0.131543537 | Cerebral cortex: 72.9%, Amygdala: 10.5%, Hippocampus: 9.0% | Cerebral cortex (Cx) - Precentral gyrus (PrCG) - Primary motor cortex - M1C | CGE interneuron | GABA |
| 138 | 4.67181E-05 | 0.000813992 | Cerebral cortex: 94.4%, Basal forebrain: 2.1%, Hippocampus: 1.9% | Cerebral cortex (Cx) - Middle Temporal Gyrus - MTG | Deep-layer intratelencephalic | VGLUT1 VGLUT2 |
| 272 | 4.7092E-05 | 0.022244782 | Cerebral cortex: 63.9%, Amygdala: 16.6%, Hippocampus: 9.9% | Cerebral cortex (Cx) - Anterior parahippocampal gyrus, posterior part (APH) - Medial entorhinal cortex - MEC | LAMP5-LHX6 and Chandelier | GABA |
| 277 | 4.9357E-05 | 0.110716862 | Hippocampus: 73.7%, Amygdala: 13.0%, Cerebral cortex: 10.8% | Body of hippocampus (HiB) - Rostral CA3 | CGE interneuron | GABA VGLUT3 |
| 107 | 5.0236E-05 | 0.000273113 | Cerebral cortex: 99.0%, Hippocampus: 0.6%, Basal forebrain: 0.4% | Cerebral cortex (Cx) - Precentral gyrus (PrCG) - Primary motor cortex - M1C | Deep-layer corticothalamic and 6b | VGLUT1 |
| 239 | 5.11977E-05 | 0.072602863 | Basal forebrain: 57.1%, Amygdala: 27.7%, Cerebral cortex: 6.8% | Basal nuclei (BN) - Body of the Caudate - CaB | Splatter | GABA |
| 242 | 5.5136E-05 | 0.048670645 | Cerebral cortex: 90.2%, Basal forebrain: 3.5%, Hippocampus: 2.5% | Cerebral cortex (Cx) - Middle Temporal Gyrus - MTG | MGE interneuron | GABA |
| 102 | 5.62387E-05 | 0.000745931 | Cerebral cortex: 65.6%, Hippocampus: 29.5%, Amygdala: 4.8% | Cerebral cortex (Cx) - Anterior parahippocampal gyrus, posterior part (APH) - Medial entorhinal cortex - MEC | Deep-layer corticothalamic and 6b | VGLUT1 |
| 289 | 5.83231E-05 | 0.115632768 | Cerebral cortex: 70.6%, Amygdala: 13.7%, Hippocampus: 6.3% | Cerebral cortex (Cx) - Precentral gyrus (PrCG) - Primary motor cortex - M1C | CGE interneuron | GABA |
| 152 | 6.00788E-05 | 0.000743544 | Cerebral cortex: 44.8%, Amygdala: 33.6%, Basal forebrain: 21.4% | Claustrum - Cla | Deep-layer intratelencephalic | VGLUT1 VGLUT2 VGLUT3 |
| 136 | 6.21699E-05 | 0.011102119 | Cerebral cortex: 95.4%, Basal forebrain: 2.7%, Hippocampus: 1.0% | Cerebral cortex (Cx) - Precentral gyrus (PrCG) - Primary motor cortex - M1C | Upper-layer intratelencephalic | VGLUT1 VGLUT2 |
| 120 | 7.46464E-05 | 0.003732703 | Hippocampus: 97.3%, Amygdala: 1.5%, Cerebral cortex: 1.3% | Head of hippocampus (HiH) - CA1 | Hippocampal CA1-3 | VGLUT1 |
| 104 | 7.54175E-05 | 0.000470733 | Cerebral cortex: 62.0%, Hippocampus: 25.8%, Amygdala: 12.1% | Cerebral cortex (Cx) - Anterior parahippocampal gyrus, posterior part (APH) - Medial entorhinal cortex - MEC | Deep-layer corticothalamic and 6b | VGLUT1 |
| 191 | 8.2353E-05 | 0.058375917 | Hippocampus: 100.0%, Cerebral cortex: 0.0%, Amygdala: 0.0% | Body of hippocampus (HiB) - Rostral CA3 | Hippocampal CA4 | VGLUT1 |
| 115 | 8.83364E-05 | 0.242968411 | Cerebral cortex: 98.4%, Hippocampus: 1.2%, Basal forebrain: 0.3% | Cerebral cortex (Cx) - Precentral gyrus (PrCG) - Primary motor cortex - M1C | Miscellaneous | VGLUT1 |
| 244 | 8.87851E-05 | 0.006254309 | Cerebral cortex: 96.3%, Hippocampus: 1.4%, Basal forebrain: 1.1% | Cerebral cortex (Cx) - Middle Temporal Gyrus - MTG | MGE interneuron | GABA |
| 223 | 9.45585E-05 | 0.01313047 | Basal forebrain: 93.9%, Thalamus: 1.9%, Cerebral cortex: 1.8% | Basal nuclei (BN) - Putamen - Pu | Eccentric medium spiny neuron | GABA |
| 284 | 9.85586E-05 | 0.090419758 | Cerebral cortex: 81.0%, Hippocampus: 9.3%, Amygdala: 5.0% | Cerebral cortex (Cx) - Middle Temporal Gyrus - MTG | CGE interneuron | GABA |
| 118 | 9.86076E-05 | 0.015084964 | Cerebral cortex: 82.3%, Basal forebrain: 7.9%, Hypothalamus: 7.7% | Cerebral cortex (Cx) - Anterior cingulate cortex - ACC | Miscellaneous | VGLUT1 |
| 91 | 0.000106561 | 0.00018995 | Cerebral cortex: 95.7%, Basal forebrain: 3.4%, Hypothalamus: 0.6% | Cerebral cortex (Cx) - Precentral gyrus (PrCG) - Primary motor cortex - M1C | Deep-layer near-projecting | VGLUT1 |
| 86 | 0.00012647 | 0.000171287 | Cerebral cortex: 47.1%, Basal forebrain: 37.0%, Hypothalamus: 10.9% | Cerebral cortex (Cx) - Anterior cingulate cortex - ACC | Deep-layer near-projecting | VGLUT1 VGLUT2 VGLUT3 |
| 162 | 0.00012758 | 0.009949619 | Amygdala: 66.2%, Cerebral cortex: 31.6%, Hippocampus: 1.3% | Amygdaloid complex (AMY) - corticomedial nuclear group - CMN | Amygdala excitatory | VGLUT1 VGLUT2 |
| 147 | 0.000130989 | 0.000788471 | Amygdala: 65.4%, Cerebral cortex: 33.1%, Hypothalamus: 0.8% | Paleocortex (PalCx) - Piriform cortex - Pir | Deep-layer intratelencephalic | VGLUT1 VGLUT2 |
| 255 | 0.000137683 | 0.10186244 | Cerebral cortex: 77.9%, Hippocampus: 13.0%, Amygdala: 4.5% | Cerebral cortex (Cx) - Precentral gyrus (PrCG) - Primary motor cortex - M1C | MGE interneuron | GABA VGLUT3 |
| 135 | 0.000146092 | 0.211546975 | Cerebral cortex: 99.3%, Hippocampus: 0.5%, Basal forebrain: 0.2% | Cerebral cortex (Cx) - Precentral gyrus (PrCG) - Primary motor cortex - M1C | Upper-layer intratelencephalic | VGLUT1 VGLUT2 |
| 88 | 0.000146206 | 5.57507E-05 | Cerebral cortex: 83.9%, Basal forebrain: 12.2%, Hypothalamus: 2.8% | Cerebral cortex (Cx) - Anterior cingulate cortex - ACC | Deep-layer near-projecting | VGLUT1 VGLUT2 |
| 197 | 0.000147249 | 0.062632663 | Hippocampus: 99.9%, Midbrain: 0.1%, Cerebral cortex: 0.0% | Tail of Hippocampus (HiT) - Caudal Hippocampus - CA4-DGC | Hippocampal CA4 | VGLUT1 |
| 144 | 0.000148245 | 0.009096421 | Cerebral cortex: 89.0%, Basal forebrain: 4.2%, Amygdala: 4.2% | Cerebral cortex (Cx) - Anterior parahippocampal gyrus, posterior part (APH) - Medial entorhinal cortex - MEC | Deep-layer intratelencephalic | VGLUT1 |
| 419 | 0.000159085 | 0.030596962 | Hypothalamus: 99.3%, Midbrain: 0.5%, Thalamus: 0.1% | Hypothalamus (HTH) - supraoptic region of HTH - HTHso (anterior hypothalamic nucleus, AHN) - tuberal region of HTH - HTHtub (ventromedial and dorsomedial hypothalamic nucleic nuclei, VMH, DMH) | Splatter | HDC VGLUT2 |
| 225 | 0.000163756 | 0.027775236 | Basal forebrain: 70.9%, Amygdala: 13.6%, Cerebral cortex: 8.2% | Basal forebrain (BF) - substantia innominata and nearby nuclei - SI | Eccentric medium spiny neuron | GABA |
| 422 | 0.000165666 | 0.033191274 | Hypothalamus: 64.1%, Amygdala: 19.7%, Thalamus: 14.8% | Hypothalamus (HTH) - preoptic region of HTH - HTHpo | Splatter | GABA |
| 220 | 0.000166159 | 0.037506396 | Amygdala: 33.2%, Basal forebrain: 32.8%, Hypothalamus: 18.6% | Basal forebrain (BF) - substantia innominata and nearby nuclei - SI | Medium spiny neuron | GABA |
| 218 | 0.000168056 | 0.001523913 | Amygdala: 69.5%, Basal forebrain: 15.7%, Hypothalamus: 7.4% | Amygdaloid complex (AMY) - Central nuclear group - CEN | Medium spiny neuron | GABA |
| 200 | 0.000180502 | 0.145145784 | Hippocampus: 96.8%, Thalamus: 3.2%, Cerebral cortex: 0.0% | Tail of Hippocampus (HiT) - Caudal Hippocampus - CA4-DGC | Hippocampal dentate gyrus | VGLUT1 |
| 388 | 0.000182366 | 0.023303162 | Pons: 89.9%, Midbrain: 8.9%, Medulla: 0.7% | Pons (Pn) - Pontine reticular formation (PnRF) - parabrachial nuclei - PB | Splatter | VGLUT2 |
| 291 | 0.000182713 | 0.034302395 | Cerebral cortex: 83.0%, Hippocampus: 8.5%, Amygdala: 5.2% | Cerebral cortex (Cx) - Middle Temporal Gyrus - MTG | CGE interneuron | GABA |
| 276 | 0.00019302 | 0.049478139 | Cerebral cortex: 66.0%, Amygdala: 16.5%, Basal forebrain: 9.9% | Claustrum - Cla | LAMP5-LHX6 and Chandelier | GABA |
| 285 | 0.000194117 | 0.080018418 | Cerebral cortex: 74.7%, Amygdala: 9.9%, Hippocampus: 9.3% | Cerebral cortex (Cx) - Precentral gyrus (PrCG) - Primary motor cortex - M1C | CGE interneuron | GABA |
| 149 | 0.000209231 | 0.00031708 | Cerebral cortex: 85.2%, Basal forebrain: 6.0%, Amygdala: 5.0% | Cerebral cortex (Cx) - Middle Temporal Gyrus - MTG | Deep-layer intratelencephalic | VGLUT1 |
| 240 | 0.000211123 | 0.008325504 | Cerebral cortex: 95.4%, Hippocampus: 1.5%, Basal forebrain: 1.2% | Cerebral cortex (Cx) - Precentral gyrus (PrCG) - Primary motor cortex - M1C | MGE interneuron | GABA |
| 156 | 0.000212815 | 0.005469515 | Amygdala: 99.6%, Cerebral cortex: 0.2%, Hippocampus: 0.2% | Amygdaloid complex (AMY) - Basolateral nuclear group (BLN) - lateral nucleus - La | Amygdala excitatory | VGLUT1 VGLUT2 |
| 425 | 0.000222907 | 0.046389817 | Amygdala: 34.9%, Hypothalamus: 32.8%, Thalamus: 29.6% | Thalamus (THM) - Anterior nuclear complex - ANC | Splatter | GABA |
| 203 | 0.000238037 | 0.024330121 | Hippocampus: 100.0%, Cerebral cortex: 0.0%, Amygdala: 0.0% | Body of hippocampus (HiB) - Rostral DG-CA4 | Hippocampal dentate gyrus | VGLUT1 |
| 251 | 0.000238267 | 0.003078753 | Cerebral cortex: 92.5%, Hippocampus: 2.4%, Basal forebrain: 2.2% | Cerebral cortex (Cx) - Anterior parahippocampal gyrus, posterior part (APH) - Medial entorhinal cortex - MEC | MGE interneuron | GABA |
| 408 | 0.00024442 | 0.000851742 | Amygdala: 72.6%, Cerebral cortex: 23.5%, Hippocampus: 2.7% | Amygdaloid complex (AMY) - Central nuclear group - CEN | Amygdala excitatory | VGLUT2 VGLUT3 |
| 97 | 0.000250655 | 0.000272341 | Cerebral cortex: 85.1%, Amygdala: 7.0%, Basal forebrain: 5.7% | Cerebral cortex (Cx) - Temporal pole (TP) - Temporopolar area - A38 | Deep-layer near-projecting | VGLUT1 VGLUT2 |
| 199 | 0.000260143 | 0.008768775 | Hippocampus: 89.9%, Midbrain: 10.1%, Cerebral cortex: 0.0% | Tail of Hippocampus (HiT) - Caudal Hippocampus - CA4-DGC | Hippocampal CA4 | VGLUT1 VGLUT2 |
| 271 | 0.000272738 | 0.04249231 | Cerebral cortex: 86.2%, Hippocampus: 6.7%, Thalamus: 3.3% | Cerebral cortex (Cx) - Precentral gyrus (PrCG) - Primary motor cortex - M1C | LAMP5-LHX6 and Chandelier | GABA |
| 252 | 0.000278835 | 0.128125324 | Cerebral cortex: 94.0%, Hippocampus: 2.5%, Basal forebrain: 2.4% | Cerebral cortex (Cx) - Middle Temporal Gyrus - MTG | MGE interneuron | GABA |
| 458 | 0.000290865 | 0.005111529 | Thalamus: 98.3%, Amygdala: 1.4%, Cerebral cortex: 0.3% | Thalamus (THM) - Anterior nuclear complex - ANC | Thalamic excitatory | VGLUT2 |
| 292 | 0.000291209 | 0.191091215 | Cerebral cortex: 75.1%, Hippocampus: 14.3%, Amygdala: 7.1% | Cerebral cortex (Cx) - Middle Temporal Gyrus - MTG | CGE interneuron | GABA |
| 198 | 0.000293638 | 0.060454954 | Hippocampus: 99.7%, Midbrain: 0.3%, Cerebral cortex: 0.0% | Body of hippocampus (HiB) - Rostral DG-CA4 | Hippocampal CA4 | VGLUT1 |
| 372 | 0.000295206 | 0.201623113 | Midbrain: 99.7%, Cerebellum: 0.3%, Cerebral cortex: 0.0% | Midbrain (M) - Superior colliculus and nearby nuclei - SC | Splatter | VGLUT2 |
| 283 | 0.000300465 | 0.047433654 | Cerebral cortex: 82.6%, Hippocampus: 11.9%, Amygdala: 2.2% | Cerebral cortex (Cx) - Rostral gyrus (RoG) - Dorsal division of MFC - A32 | CGE interneuron | GABA |
| 269 | 0.000314407 | 0.021020492 | Hippocampus: 95.5%, Cerebral cortex: 2.4%, Amygdala: 1.1% | Head of hippocampus (HiH) - Uncal CA1-CA3 | LAMP5-LHX6 and Chandelier | GABA |
| 259 | 0.000315555 | 0.116298507 | Cerebral cortex: 80.1%, Hippocampus: 6.7%, Amygdala: 6.3% | Cerebral cortex (Cx) - Precentral gyrus (PrCG) - Primary motor cortex - M1C | MGE interneuron | GABA VGLUT3 |
| 106 | 0.00031745 | 0.004543909 | Cerebral cortex: 80.5%, Basal forebrain: 8.4%, Hippocampus: 7.2% | Cerebral cortex (Cx) - Anterior cingulate cortex - ACC | Deep-layer corticothalamic and 6b | VGLUT1 |
| 94 | 0.000330651 | 0.000862911 | Cerebral cortex: 97.2%, Basal forebrain: 1.1%, Hypothalamus: 0.7% | Cerebral cortex (Cx) - Precentral gyrus (PrCG) - Primary motor cortex - M1C | Deep-layer near-projecting | VGLUT1 |
| 148 | 0.000379522 | 0.000533631 | Cerebral cortex: 65.1%, Amygdala: 19.4%, Basal forebrain: 11.8% | Claustrum - Cla | Deep-layer intratelencephalic | VGLUT1 |
| 229 | 0.000402547 | 0.00098024 | Amygdala: 83.7%, Cerebral cortex: 12.2%, Thalamus: 1.9% | Amygdaloid complex (AMY) - Basolateral nuclear group (BLN) - lateral nucleus - La | Eccentric medium spiny neuron | GABA |
| 143 | 0.000408339 | 0.017398699 | Cerebral cortex: 96.0%, Hippocampus: 1.3%, Basal forebrain: 1.2% | Cerebral cortex (Cx) - Precentral gyrus (PrCG) - Primary motor cortex - M1C | Deep-layer intratelencephalic | VGLUT1 |
| 150 | 0.000416773 | 0.001305266 | Cerebral cortex: 98.6%, Basal forebrain: 0.6%, Hippocampus: 0.5% | Cerebral cortex (Cx) - Precentral gyrus (PrCG) - Primary motor cortex - M1C | Deep-layer intratelencephalic | VGLUT1 |
| 222 | 0.000431668 | 0.004065926 | Basal forebrain: 78.8%, Amygdala: 9.0%, Cerebral cortex: 6.5% | Basal forebrain (BF) - substantia innominata and nearby nuclei - SI | Medium spiny neuron | GABA |
| 117 | 0.000444356 | 0.154611734 | Cerebral cortex: 93.4%, Amygdala: 3.3%, Basal forebrain: 2.6% | Cerebral cortex (Cx) - Long insular gyri (LIG) - Dysgranular insular cortex - Idg | Miscellaneous | VGLUT1 |
| 89 | 0.000452362 | 0.000396443 | Cerebral cortex: 92.2%, Basal forebrain: 4.8%, Amygdala: 1.9% | Cerebral cortex (Cx) - Middle Temporal Gyrus - MTG | Deep-layer near-projecting | VGLUT1 |
| 196 | 0.000473482 | 0.111205438 | Hippocampus: 99.4%, Thalamus: 0.5%, Cerebral cortex: 0.1% | Tail of Hippocampus (HiT) - Caudal Hippocampus - CA4-DGC | Hippocampal CA4 | VGLUT1 |
| 172 | 0.000520918 | 0.011344692 | Hippocampus: 93.7%, Amygdala: 5.7%, Cerebral cortex: 0.3% | Head of hippocampus (HiH) - CA1 | Amygdala excitatory | VGLUT1 VGLUT2 |
| 154 | 0.000540163 | 0.024696694 | Amygdala: 93.4%, Hippocampus: 5.3%, Cerebral cortex: 1.3% | Amygdaloid complex (AMY) - basolateral nuclear group (BLN) - basolateral nucleus (basal nucleus) - BL | Amygdala excitatory | VGLUT1 VGLUT2 |
| 109 | 0.000553203 | 0.004019824 | Cerebral cortex: 89.4%, Hippocampus: 6.0%, Basal forebrain: 2.1% | Cerebral cortex (Cx) - Precentral gyrus (PrCG) - Primary motor cortex - M1C | Deep-layer corticothalamic and 6b | VGLUT1 |
| 219 | 0.000559683 | 0.038787982 | Basal forebrain: 50.9%, Amygdala: 22.3%, Hypothalamus: 18.5% | Basal forebrain (BF) - substantia innominata and nearby nuclei - SI | Medium spiny neuron |  |
| 403 | 0.000591354 | 0.344360232 | Midbrain: 58.2%, Hypothalamus: 41.6%, Amygdala: 0.1% | Midbrain (M) - Substantia Nigra - SN | Splatter | GABA |
| 151 | 0.000598638 | 0.014851316 | Cerebral cortex: 95.3%, Hippocampus: 2.2%, Basal forebrain: 1.3% | Cerebral cortex (Cx) - Precentral gyrus (PrCG) - Primary motor cortex - M1C | Deep-layer intratelencephalic | VGLUT1 VGLUT2 |
| 127 | 0.000605018 | 0.000762922 | Cerebral cortex: 90.8%, Basal forebrain: 3.3%, Hypothalamus: 2.9% | Cerebral cortex (Cx) - Frontal agranular insular cortex - FI | Upper-layer intratelencephalic | VGLUT1 VGLUT2 |
| 167 | 0.000613372 | 0.007298849 | Cerebral cortex: 79.4%, Hippocampus: 16.1%, Amygdala: 3.1% | Cerebral cortex (Cx) - Anterior parahippocampal gyrus, posterior part (APH) - Medial entorhinal cortex - MEC | Miscellaneous | VGLUT1 VGLUT2 |
| 121 | 0.000629231 | 0.022154087 | Cerebral cortex: 91.5%, Hypothalamus: 4.6%, Basal forebrain: 2.9% | Cerebral cortex (Cx) - Frontal agranular insular cortex - FI | Upper-layer intratelencephalic | VGLUT1 VGLUT2 |
| 111 | 0.00064417 | 0.001636365 | Cerebral cortex: 84.7%, Basal forebrain: 6.3%, Amygdala: 5.0% | Cerebral cortex (Cx) - Frontal agranular insular cortex - FI | Deep-layer corticothalamic and 6b | VGLUT1 |
| 258 | 0.000658019 | 0.15330048 | Cerebral cortex: 73.5%, Hippocampus: 19.7%, Amygdala: 3.5% | Cerebral cortex (Cx) - Precentral gyrus (PrCG) - Primary motor cortex - M1C | MGE interneuron | GABA |
| 248 | 0.000669106 | 0.089991048 | Hippocampus: 91.0%, Amygdala: 4.8%, Cerebral cortex: 3.4% | Tail of Hippocampus (HiT) - Caudal Hippocampus - CA1-CA3 | MGE interneuron | GABA |
| 295 | 0.000684328 | 0.060597054 | Cerebral cortex: 64.4%, Hippocampus: 14.7%, Amygdala: 10.8% | Cerebral cortex (Cx) - Anterior parahippocampal gyrus, posterior part (APH) - Medial entorhinal cortex - MEC | CGE interneuron | GABA |
| 226 | 0.00071828 | 0.067341481 | Amygdala: 47.2%, Basal forebrain: 33.3%, Cerebral cortex: 10.2% | Basal forebrain (BF) - substantia innominata and nearby nuclei - SI | Eccentric medium spiny neuron | GABA |
| 429 | 0.000739063 | 0.032437767 | Basal forebrain: 59.7%, Amygdala: 16.1%, Hypothalamus: 13.8% | Basal forebrain (BF) - septal nuclei - SEP | Splatter | GABA |
| 397 | 0.000741324 | 0.098179066 | Pons: 66.2%, Midbrain: 31.2%, Hypothalamus: 1.2% | Pons (Pn) - other nuclei in pontine tegmentum (XPnTg) - dorsal tegmental nucleus - DTg | Splatter | VGLUT3 |
| 404 | 0.000758321 | 0.009626708 | Cerebral cortex: 45.6%, Amygdala: 33.5%, Hypothalamus: 9.8% | Paleocortex (PalCx) - Anterior Olfactory Nucleus - AON | Splatter | GABA |
| 93 | 0.000796217 | 0.000756523 | Cerebral cortex: 94.7%, Hippocampus: 2.5%, Basal forebrain: 2.2% | Cerebral cortex (Cx) - Middle Temporal Gyrus - MTG | Deep-layer near-projecting | VGLUT1 |
| 133 | 0.000834985 | 0.115931607 | Cerebral cortex: 98.4%, Hippocampus: 0.8%, Midbrain: 0.8% | Cerebral cortex (Cx) - Cingulate gyrus, retrosplenial (CgGrs) - A29-A30 | Miscellaneous | VGLUT1 VGLUT2 |
| 181 | 0.000858118 | 0.114630973 | Hippocampus: 99.6%, Cerebral cortex: 0.4%, Amygdala: 0.0% | Tail of Hippocampus (HiT) - Caudal Hippocampus - CA1-CA3 | Hippocampal CA1-3 | VGLUT1 |
| 400 | 0.000870302 | 0.274818313 | Medulla: 99.3%, Pons: 0.5%, Cerebral cortex: 0.1% | Myelencephalon (medulla oblongata) (Mo) - afferent nuclei of cranial nerves in medulla oblongata - MoAN | Splatter | VGLUT2 |
| 146 | 0.000901158 | 0.003455492 | Cerebral cortex: 65.1%, Hippocampus: 19.6%, Amygdala: 15.0% | Cerebral cortex (Cx) - Anterior parahippocampal gyrus, posterior part (APH) - Medial entorhinal cortex - MEC | Deep-layer intratelencephalic | VGLUT1 VGLUT2 |
| 53 | 0.000903258 | 0.494650232 | Cerebral cortex: 90.3%, Hypothalamus: 3.9%, Hippocampus: 2.2% | Cerebral cortex (Cx) - Middle Temporal Gyrus - MTG | Astrocyte |  |
| 114 | 0.000925651 | 0.315987484 | Cerebral cortex: 100.0%, Amygdala: 0.0%, Hippocampus: 0.0% | Cerebral cortex (Cx) - Precentral gyrus (PrCG) - Primary motor cortex - M1C | Miscellaneous | VGLUT1 |
| 96 | 0.000964696 | 0.000512473 | Cerebral cortex: 89.0%, Basal forebrain: 7.0%, Hypothalamus: 3.3% | Cerebral cortex (Cx) - Subcallosal Gyrus (SCG) - Subgenual (subcallosal) division of MFC - A25 | Deep-layer near-projecting | VGLUT1 |
| 280 | 0.000973811 | 0.549929413 | Cerebral cortex: 54.6%, Hippocampus: 26.0%, Amygdala: 14.6% | Cerebral cortex (Cx) - Anterior parahippocampal gyrus, posterior part (APH) - Medial entorhinal cortex - MEC | CGE interneuron | GABA VGLUT3 |
| 170 | 0.001034058 | 0.115720248 | Hippocampus: 98.4%, Cerebral cortex: 1.1%, Amygdala: 0.2% | Head of hippocampus (HiH) - Uncal CA1-CA3 | Hippocampal CA1-3 | VGLUT1 |
| 296 | 0.001040978 | 0.205016618 | Cerebral cortex: 67.0%, Amygdala: 17.8%, Hippocampus: 8.6% | Cerebral cortex (Cx) - Precentral gyrus (PrCG) - Primary motor cortex - M1C | CGE interneuron | GABA |
| 281 | 0.001065152 | 0.264283481 | Cerebral cortex: 73.7%, Amygdala: 13.3%, Hippocampus: 7.8% | Cerebral cortex (Cx) - Anterior parahippocampal gyrus, posterior part (APH) - Medial entorhinal cortex - MEC | CGE interneuron | GABA |
| 450 | 0.001088975 | 0.049190493 | Thalamus: 98.7%, Amygdala: 1.0%, Midbrain: 0.3% | Thalamus (THM) - lateral nuclear complex of thalamus (LNC) - lateral posterior nucleus of thalamus - LP | Thalamic excitatory | VGLUT2 |
| 202 | 0.001093783 | 0.158052059 | Hippocampus: 99.9%, Midbrain: 0.1%, Thalamus: 0.0% | Head of hippocampus (HiH) - Uncal DG-CA4 | Hippocampal dentate gyrus | VGLUT1 |
| 423 | 0.001094924 | 0.045798093 | Hypothalamus: 44.2%, Amygdala: 23.2%, Thalamus: 20.2% | Extended amygdala (EXA) - Bed nucleus of stria terminalis and nearby - BNST | Splatter | GABA |
| 316 | 0.00112139 | 0.657871626 | Pons: 95.9%, Medulla: 1.9%, Cerebral cortex: 1.5% | Pons (Pn) - efferent nuclei of cranial nerves in pons - PnEN | Lower rhombic lip | VGLUT1 VGLUT2 |
| 179 | 0.001171079 | 0.021040127 | Hippocampus: 43.8%, Midbrain: 29.4%, Cerebral cortex: 26.8% | Head of hippocampus (HiH) - Tail of Hippocampus (HiT) - Subicular cortex - Sub | Miscellaneous | VGLUT1 VGLUT2 |
| 108 | 0.001194075 | 0.048655033 | Cerebral cortex: 100.0%, Amygdala: 0.0%, Hippocampus: 0.0% | Cerebral cortex (Cx) - Lingual gyrus (LiG) - Primary Visual Cortex - V1C | Deep-layer corticothalamic and 6b | VGLUT1 |
| 174 | 0.001215398 | 0.034474042 | Hippocampus: 55.8%, Amygdala: 36.5%, Cerebral cortex: 7.7% | Head of hippocampus (HiH) - Tail of Hippocampus (HiT) - Subicular cortex - Sub | Amygdala excitatory | VGLUT1 VGLUT2 |
| 302 | 0.001370238 | 0.267613784 | Pons: 92.6%, Medulla: 6.6%, Cerebral cortex: 0.7% | Pons (Pn) - efferent nuclei of cranial nerves in pons - PnEN | Cerebellar inhibitory | GLY |
| 157 | 0.001376519 | 0.049898309 | Amygdala: 78.4%, Hippocampus: 19.2%, Cerebral cortex: 2.3% | Amygdaloid complex (AMY) - corticomedial nuclear group - CMN | Amygdala excitatory | VGLUT1 VGLUT2 |
| 399 | 0.00141104 | 0.39471744 | Medulla: 86.6%, Pons: 12.8%, Midbrain: 0.2% | Myelencephalon (medulla oblongata) (Mo) - medullary reticular formation (MoRF) & efferent nuclei of cranial nerves in the medulla oblongata (MoEN) - MoRF-MoEN | Splatter | VGLUT2 |
| 234 | 0.001481866 | 0.12222986 | Amygdala: 73.7%, Thalamus: 9.5%, Hypothalamus: 5.8% | Amygdaloid complex (AMY) - Basolateral nuclear group (BLN) - lateral nucleus - La | Eccentric medium spiny neuron | GABA |
| 116 | 0.00148759 | 0.324446238 | Cerebral cortex: 100.0%, Amygdala: 0.0%, Hippocampus: 0.0% | Cerebral cortex (Cx) - Precentral gyrus (PrCG) - Primary motor cortex - M1C | Hippocampal CA1-3 | VGLUT1 |
| 451 | 0.001551358 | 0.078691091 | Thalamus: 99.9%, Basal forebrain: 0.0%, Hypothalamus: 0.0% | Thalamus (THM) - medial nuclear complex of thalamus (MNC) - mediodorsal nucleus of thalamus - MD | Thalamic excitatory | VGLUT2 |
| 347 | 0.001613268 | 0.083237998 | Medulla: 60.2%, Spinal cord: 39.4%, Pons: 0.3% | Spinal cord - SpC | Splatter | VGLUT2 |
| 407 | 0.001629673 | 0.007839639 | Amygdala: 50.5%, Cerebral cortex: 46.9%, Hypothalamus: 2.4% | Paleocortex (PalCx) - Piriform cortex - Pir | Amygdala excitatory | VGLUT1 VGLUT2 |
| 417 | 0.00163267 | 0.025879226 | Hypothalamus: 65.4%, Thalamus: 17.9%, Amygdala: 8.1% | Hypothalamus (HTH) - supraoptic region of HTH - HTHso (anterior hypothalamic nucleus, AHN) - tuberal region of HTH - HTHtub (ventromedial and dorsomedial hypothalamic nucleic nuclei, VMH, DMH) | Splatter | GABA |
| 456 | 0.001633583 | 0.026506613 | Thalamus: 97.9%, Midbrain: 2.1%, Cerebral cortex: 0.0% | Thalamus (THM) - lateral nuclear complex of thalamus (LNC) - Pulvinar of thalamus - Pul | Thalamic excitatory | VGLUT2 |
| 34 | 0.001655158 | 0.369125016 | Pons: 23.8%, Midbrain: 22.9%, Thalamus: 19.2% | Pons (Pn) - Pontine reticular formation - PnRF | Oligodendrocyte precursor |  |
| 410 | 0.001659835 | 0.716042054 | Basal forebrain: 66.5%, Amygdala: 14.2%, Thalamus: 9.9% | Basal nuclei (BN) - Body of the Caudate - CaB | Splatter | GABA |
| 103 | 0.001662556 | 0.002418689 | Amygdala: 66.6%, Cerebral cortex: 28.9%, Hippocampus: 4.2% | Amygdaloid complex (AMY) - Basolateral nuclear group (BLN) - lateral nucleus - La | Deep-layer corticothalamic and 6b | VGLUT1 |
| 105 | 0.001756598 | 0.000373916 | Cerebral cortex: 47.1%, Hippocampus: 34.6%, Amygdala: 18.1% | Cerebral cortex (Cx) - Anterior parahippocampal gyrus, posterior part (APH) - Medial entorhinal cortex - MEC | Deep-layer corticothalamic and 6b | VGLUT1 |
| 411 | 0.00175819 | 0.125000342 | Hypothalamus: 82.0%, Thalamus: 11.6%, Amygdala: 5.2% | Hypothalamus (HTH) - preoptic region of HTH - HTHpo (medial preoptic nucleus, MPN) - supraoptic region of HTH - HTHso (paraventricular nucleus, PV) | Splatter | VGLUT2 |
| 430 | 0.001884685 | 0.02139776 | Basal forebrain: 55.9%, Amygdala: 24.1%, Thalamus: 12.9% | Basal forebrain (BF) - septal nuclei - SEP | Splatter | GABA |
| 330 | 0.001898981 | 0.010592763 | Hypothalamus: 99.7%, Midbrain: 0.3%, Thalamus: 0.1% | Hypothalamus (HTH) - mammillary region of HTH (HTHma) - tuberal region of HTH - HTHtub | Mammillary body | HDC VGLUT2 |
| 286 | 0.001931761 | 0.106424368 | Cerebral cortex: 62.9%, Hippocampus: 22.3%, Amygdala: 9.5% | Cerebral cortex (Cx) - Precentral gyrus (PrCG) - Primary motor cortex - M1C | CGE interneuron | GABA |
| 110 | 0.00194218 | 0.027949331 | Cerebral cortex: 94.0%, Hippocampus: 3.3%, Basal forebrain: 1.8% | Cerebral cortex (Cx) - Precentral gyrus (PrCG) - Primary motor cortex - M1C | Deep-layer corticothalamic and 6b | VGLUT1 |
| 301 | 0.001993284 | 0.544315581 | Cerebellum: 61.4%, Pons: 32.9%, Midbrain: 3.2% | Cerebellum (CB) - Cerebellar deep nuclei - CbDN | Cerebellar inhibitory | GABA |
| 339 | 0.002002987 | 0.218313621 | Medulla: 99.1%, Basal forebrain: 0.6%, Pons: 0.3% | Myelencephalon (medulla oblongata) (Mo) - afferent nuclei of cranial nerves in medulla oblongata - MoAN | Splatter | VGLUT2 |
| 364 | 0.002027709 | 0.004996252 | Thalamus: 64.4%, Basal forebrain: 20.9%, Hypothalamus: 7.4% | Thalamus (THM) - Anterior nuclear complex - ANC | Splatter | VGLUT2 |
| 90 | 0.002059142 | 0.008639101 | Cerebral cortex: 98.1%, Basal forebrain: 1.6%, Amygdala: 0.1% | Cerebral cortex (Cx) - Precentral gyrus (PrCG) - Primary motor cortex - M1C | Deep-layer near-projecting | VGLUT1 VGLUT2 |
| 326 | 0.002130056 | 0.478922341 | Hypothalamus: 100.0%, Cerebral cortex: 0.0%, Amygdala: 0.0% | Hypothalamus (HTH) - mammillary region of HTH (HTHma) - mammillary nucleus - MN | Mammillary body | VGLUT2 |
| 459 | 0.002179358 | 0.082350241 | Thalamus: 93.6%, Midbrain: 6.4%, Amygdala: 0.0% | Epithalamus - ETH | Thalamic excitatory | VGLUT2 |
| 323 | 0.002252056 | 0.94672102 | Pons: 83.9%, Medulla: 15.5%, Cerebral cortex: 0.4% | Pons (Pn) - efferent nuclei of cranial nerves in pons - PnEN | Lower rhombic lip | VGLUT1 VGLUT2 |
| 38 | 0.002262631 | 0.241049436 | Cerebral cortex: 35.6%, Hippocampus: 19.5%, Amygdala: 8.2% | Perirhinal cortex (area 35) (A35) - rostral subdivision of area 35 - A35r | Committed oligodendrocyte precursor |  |
| 449 | 0.002263741 | 0.273751268 | Thalamus: 99.7%, Amygdala: 0.3%, Basal forebrain: 0.0% | Thalamus (THM) - Lateral nuclear complex of thalamus (LNC) - ventral group of lateral nucleus - VLN | Thalamic excitatory | VGLUT2 |
| 87 | 0.002309801 | 0.015225327 | Cerebral cortex: 97.0%, Basal forebrain: 1.8%, Hippocampus: 1.1% | Cerebral cortex (Cx) - Precentral gyrus (PrCG) - Primary motor cortex - M1C | Deep-layer near-projecting | VGLUT1 VGLUT2 |
| 394 | 0.002336025 | 0.106674994 | Thalamus: 76.5%, Hypothalamus: 10.6%, Midbrain: 6.0% | Thalamus (THM) - Anterior nuclear complex - ANC | Splatter | GABA |
| 169 | 0.002357194 | 0.089028043 | Cerebral cortex: 92.4%, Hippocampus: 7.6%, Amygdala: 0.0% | Cerebral cortex (Cx) - Posterior parahippocampal gyrus (PPH) - TH-TL | Miscellaneous | VGLUT1 VGLUT2 |
| 224 | 0.00236302 | 0.043279303 | Basal forebrain: 74.1%, Amygdala: 12.2%, Thalamus: 9.6% | Basal nuclei (BN) - Body of the Caudate - CaB | Eccentric medium spiny neuron | GABA |
| 355 | 0.002373107 | 0.330881718 | Midbrain: 86.4%, Thalamus: 9.6%, Pons: 2.2% | Midbrain (M) - Pretectal region - PTR | Splatter | GABA |
| 232 | 0.00238609 | 0.048041135 | Amygdala: 60.2%, Basal forebrain: 16.4%, Cerebral cortex: 13.7% | Amygdaloid complex (AMY) - corticomedial nuclear group - CMN | Eccentric medium spiny neuron | GABA |
| 431 | 0.002401799 | 0.140820439 | Amygdala: 87.9%, Hypothalamus: 6.5%, Thalamus: 3.4% | Amygdaloid complex (AMY) - Central nuclear group - CEN | Medium spiny neuron | GABA |
| 424 | 0.002438205 | 0.04581501 | Amygdala: 50.6%, Thalamus: 19.8%, Hypothalamus: 15.4% | Thalamus (THM) - Anterior nuclear complex - ANC | Splatter | GABA |
| 329 | 0.002487296 | 0.03715522 | Hypothalamus: 100.0%, Cerebral cortex: 0.0%, Amygdala: 0.0% | Hypothalamus (HTH) - mammillary region of HTH (HTHma) - mammillary nucleus - MN | Mammillary body | HDC VGLUT2 |
| 396 | 0.002741551 | 0.457851419 | Midbrain: 92.6%, Hypothalamus: 6.5%, Thalamus: 0.7% | Midbrain (M) - Substantia Nigra, Red Nucleus, and nearby nuclei - SN-RN | Splatter | DA VGLUT2 |
| 325 | 0.002780287 | 0.658310692 | Hypothalamus: 100.0%, Cerebral cortex: 0.0%, Amygdala: 0.0% | Hypothalamus (HTH) - mammillary region of HTH (HTHma) - mammillary nucleus - MN | Mammillary body | VGLUT2 |
| 164 | 0.002796602 | 0.052206931 | Amygdala: 93.5%, Hippocampus: 5.9%, Hypothalamus: 0.4% | Amygdaloid complex (AMY) - corticomedial nuclear group - CMN | Hippocampal CA1-3 | VGLUT1 VGLUT2 |
| 175 | 0.002811013 | 0.025204248 | Cerebral cortex: 84.3%, Amygdala: 10.2%, Hippocampus: 5.2% | Cerebral cortex (Cx) - Anterior parahippocampal gyrus (AG) - Lateral entorhinal cortex - LEC | Amygdala excitatory | VGLUT1 VGLUT2 |
| 153 | 0.003079476 | 0.012366805 | Basal forebrain: 76.2%, Amygdala: 19.9%, Cerebral cortex: 3.8% | Claustrum - Cla | Deep-layer intratelencephalic | VGLUT1 VGLUT2 VGLUT3 |
| 230 | 0.00309395 | 0.188724892 | Amygdala: 56.1%, Basal forebrain: 28.8%, Cerebral cortex: 11.2% | Amygdaloid complex (AMY) - Central nuclear group - CEN | Eccentric medium spiny neuron | GABA |
| 64 | 0.003120593 | 0.554618482 | Medulla: 38.6%, Spinal cord: 20.0%, Pons: 15.4% | Spinal cord - SpC | Astrocyte |  |
| 59 | 0.003222102 | 0.816597775 | Cerebral cortex: 57.1%, Hippocampus: 37.8%, Amygdala: 3.0% | Tail of Hippocampus (HiT) - Caudal Hippocampus - CA4-DGC | Astrocyte |  |
| 402 | 0.003258185 | 0.046493437 | Hippocampus: 76.2%, Amygdala: 19.4%, Cerebral cortex: 4.4% | Head of hippocampus (HiH) - Uncal CA1-CA3 | Miscellaneous | VGLUT2 |
| 180 | 0.003315928 | 0.088302624 | Hippocampus: 97.2%, Cerebral cortex: 2.8%, Amygdala: 0.0% | Tail of Hippocampus (HiT) - Caudal Hippocampus - CA1-CA3 | Hippocampal CA1-3 | VGLUT1 |
| 305 | 0.003325217 | 0.549920137 | Cerebellum: 95.5%, Pons: 2.4%, Medulla: 1.8% | Cerebellum (CB) - Cerebellar Vermis - CBV | Cerebellar inhibitory | GABA |
| 95 | 0.003482354 | 0.008464369 | Cerebral cortex: 94.1%, Hippocampus: 2.6%, Amygdala: 2.0% | Cerebral cortex (Cx) - Middle Temporal Gyrus - MTG | Deep-layer near-projecting | VGLUT1 |
| 337 | 0.003706033 | 0.891866819 | Midbrain: 91.5%, Thalamus: 8.1%, Medulla: 0.3% | Midbrain (M) - Substantia Nigra, Red Nucleus, and nearby nuclei - SN-RN | Splatter | VGLUT2 VGLUT3 |
| 213 | 0.003923741 | 0.022826321 | Basal forebrain: 90.4%, Cerebral cortex: 3.6%, Amygdala: 3.6% | Basal nuclei (BN) - Body of the Caudate - CaB | Medium spiny neuron | GABA |
| 264 | 0.003968952 | 0.271185813 | Cerebral cortex: 98.5%, Basal forebrain: 0.6%, Hippocampus: 0.4% | Cerebral cortex (Cx) - Precentral gyrus (PrCG) - Primary motor cortex - M1C | MGE interneuron | GABA |
| 309 | 0.004376507 | 0.038661652 | Cerebellum: 84.5%, Pons: 10.2%, Medulla: 3.6% | Cerebellum (CB) - Cerebellar Vermis - CBV | Upper rhombic lip | VGLUT1 |
| 452 | 0.004518997 | 0.033785026 | Thalamus: 99.9%, Midbrain: 0.1%, Hypothalamus: 0.0% | Thalamus (THM) - medial nuclear complex of thalamus (MNC) - mediodorsal nucleus of thalamus - MD | Thalamic excitatory | VGLUT2 |
| 344 | 0.00473351 | 0.10831531 | Medulla: 57.2%, Spinal cord: 34.1%, Pons: 5.9% | Myelencephalon (medulla oblongata) (Mo) - afferent nuclei of cranial nerves in medulla oblongata - MoAN | Splatter | VGLUT2 |
| 385 | 0.004863376 | 0.148904147 | Hypothalamus: 43.2%, Midbrain: 39.3%, Pons: 9.4% | Midbrain (M) - Substantia Nigra - SN | Splatter | VGLUT2 |
| 204 | 0.004933833 | 0.24420975 | Hippocampus: 98.3%, Midbrain: 1.4%, Cerebral cortex: 0.2% | Head of hippocampus (HiH) - Uncal DG-CA4 | Hippocampal dentate gyrus | VGLUT1 |
| 382 | 0.005070161 | 0.982556995 | Pons: 83.9%, Midbrain: 15.4%, Medulla: 0.5% | Pons (Pn) - other nuclei in pontine tegmentum (XPnTg) - dorsal tegmental nucleus - DTg | Splatter | GABA |
| 173 | 0.005076636 | 0.063547479 | Cerebral cortex: 90.6%, Hippocampus: 8.8%, Amygdala: 0.6% | Cerebral cortex (Cx) - Anterior parahippocampal gyrus, posterior part (APH) - Medial entorhinal cortex - MEC | Amygdala excitatory | VGLUT1 VGLUT2 |
| 333 | 0.005119479 | 0.093333139 | Hypothalamus: 99.9%, Thalamus: 0.1%, Cerebral cortex: 0.0% | Hypothalamus (HTH) - mammillary region of HTH (HTHma) - mammillary nucleus - MN | Mammillary body | VGLUT2 |
| 334 | 0.005178613 | 0.164208408 | Hypothalamus: 97.7%, Cerebral cortex: 1.0%, Thalamus: 0.8% | Hypothalamus (HTH) - mammillary region of HTH (HTHma) - tuberal region of HTH - HTHtub | Mammillary body | HDC VGLUT2 |
| 227 | 0.005248183 | 0.119433238 | Basal forebrain: 33.6%, Amygdala: 33.3%, Hypothalamus: 19.3% | Basal forebrain (BF) - substantia innominata and nearby nuclei - SI | Eccentric medium spiny neuron | GABA |
| 176 | 0.005249697 | 0.021330342 | Cerebral cortex: 91.3%, Hippocampus: 7.3%, Amygdala: 1.2% | Cerebral cortex (Cx) - Anterior parahippocampal gyrus, posterior part (APH) - Medial entorhinal cortex - MEC | Amygdala excitatory | VGLUT1 VGLUT2 |
| 314 | 0.005350061 | 0.116909007 | Hippocampus: 41.6%, Cerebral cortex: 36.5%, Amygdala: 7.6% | Cerebral cortex (Cx) - Anterior parahippocampal gyrus, posterior part (APH) - Medial entorhinal cortex - MEC | Splatter | VGLUT2 |
| 353 | 0.005398601 | 0.268936159 | Midbrain: 97.3%, Pons: 0.9%, Medulla: 0.8% | Midbrain (M) - Superior colliculus and nearby nuclei - SC | Splatter | GABA |
| 328 | 0.005495539 | 0.092537657 | Hypothalamus: 99.9%, Midbrain: 0.1%, Cerebral cortex: 0.0% | Hypothalamus (HTH) - mammillary region of HTH (HTHma) - mammillary nucleus - MN | Mammillary body | VGLUT2 |
| 125 | 0.005514787 | 0.048328289 | Cerebral cortex: 98.9%, Basal forebrain: 0.5%, Hippocampus: 0.2% | Cerebral cortex (Cx) - Middle Temporal Gyrus - MTG | Upper-layer intratelencephalic | VGLUT1 VGLUT2 |
| 233 | 0.005668182 | 0.174042722 | Basal forebrain: 32.8%, Amygdala: 30.3%, Hypothalamus: 20.9% | Basal forebrain (BF) - substantia innominata and nearby nuclei - SI | Eccentric medium spiny neuron | GABA |
| 130 | 0.005707074 | 0.065951623 | Cerebral cortex: 99.1%, Hippocampus: 0.4%, Basal forebrain: 0.4% | Cerebral cortex (Cx) - Middle Temporal Gyrus - MTG | Upper-layer intratelencephalic | VGLUT1 VGLUT2 |
| 442 | 0.005909802 | 0.401174703 | Thalamus: 89.9%, Midbrain: 8.8%, Pons: 0.7% | Thalamus (THM) - lateral nuclear complex of thalamus (LNC) - Pulvinar of thalamus - Pul | Midbrain-derived inhibitory | GABA |
| 177 | 0.006064768 | 0.136560138 | Hippocampus: 98.8%, Cerebral cortex: 1.0%, Thalamus: 0.2% | Head of hippocampus (HiH) - Uncal CA1-CA3 | Miscellaneous | VGLUT1 VGLUT2 |
| 221 | 0.006305277 | 0.127236533 | Amygdala: 91.6%, Thalamus: 8.4%, Cerebral cortex: 0.0% | Amygdaloid complex (AMY) - Central nuclear group - CEN | Medium spiny neuron | GABA |
| 36 | 0.006337584 | 0.152707476 | Cerebral cortex: 62.3%, Hippocampus: 21.4%, Basal forebrain: 6.5% | Perirhinal cortex (area 35) (A35) - rostral subdivision of area 35 - A35r | Oligodendrocyte precursor |  |
| 257 | 0.006399333 | 0.606520735 | Cerebral cortex: 68.6%, Hippocampus: 21.8%, Basal forebrain: 5.0% | Cerebral cortex (Cx) - Precentral gyrus (PrCG) - Primary motor cortex - M1C | MGE interneuron | GABA VGLUT3 |
| 58 | 0.006530065 | 0.932171863 | Hippocampus: 66.8%, Basal forebrain: 25.3%, Cerebral cortex: 4.0% | Body of hippocampus (HiB) - Rostral DG-CA4 | Astrocyte |  |
| 320 | 0.006645131 | 0.879155852 | Pons: 97.8%, Midbrain: 1.2%, Medulla: 1.0% | Pons (Pn) - Pontine Nucleus - PN | Lower rhombic lip | VGLUT1 VGLUT2 |
| 126 | 0.007153298 | 0.138461528 | Cerebral cortex: 98.3%, Basal forebrain: 0.9%, Amygdala: 0.4% | Cerebral cortex (Cx) - Middle Temporal Gyrus - MTG | Upper-layer intratelencephalic | VGLUT1 VGLUT2 |
| 194 | 0.007256269 | 0.386024587 | Hippocampus: 98.3%, Amygdala: 0.9%, Cerebral cortex: 0.5% | Tail of Hippocampus (HiT) - Caudal Hippocampus - CA4-DGC | Hippocampal CA4 | VGLUT1 |
| 457 | 0.007283643 | 0.211091838 | Thalamus: 99.8%, Hippocampus: 0.2%, Cerebral cortex: 0.0% | Thalamus (THM) - posterior nuclear complex of thalamus (PoN) - lateral geniculate nucleus (LG) | Thalamic excitatory | VGLUT2 |
| 215 | 0.007536256 | 0.082748258 | Basal forebrain: 61.9%, Amygdala: 15.7%, Cerebral cortex: 13.3% | Basal forebrain (BF) - substantia innominata and nearby nuclei - SI | Medium spiny neuron | GABA |
| 187 | 0.00764517 | 0.041394543 | Hippocampus: 98.7%, Cerebral cortex: 1.1%, Amygdala: 0.2% | Body of hippocampus (HiB) - Rostral CA1-CA3 | Hippocampal CA1-3 | VGLUT1 |
| 441 | 0.007868311 | 0.283282225 | Thalamus: 87.1%, Midbrain: 10.2%, Pons: 1.9% | Epithalamus - ETH | Midbrain-derived inhibitory | GABA |
| 190 | 0.008237485 | 0.109012692 | Hippocampus: 73.4%, Amygdala: 24.4%, Hypothalamus: 0.9% | Head of hippocampus (HiH) - CA1 | Hippocampal CA1-3 | VGLUT1 |
| 384 | 0.008321872 | 0.305775533 | Hypothalamus: 68.0%, Midbrain: 24.6%, Thalamus: 6.6% | Hypothalamus (HTH) - mammillary region of HTH (HTHma) | Splatter | VGLUT2 VGLUT3 |
| 371 | 0.008415425 | 0.396778998 | Midbrain: 98.8%, Pons: 0.5%, Thalamus: 0.5% | Midbrain (M) - Superior colliculus and nearby nuclei - SC | Splatter | VGLUT2 |
| 235 | 0.008659473 | 0.154650841 | Amygdala: 75.9%, Cerebral cortex: 14.6%, Thalamus: 5.4% | Amygdaloid complex (AMY) - basolateral nuclear group (BLN) - basomedial nucleus (accessory basal nucleus) - BM | Eccentric medium spiny neuron | GABA |
| 113 | 0.008974763 | 0.019567967 | Cerebral cortex: 65.2%, Hippocampus: 33.8%, Amygdala: 0.6% | Cerebral cortex (Cx) - Anterior parahippocampal gyrus, posterior part (APH) - Medial entorhinal cortex - MEC | Deep-layer corticothalamic and 6b | VGLUT1 |
| 415 | 0.009267569 | 0.918098659 | Hypothalamus: 90.6%, Midbrain: 8.5%, Thalamus: 0.8% | Hypothalamus (HTH) - mammillary region of HTH (HTHma) - tuberal region of HTH - HTHtub | Splatter | VGLUT2 |
| 207 | 0.009267774 | 0.095229855 | Hippocampus: 97.1%, Thalamus: 2.3%, Cerebral cortex: 0.6% | Tail of Hippocampus (HiT) - Caudal Hippocampus - CA4-DGC | Medium spiny neuron |  |
| 365 | 0.009507364 | 0.152662489 | Midbrain: 99.6%, Thalamus: 0.2%, Medulla: 0.1% | Midbrain (M) - Inferior colliculus and nearby nuclei - IC | Splatter | VGLUT2 |
| 193 | 0.009557655 | 0.256205285 | Hippocampus: 99.6%, Cerebral cortex: 0.4%, Amygdala: 0.0% | Tail of Hippocampus (HiT) - Caudal Hippocampus - CA4-DGC | Hippocampal CA4 | VGLUT1 |
| 368 | 0.009820077 | 0.027284269 | Midbrain: 96.9%, Thalamus: 2.6%, Pons: 0.3% | Midbrain (M) - Superior colliculus and nearby nuclei - SC | Splatter | VGLUT2 |
| 401 | 0.010130043 | 0.127878482 | Basal forebrain: 54.0%, Amygdala: 22.9%, Hypothalamus: 14.7% | Basal forebrain (BF) - substantia innominata and nearby nuclei - SI | Splatter | CHOL VGLUT3 |
| 427 | 0.010159944 | 0.002665804 | Amygdala: 81.9%, Cerebral cortex: 10.0%, Thalamus: 3.2% | Amygdaloid complex (AMY) - Central nuclear group - CEN | Eccentric medium spiny neuron | GABA |
| 373 | 0.010182981 | 0.018227252 | Midbrain: 53.2%, Thalamus: 44.2%, Cerebral cortex: 2.5% | Midbrain (M) - Pretectal region - PTR | Thalamic excitatory | VGLUT2 |
| 55 | 0.010617163 | 0.488222129 | Cerebral cortex: 38.8%, Hippocampus: 33.1%, Amygdala: 22.1% | Perirhinal cortex (area 35) (A35) - rostral subdivision of area 35 - A35r | Astrocyte |  |
| 35 | 0.010635484 | 0.320008797 | Hippocampus: 33.6%, Cerebral cortex: 30.9%, Amygdala: 16.5% | Head of hippocampus (HiH) - CA1 | Oligodendrocyte precursor |  |
| 245 | 0.010766341 | 0.017430571 | Cerebral cortex: 77.4%, Amygdala: 13.7%, Hippocampus: 6.5% | Cerebral cortex (Cx) - Anterior parahippocampal gyrus, posterior part (APH) - Medial entorhinal cortex - MEC | MGE interneuron | GABA |
| 370 | 0.010891188 | 0.26608162 | Midbrain: 96.5%, Thalamus: 2.3%, Pons: 0.6% | Midbrain (M) - Pretectal region - PTR | Splatter | VGLUT2 |
| 134 | 0.011173718 | 0.224852171 | Cerebral cortex: 100.0%, Basal forebrain: 0.0%, Hypothalamus: 0.0% | Cerebral cortex (Cx) - Lingual gyrus (LiG) - Primary Visual Cortex - V1C | Upper-layer intratelencephalic | VGLUT1 VGLUT2 |
| 33 | 0.011254224 | 0.949768395 | Hypothalamus: 54.7%, Thalamus: 16.6%, Amygdala: 8.5% | Hypothalamus (HTH) - mammillary region of HTH (HTHma) - mammillary nucleus - MN | Oligodendrocyte precursor |  |
| 3 | 0.011284325 | 0.049695553 | Thalamus: 17.6%, Cerebral cortex: 15.5%, Basal forebrain: 14.8% | Basal nuclei (BN) - Globus pallidus (GP) - External segment of globus pallidus - GPe | Miscellaneous |  |
| 413 | 0.011730693 | 0.141301327 | Hypothalamus: 97.4%, Midbrain: 2.2%, Thalamus: 0.3% | Hypothalamus (HTH) - mammillary region of HTH (HTHma) - mammillary nucleus - MN | Splatter | GABA HDC VGLUT2 |
| 228 | 0.012444805 | 0.294737372 | Amygdala: 52.6%, Basal forebrain: 20.6%, Hypothalamus: 20.3% | Amygdaloid complex (AMY) - corticomedial nuclear group - CMN | Eccentric medium spiny neuron | GABA |
| 383 | 0.01249091 | 0.056948516 | Amygdala: 32.1%, Hypothalamus: 30.4%, Basal forebrain: 21.9% | Extended amygdala (EXA) - Bed nucleus of stria terminalis and nearby - BNST | Splatter | VGLUT2 |
| 123 | 0.012666473 | 0.07257631 | Cerebral cortex: 93.9%, Basal forebrain: 2.6%, Hippocampus: 1.7% | Cerebral cortex (Cx) - Middle Temporal Gyrus - MTG | Upper-layer intratelencephalic | VGLUT1 VGLUT2 |
| 98 | 0.01299271 | 0.007031447 | Cerebral cortex: 54.4%, Hippocampus: 42.5%, Amygdala: 2.1% | Cerebral cortex (Cx) - Anterior parahippocampal gyrus, posterior part (APH) - Medial entorhinal cortex - MEC | Deep-layer corticothalamic and 6b | VGLUT1 |
| 443 | 0.013355919 | 0.556024105 | Thalamus: 99.1%, Midbrain: 0.8%, Pons: 0.1% | Thalamus (THM) - lateral nuclear complex of thalamus (LNC) - Pulvinar of thalamus - Pul | Midbrain-derived inhibitory | GABA |
| 304 | 0.013451163 | 0.678860152 | Cerebellum: 96.8%, Pons: 1.5%, Medulla: 1.0% | Cerebellum (CB) - Cerebellar Vermis - CBV | Cerebellar inhibitory | GABA |
| 122 | 0.013552498 | 0.096177564 | Cerebral cortex: 91.7%, Amygdala: 3.9%, Basal forebrain: 2.7% | Cerebral cortex (Cx) - Temporal pole (TP) - Temporopolar area - A38 | Upper-layer intratelencephalic | VGLUT1 VGLUT2 |
| 460 | 0.013784178 | 0.393056019 | Thalamus: 89.9%, Amygdala: 9.2%, Midbrain: 0.6% | Thalamus (THM) - medial nuclear complex of thalamus (MNC) - mediodorsal nucleus of thalamus - MD | Thalamic excitatory | VGLUT2 |
| 387 | 0.013832372 | 0.138329363 | Medulla: 98.2%, Pons: 1.2%, Midbrain: 0.6% | Myelencephalon (medulla oblongata) (Mo) - medullary reticular formation (MoRF) & efferent nuclei of cranial nerves in the medulla oblongata (MoEN) - MoRF-MoEN | Splatter | VGLUT2 |
| 461 | 0.0138603 | 0.278045475 | Thalamus: 98.9%, Midbrain: 0.9%, Cerebral cortex: 0.1% | Thalamus (THM) - intralaminar nuclear complex (ILN) - posterior group of intralaminar nuclei (PILN) - centromedian and parafasicular nuclei - CM and Pf | Thalamic excitatory | VGLUT2 |
| 367 | 0.014193466 | 0.666272343 | Midbrain: 57.4%, Pons: 19.9%, Thalamus: 15.2% | Midbrain (M) - Periaqueductal gray and nearby nuclei - PAG | Splatter | VGLUT2 |
| 212 | 0.014532877 | 0.083363244 | Basal forebrain: 85.7%, Amygdala: 6.3%, Cerebral cortex: 4.2% | Basal nuclei (BN) - Body of the Caudate - CaB | Medium spiny neuron | GABA |
| 420 | 0.014585116 | 0.105322329 | Amygdala: 78.9%, Cerebral cortex: 14.0%, Thalamus: 5.1% | Amygdaloid complex (AMY) - corticomedial nuclear group - CMN | Amygdala excitatory | VGLUT1 VGLUT2 |
| 444 | 0.015276094 | 0.434481227 | Thalamus: 98.3%, Midbrain: 1.6%, Pons: 0.1% | Thalamus (THM) - lateral nuclear complex of thalamus (LNC) - Pulvinar of thalamus - Pul | Midbrain-derived inhibitory | GABA |
| 112 | 0.015857912 | 0.007104079 | Hippocampus: 92.5%, Amygdala: 6.3%, Cerebral cortex: 1.2% | Head of hippocampus (HiH) - CA1 | Deep-layer corticothalamic and 6b | VGLUT1 VGLUT2 |
| 421 | 0.0161651 | 0.103105353 | Thalamus: 46.3%, Hypothalamus: 26.8%, Amygdala: 24.7% | Thalamus (THM) - Anterior nuclear complex - ANC | Splatter | VGLUT2 |
| 166 | 0.016179502 | 0.019149527 | Cerebral cortex: 92.1%, Hippocampus: 5.0%, Amygdala: 2.8% | Cerebral cortex (Cx) - Anterior parahippocampal gyrus, posterior part (APH) - Medial entorhinal cortex - MEC | Miscellaneous | VGLUT1 VGLUT2 |
| 63 | 0.016264799 | 0.436100563 | Basal forebrain: 40.2%, Cerebellum: 19.3%, Midbrain: 16.2% | Basal nuclei (BN) - Globus pallidus (GP) - Internal segment of globus pallidus - GPi | Astrocyte |  |
| 236 | 0.016318142 | 0.800428759 | Cerebral cortex: 31.8%, Amygdala: 28.5%, Basal forebrain: 25.2% | Basal forebrain (BF) - substantia innominata and nearby nuclei - SI | Splatter | GABA |
| 247 | 0.016533341 | 0.468004713 | Hippocampus: 72.3%, Cerebral cortex: 16.5%, Amygdala: 8.9% | Tail of Hippocampus (HiT) - Caudal Hippocampus - CA4-DGC | MGE interneuron | GABA |
| 346 | 0.016668436 | 0.250368373 | Medulla: 52.3%, Spinal cord: 35.4%, Midbrain: 6.8% | Myelencephalon (medulla oblongata) (Mo) - afferent nuclei of cranial nerves in medulla oblongata - MoAN | Splatter | VGLUT2 |
| 432 | 0.016778319 | 0.256724943 | Hypothalamus: 99.7%, Midbrain: 0.2%, Cerebral cortex: 0.0% | Hypothalamus (HTH) - mammillary region of HTH (HTHma) - mammillary nucleus - MN | Splatter | HDC VGLUT2 |
| 331 | 0.016817759 | 0.265020635 | Hypothalamus: 100.0%, Cerebral cortex: 0.0%, Amygdala: 0.0% | Hypothalamus (HTH) - mammillary region of HTH (HTHma) - mammillary nucleus - MN | Mammillary body | VGLUT2 |
| 345 | 0.016963616 | 0.275386018 | Medulla: 61.0%, Spinal cord: 21.9%, Pons: 14.9% | Myelencephalon (medulla oblongata) (Mo) - afferent nuclei of cranial nerves in medulla oblongata - MoAN | Splatter | VGLUT2 |
| 426 | 0.017093641 | 0.03358491 | Thalamus: 35.4%, Hypothalamus: 29.5%, Amygdala: 29.1% | Thalamus (THM) - Anterior nuclear complex - ANC | Splatter | GABA |
| 377 | 0.017143424 | 0.412105847 | Hypothalamus: 51.0%, Midbrain: 37.9%, Thalamus: 6.6% | Midbrain (M) - Substantia Nigra, Red Nucleus, and nearby nuclei - SN-RN | Splatter | VGLUT2 |
| 184 | 0.017190714 | 0.121018719 | Hippocampus: 99.9%, Amygdala: 0.0%, Cerebral cortex: 0.0% | Head of hippocampus (HiH) - CA2-3 | Hippocampal CA1-3 | VGLUT1 |
| 324 | 0.017281718 | 0.414891246 | Hypothalamus: 99.7%, Thalamus: 0.2%, Midbrain: 0.2% | Hypothalamus (HTH) - mammillary region of HTH (HTHma) - mammillary nucleus - MN | Mammillary body | VGLUT2 |
| 312 | 0.017380646 | 0.033773064 | Cerebellum: 72.7%, Medulla: 10.3%, Pons: 8.2% | Cerebellum (CB) - Cerebellar Vermis - CBV | Upper rhombic lip | VGLUT1 |
| 192 | 0.017665554 | 0.593131083 | Hippocampus: 98.1%, Cerebral cortex: 1.6%, Midbrain: 0.2% | Tail of Hippocampus (HiT) - Caudal Hippocampus - CA4-DGC | Hippocampal CA4 | VGLUT1 |
| 124 | 0.018395743 | 0.067712896 | Cerebral cortex: 99.2%, Hippocampus: 0.4%, Basal forebrain: 0.3% | Cerebral cortex (Cx) - Supraparietal lobule (SPL) - Posterosuperior (dorsal) parietal cortex - A5-A7 | Upper-layer intratelencephalic | VGLUT1 |
| 158 | 0.020313134 | 0.055500593 | Amygdala: 65.8%, Cerebral cortex: 28.7%, Hippocampus: 5.5% | Amygdaloid complex (AMY) - basolateral nuclear group (BLN) - basolateral nucleus (basal nucleus) - BL | Amygdala excitatory | VGLUT1 VGLUT2 |
| 2 | 0.020422873 | 0.334196375 | Midbrain: 15.0%, Basal forebrain: 14.0%, Pons: 13.2% | Spinal cord - SpC | Miscellaneous |  |
| 161 | 0.020525314 | 0.120601118 | Amygdala: 90.3%, Cerebral cortex: 8.9%, Hippocampus: 0.8% | Amygdaloid complex (AMY) - basolateral nuclear group (BLN) - basolateral nucleus (basal nucleus) - BL | Amygdala excitatory | VGLUT1 VGLUT2 |
| 195 | 0.021107791 | 0.448060315 | Hippocampus: 99.8%, Cerebral cortex: 0.2%, Amygdala: 0.0% | Tail of Hippocampus (HiT) - Caudal Hippocampus - CA4-DGC | Hippocampal CA4 | VGLUT1 |
| 336 | 0.021197159 | 0.133182023 | Medulla: 78.3%, Pons: 15.9%, Midbrain: 4.8% | Myelencephalon (medulla oblongata) (Mo) - precerebellar nuclei (PrCbN) - inferior olive - IO | Splatter | VGLUT2 |
| 455 | 0.021779131 | 0.405925095 | Thalamus: 99.5%, Midbrain: 0.5%, Cerebral cortex: 0.0% | Thalamus (THM) - lateral nuclear complex of thalamus (LNC) - Pulvinar of thalamus - Pul | Thalamic excitatory | VGLUT2 |
| 208 | 0.022550331 | 0.011344748 | Basal forebrain: 92.9%, Thalamus: 4.1%, Amygdala: 1.9% | Basal nuclei (BN) - Putamen - Pu | Medium spiny neuron | GABA |
| 186 | 0.022588904 | 0.251786423 | Hippocampus: 100.0%, Cerebral cortex: 0.0%, Amygdala: 0.0% | Tail of Hippocampus (HiT) - Caudal Hippocampus - CA1-CA3 | Hippocampal CA1-3 | VGLUT1 |
| 439 | 0.023801436 | 0.163776146 | Midbrain: 44.9%, Thalamus: 38.0%, Hypothalamus: 8.8% | Midbrain (M) - Substantia Nigra, Red Nucleus, and nearby nuclei - SN-RN | Midbrain-derived inhibitory | GABA |
| 437 | 0.023889784 | 0.305093436 | Thalamus: 82.2%, Midbrain: 17.5%, Hypothalamus: 0.1% | Thalamus (THM) - posterior nuclear complex of thalamus (PoN) - lateral geniculate nucleus (LG) | Midbrain-derived inhibitory | GABA |
| 378 | 0.023968352 | 0.320618063 | Hypothalamus: 33.0%, Amygdala: 32.4%, Thalamus: 31.4% | Extended amygdala (EXA) - Bed nucleus of stria terminalis and nearby - BNST | Splatter | VGLUT2 |
| 418 | 0.024158935 | 0.085806437 | Hypothalamus: 95.8%, Midbrain: 4.0%, Amygdala: 0.1% | Hypothalamus (HTH) - tuberal region of hypothalamus - HTHtub | Splatter | GABA HDC |
| 205 | 0.024673902 | 0.751503419 | Hippocampus: 98.3%, Thalamus: 1.7%, Cerebral cortex: 0.0% | Body of hippocampus (HiB) - Rostral DG-CA4 | Hippocampal dentate gyrus | VGLUT1 |
| 354 | 0.024776111 | 0.09487542 | Thalamus: 97.2%, Midbrain: 2.2%, Hypothalamus: 0.5% | Thalamus (THM) - posterior nuclear complex of thalamus (PoN) - lateral geniculate nucleus (LG) | Splatter | GABA |
| 39 | 0.025167169 | 0.681874142 | Hippocampus: 29.9%, Cerebral cortex: 29.5%, Pons: 7.8% | Head of hippocampus (HiH) - CA1 | Committed oligodendrocyte precursor |  |
| 398 | 0.025313676 | 0.384931179 | Pons: 80.1%, Medulla: 8.1%, Midbrain: 7.6% | Pons (Pn) - Pontine reticular formation - PnRF | Splatter | SER VGLUT3 |
| 351 | 0.025424969 | 0.113447362 | Medulla: 98.2%, Spinal cord: 1.0%, Pons: 0.7% | Myelencephalon (medulla oblongata) (Mo) - medullary reticular formation (MoRF) & efferent nuclei of cranial nerves in the medulla oblongata (MoEN) - MoRF-MoEN | Splatter | GLY |
| 119 | 0.025442226 | 0.635183677 | Hippocampus: 60.6%, Cerebral cortex: 26.6%, Midbrain: 11.9% | Head of hippocampus (HiH) - Tail of Hippocampus (HiT) - Subicular cortex - Sub | Miscellaneous | VGLUT1 |
| 21 | 0.025825291 | 0.665024922 | Thalamus: 21.8%, Cerebral cortex: 13.7%, Medulla: 12.4% | Hypothalamus (HTH) - mammillary region of HTH (HTHma) - mammillary nucleus - MN | Vascular |  |
| 52 | 0.026416509 | 0.825798667 | Cerebellum: 95.8%, Pons: 1.9%, Medulla: 1.9% | Cerebellum (CB) - Cerebellar Vermis - CBV | Bergmann glia |  |
| 189 | 0.026842297 | 0.246251971 | Hippocampus: 98.4%, Cerebral cortex: 1.2%, Amygdala: 0.3% | Head of hippocampus (HiH) - Uncal CA1-CA3 | Hippocampal CA1-3 | VGLUT1 |
| 56 | 0.027493629 | 0.780822832 | Basal forebrain: 82.4%, Thalamus: 7.0%, Hypothalamus: 5.0% | Basal nuclei (BN) - Putamen - Pu | Astrocyte |  |
| 209 | 0.027804443 | 0.045613365 | Basal forebrain: 89.6%, Amygdala: 5.6%, Cerebral cortex: 3.5% | Basal nuclei (BN) - Body of the Caudate - CaB | Medium spiny neuron | GABA |
| 332 | 0.028543721 | 0.185008961 | Hypothalamus: 96.6%, Midbrain: 2.3%, Cerebral cortex: 1.1% | Hypothalamus (HTH) - mammillary region of HTH (HTHma) - mammillary nucleus - MN | Mammillary body | VGLUT2 |
| 62 | 0.028586339 | 0.045320118 | Hippocampus: 47.7%, Cerebral cortex: 21.1%, Basal forebrain: 19.2% | Head of hippocampus (HiH) - CA1 | Astrocyte |  |
| 246 | 0.029448581 | 0.094390759 | Amygdala: 50.6%, Cerebral cortex: 31.0%, Hippocampus: 15.3% | Amygdaloid complex (AMY) - corticomedial nuclear group - CMN | MGE interneuron | GABA |
| 61 | 0.030155856 | 0.369630739 | Thalamus: 37.1%, Midbrain: 31.3%, Pons: 16.8% | Midbrain (M) - Substantia Nigra, Red Nucleus, and nearby nuclei - SN-RN | Astrocyte |  |
| 453 | 0.030826475 | 0.525805412 | Thalamus: 98.9%, Midbrain: 1.1%, Amygdala: 0.0% | Thalamus (THM) - posterior nuclear complex of thalamus (PoN) - lateral geniculate nucleus (LG) | Thalamic excitatory | VGLUT2 |
| 299 | 0.030888924 | 0.926541114 | Pons: 82.7%, Cerebellum: 11.0%, Medulla: 4.1% | Pons (Pn) - efferent nuclei of cranial nerves in pons - PnEN | Cerebellar inhibitory | GLY |
| 335 | 0.032331307 | 0.353785028 | Thalamus: 91.7%, Midbrain: 8.2%, Pons: 0.1% | Epithalamus - ETH | Splatter | CHOL VGLUT1 VGLUT2 |
| 303 | 0.032683237 | 0.3215936 | Cerebellum: 60.1%, Medulla: 19.0%, Pons: 15.3% | Cerebellum (CB) - Cerebellar Vermis - CBV | Cerebellar inhibitory | GLY |
| 37 | 0.032735562 | 0.896124286 | Cerebral cortex: 33.7%, Thalamus: 12.5%, Midbrain: 11.5% | Perirhinal cortex (area 35) (A35) - rostral subdivision of area 35 - A35r | Oligodendrocyte precursor |  |
| 168 | 0.03548574 | 0.134526924 | Hippocampus: 76.8%, Cerebral cortex: 14.3%, Midbrain: 8.2% | Head of hippocampus (HiH) - Tail of Hippocampus (HiT) - Subicular cortex - Sub | Miscellaneous | VGLUT1 |
| 129 | 0.037005006 | 0.532326453 | Cerebral cortex: 99.9%, Hypothalamus: 0.0%, Basal forebrain: 0.0% | Cerebral cortex (Cx) - Precentral gyrus (PrCG) - Primary motor cortex - M1C | Upper-layer intratelencephalic | VGLUT1 VGLUT2 |
| 12 | 0.039443019 | 0.644741402 | Cerebral cortex: 37.4%, Hippocampus: 24.2%, Thalamus: 11.1% | Body of hippocampus (HiB) - Rostral DG-CA4 | Microglia |  |
| 231 | 0.040847834 | 0.329910111 | Amygdala: 41.5%, Basal forebrain: 26.8%, Cerebral cortex: 15.8% | Basal forebrain (BF) - substantia innominata and nearby nuclei - SI | Eccentric medium spiny neuron | GABA |
| 341 | 0.042736238 | 0.417353696 | Pons: 88.3%, Midbrain: 9.5%, Thalamus: 1.6% | Pons (Pn) - Pontine reticular formation (PnRF) - parabrachial nuclei - PB | Splatter | VGLUT2 |
| 389 | 0.04276322 | 0.293562262 | Pons: 90.9%, Hypothalamus: 4.1%, Amygdala: 3.6% | Pons (Pn) - efferent nuclei of cranial nerves in pons - PnEN | Splatter | VGLUT2 |
| 188 | 0.044790525 | 0.275945292 | Hippocampus: 92.4%, Amygdala: 6.4%, Cerebral cortex: 1.1% | Head of hippocampus (HiH) - CA1 | Hippocampal CA1-3 | VGLUT1 |
| 19 | 0.044927148 | 0.868894659 | Cerebral cortex: 23.3%, Medulla: 16.4%, Thalamus: 14.6% | Perirhinal cortex (area 35) (A35) - rostral subdivision of area 35 - A35r | Vascular |  |
| 386 | 0.045260537 | 0.73662795 | Hypothalamus: 93.4%, Midbrain: 4.5%, Amygdala: 0.8% | Hypothalamus (HTH) - mammillary region of HTH (HTHma) - tuberal region of HTH - HTHtub | Splatter | VGLUT2 |
| 369 | 0.045405442 | 0.665692293 | Midbrain: 84.5%, Thalamus: 13.4%, Pons: 1.5% | Midbrain (M) - Pretectal region - PTR | Splatter | VGLUT2 |
| 211 | 0.047184388 | 0.190982172 | Basal forebrain: 97.9%, Thalamus: 1.9%, Hippocampus: 0.2% | Basal nuclei (BN) - Putamen - Pu | Medium spiny neuron | GABA |
| 440 | 0.049164725 | 0.418674638 | Midbrain: 52.8%, Thalamus: 31.3%, Pons: 10.7% | Midbrain (M) - Superior colliculus and nearby nuclei - SC | Midbrain-derived inhibitory | GABA |
| 57 | 0.050303764 | 0.653861907 | Hypothalamus: 30.0%, Thalamus: 21.5%, Amygdala: 14.3% | Hypothalamus (HTH) - supraoptic region of HTH - HTHso | Astrocyte |  |
| 131 | 0.050820431 | 0.159058187 | Cerebral cortex: 97.8%, Amygdala: 0.7%, Basal forebrain: 0.7% | Cerebral cortex (Cx) - Cingulate gyrus, retrosplenial (CgGrs) - A29-A30 | Upper-layer intratelencephalic | VGLUT1 VGLUT2 |
| 416 | 0.052312818 | 0.339808578 | Hypothalamus: 78.7%, Midbrain: 7.6%, Thalamus: 3.7% | Hypothalamus (HTH) - preoptic region of HTH - HTHpo (medial preoptic nucleus, MPN) - supraoptic region of HTH - HTHso (paraventricular nucleus, PV) | Splatter | GABA VGLUT2 |
| 327 | 0.053116655 | 0.487519505 | Hypothalamus: 100.0%, Cerebral cortex: 0.0%, Amygdala: 0.0% | Hypothalamus (HTH) - mammillary region of HTH (HTHma) - mammillary nucleus - MN | Mammillary body | VGLUT2 |
| 366 | 0.053137799 | 0.65005745 | Pons: 43.0%, Midbrain: 30.2%, Medulla: 17.1% | Pons (Pn) - other nuclei in pontine tegmentum (XPnTg) - dorsal tegmental nucleus - DTg | Splatter | GABA VGLUT2 |
| 54 | 0.053790606 | 0.773280656 | Hippocampus: 96.3%, Cerebral cortex: 1.5%, Basal forebrain: 1.3% | Tail of Hippocampus (HiT) - Caudal Hippocampus - CA4-DGC | Astrocyte |  |
| 379 | 0.056277722 | 0.55998318 | Amygdala: 29.4%, Basal forebrain: 25.2%, Hypothalamus: 22.3% | Extended amygdala (EXA) - Bed nucleus of stria terminalis and nearby - BNST | Splatter | HDC VGLUT2 |
| 182 | 0.057467033 | 0.484744017 | Hippocampus: 100.0%, Cerebral cortex: 0.0%, Amygdala: 0.0% | Tail of Hippocampus (HiT) - Caudal Hippocampus - CA1-CA3 | Hippocampal CA1-3 | VGLUT1 |
| 392 | 0.057482483 | 0.498954333 | Hypothalamus: 92.3%, Thalamus: 3.3%, Midbrain: 2.8% | Hypothalamus (HTH) - preoptic region of HTH - HTHpo (medial preoptic nucleus, MPN) - supraoptic region of HTH - HTHso (paraventricular nucleus, PV) | Splatter | GABA |
| 448 | 0.058772424 | 0.819929459 | Thalamus: 100.0%, Cerebral cortex: 0.0%, Amygdala: 0.0% | Thalamus (THM) - posterior nuclear complex of thalamus (PoN) - lateral geniculate nucleus (LG) | Thalamic excitatory | VGLUT2 |
| 217 | 0.059377207 | 0.056129464 | Basal forebrain: 40.9%, Amygdala: 26.3%, Hypothalamus: 18.3% | Basal forebrain (BF) - substantia innominata and nearby nuclei - SI | Medium spiny neuron | GABA |
| 22 | 0.060090352 | 0.817450567 | Thalamus: 28.3%, Pons: 17.9%, Basal forebrain: 9.8% | Thalamus (THM) - posterior nuclear complex of thalamus (PoN) - lateral geniculate nucleus (LG) | Vascular |  |
| 381 | 0.063168751 | 0.685416373 | Pons: 62.5%, Medulla: 34.7%, Spinal cord: 1.1% | Pons (Pn) - efferent nuclei of cranial nerves in pons - PnEN | Splatter | GLY |
| 300 | 0.063954336 | 0.158841225 | Cerebellum: 71.1%, Pons: 20.4%, Midbrain: 4.5% | Cerebellum (CB) - Cerebellar deep nuclei - CbDN | Cerebellar inhibitory | GLY |
| 375 | 0.064294456 | 0.106983593 | Pons: 77.9%, Midbrain: 19.6%, Medulla: 2.0% | Pons (Pn) - Pontine reticular formation (PnRF) - parabrachial nuclei - PB | Splatter | VGLUT2 |
| 311 | 0.065442665 | 0.05912794 | Cerebellum: 52.4%, Pons: 26.2%, Medulla: 15.1% | Cerebellum (CB) - Cerebellar Vermis - CBV | Upper rhombic lip | VGLUT1 |
| 67 | 0.06583432 | 0.682824329 | Medulla: 83.1%, Pons: 16.0%, Midbrain: 0.4% | Myelencephalon (medulla oblongata) (Mo) - afferent nuclei of cranial nerves in medulla oblongata - MoAN | Ependymal |  |
| 445 | 0.068831863 | 0.73642676 | Thalamus: 83.2%, Midbrain: 14.0%, Pons: 1.9% | Thalamus (THM) - posterior nuclear complex of thalamus (PoN) - lateral geniculate nucleus (LG) | Midbrain-derived inhibitory | GABA |
| 214 | 0.070021423 | 0.102526287 | Basal forebrain: 75.5%, Cerebral cortex: 12.2%, Amygdala: 9.4% | Basal forebrain (BF) - substantia innominata and nearby nuclei - SI | Medium spiny neuron | GABA |
| 132 | 0.071441863 | 0.657820107 | Cerebral cortex: 99.9%, Hypothalamus: 0.0%, Basal forebrain: 0.0% | Cerebral cortex (Cx) - Precentral gyrus (PrCG) - Primary motor cortex - M1C | Upper-layer intratelencephalic | VGLUT1 VGLUT2 |
| 69 | 0.07467156 | 0.444969365 | Medulla: 29.8%, Pons: 23.1%, Thalamus: 13.7% | Myelencephalon (medulla oblongata) (Mo) - medullary reticular formation (MoRF) & efferent nuclei of cranial nerves in the medulla oblongata (MoEN) - MoRF-MoEN | Ependymal |  |
| 24 | 0.075611096 | 0.05629494 | Thalamus: 48.8%, Cerebral cortex: 25.0%, Basal forebrain: 10.7% | Thalamus (THM) - lateral nuclear complex of thalamus (LNC) - Pulvinar of thalamus - Pul | Vascular |  |
| 20 | 0.075639929 | 0.110103417 | Thalamus: 28.4%, Midbrain: 12.2%, Medulla: 11.8% | Thalamus (THM) - Anterior nuclear complex - ANC | Vascular |  |
| 210 | 0.078310158 | 0.216267552 | Basal forebrain: 96.3%, Thalamus: 2.9%, Amygdala: 0.6% | Basal nuclei (BN) - Body of the Caudate - CaB | Medium spiny neuron | GABA |
| 435 | 0.083051046 | 0.384335177 | Thalamus: 89.3%, Midbrain: 8.9%, Pons: 1.3% | Thalamus (THM) - Lateral nuclear complex of thalamus (LNC) - ventral group of lateral nucleus - VLN | Midbrain-derived inhibitory | GABA |
| 32 | 0.084239081 | 0.692825402 | Basal forebrain: 47.3%, Medulla: 20.3%, Hypothalamus: 12.4% | Basal forebrain (BF) - septal nuclei - SEP | Fibroblast |  |
| 361 | 0.087820019 | 0.671686983 | Midbrain: 99.1%, Pons: 0.8%, Thalamus: 0.1% | Midbrain (M) - Superior colliculus and nearby nuclei - SC | Splatter | GABA |
| 313 | 0.096371545 | 0.371255398 | Medulla: 61.0%, Pons: 34.8%, Cerebellum: 3.6% | Myelencephalon (medulla oblongata) (Mo) - afferent nuclei of cranial nerves in medulla oblongata - MoAN | Upper rhombic lip | VGLUT1 |
| 11 | 0.09714066 | 0.974650473 | Basal forebrain: 27.7%, Midbrain: 22.1%, Cerebral cortex: 10.3% | Basal nuclei (BN) - Globus pallidus (GP) - Internal segment of globus pallidus - GPi | Microglia |  |
| 349 | 0.097838428 | 0.664233268 | Medulla: 65.6%, Spinal cord: 26.9%, Pons: 6.0% | Myelencephalon (medulla oblongata) (Mo) - afferent nuclei of cranial nerves in medulla oblongata - MoAN | Splatter | GLY |
| 60 | 0.098791865 | 0.3497836 | Pons: 36.2%, Cerebellum: 36.1%, Medulla: 10.9% | Cerebellum (CB) - Cerebellar deep nuclei - CbDN | Astrocyte |  |
| 434 | 0.103916099 | 0.229174664 | Midbrain: 89.4%, Thalamus: 8.3%, Hypothalamus: 1.3% | Midbrain (M) - Superior colliculus and nearby nuclei - SC | Midbrain-derived inhibitory | GABA |
| 23 | 0.106946096 | 0.267637088 | Thalamus: 34.2%, Cerebral cortex: 15.9%, Basal forebrain: 10.2% | Cerebellum (CB) - Cerebellar Vermis - CBV | Vascular |  |
| 85 | 0.107915615 | 0.090900044 | Hippocampus: 98.5%, Amygdala: 0.8%, Cerebral cortex: 0.7% | Head of hippocampus (HiH) - Uncal CA1-CA3 | Deep-layer corticothalamic and 6b | VGLUT1 VGLUT2 VGLUT3 |
| 391 | 0.109397804 | 0.270629352 | Thalamus: 81.4%, Amygdala: 5.6%, Pons: 5.3% | Thalamus (THM) - posterior nuclear complex of thalamus (PoN) - lateral geniculate nucleus (LG) | Splatter | GABA |
| 26 | 0.110726153 | 0.156789598 | Thalamus: 22.3%, Cerebral cortex: 20.0%, Pons: 15.0% | Hypothalamus (HTH) - mammillary region of HTH (HTHma) - mammillary nucleus - MN | Fibroblast |  |
| 8 | 0.111747143 | 0.955517972 | Basal forebrain: 32.0%, Midbrain: 19.4%, Pons: 12.5% | Basal nuclei (BN) - Globus pallidus (GP) - Internal segment of globus pallidus - GPi | Microglia |  |
| 359 | 0.11264309 | 0.458931442 | Midbrain: 91.5%, Thalamus: 7.8%, Pons: 0.4% | Midbrain (M) - Superior colliculus and nearby nuclei - SC | Splatter | GABA |
| 395 | 0.115893969 | 0.288259382 | Pons: 92.2%, Midbrain: 6.7%, Medulla: 0.5% | Pons (Pn) - Pontine reticular formation (PnRF) - parabrachial nuclei - PB | Splatter | VGLUT2 |
| 380 | 0.116968159 | 0.337296122 | Pons: 99.2%, Medulla: 0.8%, Cerebral cortex: 0.0% | Pons (Pn) - efferent nuclei of cranial nerves in pons - PnEN | Splatter | GLY |
| 9 | 0.12007451 | 0.780325225 | Hippocampus: 19.2%, Midbrain: 18.4%, Basal forebrain: 11.1% | Midbrain (M) - Substantia Nigra - SN | Microglia |  |
| 390 | 0.12521802 | 0.306903514 | Pons: 67.3%, Midbrain: 29.6%, Thalamus: 2.4% | Pons (Pn) - efferent nuclei of cranial nerves in pons - PnEN | Splatter | GABA |
| 342 | 0.128855561 | 0.999993012 | Pons: 98.8%, Medulla: 1.2%, Cerebral cortex: 0.0% | Pons (Pn) - other nuclei in pontine tegmentum (XPnTg) - dorsal tegmental nucleus - DTg | Splatter | VGLUT2 |
| 350 | 0.135435806 | 0.108711796 | Medulla: 84.7%, Spinal cord: 12.7%, Pons: 1.7% | Myelencephalon (medulla oblongata) (Mo) - medullary reticular formation (MoRF) & efferent nuclei of cranial nerves in the medulla oblongata (MoEN) - MoRF-MoEN | Splatter | GLY |
| 436 | 0.139804335 | 0.303354511 | Midbrain: 54.9%, Thalamus: 43.2%, Pons: 0.8% | Midbrain (M) - Substantia Nigra, Red Nucleus, and nearby nuclei - SN-RN | Midbrain-derived inhibitory | GABA |
| 70 | 0.141061719 | 0.577978603 | Hippocampus: 69.2%, Basal forebrain: 11.2%, Amygdala: 7.6% | Head of hippocampus (HiH) - CA1 | Ependymal |  |
| 321 | 0.147522058 | 0.227730392 | Pons: 90.4%, Medulla: 6.2%, Basal forebrain: 3.2% | Pons (Pn) - Pontine Nucleus - PN | Lower rhombic lip | VGLUT1 VGLUT2 |
| 40 | 0.154340463 | 0.631700044 | Cerebral cortex: 28.1%, Hippocampus: 19.7%, Amygdala: 14.6% | Head of hippocampus (HiH) - CA1 | Committed oligodendrocyte precursor |  |
| 319 | 0.15541751 | 0.513157932 | Pons: 98.8%, Medulla: 0.7%, Midbrain: 0.5% | Pons (Pn) - Pontine Nucleus - PN | Lower rhombic lip | VGLUT1 VGLUT2 |
| 183 | 0.161409756 | 0.592538277 | Hippocampus: 100.0%, Cerebral cortex: 0.0%, Amygdala: 0.0% | Tail of Hippocampus (HiT) - Caudal Hippocampus - CA1-CA3 | Hippocampal CA1-3 | VGLUT1 |
| 317 | 0.169690463 | 0.482982862 | Pons: 76.6%, Medulla: 22.0%, Midbrain: 0.6% | Pons (Pn) - efferent nuclei of cranial nerves in pons - PnEN | Lower rhombic lip | VGLUT1 VGLUT2 |
| 10 | 0.174447726 | 0.741484994 | Cerebral cortex: 27.2%, Midbrain: 16.6%, Hippocampus: 15.0% | Midbrain (M) - Substantia Nigra, Red Nucleus, and nearby nuclei - SN-RN | Microglia |  |
| 75 | 0.185599553 | 0.447221589 | Basal forebrain: 68.3%, Thalamus: 13.3%, Medulla: 7.3% | Basal forebrain (BF) - septal nuclei - SEP | Ependymal |  |
| 216 | 0.186360758 | 0.328210536 | Basal forebrain: 85.0%, Amygdala: 9.2%, Cerebral cortex: 3.6% | Basal nuclei (BN) - Nucleus Accumbens - NAC | Medium spiny neuron | GABA |
| 360 | 0.190302265 | 0.343646817 | Midbrain: 99.1%, Thalamus: 0.4%, Pons: 0.3% | Midbrain (M) - Superior colliculus and nearby nuclei - SC | Splatter | GABA |
| 1 | 0.19060705 | 0.581758964 | Midbrain: 21.0%, Basal forebrain: 19.0%, Pons: 14.3% | Basal nuclei (BN) - Globus pallidus (GP) - Internal segment of globus pallidus - GPi | Miscellaneous |  |
| 438 | 0.191300525 | 0.50121888 | Thalamus: 85.5%, Midbrain: 13.8%, Pons: 0.5% | Thalamus (THM) - lateral nuclear complex of thalamus (LNC) - Pulvinar of thalamus - Pul | Midbrain-derived inhibitory | GABA |
| 27 | 0.194830382 | 0.07119184 | Thalamus: 20.7%, Basal forebrain: 15.4%, Pons: 13.1% | Basal forebrain (BF) - septal nuclei - SEP | Fibroblast |  |
| 358 | 0.19593788 | 0.363560702 | Midbrain: 97.4%, Pons: 1.6%, Thalamus: 0.9% | Midbrain (M) - Superior colliculus and nearby nuclei - SC | Splatter | GABA |
| 433 | 0.198702602 | 0.565225518 | Hypothalamus: 79.8%, Midbrain: 19.6%, Thalamus: 0.6% | Hypothalamus (HTH) - mammillary region of HTH (HTHma) - tuberal region of HTH - HTHtub | Splatter | HDC |
| 393 | 0.20002195 | 0.253812103 | Amygdala: 39.6%, Basal forebrain: 32.7%, Hypothalamus: 21.2% | Extended amygdala (EXA) - Bed nucleus of stria terminalis and nearby - BNST | Splatter | GABA |
| 18 | 0.207408892 | 0.944106959 | Hypothalamus: 26.7%, Medulla: 21.8%, Pons: 15.8% | Hypothalamus (HTH) - mammillary region of HTH (HTHma) - mammillary nucleus - MN | Vascular |  |
| 343 | 0.215483302 | 0.049125688 | Cerebellum: 97.0%, Pons: 1.8%, Midbrain: 0.8% | Cerebellum (CB) - Cerebellar deep nuclei - CbDN | Splatter | VGLUT2 |
| 68 | 0.22134428 | 0.068697602 | Medulla: 52.7%, Spinal cord: 22.3%, Pons: 17.8% | Spinal cord - SpC | Ependymal |  |
| 374 | 0.229532803 | 0.318516122 | Midbrain: 61.4%, Thalamus: 36.6%, Amygdala: 0.7% | Midbrain (M) - Pretectal region - PTR | Splatter | VGLUT2 |
| 310 | 0.232276812 | 0.672686935 | Pons: 89.0%, Cerebellum: 8.6%, Medulla: 1.6% | Pons (Pn) - efferent nuclei of cranial nerves in pons - PnEN | Upper rhombic lip | VGLUT1 VGLUT2 |
| 340 | 0.238382276 | 0.537861415 | Pons: 99.4%, Medulla: 0.4%, Midbrain: 0.2% | Pons (Pn) - other nuclei in pontine tegmentum (XPnTg) - dorsal tegmental nucleus - DTg | Splatter | GLY VGLUT2 |
| 356 | 0.249502572 | 0.563723246 | Midbrain: 87.7%, Pons: 9.3%, Thalamus: 2.4% | Midbrain (M) - Inferior colliculus and nearby nuclei - IC | Splatter | GABA |
| 357 | 0.260578211 | 0.994448811 | Midbrain: 67.5%, Pons: 20.1%, Thalamus: 9.5% | Midbrain (M) - Pretectal region - PTR | Splatter | GABA |
| 412 | 0.262768646 | 0.447787693 | Hypothalamus: 99.2%, Amygdala: 0.2%, Midbrain: 0.2% | Hypothalamus (HTH) - mammillary region of HTH (HTHma) - tuberal region of HTH - HTHtub | Splatter | VGLUT2 |
| 47 | 0.269732525 | 0.753243547 | Thalamus: 22.7%, Midbrain: 19.9%, Cerebral cortex: 13.7% | Midbrain (M) - Substantia Nigra, Red Nucleus, and nearby nuclei - SN-RN | Oligodendrocyte |  |
| 4 | 0.278386539 | 0.4012777 | Thalamus: 21.9%, Pons: 16.1%, Basal forebrain: 13.9% | Pons (Pn) - efferent nuclei of cranial nerves in pons - PnEN | Miscellaneous |  |
| 318 | 0.281120135 | 0.408137514 | Pons: 97.6%, Medulla: 2.1%, Midbrain: 0.2% | Pons (Pn) - Pontine reticular formation - PnRF | Lower rhombic lip | VGLUT1 VGLUT2 |
| 454 | 0.286052279 | 0.983574934 | Thalamus: 82.7%, Midbrain: 17.3%, Cerebral cortex: 0.0% | Thalamus (THM) - posterior nuclear complex of thalamus (PoN) - lateral geniculate nucleus (LG) | Thalamic excitatory | VGLUT2 |
| 82 | 0.29148185 | 0.826487468 | Basal forebrain: 73.7%, Medulla: 25.6%, Pons: 0.6% | Basal forebrain (BF) - septal nuclei - SEP | Choroid plexus |  |
| 43 | 0.292388415 | 0.579929818 | Thalamus: 92.7%, Midbrain: 2.3%, Pons: 1.5% | Thalamus (THM) - Lateral nuclear complex of thalamus (LNC) - ventral group of lateral nucleus - VLN | Committed oligodendrocyte precursor |  |
| 206 | 0.317930239 | 0.71091929 | Hippocampus: 96.5%, Midbrain: 3.4%, Thalamus: 0.0% | Head of hippocampus (HiH) - Uncal DG-CA4 | Hippocampal dentate gyrus | VGLUT1 |
| 72 | 0.33215646 | 0.193464108 | Basal forebrain: 41.0%, Hypothalamus: 26.7%, Thalamus: 14.5% | Basal nuclei (BN) - Body of the Caudate - CaB | Ependymal |  |
| 185 | 0.33843537 | 0.274868756 | Hippocampus: 100.0%, Amygdala: 0.0%, Cerebral cortex: 0.0% | Head of hippocampus (HiH) - Uncal CA1-CA3 | Hippocampal CA1-3 | VGLUT1 |
| 338 | 0.341306926 | 0.705479733 | Basal forebrain: 81.9%, Midbrain: 17.6%, Hypothalamus: 0.5% | Basal nuclei (BN) - Globus pallidus (GP) - Internal segment of globus pallidus - GPi | Splatter | GABA |
| 322 | 0.342202575 | 0.129113216 | Pons: 95.9%, Medulla: 3.3%, Midbrain: 0.4% | Pons (Pn) - efferent nuclei of cranial nerves in pons - PnEN | Lower rhombic lip | VGLUT1 VGLUT2 |
| 41 | 0.348394521 | 0.345615592 | Hippocampus: 24.8%, Cerebral cortex: 23.4%, Amygdala: 10.0% | Tail of Hippocampus (HiT) - Caudal Hippocampus - CA4-DGC | Oligodendrocyte |  |
| 362 | 0.350252284 | 0.217664462 | Midbrain: 83.4%, Thalamus: 14.3%, Pons: 1.8% | Midbrain (M) - Pretectal region - PTR | Splatter | GABA |
| 51 | 0.364642293 | 0.060266178 | Hypothalamus: 90.7%, Thalamus: 3.6%, Basal forebrain: 3.6% | Hypothalamus (HTH) - mammillary region of HTH (HTHma) - mammillary nucleus - MN | Oligodendrocyte |  |
| 7 | 0.367354538 | 0.509068289 | Hippocampus: 25.1%, Basal forebrain: 15.2%, Thalamus: 11.0% | Head of hippocampus (HiH) - CA1 | Microglia |  |
| 66 | 0.369459199 | 0.012930393 | Midbrain: 28.4%, Hippocampus: 12.5%, Pons: 12.5% | Pons (Pn) - efferent nuclei of cranial nerves in pons - PnEN | Ependymal |  |
| 363 | 0.381289211 | 0.717777585 | Pons: 39.4%, Cerebellum: 34.7%, Midbrain: 21.7% | Cerebellum (CB) - Cerebellar deep nuclei - CbDN | Splatter | GABA |
| 74 | 0.383166189 | 0.231917183 | Hippocampus: 85.9%, Amygdala: 8.2%, Basal forebrain: 1.9% | Head of hippocampus (HiH) - CA1 | Ependymal |  |
| 81 | 0.387518616 | 0.843584137 | Basal forebrain: 94.8%, Medulla: 4.5%, Hippocampus: 0.4% | Basal forebrain (BF) - septal nuclei - SEP | Choroid plexus |  |
| 50 | 0.399095339 | 0.87423809 | Pons: 30.3%, Midbrain: 20.2%, Medulla: 12.2% | Pons (Pn) - Pontine reticular formation - PnRF | Oligodendrocyte |  |
| 45 | 0.39914172 | 0.802881423 | Thalamus: 20.9%, Pons: 19.2%, Midbrain: 19.1% | Midbrain (M) - Substantia Nigra - SN | Oligodendrocyte |  |
| 6 | 0.4162229 | 0.495898118 | Hippocampus: 40.4%, Medulla: 29.6%, Basal forebrain: 20.1% | Myelencephalon (medulla oblongata) (Mo) - afferent nuclei of cranial nerves in medulla oblongata - MoAN | Microglia |  |
| 44 | 0.425361801 | 0.773319028 | Thalamus: 36.2%, Hypothalamus: 18.0%, Amygdala: 10.3% | Thalamus (THM) - Lateral nuclear complex of thalamus (LNC) - ventral group of lateral nucleus - VLN | Committed oligodendrocyte precursor |  |
| 30 | 0.428233663 | 0.130166635 | Medulla: 41.3%, Spinal cord: 39.7%, Thalamus: 4.8% | Spinal cord - SpC | Fibroblast |  |
| 42 | 0.429699553 | 0.631332237 | Amygdala: 24.7%, Hypothalamus: 23.3%, Basal forebrain: 20.2% | Hypothalamus (HTH) - supraoptic region of HTH - HTHso | Committed oligodendrocyte precursor |  |
| 5 | 0.435485603 | 0.978730559 | Spinal cord: 31.8%, Pons: 26.0%, Medulla: 13.2% | Spinal cord - SpC | Microglia |  |
| 298 | 0.471276684 | 0.848781498 | Midbrain: 97.1%, Thalamus: 2.9%, Cerebral cortex: 0.0% | Midbrain (M) - Superior colliculus and nearby nuclei - SC | Upper rhombic lip | VGLUT1 |
| 446 | 0.478428275 | 0.6724271 | Thalamus: 99.3%, Hippocampus: 0.7%, Cerebral cortex: 0.0% | Thalamus (THM) - posterior nuclear complex of thalamus (PoN) - lateral geniculate nucleus (LG) | Thalamic excitatory | VGLUT2 |
| 29 | 0.493960396 | 0.009895497 | Thalamus: 36.4%, Cerebral cortex: 12.1%, Pons: 11.0% | Thalamus (THM) - posterior nuclear complex of thalamus (PoN) - lateral geniculate nucleus (LG) | Fibroblast |  |
| 48 | 0.495065156 | 0.487293937 | Thalamus: 24.9%, Cerebral cortex: 20.9%, Hippocampus: 13.8% | Basal nuclei (BN) - Globus pallidus (GP) - External segment of globus pallidus - GPe | Oligodendrocyte |  |
| 76 | 0.500314476 | 0.860875858 | Medulla: 38.6%, Basal forebrain: 32.9%, Thalamus: 15.7% | Basal forebrain (BF) - septal nuclei - SEP | Committed oligodendrocyte precursor |  |
| 376 | 0.50973994 | 0.352659552 | Thalamus: 94.3%, Midbrain: 5.4%, Pons: 0.1% | Epithalamus - ETH | Splatter | VGLUT2 VGLUT3 |
| 28 | 0.530304471 | 0.060456661 | Thalamus: 34.3%, Cerebral cortex: 15.0%, Pons: 13.3% | Thalamus (THM) - intralaminar nuclear complex (ILN) - posterior group of intralaminar nuclei (PILN) - centromedian and parafasicular nuclei - CM and Pf | Fibroblast |  |
| 315 | 0.553136778 | 0.361919577 | Medulla: 35.7%, Pons: 33.3%, Cerebellum: 26.2% | Cerebellum (CB) - Cerebellar Vermis - CBV | Miscellaneous | GABA |
| 25 | 0.569767804 | 0.033105912 | Basal forebrain: 38.0%, Medulla: 32.3%, Hippocampus: 25.1% | Basal forebrain (BF) - septal nuclei - SEP | Fibroblast |  |
| 16 | 0.587992875 | 0.448330954 | Cerebral cortex: 23.2%, Hippocampus: 15.7%, Thalamus: 15.3% | Perirhinal cortex (area 35) (A35) - rostral subdivision of area 35 - A35r | Vascular |  |
| 77 | 0.618920861 | 0.373386905 | Basal forebrain: 55.4%, Medulla: 31.7%, Thalamus: 8.9% | Basal forebrain (BF) - septal nuclei - SEP | Choroid plexus |  |
| 73 | 0.625314054 | 0.237874955 | Hippocampus: 53.2%, Amygdala: 20.2%, Basal forebrain: 7.9% | Head of hippocampus (HiH) - CA1 | Ependymal |  |
| 80 | 0.649497949 | 0.379291565 | Basal forebrain: 94.0%, Medulla: 5.0%, Hippocampus: 0.5% | Basal forebrain (BF) - septal nuclei - SEP | Choroid plexus |  |
| 17 | 0.650038296 | 0.474853052 | Cerebral cortex: 25.5%, Hippocampus: 17.8%, Midbrain: 8.4% | Perirhinal cortex (area 35) (A35) - rostral subdivision of area 35 - A35r | Vascular |  |
| 414 | 0.683165436 | 0.844842929 | Hypothalamus: 92.1%, Midbrain: 5.8%, Pons: 1.4% | Hypothalamus (HTH) - mammillary region of HTH (HTHma) - mammillary nucleus - MN | Splatter | VGLUT2 |
| 79 | 0.707235264 | 0.393610792 | Basal forebrain: 92.5%, Medulla: 6.3%, Thalamus: 0.6% | Basal forebrain (BF) - septal nuclei - SEP | Choroid plexus |  |
| 78 | 0.736826208 | 0.566384142 | Basal forebrain: 49.7%, Medulla: 46.2%, Hippocampus: 3.0% | Basal forebrain (BF) - septal nuclei - SEP | Choroid plexus |  |
| 46 | 0.782965979 | 0.635678123 | Cerebral cortex: 20.0%, Basal forebrain: 18.5%, Midbrain: 15.8% | Basal nuclei (BN) - Globus pallidus (GP) - External segment of globus pallidus - GPe | Oligodendrocyte |  |
| 71 | 0.809723508 | 0.252410179 | Thalamus: 41.0%, Amygdala: 21.3%, Midbrain: 17.1% | Thalamus (THM) - medial nuclear complex of thalamus (MNC) - mediodorsal nucleus of thalamus - MD | Ependymal |  |
| 348 | 0.830159517 | 0.444196616 | Medulla: 68.6%, Spinal cord: 29.3%, Pons: 0.9% | Myelencephalon (medulla oblongata) (Mo) - afferent nuclei of cranial nerves in medulla oblongata - MoAN | Splatter | GLY |
| 65 | 0.847393239 | 3.19574E-05 | Hypothalamus: 93.0%, Amygdala: 1.6%, Thalamus: 1.6% | Hypothalamus (HTH) - mammillary region of HTH (HTHma) - mammillary nucleus - MN | Astrocyte |  |
| 409 | 0.85768507 | 0.658798543 | Cerebral cortex: 82.2%, Amygdala: 13.3%, Hypothalamus: 4.4% | Paleocortex (PalCx) - Anterior Olfactory Nucleus - AON | Amygdala excitatory | VGLUT1 VGLUT2 |
| 14 | 0.864631186 | 0.819599715 | Basal forebrain: 74.0%, Medulla: 23.3%, Hippocampus: 1.4% | Basal forebrain (BF) - septal nuclei - SEP | Vascular |  |
| 447 | 0.866172889 | 0.216902806 | Thalamus: 99.9%, Hippocampus: 0.0%, Midbrain: 0.0% | Thalamus (THM) - posterior nuclear complex of thalamus (PoN) - lateral geniculate nucleus (LG) | Thalamic excitatory | VGLUT2 |
| 15 | 0.874370482 | 0.487212373 | Hippocampus: 23.8%, Cerebral cortex: 23.3%, Thalamus: 10.3% | Perirhinal cortex (area 35) (A35) - rostral subdivision of area 35 - A35r | Vascular |  |
| 83 | 0.893454984 | 0.597413251 | Basal forebrain: 76.6%, Medulla: 22.2%, Pons: 1.2% | Basal forebrain (BF) - septal nuclei - SEP | Choroid plexus |  |
| 352 | 0.921339803 | 0.06017857 | Midbrain: 90.0%, Thalamus: 8.2%, Hypothalamus: 1.3% | Midbrain (M) - Substantia Nigra, Red Nucleus, and nearby nuclei - SN-RN | Splatter | GABA |
| 13 | 0.925492437 | 0.856468416 | Basal forebrain: 29.4%, Cerebral cortex: 14.0%, Cerebellum: 12.2% | Basal nuclei (BN) - Globus pallidus (GP) - Internal segment of globus pallidus - GPi | Microglia |  |
| 31 | 0.931704496 | 0.005019906 | Cerebral cortex: 35.3%, Basal forebrain: 29.7%, Medulla: 10.6% | Basal forebrain (BF) - septal nuclei - SEP | Fibroblast |  |
| 49 | 0.939835117 | 0.076093252 | Basal forebrain: 47.8%, Thalamus: 42.2%, Hypothalamus: 5.3% | Basal nuclei (BN) - Globus pallidus (GP) - External segment of globus pallidus - GPe | Oligodendrocyte |  |

Note:

Cluster: cell cluster identifier from the human brain single-cell transcriptomic atlas (Siletti et al., 2023);

P1: TDEP enrichment P-value using genome-wide GERD GWAS summary statistics;

P2: TDEP enrichment P-value using GERD GWAS signals restricted to genomic regions shared with F1-loading internalizing psychiatric disorders;

Top Brain Regions: top three brain regions by proportion of cells in the cluster;

Anatomical Detail: detailed anatomical dissection and cortical area annotation;

Supercluster: supercluster cell-type category as defined in Siletti et al., 2023;

Neurotransmitter: neurotransmitter auto-annotation (e.g., GABA, VGLUT1, VGLUT2).
